# scEPS integrates genetic and single-cell disease atlas data to provide granular mechanistic insights into complex human diseases

**DOI:** 10.64898/2026.06.26.26356714

**Authors:** Luli Zou, Owen Whitley, Hsin-Wei Tseng, Caitlin Simopoulos, Diana Chang, Ruoyu Zhang, Amy Stockwell, Wuming Gong, Kipper Fletez-Brant, Tawaun Lucas, Ephraim Kenigsberg, Alsu Missarova, David Garfield, Brian Yaspan, Mark McCarthy, Anubha Mahajan, Josh Kaminker, Huwenbo Shi

## Abstract

Integrating GWAS and single-cell data holds great potential for prioritizing causal disease biology at cellular resolution. Recent integrative approaches typically assess the enrichment of disease genetic signals in cell types or individual cells, without directly modeling disease phenotypes. We develop a new method, single-cell Expression exPlainability Statistics (scEPS), for identifying disease-associated cell neighborhoods, by explicitly testing whether the expression of GWAS-prioritized genes explains more variance in a disease than randomly selected, mean-expression-matched control genes. Crucially, when applied to PRSs of healthy donors, scEPS captures the genetic covariance between gene expression and diseases, mitigating the effect of reverse causation and prioritizing cell populations mediating the effects of GWAS genes. We applied scEPS to clinical diagnoses and PRSs of 4 neurological and 4 respiratory disorders, integrating brain and lung cell atlas data, respectively, with respective GWAS summary statistics data. scEPS recapitulated known and uncovered novel disease-associated cell populations, identifying 1.77× (s.e. 1.21) and 5.13× (s.e. 3.08) more significant associations than a CNA-based approach and scDRS, respectively. Furthermore, scEPS detected different cell populations, contrasting clinical diagnoses vs. their PRSs, revealing distinct biology for the active/symptomatic vs. preclinical/asymptomatic states of the disease. Finally, we observed limited concordance across methods using distinct definitions of disease association, underscoring the need to integrate complementary insights for holistic understanding of disease biology.

## Introduction

Understanding the causal biological mechanisms underlying diseases is critical for the development of successful therapeutics [1, 2, 3, 4, 5, 6, 7, 8]. Genome-wide association study (GWAS) provides a powerful tool for discovering disease-causal variants [8, 9, 10, 11, 12, 13, 14, 15, 16]. However, the complex biological and cellular processes connecting genetic and phenotypic variation renders it difficult to derive actionable mechanistic disease insights from GWAS data alone. Conversely, while single-cell data enables investigating diseases at cellular resolution [17, 18, 19, 20, 21, 22, 23], it remains challenging to distinguish causal upstream biological processes from downstream responses of diseases. Integrating the causal signals from GWAS with high resolution single-cell data, thus, holds great potential for discovering causal disease biology at cellular resolution.

Several recent methods integrating GWAS with single-cell RNA-seq (scRNA-seq) and ATAC-seq (scATAC-seq) data have yielded tremendous insights into causal disease biology at cellular resolution [24, 25, 26, 27]. Fundamentally, these methods test for the excess overlap of GWAS signals (e.g., GWAS-prioritized genes, fine-mapped variants) with single-cell data (e.g., gene expression, chromatin accessibility peaks), but do not directly model the disease phenotypes of the donors represented in the single-cell data. And it remains challenging to determine the cell populations that translate the expression of GWAS-prioritized genes to differences in disease phenotypes across patients with these methods.

Here, we developed <u>s</u>ingle-<u>c</u>ell <u>E</u>xpression ex<u>P</u>lainability <u>S</u>tatistics (scEPS), for identifying cell neighborhoods, a small group of cells with similar gene expression profile [28, 29, 30], in scRNA-seq disease atlas data, in which the neighborhood-specific expression of disease GWAS genes (e.g., prioritized by MAGMA [13]) explains excess differences in the disease phenotype across patients, by directly modeling the disease as the outcome of gene expression. Specifically, for each cell neighborhood, scEPS estimates a *d statistic*, the difference in the variance in disease explained by the variation (both up and down regulation) in the expression of each GWAS gene vs. each randomly selected mean-expression-matched control gene. For a group of cell neighborhoods (e.g., within a specific cell type), scEPS estimates the average *d* statistics across the cell neighborhoods constituting the group. Compared to existing integrative approaches (e.g., scDRS [24], sc-linker [31]) that test for the enrichment of GWAS signals (e.g., gene expression, heritability, etc.) in scRNA-seq data, scEPS aims to identify cell populations, where differential expression of GWAS genes directly translates to meaningful differences in disease phenotypes. Crucially, when applied to the disease polygenic risk scores (PRSs) of control donors, scEPS captures the genetic covariance between gene expression and the disease. This mitigates the impact of reverse causation and prioritizes cell populations that likely mediate the effects of GWAS genes.

We confirmed via extensive simulations that scEPS is unbiased and yields well-calibrated test statistics. We then applied scEPS to 4 neurological and 4 respiratory disorders (including clinical diagnoses and PRSs), integrating the respective GWAS summary statistics data (average N=434K) with single-cell brain atlas [32] (1,203,090 cells from 81 donors) and lung atlas [33] (475,047 cells from 59 donors) data. Next, we investigated whether scEPS recapitulated known and uncovered novel disease-associated cell populations and biological processes, by explicitly modeling the disease. We also sought to distinguish biological processes that are likely upstream vs. downstream of the diseases, by contrasting analyses of diseases with their respective PRSs. Finally, we investigated whether different metrics of disease association could yield distinct biological insights, by comparing the cell types implicated by scEPS vs. those implicated by existing single-cell methods, CNA [34] and scDRS [24].

## Results

### Overview of the methods

scEPS is a method to identify disease-associated cell neighborhood, a small group of cells with similar transcriptional profile [28, 29, 30], incorporating GWAS summary statistics and single-cell disease atlas data (Figure 1, Supplementary Table 1). scEPS determines that a cell neighborhood is associated with the disease if the expression of each GWAS gene (more likely to be disease causal [9, 35, 36]) explains more variance in the disease across donors than each randomly selected control gene matched on mean expression. When applied to polygenic risk scores (PRS) of control donors, scEPS captures the genetic covariance between gene expression and the disease (“Supplementary notes”).

**Figure 1:**
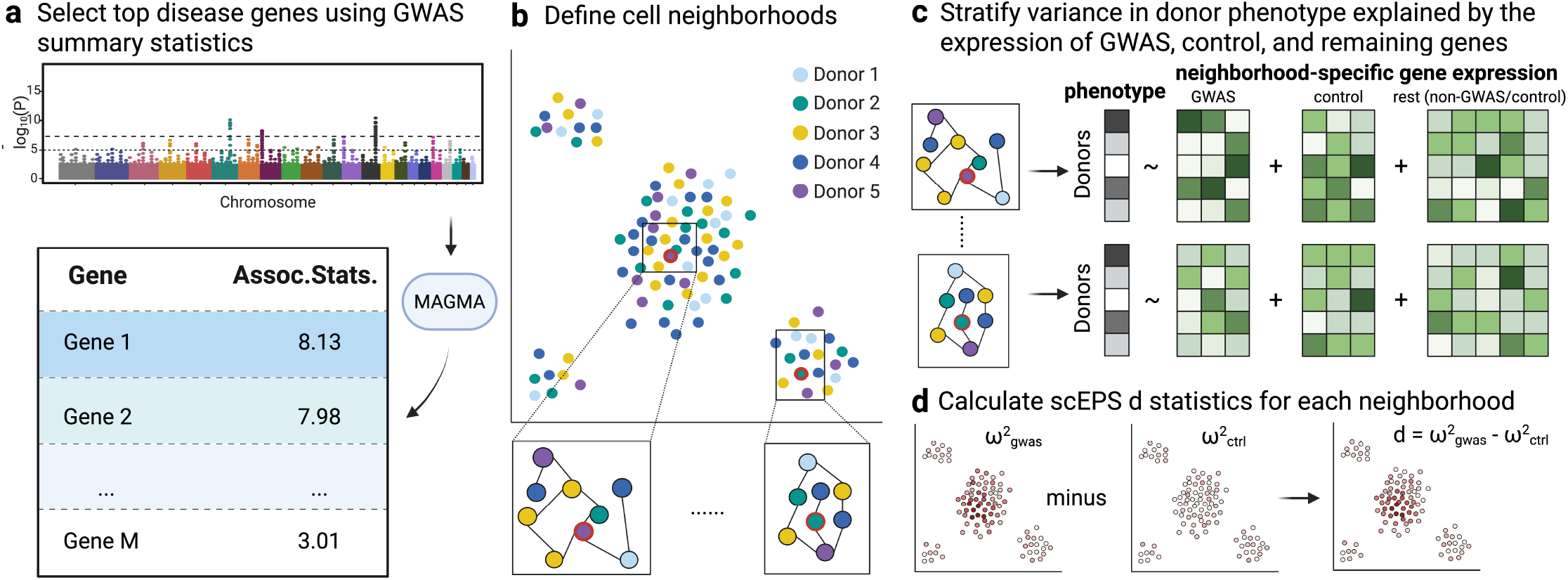
Overview of the scEPS methods. (**a**) scEPS uses MAGMA to prioritize disease-associated genes using GWAS summary statistics data (i.e., GWAS genes). (**b**) scEPS defines a neighborhood anchored at each individual cell based on the k-NN graph for cells in a single-cell data. We highlight 2 neighborhoods in boxes on the UMAP plot for a sample scRNA-seq data, where the anchor cells are circled in red. (**c**) At each cell neighborhood, scEPS models donors’ phenotypes (e.g., clinical diagnoses, PRSs, etc.) as a linear function of the neighborhood-specific expression of GWAS genes, randomly selected control genes matched on mean expression of the GWAS genes, and the remaining non-GWAS/control genes. Here, scEPS will yield a high and low *d* statistics for the first and second neighborhood, respectively, as the expression of GWAS genes better captures donors’ phenotypes in the first vs. the second neighborhood. (**d**) For each neighborhood, scEPS calculates *d* statistic as the difference between *ω*^2^_*gwas*_ and *ω*^2^_*ctrl*_.

We choose the set of disease GWAS genes based on the MAGMA [13] gene-level association statistics derived from the GWAS summary statistics data. By default, we include genes with MAGMA FDR < 0.05 (“Methods”). To avoid including too few or too many genes, we set the minimum and maximum number of genes to 500 and 2,000, respectively, by default.

At each cell neighborhood, we model the relationship between the pseudo-bulk expression of protein coding genes and the disease phenotype using a linear model. Let *y*_*i*_ and *y*_*i*′_ represent the mean-centered disease phenotypes, of individual *i* and *i*′, respectively, the expected value of *y*_*i*_*y*_*i*′_ is then,

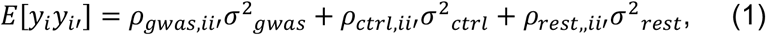

where *ρ*_*gwas,ii*′_ = (∑_*j*_ *x*_*gwas,ij*_*x*_*gwas,i*′*j*_/*m, ρ*_*ctrl,ii*′_ = (∑_*j*_ *x*_*ctrl,ij*_*x*_*ctrl,i*′*j*_/*m*, and *ρ*_*rest,ii*′_ = (∑_*j*_ *x*_*rest,ij*_*x*_*rest,i*′*j*_/*m*_*rest*_, represent the covariance between individual *i* and *i*′ across the mean centered expression of *m* GWAS genes (*x*_*gwas,ij*_, *x*_*gwas,i*′*j*_), *m* randomly selected control genes (*x*_*ctrl,ij*_, *x*_*ctrl,i*′*j*_) matched on mean expression of the GWAS genes prior to centering, and *m*_*rest*_ remaining genes (*x*_*rest,ij*_, *x*_*rest,i*′*j*_); *σ*^2^_*gwas*_, *σ*^2^_*ctrl*_, and *σ*^2^_*rest*_ represent the variances of the effect sizes of the expression of each GWAS, control, and remaining gene on the disease phenotype, respectively.

We define the scEPS *d* statistic at a neighborhood as,

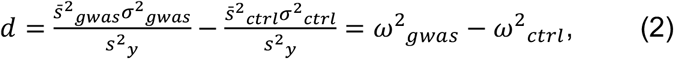

where 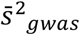 and 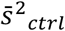 represent the average variance of the expression of GWAS and control genes, respectively; *s*^2^_*y*_ is the variance of the disease phenotype; *ω*^2^_*gwas*_ and *ω*^2^_*ctrl*_ represent the variance in disease explained by each GWAS and control gene, respectively. We determine that a cell neighborhood is associated with the disease if *d* > 0. We also define 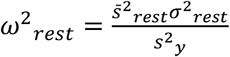, representing the variance in disease explained by each of the remaining genes, where 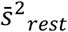 is the average variance of the expression of these genes; and 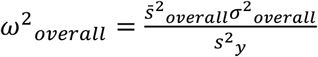, representing the variance in disease explained by each gene (both GWAS and non-GWAS genes), where 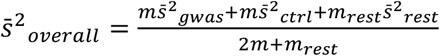 and 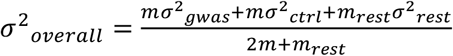.

For a group of cell neighborhoods (e.g., from the same cell type), we aggregate the *d* statistics by averaging across the cell neighborhoods constituting the group, i.e., aggregated *d*, 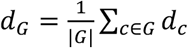, where *G* represents the group of cell neighborhoods. We aggregate other scEPS statistics, *ω*^2^_*gwas*_, *ω*^2^_*ctrl*_, *ω*^2^_*rest*_, and *ω*^2^_*overall*_, similarly.

scEPS defines a neighborhood of cells, anchored at each individual cell, based on the k-nearest-neighbor (k-NN) graph, balancing between neighborhood size and even contribution of cells from all donors. Briefly, we first calculate a neighborhood abundance matrix, ***Q***^*s*^, representing the proportions of cells from each donor in each cell neighborhood, by performing a random walk of length *s* on the k-NN graph, starting from each cell. We determine the parameter, *s*, using the heuristic approach described in ref. [34], which promotes a uniform contribution of cells from the donors across the cell neighborhoods. Within the neighborhood of each anchor cell, we include all cells that can be reached within a random walk of length *s* with a probability greater than *θ* from the anchor cell. We determine the optimal value for *θ, θ*^*\**^, for each cell neighborhood *c*, by minimizing the difference between the target neighborhood abundance, ***Q***_*c*_^*s*^, and the realized neighborhood abundance, while ensuring sufficient numbers of donors and cells are included (“Methods”). For sparse local regions of the k-NN graph, where a globally determined *s* fails to capture sufficient numbers of cells or donors in a neighborhood at any level of *θ*, we adaptively increase *s* and select *θ* based on the updated *s* (“Methods”).

We provide further details of the scEPS methods, including derivation of the scEPS model, methods of moment approach for estimating the scEPS statistics, statistical testing procedures, default parameter settings, and relationship between scEPS statistics and genetic covariance when analyzing PRS, in the “Methods” section and “Supplementary notes”. We have publicly released the software implementing the method (see “Code availability”).

### Performance of scEPS in simulations

We evaluated the performance of scEPS in simulations using the publicly available single-nucleus (snRNA-seq) data for 37,720 microglia and perivascular macrophages (microglia-PVM) from 81 donors of self-reported European ancestry from the Seattle Alzheimer’s Disease Brain Cell Atlas (SEA-AD) consortium [32]. For each set of simulation parameters, we simulated phenotypes as a linear function of the pseudo-bulk expression of 17,095 (out of 36,601) protein-coding genes, at cell neighborhoods anchored at randomly selected cells (“Methods”). We randomly selected a fraction, *p*_*gwas,causal*_, of causal GWAS genes from the top *m*_*sim*_ Alzheimer’s disease (AD) GWAS genes with the highest MAGMA Z-scores, derived from the AD GWAS summary statistics data [37]. We also randomly selected a fraction, *p*_*non*−*gwas,causal*_, of causal non-GWAS genes from the remaining genes. We sampled the effect sizes of the causal GWAS and non-GWAS genes from Gaussian distributions with mean zero, varying the variance parameters to attain the desired *λ*_*gwas*_ (enrichment of variance in phenotype explained by each GWAS gene) and *η*^2^_*total*_ (total variance in the phenotype explained by all genes). We assessed the performance of scEPS for individual cell neighborhoods and for groups of cell neighborhoods. We performed full null (*η*^2^_*total*_ = 0), semi null (*η*^2^_*total*_ > 0, *λ*^2^_*gwas*_ = 1), and causal (*η*^2^_*total*_ > 0, *λ*^2^_*gwas*_ > 1) simulations. We provide detailed descriptions of the simulations in the “Methods” section.

We report the results in Figure 2, Supplementary Figure 2–28, and Supplementary Table 2–5. First, we evaluated the performance of scEPS for individual cell neighborhoods in simulations without model misspecification. scEPS yielded approximately unbiased estimates of *d* across a wide range of *λ*_*gwas*_, *η*^2^_*total*_, *p*_*gwas,causal*_, *p*_*non*−*gwas,causal*_, neighborhood size, and number of donors in the neighborhoods (Figure 2**a**, Supplementary Figure 2**c, e, g, i, k, m**, Supplementary Table 2**a**). The p-values for *d* were generally calibrated with controlled type I errors (at *α* < 0.05) in both full null and semi null simulations (Figure 2**b**, Supplementary Figure 2**b, d, o, p**, Supplementary Table 2**a**). The statistical power of scEPS generally increased with *λ*_*gwas*_, *η*^2^_*gwas*_, neighborhood size, and size of GWAS gene set (Figure 2; Supplementary Figure 2**d, l, n**, Supplementary Table 2**a**). We note that scEPS underestimated *d* in simulations, where a total 10 GWAS genes were randomly selected to be causal (Supplementary Figure 2**m**, Supplementary Table 2**a**), although the estimated *d* still correlated well with the simulated *d*. This downward bias was not driven by, *m*, the number of GWAS genes analyzed (Supplementary Figure 24, Supplementary Table 5**a**, also see below). In practice, we recommend applying scEPS to analyze diseases, for which at least 50 GWAS genes are expected to be associated with the disease. We confirmed that scEPS also yielded approximately unbiased estimates of *ω*^2^_*gwas*_, *ω*^2^_*ctrl*_, *ω*^2^_*rest*_ and *ω*^2^_*overall*_ across a wide range of simulations, with calibrated p-values and controlled type I errors in full null simulations (Supplementary Figure 3–6, Supplementary Table 2**a**).

**Figure 2:**
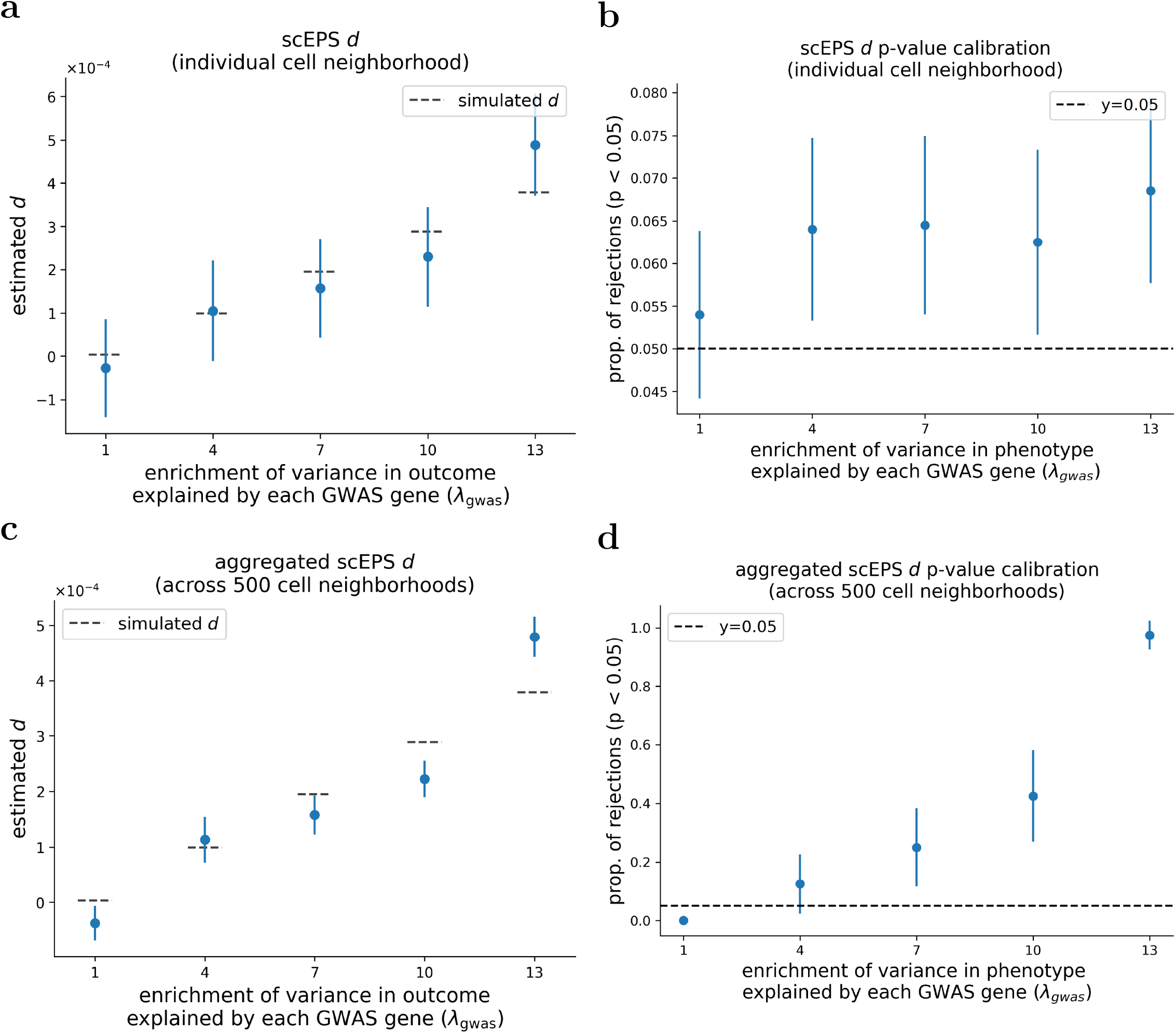
Performance of scEPS in estimating the *d* statistics in simulations. (**a, b**) Average estimated *d* statistics and proportion of rejected null hypotheses, respectively, for individual cell neighborhoods across different simulated *λ*^2^_*gwas*_. Mean and standard errors were obtained based on 2,000 simulations. (**c, d**) Average estimated aggregated *d* statistics and proportion of rejected null hypotheses, respectively, for groups of cell neighborhoods of size 500, across different simulated *λ*^2^_*gwas*_. Mean and standard errors were based on 40 simulations. Error bars represent 1.96× standard errors on both sides. Numerical results are reported in Supplementary Table 2.

Next, we evaluated the performance of scEPS for groups of cell neighborhoods. scEPS yielded approximately unbiased estimates of aggregated *d* across groups of different sizes, in a wide range of simulations (Figure 2**c**, Supplementary Figure 7–9, Supplementary Table 3). For cell neighborhood groups of size greater than 100 p-values for aggregated *d* were generally calibrated with controlled type I errors in both full null and semi null simulations (Supplementary Figure 7–9, Supplementary Table 3**b, c**). However, we observed less calibrated p-values and elevated type I errors in null simulations, in which the size of the group was 20 (Supplementary Figure 7**b, d, k, l**, Supplementary Table 3**a**). Thus, when aggregating scEPS *d* statistics, we recommend restricting to groups with a size of at least 100 neighborhoods. The statistical power of scEPS generally increased with *λ*_*gwas*_, *η*^2^_*gwas*_, and the size of GWAS gene set (Figure 2**b**; Supplementary Figure 2**d, l, n**, Supplementary Table 3). We confirmed that for groups with at least 100 cell neighborhoods, scEPS also yielded approximately unbiased estimates of aggregated *ω*^2^_*gwas*_, *ω*^2^_*ctrl*_, *ω*^2^_*rest*_ and *ω*^2^_*overall*_ across a wide range of simulations, with calibrated p-values and controlled type I errors in full null simulations (Supplementary Figure 10–21, Supplementary Table 3). We also confirmed that the calibration of the scEPS’ p-values in null simulations was robust to the number of approximately independent cell neighborhood blocks used in statistical bootstrap (Supplementary Figure 22, 23, Supplementary Table 4).

We performed 3 simulations to assess the performance of scEPS for individual cell neighborhoods, in simulations with model misspecification. First, we ran scEPS to analyze GWAS gene sets consisting of 300 genes (i.e., *m* = 300), using simulated data with *m*_*sim*_ = 600. scEPS was approximately unbiased and yielded controlled type I errors in null simulation (Supplementary Figure 24, Supplementary Table 5**a**), suggesting that scEPS was robust when analyzing smaller but more stringent GWAS gene sets. Second, we ran scEPS with *m* = 900 on the same simulated data. As expected, the estimated *d* was biased downwards (Supplementary Figure 25, Supplementary Table 5**b**), as the set of GWAS genes analyzed by scEPS contained genes with smaller effect sizes. In practice, we recommend applying stringent thresholds for selecting GWAS genes (MAGMA FDR < 0.05 by default, “Methods”). Third, we ran scEPS on simulated data in which 50% of the cells in each neighborhood were randomly removed. scEPS was still approximately unbiased with controlled type I errors (Supplementary Figure 26, Supplementary Table 5**c**). As expected, the statistical power decreased due to lower number of cells in the cell neighborhoods, contributing to decreased signal-to-noise ratio in the pseudo-bulk expression (Supplementary Figure 2**d**, Supplementary Figure 26**d**, Supplementary Table 5**c**).

We also assessed the robustness of scEPS’ statistical bootstrap testing procedure for individual cell neighborhoods. First, we compared the p-values for individual cell neighborhoods obtained using bootstrap vs. permutation. We confirmed that the p-values obtained using both approaches were highly concordant across a wide range of simulations (Supplementary Figure 27), justifying the choice of using the more computationally and statistically efficient bootstrap approach. Second, we ran scEPS using 100 (instead of the default 1,000) bootstrapped samples for statistical testing. scEPS still yielded generally calibrated p-values with controlled type I errors in null simulations with much fewer bootstrap samples (Supplementary Figure 28, Supplementary Table 2**b**). In practice, we recommend using the default 1,000 bootstrapped samples for increased robustness, and using a smaller number of bootstrap samples, e.g., when computational resources are limited.

In summary, scEPS was approximately unbiased and yielded calibrated p-values with controlled type I errors across a wide range of simulations, for both individual cell neighborhoods and groups of cell neighborhoods.

### scEPS recapitulated known and discovered novel disease-associated cell populations

We applied scEPS to analyze cognitive status (CS, defined as clinical diagnosis of dementia), and PRSs of AD, multiple sclerosis (MS), and Parkinson’s disease (PD), using the respective GWAS summary statistics data (Table 1, Supplementary Table 6) and the SEA-AD snRNA-seq brain cell atlas data [32] (Ncell=1,378,211, N_donor_=89, Supplementary Figure 29–31). We note that because the SEA-AD cohort is an AD-specific cohort, cognitive decline in this cohort primarily reflects AD pathology, justifying the use of AD GWAS genes to analyze CS. We performed analogous analyses of idiopathic pulmonary fibrosis (IPF), and PRSs of IPF, chronic obstructive pulmonary disease (COPD), and FEV1/FVC ratio (Table 1, Supplementary Table 6), using the lung cell atlas data [33] from the Translational Genomics Research Institute (TGen) (N_cell_=475,047, N_donor_=120, Supplementary Figure 32, 33). We obtained all PRSs using PGS Catalog Calculator [38, 39]. We restricted our analyses to European donors (Table 1); PRSs for non-Europeans have limited accuracy [40, 41, 42], complicating comparisons of the analysis of the disease vs. its PRS. For the analyses of PRSs, we only used the control donors to reduce consequential effects of the disease. For diseases with multiple publicly available PRS weights, we analyzed the PRS that yielded the highest aggregated *ω*^2^_*overall*_ across all cell neighborhoods (Supplementary Table 7). In all analyses, we ad*j*usted for age, sex, and neighborhood abundance as the covariates. We elected not to ad*j*ust for smoking status, a known risk factor [43, 44, 45], in the analyses of respiratory disorders, due to high missingness (12% of donors) of this variable. We elected to include genes in the MHC region, known to be involved in many complex human diseases [46, 47], in our analyses. The average size of the cell neighborhoods ranges from 430 to 1,310 for the brain cell atlas, and from 581 to 1,168 for the lung cell atlas (Supplementary Table 8).

**Table 1:**
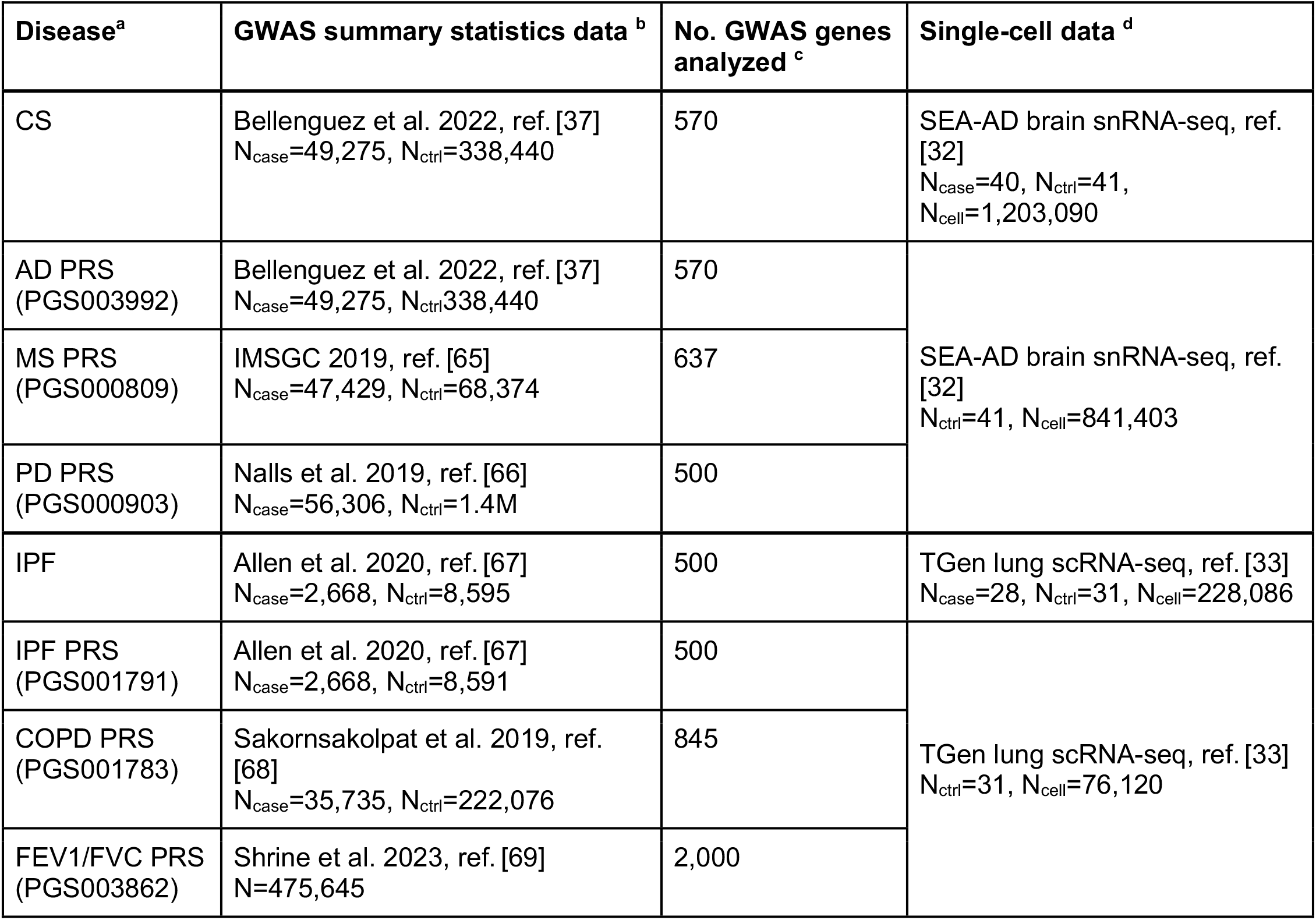
Overview of the 4 neurological and 4 respiratory disorders, GWAS summary statistics, and single-cell RNA-seq data analyzed. ^**a**^ PGS catalog numbers of the PRS weights used to calculate the PRSs are shown in parentheses. All PRS weights are obtained based on GWASs of European ancestry. ^**b**^ The GWAS sample sizes for AD and PD include the numbers of both clinically diagnosed and proxy cases. We note that FEV1/FVC is a quantitative phenotype. ^**c**^ Number of top GWAS genes with the highest MAGMA gene-level association statistics analyzed by scEPS. ^**d**^ The sample sizes of the single-cell data represent the numbers of individuals of European ancestry. Analyses of PRSs were restricted to the control donors.

#### Analysis of 4 neurological disorders

We report the results of the analyses of 4 neurological disorders in Figure 3**a, b**, Supplementary Figure 34–38, and Supplementary Table 9. At the global level, aggregating the *d* statistics across *all* cell neighborhoods (Figure 3**a**), we obtained significantly (p < 0.0125) positive aggregated *d* for 3 out of the 4 disorders (Supplementary Table 9**a, c, e, g**), with the highest aggregated *d* observed for PD PRS (*d*=1.3×10^-3^ (s.e. 1.4×10^-4^)). At the cell-type level, aggregating the *d* statistics across cell neighborhoods of each of the 139 brain cell subtypes, we identified 322 (out of 556) significant (FDR < 0.1) disease-cell-subtype associations with aggregated *d* > 0 (Figure 3**b**, Supplementary Table 9**b, d, f, h**). At individual cell neighborhood level, we identified 1,696 significant associations (*d* > 0, FDR < 0.1) only for CS, with neighborhoods in oligodendrocyte, OPC, and microglia-PVM having the highest *d*; analyses of PRSs didn’t yield any significant result, except for one L2/3 IT neighborhood implicated for PD PRS.

**Figure 3:**
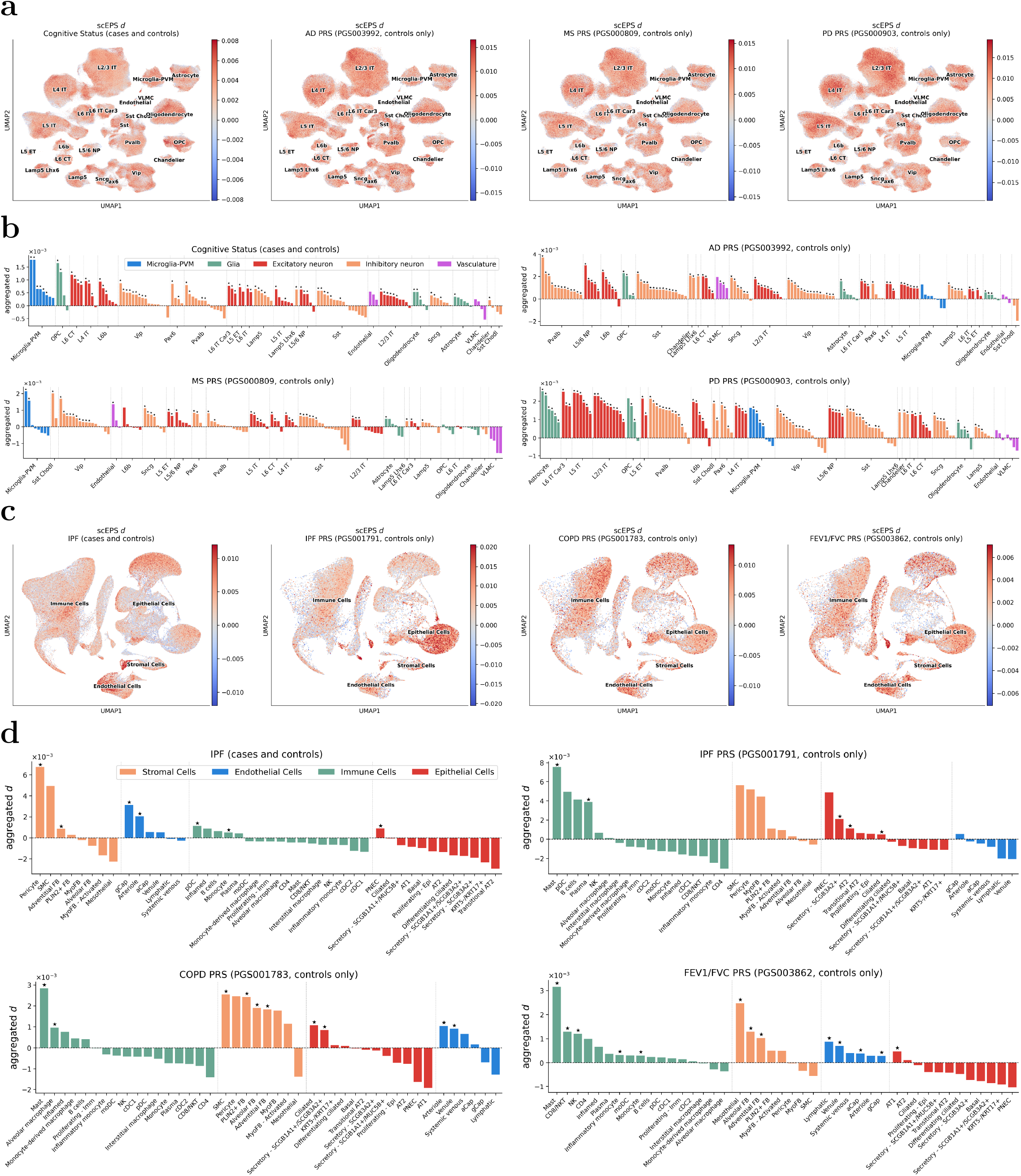
Results for the analysis of 4 neurological and 4 respiratory disorders using scEPS. (**a**) UMAP plots showing the scEPS *d* statistics at individual cell neighborhood level for (left to right) CS, PRSs of AD, MS, and PD. (**b**) We report the aggregated *d* statistics at cell subtype level for CS (top left), PRSs of AD (top right), MS (bottom left), and PD (bottom right). (**c**) UMAP plots showing the scEPS *d* statistics at individual cell neighborhood level for (left to right) IPF, PRSs of IPF, COPD, and FEV1/FVC. (**d**) We report the aggregated *d* statistics at cell subtype level for IPF (top left), PRSs of IPF (top right), COPD (bottom left), and FEV1/FVC (bottom right). Each cell in the UMAP plots represents a cell neighborhood. We sort the cell subtypes based on the aggregated *d* statistics within each cell type; we sort the cell types based on the highest aggregated *d* statistic of their constituent subtypes. Stars (⋆) denote number of cell neighborhoods > 100, aggregated *d* > 0, aggregated *ω*^2^_*gwas*_ > 0, and FDR < 0.1. Numerical results are reported in Supplementary Table 9, 10.

We recapitulated several well-studied disease-cell-type associations, including the associations linking microglia and astrocytes subtypes with CS, and PRSs of MS and PD [48, 49, 50, 51, 52, 53] (Figure 3**b**, Supplementary Table 9**b, f, h**). We also identified less well-studied but biologically plausible associations linking OPC with CS, and PRSs of AD and PD [54, 55, 56, 57] (Figure 3**b**, Supplementary Table 9**b, d, h**).

Interestingly, we observed limited correlation (R=0.071 (s.e. 0.11)) between the aggregated *d* for CS vs. AD PRS across the cell subtypes (Supplementary Figure 38**a**), primarily due to the negative (although non-significant) correlation (R=-0.21 (s.e. 0.26)) for non-neuronal cell subtypes (Supplementary Figure 38**g**). Notably, microglia-PVM subtypes had the highest and significant aggregated *d* in the analysis of CS, but were not significant in the analysis of AD PRS (Figure 3**b**, Supplementary Table 9**b, d**); the aggregated *ω*^2^_*overall*_ for microglia-PVM subtypes, despite being significantly positive (FDR < 0.1), were also comparably lower than those for neuronal cell subtypes in the analysis of AD PRS (Supplementary Figure 35**h**, Supplementary Table 9**b, d**). Interestingly, the aggregated *d* for VLMC subtypes were non-significant in the analysis of CS but were significantly positive in the analysis of AD PRS (Figure 3**b**, Supplementary Table 9**b, d**). Nevertheless, we also observed that subtypes of OPC and L6b had highly positive and significant aggregated *d* in both the analysis of CS and the analysis of AD PRS (Figure 3**b**, Supplementary Table 9**b, d**). Taken together, these results suggest that the disease mechanisms were largely different for active vs. preclinical stages of AD.

#### Analysis of 4 respiratory disorders

The results of the analyses of 4 respiratory disorders are reported in Figure 3**c, d**, Supplementary Figure 39–43, and Supplementary Table 10. At the global level, aggregating the *d* statistics across *all* cell neighborhoods (Figure 3**c**), we observed a lack of significantly (p < 0.0125) positive aggregated *d* (Figure 3**b**, Supplementary Table 10**a, c, e, g**). At the cell-type level, aggregating the *d* statistics across cell neighborhoods of each of the 43 lung cell subtypes, we identified 35 (out of 172) significant (FDR < 0.1) disease-cell-subtype associations with aggregated *d* > 0 (Figure 3**d**, Supplementary Table 10**b, d, f, h**). At individual cell neighborhood level, we identified 861 significant associations (*d* > 0, FDR < 0.1) only for IPF, with neighborhoods in lung arteriole cells, inflammatory monocytes, and smooth muscle cells having the highest *d*; analyses of PRSs didn’t yield any significant results at individual cell neighborhood resolution.

Our results highlighted the association linking pericytes and AT2 cells to both IPF and IPF PRS, and the association linking mast cells to the PRSs of IPF and COPD (Figure 3**b**, Supplementary Table 10**b, d, f**). We note that the roles of pericytes, AT2, and mast cells are well documented in the pathogenesis of IPF [58, 59, 60, 59]. However, the role of mast cells in COPD is less well studied [61].

We observed moderate but significant correlation (R=0.44 (s.e. 0.15)) between aggregated *d* for IPF vs. IPF PRS across the lung cell subtypes (Figure 3**d**, Supplementary Figure 43**a**, Supplementary Table 10**b, d**). Further, this correlation was generally consistent across immune (R=0.42 (s.e. 0.23)) vs. non-immune (R=0.56 (s.e. 0.21)) cell subtypes (Supplementary Figure 43**d, g**). Despite these similarities, we also observed that analysis of IPF PRS yielded much higher aggregated *d* for immune and epithelial cell subtypes than in the analysis of IPF (Figure 3**d**). Taken together, these results suggest that the active and preclinical stages of IPF involved shared disease mechanisms, while the preclinical stage of IPF was more heavily impacted by immune and epithelial cell biology.

#### Sensitivity analysis

We performed four secondary analyses. First, we assessed the robustness of our approach to selecting control genes. Since the control genes were selected at random, we expect limited contributions of these genes to the diseases. Indeed, across all diseases, the aggregated *ω*^2^_*ctrl*_ statistics were magnitudes smaller than the aggregated *d* and *ω*^2^_*gwas*_, with few reaching statistical significance at FDR < 0.1. (see subfigures **e**–**g** of Supplementary Figure 34–37, 39–42). We note that significantly positive *ω*^2^_*ctrl*_ is still possible, e.g., for diseases regulated by many genes. We also compared the average variance of the expression of GWAS vs. control genes in each cell neighborhood, and observed strong consistency, with R ranging from 0.84 to 0.99 (Supplementary Figure 44). These results suggest that our approach to selecting control genes also resulted in similar expression variance for GWAS and control genes.

Second, we investigated whether scEPS’ neighborhood-based approach was able to detect more disease-associated cell populations than a pseudo-bulk approach. We applied scEPS to analyze the 4 neurological and 4 respiratory disorders, using the pseudo-bulk gene expression of the 139 brain and 43 lung cell subtypes, respectively, effectively treating each cell subtype as a single cell neighborhood (Supplementary Figure 45–52, Supplementary Table 11, 12). Compared with the pseudo-bulk-based analysis, scEPS’ neighborhood-based analysis detected substantially more cell subtypes significantly (FDR < 0.1) associated with the diseases (Supplementary Table 13), as the neighborhood-based approach was more powerful at capturing subtle associations among disease, gene, and granular cell populations.

Third, we assessed the impact of analyzing a disease/phenotype with a mismatched disease GWAS. Specifically, we applied scEPS to analyze the 4 neurological and 4 respiratory disorders using COPD and AD GWAS, respectively (Supplementary Figure 53–60, Supplementary Table 14, 15). As expected, the aggregated *d* across all cell neighborhoods were significantly lower (p < 0.0125) in the analyses using mismatched GWASs, compared with using matched GWASs, for all diseases except IPF PRS (Supplementary Table 16). Interestingly, even in the mismatched analyses, certain cell subtypes still had significantly (FDR < 0.1) positive aggregated *d*. For example, Microglia-PVM subtypes had significantly (FDR < 0.1) positive aggregated *d*, in the analyses of CS using both AD and COPD GWASs (Figure 3**b**, Supplementary Figure 53**e**, Supplementary Table 9**b**, 14**a**). We note that this is likely driven by shared immune-related biology across AD [62] and COPD [63]. Indeed, gene set enrichment analysis of top AD and COPD GWAS genes analyzed by scEPS both highlighted immune-related pathways (Supplementary Table 17).

Fourth, we investigated whether cell types identified by scEPS could be replicated in an external data. We applied scEPS to analyze CS (dementia vs. normal) using the ROSMAP [64] brain cell atlas data (N_donor_=397, Supplementary Figure 61, also see “Supplementary notes” for more details). Here, we defined dementia as cognitive decline caused specifically by AD with Mini-Mental State Examination (MMSE) score less than 25. Reassuringly, we observed that similar brain cell subtypes, including neuron and microglia subtypes, were implicated for CS using the ROSMAP data (Supplementary Figure 62**e**, Supplementary Table 18). We also note that the aggregated *ω*^2^_*overall*_ statistics were lower using the ROSMAP data (*ω*^2^_*overall*_=1.30×10^-5^ (s.e. 1.27×10^-6^)) vs. the SEA-AD data (*ω*^2^_*overall*_=1.78×10^-5^ (s.e. 1.81×10^-6^)). This is likely because the number of cells from each donor is smaller for ROSMAP (~1095 cells per donor) than SEA-AD (14,852 cells per donor), leading to increased statistical noise in neighborhood-level gene expression and decreased *ω*^2^_*overall*_ statistics (consistent with simulation, Supplementary Figure 2**d**, Supplementary Figure 26**d**, Supplementary Table 5**c**).

In summary, our results revealed both established and potentially novel disease-cell-type associations for 4 neurological and 4 respiratory disorders and highlighted the discrepancies between analysis of case/control status and PRSs. We provide further interpretation of these findings in the “Discussion” section. We have publicly released the scEPS statistics at individual cell neighborhood resolution for the 8 disorders (see “Data availability”).

### scEPS recapitulated known and discovered novel and more granular disease-associated biological processes

We investigated the disease-associated biological processes implicated by scEPS using gene set enrichment analysis (GSEA). We performed GSEA over 2,785 Reactome [70] biological pathway gene sets using GSEApy [71], based on the rank correlations (Spearman’s *ρ*) between scEPS *d* statistics and the expression of each protein coding gene (27,033 in total), across cells where the gene is expressed. We performed both global and cell-type level GSEA, based on the correlations calculated across all cells and across cells of individual cell types, respectively. The global GSEA highlights disease-associated biological processes in the most disease-associated cell-types, while the cell-type level GSEA captures more granular associations within each cell type.

We report the results in Figure 4–6, and Supplementary Table 19, 20. scEPS recapitulated known disease-associated biology for the 4 neurological disorders via global GSEA. For CS, scEPS highlighted several T cell related biological processes, including “Co-inhibition by PD-1”, “Phosphorylation of CD3 and TCR *ζ*-chains”, and “Regulation of T cell activation by CD28 family” (Figure 6**a**, Supplementary Table 19**a**), concordant with recent studies highlighting the role of T cells in AD [72, 73, 74]. We note that these results were consistent with the high lymphocytes *d* statistics for CS (aggregated *d*=0.0017 (s.e. 0.00073, p=0.014), Figure 7**b**). For AD PRS, scEPS implicated several RNA metabolism biological processes, e.g., “Viral mRNA Translation”, “Selenocysteine synthesis”, and “Nonsense Mediated Decay (NMD)” (Figure 6**b**, Supplementary Table 19**b**), corroborating studies implicating the role of RNA metabolism in AD [75, 76]. Interestingly, immune pathway gene sets were enriched for genes negatively correlated with scEPS *d* statistics for AD PRS (Figure 6**b**, Supplementary Table 19**b**), contrary to the results for CS (Figure 6**a**, Supplementary Table 19**a**). The differences in the pathways identified for CS and AD PRS suggest that different biological processes were involved in active/symptomatic state vs. pre-disease/asymptomatic state of the disease. For MS PRS, scEPS implicated similar T cell biology as implicated for CS, which is well documented for MS (Figure 6**c**, Supplementary Table 19**c**) [77, 78, 79]. Finally, for PD PRS, scEPS highlighted several neuronal pathways, including “Trafficking of AMPA receptors” (Figure 6**d**, Supplementary Table 19**d**), which aligned with a previous finding that inhibiting AMPA receptor expression had antiparkinsonian effects in rats and monkeys [80].

**Figure 4:**
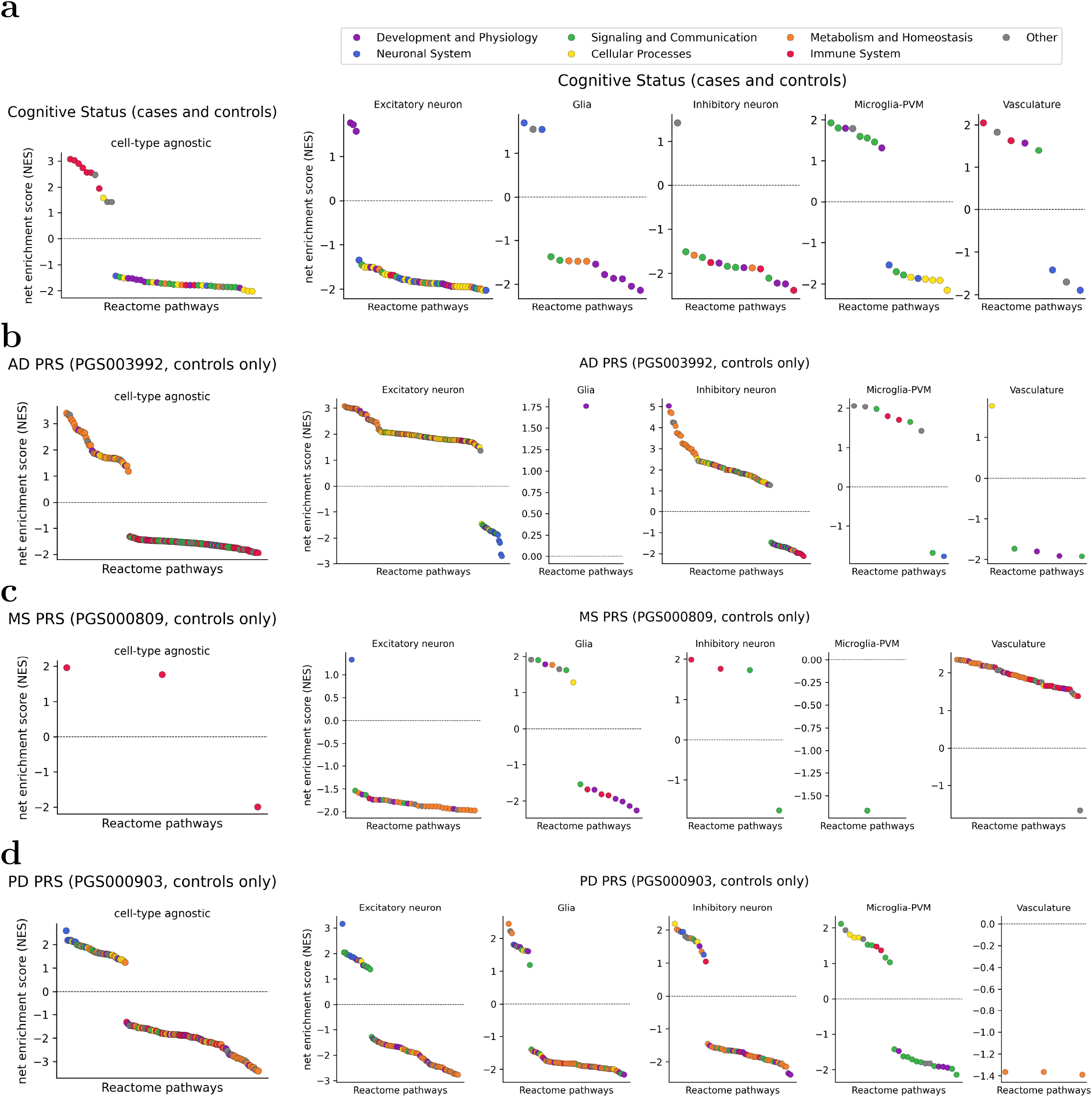
Biological processes implicated in the GSEA of genes correlated with scEPS *d* statistics across cells for the 4 neurological disorders. The leftmost plots show the Reactome pathways implicated via global GSEA (FDR < 0.05) for CS (**a**), AD PRS (**b**), MS PRS (**c**), and PD PRS (**d**); the rightmost 5 plots show analogous results obtained via cell-type level GSEA (FDR < 0.05) for excitatory neuron, glia, inhibitory neuron, microglia-PVM, and vasculature, respectively. The FDRs in the cell-type level GSEA were calculated across all (cell type, pathway) pairs. Reactome pathways are grouped into 7 broad categories based on their biological functions and are represented by different colors. Numerical results are reported in Supplementary Table 19.

**Figure 5:**
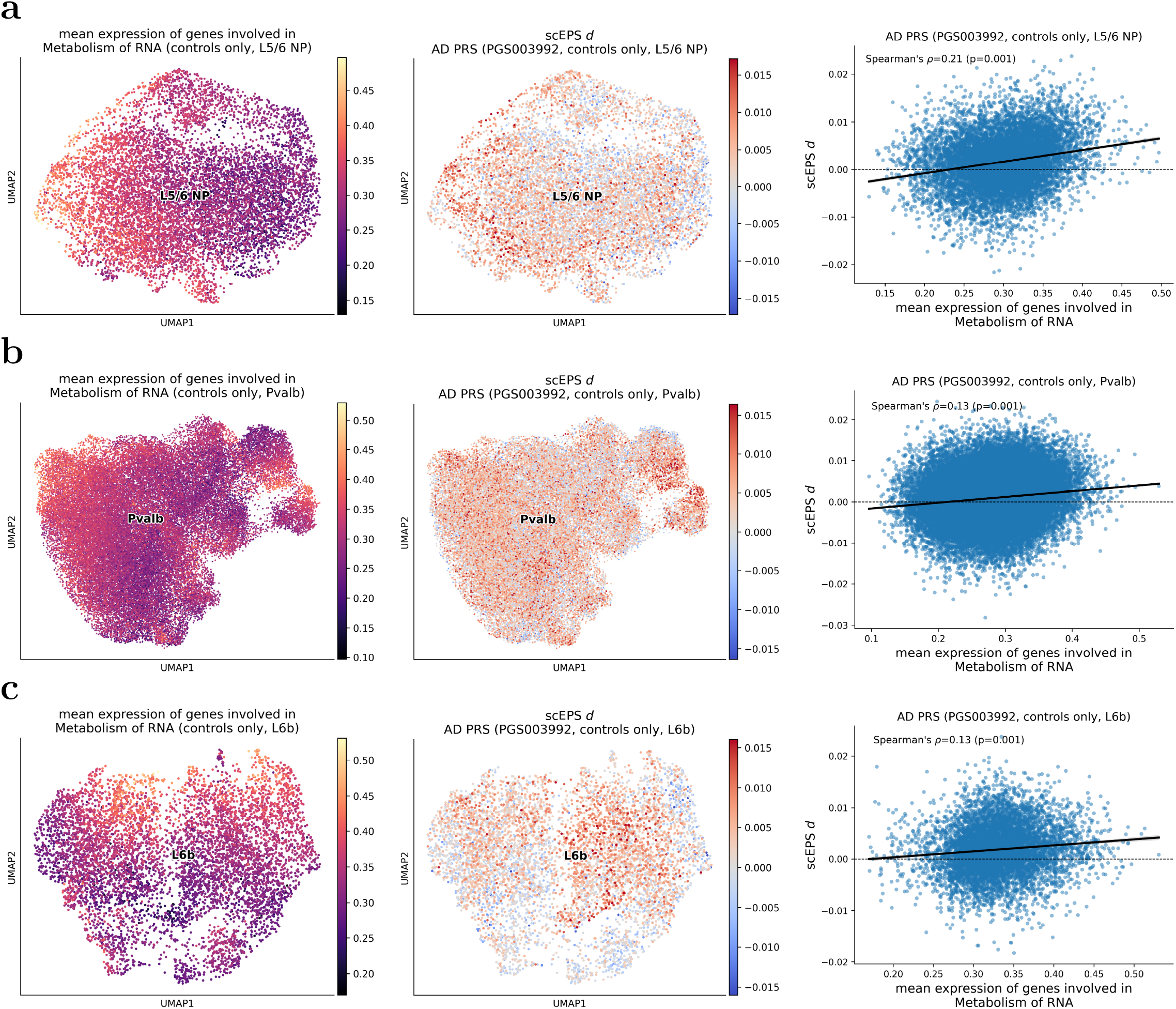
AD PRS scEPS *d* statistics vs. average expression of genes involved in the Reactome “Metabolism of RNA” pathways in neuronal cells. The leftmost and middle UMAP plots show the average expression of genes involved in the “Metabolism of RNA” pathway and AD PRS scEPS *d* statistics, respectively, in L5/6 NP (**a**), Pvalb (**b**), and L6b (**c**) neuronal cell; side-by-side comparisons of the average expression vs. scEPS *d* statistics across cells are shown in the rightmost scatter plots. P-values for the Spearman’s *ρ* in the scatter plots were obtained based on 1,000 permutations. Solid lines in the scatter plots represent the regression line, with shaded regions representing 95% confidence intervals.

**Figure 6:**
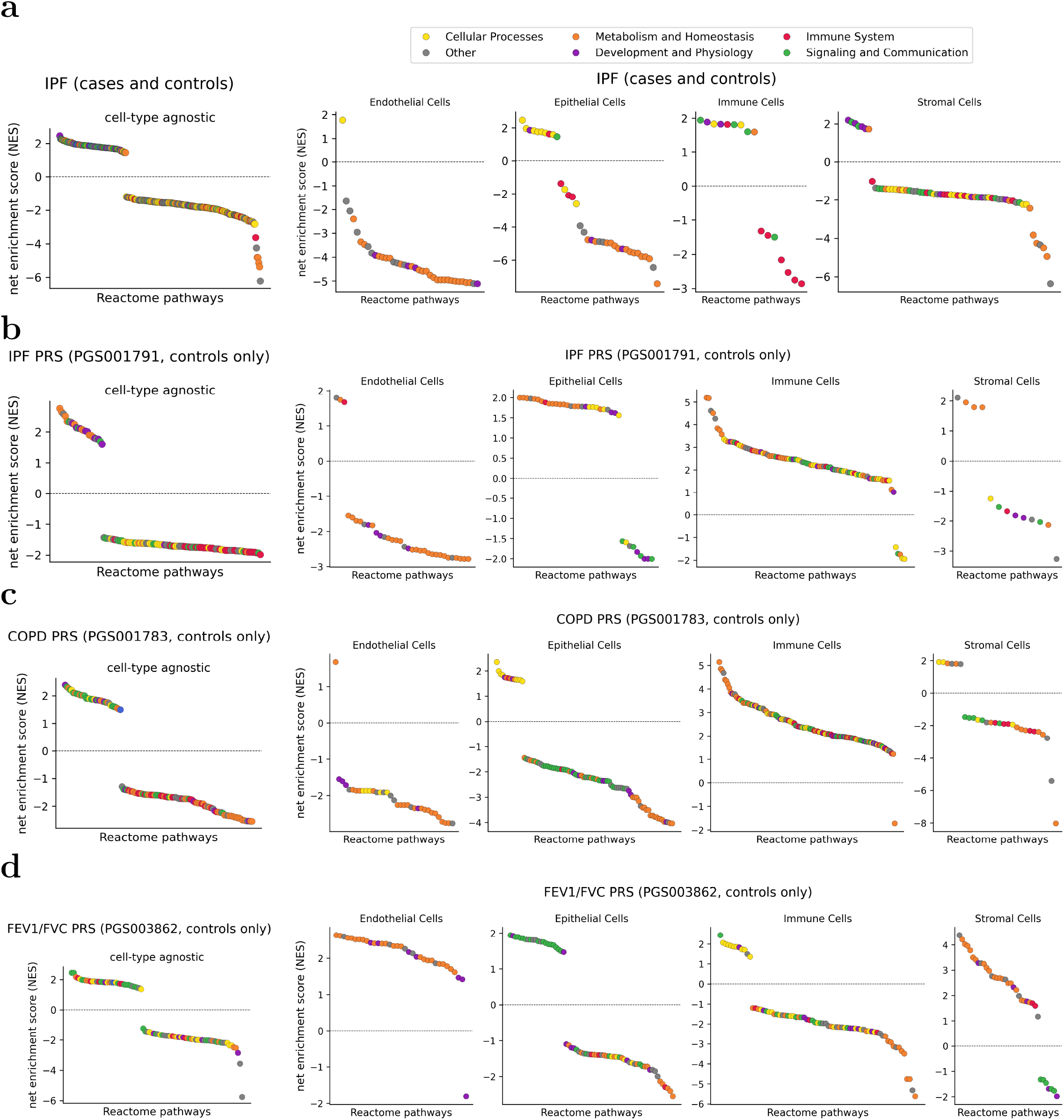
Biological processes implicated in the GSEA of genes correlated with scEPS *d* statistics across cells for the 4 respiratory disorders. The leftmost plots show the Reactome pathways implicated via global GSEA (FDR < 0.05) for IPF (**a**), IPF PRS (**b**), COPD PRS (**c**), and FEV1/FVC PRS (**d**); the rightmost 4 plots show analogous results obtained via cell-type level GSEA (FDR < 0.05) for endothelial cells, epithelial cells, immune cells, and stromal cells, respectively. The FDRs in the cell-type level GSEA were calculated across all (cell type, pathway) pairs. Reactome pathways are grouped into 6 broad categories based on their biological functions and are represented by different colors. Numerical results are reported in Supplementary Table 20.

**Figure 7:**
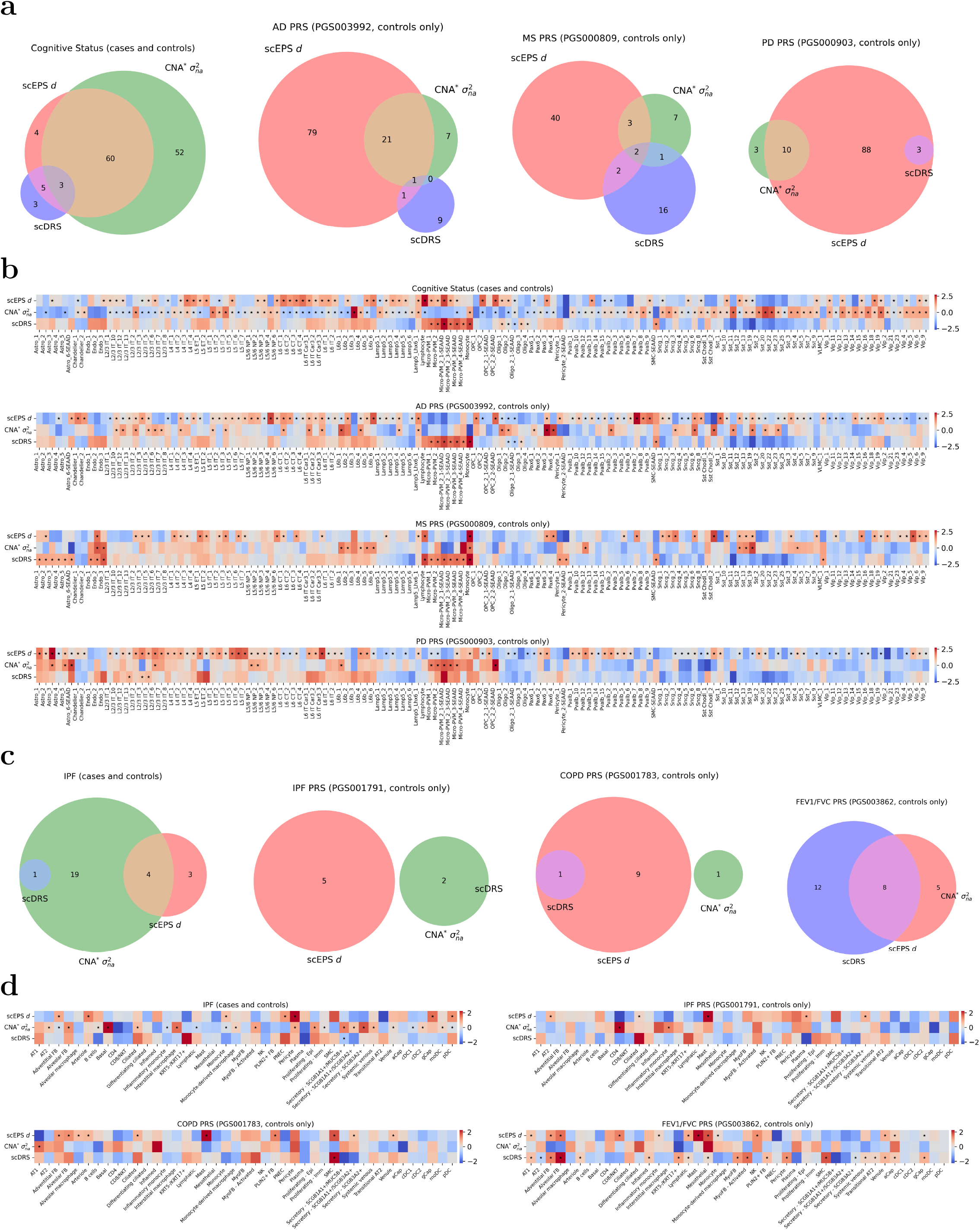
Overlap of disease-associated cell types identified by scEPS, CNA*, and scDRS. (**a**) We report the overlap of the significant associations (FDR < 0.1) identified by scEPS (aggregated *d* > 0), CNA* (*σ*^2^_*na*_ > 0), and scDRS for (left to right) cognitive status, PRSs of AD, MS, and PD, across the 139 brain cell subtypes; we report in (**c**) results for (left to right) IPF, PRSs of IPF, COPD, and FEV1/FVC, across the 43 lung cell subtypes. Note: scDRS did not report any significant association for PD. (**b**) Heat maps showing the inverse normal transformed aggregated scEPS *d* statistics, mean CNA* *σ*^2^_*na*_, and mean scDRS raw statistics for (top to bottom) cognitive status, PRSs of AD, MS, and PD, across the 139 brain cell subtypes; we report in (**d**) results for IPF (top left), PRSs of IPF (top right), COPD (bottom left), and FEV1/FVC (bottom right), across the 43 lung cell subtypes. Significant associations are denoted by stars (?) in the heat maps. Numerical results are reported in Supplementary Table 9, 10, 21–24.

scEPS provided more granular mechanistic disease insights for the 4 neurological disorders via cell-type level GSEA. For CS, scEPS implicated “Neurexins and neuroligins” in glial cells and extracellular matrix (ECM) related gene sets (e.g., “Laminin interactions”) in microglia-PVM (Figure 6**a**, Supplementary Table 19**c**). Both of these biological processes play critical roles in the formation and function of synapse and are linked to neurodegenerative disorders [81, 82, 83, 84, 85, 86]. For AD PRS, scEPS implicated multiple RNA metabolism related pathways in neurons, similar to the results from the global GSEA (Figure 6**b**, Supplementary Table 19**d**). Motivated by these results, we calculated the average expression of the genes in the Reactome “Metabolism of RNA” pathway for each L5/6 NP, Pvalb, and L6b cell, and observed significant correlation (permutation p=0.001) with scEPS *d* statistics for AD PRS (Figure 5). Notably, scEPS also implicated “Signaling by CSF3 (G-CSF)” for AD PRS in microglia-PVM, which aligned with studies demonstrating the protective effect of G-CSF on cognition in *in vitro* and *in vivo* models (Figure 6**b**, Supplementary Table 19**d**) [87, 88]. For MS PRS, scEPS highlighted multiple immune related pathways in vasculature cells, including “NOD1/2 Signaling Pathway” (Figure 6**c**, Supplementary Table 19**e**). We note that although the involvement of the immune system in MS is known, the specific roles of NOD1/2 signaling remains under-studied [89]. Finally, for PD PRS, scEPS highlighted several neuronal system pathways in neurons (Figure 6**d**, Supplementary Table 19**f**), consistent with the global analysis. Notably, scEPS also implicated gene sets involved in TGF-*β* signaling in microglia-PVM; TGF-*β* is involved in neuronal functions and could be a potential therapeutic target for PD [90, 91, 92].

scEPS also recapitulated known disease-associated biology for the 4 respiratory disorders/phenotypes via global GSEA. Notably, scEPS implicated ECM-related biological processes (e.g., “Laminin interactions” for IPF and “Collagen degradation” for COPD PRS) for all 4 respiratory disorders/phenotypes (Figure 6, Supplementary Table 20**a**–**d**). Indeed, ECM is essential for the structural integrity of the lung; dysregulations of ECM have been linked to lung function and respiratory disorders [93, 94, 95, 96]. scEPS also implicated biological processes involved in muscle contraction (e.g., “Smooth Muscle Contraction”) for IPF, IPF PRS, and COPD (Figure 6**a, b, c**, Supplementary Table 20**a, b, c**). We note that the link between respiratory muscles and COPD is well established by previous studies [97, 98, 99]; however, the link with IPF is less investigated, but has potential to yield novel therapeutic insights [100]. The shared biological pathways identified for IPF and IPF PRS also suggest that similar biological processes are involved in the active/symptomatic state and pre-disease/asymptomatic state of the disease. Interestingly, for FEV1/FVC PRS, scEPS prioritized several immune related pathways, e.g., “TNFs bind their physiological receptors”, “DAP12 signaling”, etc. (Figure 6**d**, Supplementary Table 20**d**), which were generally enriched for genes negatively correlated with scEPS *d* statistics for other respiratory disorders. Taken together, these results suggest that lung function phenotype (e.g., FEV1/FVC) and respiratory disorders (e.g., IPF, COPD) involve both shared and distinct biological processes.

scEPS also identified more granular disease-associated biological processes for the 4 respiratory disorders via cell-type level GSEA. For IPF, scEPS highlighted cellular processes in cilia cells, including “Intraflagellar transport” and “Cilium Assembly” in epithelial cells (Figure 6**a**, Supplementary Table 20**e**), consistent with recent findings that cilium biology was elevated in IPF [101, 102]. Notably, the biological process, “Immunoregulatory interactions between a Lymphoid and a non-Lymphoid cell”, was implicated in immune cells for both IPF and IPF PRS, and in epithelial cells for IPF (Figure 6**a, b**, Supplementary Table 20**e, f**), suggesting that IPF disease biology likely involved interactions between immune and epithelial cells. We note that although immune cells are known to be involved in IPF [103], their interaction with epithelium remains under-investigated and has the potential to yield novel therapeutic insights for IPF [104]. Interestingly, for both IPF PRS and COPD PRS, scEPS implicated multiple biological processes involved in protein metabolism (e.g., “Peptide chain elongation”, “Eukaryotic Translation Elongation”, etc.) in immune cells (Figure 6**b, c**, Supplementary Table 20**f, g**). Similar protein metabolism pathways were also implicated for FEV1/FVC PRS in endothelial and stromal cells (Figure 6**d**, Supplementary Table 20**h**). We note that although protein metabolism is known to be involved in respiratory disorders [105, 106, 107, 108], its specific link with respiratory disorders, for example, in immune cells, remains unclear. However, protein metabolism plays critical roles in immune cell function [109, 110, 111], and could contribute to respiratory disorders.

In summary, scEPS implicated both well-known and discovered novel and more fine-grained biological and cellular processes for the 4 neurological and 4 respiratory disorders/phenotypes. The differences in the implicated biological processes for the CS vs. AD PRS also suggest that distinct biology could be involved in active/symptomatic vs. preclinical/asymptomatic states of the disease.

### scEPS identified both shared and distinct disease-associated cell populations compared with other methods

We compared scEPS with CNA*, our modified version of CNA [34]; CNA* estimates *σ*^2^_*na*_, variance in disease phenotypes across patients explained by variations in neighborhood cell abundance (“Methods”), while CNA quantifies the correlation between neighborhood abundance and disease phenotypes. We note that *σ*^2^_*na*_ is more comparable to the scEPS statistics, also defined based on variance explained, than the CNA neighborhood correlations. In all CNA* analyses, we ad*j*usted for age and sex as covariates, consistent with the scEPS analyses. We also compared scEPS with scDRS [24], a method for identifying cells overexpressing GWAS genes. For fair comparisons, we applied both CNA* and scDRS to analyze the same sets of cells as analyzed by scEPS (Table 1). Specifically, for comparisons involving analyses of PRSs, we applied both methods to cells from control donors only. Additionally, for scDRS, we analyzed the same disease GWAS genes as analyzed by scEPS (Table 1), instead of the default top 1,000 GWAS genes. We also increased the number of control gene sets from the default 1,000 to 3,000 in the analyses using SEA-AD brain cell atlas data, to ensure sufficient statistical power in testing aggregated scDRS scores across the 139 brain cell subtypes.

We report the results in Figure 7, 8, Supplementary Figure 63–65, and Supplementary Table 21–24. Across all the 8 disorders, we identified 1.77× (s.e. 1.21) and 5.13× (s.e. 3.08) more disease-associated cell subtypes using the scEPS (aggregated *d* > 0, FDR < 0.1) than CNA* (*σ*^2^_*na*_ > 0, FDR < 0.1) and scDRS (FDR < 0.1), respectively (Figure 7**a, c**). The relative increase of scEPS was higher for the 4 neurological disorders, with scEPS identifying 1.84× (s.e. 1.52) and 6.83× (s.e. 3.83) more associations than CNA* and scDRS, respectively. For the 4 respiratory disorders, scEPS identified 1.30× (s.e. 7.82) and 1.59× (s.e. 5.66) more associations than CNA* and scDRS, respectively. We also observed a higher relative increase for scEPS vs. CNA* in the analyses of PRSs, with scEPS identifying 4.66× (s.e. 2.61) more associations than CNA* across analyses of PRSs (Figure 7**a, c**).

**Figure 8:**
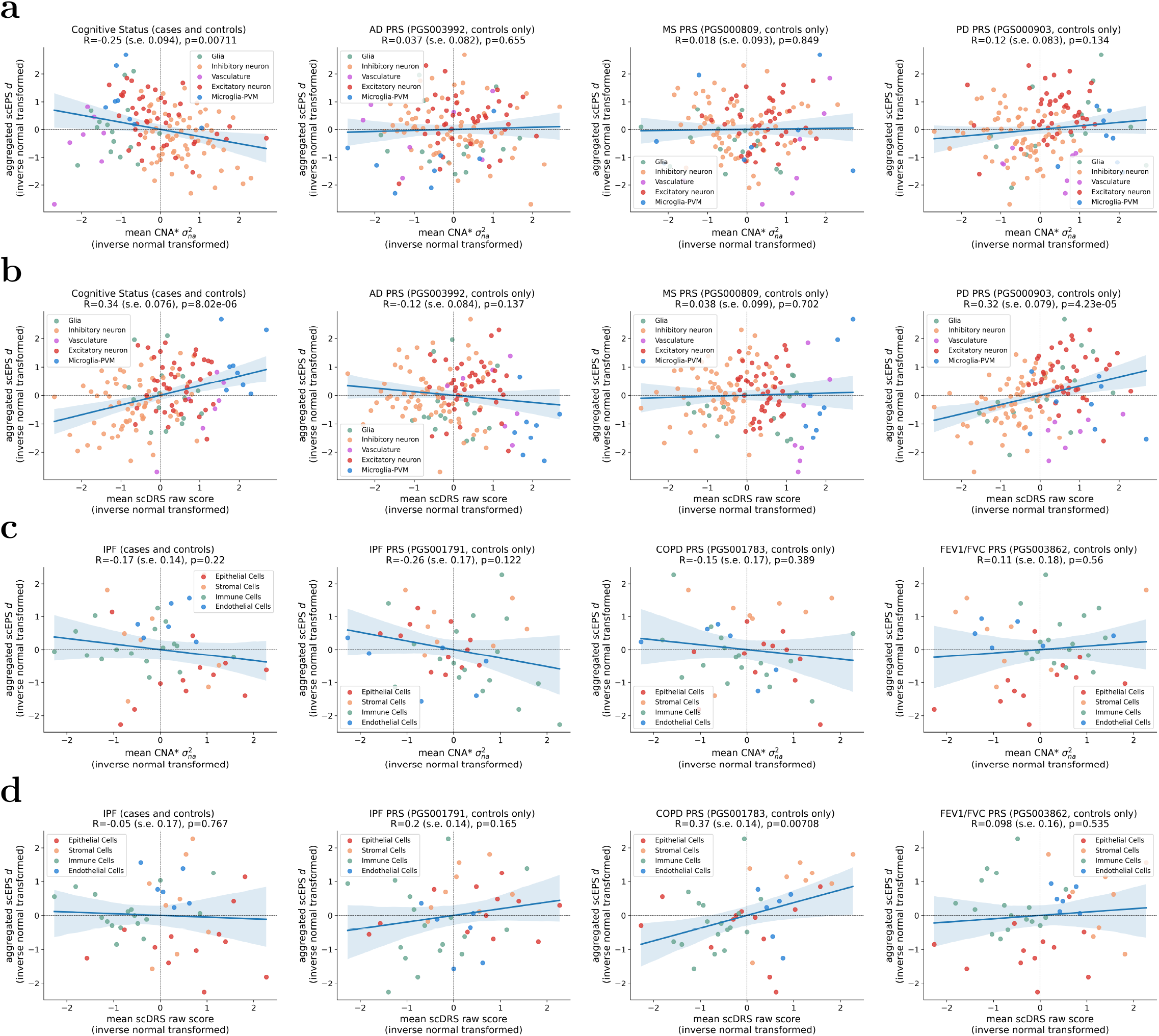
Comparisons of aggregated scEPS *d* statistics vs. CNA* σ^*2*^_*na*_ and scDRS raw scores. (**a, b**) Scatter plots showing the inverse normal transformed aggregated *d* statistics vs. CNA* *σ*^2^_*na*_ and scDRS raw scores, respectively, across 139 brain cell subtypes, for (left to right) CS, and PRSs of AD, MS, and PD. (**c, d**) Similar scatter plots across 43 lung cell subtypes, for (left to right) IPF, and PRSs of IPF, COPD, and FEV1/FVC. Standard errors of the correlations were obtained by statistical bootstrapping across the cell subtypes. Shaded regions of the regression line represent 95% confidence intervals. Numerical results are reported in Supplementary Table 21–24.

We observed both shared and distinct disease-associated cell types identified by the 3 methods (Figure 7). For example, all 3 methods implicated subtypes of microglia for CS, but only scEPS implicated a subtype of astrocytes (Figure 7**b**). We note that several recent studies have revealed the involvement of astrocytes for AD [112], and highlighted its potential for therapeutics and biomarker development [113, 114]. Interestingly, neither scEPS nor CNA* identified any microglia subtype for AD PRS, but scDRS implicated 7 (out of 8) microglia subtypes (Figure 7**b**). However, all 3 methods implicated a subtype of oligodendrocyte for AD PRS (Figure 7**b**). For IPF, both scEPS and CNA* highlighted myeloid subtypes, including monocyte and pDC, whereas scDRS only identified *SCGB1A1*^+^/*MUC5B*^+^ secretory cells (also identified by CNA* but not by scEPS) (Figure 7**d**). However, only scEPS identified endothelial cell subtypes, including gCap and arteriole cells (Figure 7**d**). We note that a recent work highlighted both the causal and consequential roles of endothelial cells in IPF [100]. For IPF PRS, scEPS and CNA* identified disjoint sets of immune cell subtypes (e.g., mast cells by scEPS, inflammatory monocyte by CNA*), but only scEPS identified epithelial cell subtypes (e.g., AT2 and ciliated) (Figure 7**d**). We note that the role of AT2 cells in IPF is well documented [60, 100, 115].

We also observed both concordance and discordance in how scEPS prioritized cell types for each disease vs. other methods (Figure 8). Meta-analyzing across all 8 disorders using inverse-variance weighted mean, we observed a weak negative correlation between inverse normal transformed aggregated scEPS *d* statistics and CNA* *σ*^2^_*na*_ (R=-0.034, s.e. 0.11), and a weak positive correlation with raw scDRS scores (R=0.17, s.e. 0.11) (Figure 8). The strongest negative correlation between scEPS and CNA* was observed for CS (R=-0.25 (s.e. 0.094)), with scEPS prioritizing microglia-PVM subtypes, and CNA* prioritizing neuronal cell subtypes (Figure 8**b**). We note, however, that both methods implicated subtypes of microglia-PVM and neurons for CS (Figure 7**b**), despite the discrepancy in how each method rank-ordered the cell subtypes. The strongest positive correlation between scEPS and scDRS was observed for COPD PRS (R=0.37 (s.e. 0.14)), with both methods prioritizing stromal cell subtypes (Figure 8**d**); specifically, both methods implicated smooth muscle cells (Figure 7**d**).

We also compared the disease-associated cell subtypes identified using the aggregated scEPS *ω*^2^_*overall*_ statistics vs. CNA* and scDRS (Supplementary Figure 63, 64). We observed substantial increase in the overlap of disease-associated cell types identified using aggregated scEPS *ω*^2^_*overall*_ and CNA* *σ*^2^_*na*_, relative to using scEPS *d* statistics, especially for CS, PD PRS, and IPF (Supplementary Figure 63**a, c**, Figure 7**a, c**). Interestingly, we also observed weak positive correlation between aggregated scEPS *ω*^2^_*overall*_ and CNA* *σ*^2^_*na*_ (R=0.12 (s.e. 0.10), meta-analyzing across 8 disorders) (Supplementary Figure 64**a, c**). On the contrary, we did not observe a similar increase in the overlap of cell types identified using aggregated scEPS *ω*^2^_*overall*_ and scDRS (Supplementary Figure 63**a, c**, Figure 7**a, c**), and observed a slight decrease in the correlation with scDRS raw scores (R=0.12 (s.e. 0.10)) (Supplementary Figure 64**b, d**).

For completeness, we also compared CNA* and scDRS with respect to prioritization of disease-associated cell types (Supplementary Figure 65). We observed a weak negative correlation (R=-0.05 (s.e. 0.11)) between the aggregated CNA* *σ*^2^_*na*_ and scDRS raw scores across the 8 disorders. The strongest negative correlation was observed for FEV1/FVC PRS (R=-0.54 (s.e. 0.11)), with CNA* prioritizing immune cell subtypes, while scDRS prioritizing stromal cell subtypes (Supplementary Figure 65**b**). And the strongest positive correlation was observed for PD PRS (R=0.35 (s.e. 0.088)), with both methods highlighting microglia-PVM subtypes (Supplementary Figure 65**a**).

In summary, scEPS identified both shared and distinct disease-associated cell subtypes, when compared with CNA* and scDRS. Our comparisons of these methods also revealed discrepancies in how each method prioritizes disease-associated cell types. We further discuss the implications of these results in the “Discussion” section.

## Discussion

We developed scEPS, an integrative approach for identifying disease-associated cell neighborhoods, where the expression of disease GWAS genes explains more variance in the disease phenotype than randomly selected mean expression matched control genes. We showed through extensive simulations that scEPS yielded unbiased estimates of the difference in the variance in disease attributable to each GWAS vs. each control gene (*d* statistics), with calibrated test statistics. We applied scEPS to analyze clinical diagnoses and PRSs of 4 neurological and 4 respiratory disorders. We recapitulated known and discovered novel and more granular disease-associated cell populations, biological, and cellular processes. We also found that analyses of clinical diagnoses and their respective disease PRSs implicated distinct cell types and biological processes. Finally, we compared scEPS with CNA [34] and scDRS [24], existing methods for identifying disease-associated cell populations, in the analysis of real traits. We found that scEPS identified substantially more disease-associated cell types compared with both CNA and scDRS. Notably, although these methods identified shared cell types, each method also implicated a substantial number of distinct cell types.

Our study has several implications. First, the differences in the cell types and biological processes identified for clinical diagnoses vs. their respective disease PRSs suggest that the active and preclinical stages of a disease can involve distinct biological mechanisms. For instance, the preclinical stage of AD is characterized by oxidative stress in the brain [116, 117, 118], whereas the symptomatic and more severe stage of AD involves neuronal inflammation [116, 117, 118]. Indeed, our results highlighting Microglia-PVM (Figure 3**b**) and immune-related pathways (Figure 4**a**) for CS and neuronal cell types (Figure 3**b**) and metabolism-related pathways (Figure 4**b**) for AD PRS are consistent with these findings. In general, understanding the biology in different stages of a disease can provide further insights into the disease mechanisms, facilitating the development of therapeutics. Second, the inconsistencies among scEPS, CNA, and scDRS (Figure 7, Figure 8) suggest that different cell types could be implicated for a disease, depending on the metric of association. Specifically, scEPS prioritizes cell types exhibiting differential expression of GWAS genes across disease phenotypic spectrums; CNA prioritizes cell types exhibiting differential proportions across phenotypic spectrums; whereas scDRS prioritizes cell types with overexpression of GWAS genes, irrespective of the disease phenotype. In practice, the choice of the method should depend on the biological question of interest, i.e., whether differential expression, differential proportion, or specific expression provides the most relevant disease insights. In general, we recommend integrating insights from different methods to provide a more comprehensive understanding of the disease. Third, the substantial relative increase in the number of disease-associated cell types implicated by scEPS over other methods (1.77× and 5.13× for CNA* and scDRS, respectively, Figure 7) suggest that there’s greater contrast across disease phenotypes in terms of the expression of GWAS genes vs. cell abundance or specific expression of GWAS genes. In other words, the disease phenotypes, analyzed in this study, manifested more strongly as changes in gene expression instead of cell type compositions.

Next, we discuss several potential extensions of scEPS for directions of future research. First, scEPS could be extended to analyze gene sets representing biological pathways and gene expression programs, instead of GWAS genes. Second, scEPS could be extended to analyze continuous gene scores (e.g., probability of loss-of-function intolerant (pLI) score [119]), instead of binary gene sets, providing a more flexible way to rank order genes. Third, scEPS could be extended to analyze scATAC-seq data [120], yielding complementary insights into diseases than scRNA-seq data. Fourth, the scEPS method could be extended to analyze neighborhoods composed of spatial spots, defined using spatial transcriptomics data [121], identifying disease-associated spatial locations, where GWAS genes play critical roles. This extension of scEPS could provide additional insights to the recent approach [122] that integrates GWAS and spatial transcriptomics data, identifying spatial spots enriched for disease heritability.

We note several limitations of the scEPS methods. First, scEPS can only be applied to analyze single-cell data composed of multiple donors showing variations in their disease phenotypes. We recommend applying scEPS to analyze data sets with at least 20 donors, with balanced disease phenotypic distributions. Second, scEPS models disease phenotype as a linear function of gene expression, assuming independence of effect sizes across genes. scEPS could yield biased estimates of the *d* statistics, if the gene expression effect sizes are non-linear and/or not independent. Third, scEPS does not account for the correlation in the MAGMA statistics across genes, e.g., driven by linkage disequilibrium (LD) to shared GWAS loci, and could be biased by the genes near few strong GWAS loci. Selecting GWAS genes accounting for correlated MAGMA statistics may alleviate this bias. Fourth, scEPS assumes equal contribution of each gene to the disease, not distinguishing between genes with higher vs. lower genetic associations with the disease. We note that applying weights to the genes based on their strength of association with the disease could potentially increase the statistical power but would complicate downstream interpretations. Thus, we elected not to apply weights on the genes for better interpretability. Fifth, scEPS does not model interactions between genes or cells, key biological processes involved in many diseases [123]. Investigating and accounting for the impact of gene-gene and cell-cell interactions on diseases may yield further insights into diseases. Sixth, scEPS defines cell neighborhoods based on an extension of the CNA approach. However, other approaches to define cell neighborhoods are also available [28, 29, 30]; we have not systematically evaluated the advantages and disadvantages of each approach. Benchmarking and selecting the optimal approach that maximizes the signal-to-noise ratio at each neighborhood may further improve the statistical power and resolution of scEPS. Seventh, scEPS corrects for batch effects in both the k-NN graph and the gene expression based on linear models. Lingering non-linear batch effects may introduce bias in scEPS. Eighth, in the analysis of PRS in controls, scEPS does not completely resolve reverse causation, as changes in the gene expression could be a response to the preclinical stage of the disease. However, scEPS is less impacted by the effects of active disease symptoms and can be used to distinguish between different disease models (“Supplementary notes”, Supplementary Figure 66). Ad*j*usting disease progression phenotypes from PRSs and incorporating ideas from Mendelian randomization approaches [124] represent promising directions for improving scEPS’ ability to distinguish causation from association.

Finally, we note several limitations and caveats of the analyses of real traits. First, our MAGMA-based approach to select GWAS genes prioritizes genes that are proximal to SNPs with strong associations with the disease; in cases where the disease-causal gene is regulated by distal variants, this proximity-based approach could select the incorrect gene. We note several approaches to prioritize GWAS genes that leverage fine-grained SNP-to-gene linking strategies [125, 126, 127, 128, 129]. However, these approaches could bias towards genes expressed in the tissues/cell types from which the SNP-to-gene links are derived, and/or genes with specific regulatory mechanisms (e.g., enhancer vs. promoter driven). Thus, we elected to use MAGMA-based approach as the default, to mitigate biases in the selection of genes. Second, we elected to include genes in the MHC regions in our analyses, as these genes play important roles in immune response [130, 131]. Excluding these genes may significantly downplay the role of immune-related cell types and pathways in our analyses. However, we note that disentangling causal genes in the MHC region has been challenging, due to complex LD structures in the region [132]. Third, our analyses focused on genes implicated by common variants, which are well powered in GWASs; the impact of rare variants implicated genes was not investigated in our study. Applying scEPS to analyze genes implicated by rare variants, e.g., via exome-wide association studies, could provide complementary insights into disease biology [133]. Fourth, our analyses of PRSs were restricted to donors of European ancestry, due to the lack of sufficient non-European donors and accurate non-European PRSs donors [40, 134]. Analyzing data from non-European donors may provide additional insights into diseases and reduce health disparities across ancestry groups. Fifth, we only analyzed 4 neurological and 4 respiratory disorders in this study. Applying scEPS to additional single-cell disease atlases (e.g., OneK1K [135]) could yield biological insights into different disease areas. Despite these limitations, our study provides additional mechanistic insights into several neurological and respiratory disorders.

## Methods

### Intuition of the single-cell Expression exPlainability Statistics (scEPS) method

scEPS relies on the assumption that a cell population is more likely to be involved in the causal pathway of a disease, if the expression of each disease-causal gene in the cell population better explains the disease phenotype than each randomly selected gene with similar characteristics. More precisely, scEPS quantifies the variance in disease phenotypes attributable to the expression of each GWAS gene (e.g., prioritized through MAGMA [13]), which are enriched for disease causal (as opposed to response) genes [10, 11, 12, 136], vs. each randomly selected gene matched on mean expression, at the resolution of individual cell neighborhoods.

### Defining cell neighborhoods

We define a cell neighborhood anchored at a cell, *c*, as the set of cells that can be reached, with a probability greater than *θ*, in a random walk of length *s* starting at *c*, on the k-NN graph defined for a set, *C*, of cells.

Let ***A*** ∈ {0,1}^|*C*|×|*C*|^ be the adjacency matrix representing the k-NN graph for the set of cells in the single-cell data, we obtain the matrix,***Ã*** ∈ [0,1]^|*C*|×|*C*|^, of probabilities of transitioning from cell *j* to cell k in one step on the k-NN graph,

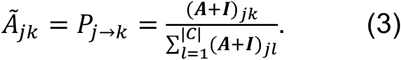

The probability that cell *i* can reach cell *j* in *s* steps is then,

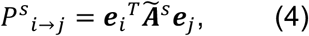

where ***e***_*i*_, ***e***_*j*_, ∈ {0,1}^|*C*|^ are vectors indexing the i-th and *j*-th cell, respectively. We define the set, *N*_*c*_^*s*^(*θ*), of neighborhood of cells anchored at cell *c* that can be reached, with probability greater than *θ*, in *s* steps of random walks on the k-NN graph starting at *c* as,

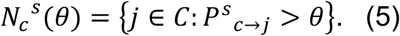

The parameters, *s* and *θ*, govern the size and resolution of the cell neighborhood. We choose these parameters in a data dependent fashion, similar to CNA [34], such that each neighborhood has a balanced contribution of cells from the donors in the single-cell dataset while attaining the smallest possible size. Detailed description of the methods to determine the parameters for *s* and *θ* from the single-cell data are outlined in the following subsections.

#### Determining the length of the random walk (*s*)

We first choose a global random walk length based on the neighborhood abundance matrix (NAM), representing the proportions of cells contributed by each individual in each cell neighborhood, using heuristics outlined in ref. [34]. Let

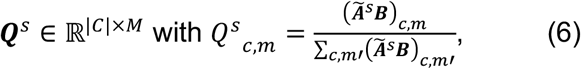

be the NAM corresponding to a random walk of length *s*, where *c* and *m* index a cell neighborhood and an individual, respectively; *s* is the length of the random walk; each column of ***B*** ∈ {0,1}^|*C*|×*M*^ indexes the cells contributed by an individual; and *M* is the total number of individuals. And let *κ*^*s*^ be the median Kurtosis across columns of ***Q***^*s*^. We increment *s* from *s* = 1, until *κ*^*s*^ is less than 8 or the difference between *κ*^*s*^ and *κ*^*s*−1^ is less than 3, as implemented in ref. [34]. This procedure of selecting *s* trades off between neighborhood granularity and balanced contribution of individuals to each neighborhood.

For sparse local regions of the graph, a larger random walk length is needed such that cells from multiple individuals are represented in the neighborhood. For these cell neighborhoods, we increment the length of the random walk, starting from the global length, until sufficient individuals are represented. This approach results in neighborhood specific random walk lengths, adapted to the local sparsity pattern of the k-NN graph.

#### Choosing the transition probability threshold (*θ*)

Given a pre-specified number of random walks, *s*, we next select the set of cells that belong to each cell neighborhood, by choosing *θ* that minimizes the difference between the contribution of cells to the neighborhood anchored at cell *c* that is entailed by *s* and those realized from a specific value of *θ*. More precisely, let ***Q***^*s*^_*c*_ ∈ ℝ^*M*^ be the vector of pre-specified proportions of cells in the neighborhood indexed by *c* from each donor, and let 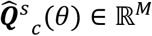 with 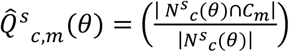, where *C*_*m*_ is the set of cells from individual *m*, be the realized counterpart, we aim to find *θ* that minimizes 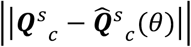 such that there are at least *M*_*min*_ (10 by default) individuals with distinct outcomes having at least *N*_*min*_ (5 by default) cells each in the neighborhood.

We perform a grid search of 1,000 values of *θ*, ranging from 0.0 to 0.01, to find the optimal *θ, θ*^*\**^. If multiple values of *θ* minimizes 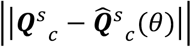 while satisfying the constraints, we choose the value of *θ* that results in the smallest neighborhood size. The set of cells in the neighborhood anchored at cell *c* is then *N*_*c*_^*s*^(*θ*^*\**^) =[*j* ∈ *C*: *P*^*s*^_*c*→*j*_ > *θ*^*\**^].

### Determining the set of disease GWAS genes

We apply MAGMA [13] to the GWAS summary statistics data of a disease, using default parameters, to obtain gene-level association statistics (i.e., Z-scores and p-values). We select genes with adjusted MAGMA p-value (specifically, FDR q-values) less than 0.05, as the set of disease-associated GWAS genes by. We set the minimum and maximum number of selected GWAS genes as 500 and 2,000, respectively, if the number of genes with FDR q-value < 0.05 is less than 500 or greater than 2,000, respectively. We elected to include genes in the MHC region, which are known to be involved in many complex human diseases [46, 47]; excluding these genes may lead to biased conclusions.

We note that while other methods for linking SNPs to genes exist (e.g., ref. [125, 137]), these methods typically use data derived from specific cell types and can bias the identification of disease-associated cell types. Therefore, we elected to not use these methods to select GWAS genes for scEPS analysis, to avoid potential biases.

### scEPS model

We model the disease phenotype of individual *i, y*_*i*_, given the pseudo-bulk gene expression at a neighborhood of cells, *x*_*i*_, using a linear model (dropping the index for cell neighborhood for notational simplicity),

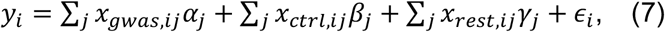

where *x*_*gwas,ij*_, *x*_*ctrl,ij*_, and *x*_*rest,ij*_ represent individual i-th’s expression of the *j*-th GWAS gene, control gene, and remaining (non-GWAS/control) gene, respectively;*α*_*j*_,*β*_*j*_, and *γ*_*j*_ represent the corresponding effect sizes; ϵ_*i*_ represents environmental effects. We select the set of control genes so that it matches the size and mean expression of the set of GWAS genes. We further assume that both the disease phenotype and gene expression are mean centered across individuals, that *E*[*α*_*j*_] = *E*[*β*_*j*_] = *E*[*γ*_*j*_] = 0 with *Var*[*α*_*j*_] = *σ*^2^_*gwas*_, *Var*[*β*_*j*_] = *σ*^2^_*ctrl*_, *Var*[*γ*_*j*_] = *σ*^2^_*rest*_, and that *E*[ϵ_*i*_] = 0 with *Var*[ϵ_*i*_] = *σ*^2^_*e*_. The parameters *σ*^2^_*gwas*_, *σ*^2^_*ctrl*_, and *σ*^2^_*rest*_ govern the magnitude of the effect sizes of the expression of GWAS, control, and remaining genes on disease phenotype; the parameter *σ*^2^_*e*_ governs the magnitude of the environmental effects on the disease phenotype.

We define the scEPS *d statistic* at the cell neighborhood as,

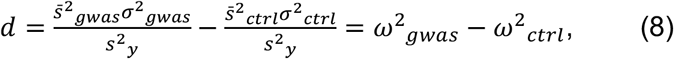

where 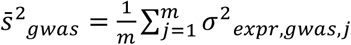 and 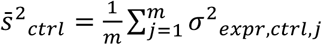 represent the average of the variance of the expression of each GWAS (*σ*^2^_*expr,gwas,j*_) and each control gene (*σ*^2^_*expr,ctrl,j*_) across donors, respectively; *s*^2^_*y*_ is the variance of the disease phenotype; *ω*^2^_*gwas*_ and *ω*^2^_*ctrl*_ represent the variance in the disease explained by each GWAS and each control gene, respectively. If there’s no phenotypic variation at the cell neighborhood (i.e., *s*^2^_*y*_ = 0), we set the scEPS *d* statistic to 0. We determine that the cell neighborhood is associated with the disease if *d* is greater than 0, i.e., if the expression of each GWAS gene explains more variation in the disease phenotype than each randomly selected control gene. Since different sets of randomly selected control genes likely have the same magnitude of per-gene effect size (i.e., same *σ*^2^_*ctrl*_), we elected to select one set of control genes to test for *d* statistics.

We also define 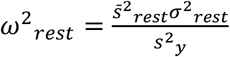, representing the variance in the disease phenotype explained by each of the remaining non-GWAS/control genes, where 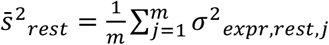 is the average of the variance of the expression of each remaining genes (*σ*^2^_*expr,rest,j*_) across donors; and 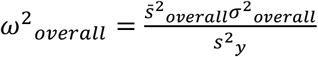, representing the variance in disease explained by each gene (both GWAS and non-GWAS genes), where 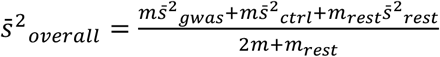 and 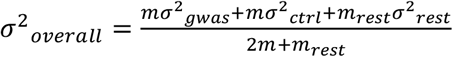. Here, 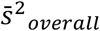 represents the average of the variance of the expression of each gene (i.e., GWAS, control, and remaining) across donors; and *σ*^2^_*overall*_ represents the overall variance of the per-gene effect size on disease phenotype.

We provide a detailed derivation of the scEPS statistics and an explanation of their relationship to genetic covariance when analyzing disease PRS and their behavior under different disease models in the “Supplementary notes”.

### Estimating the scEPS statistics

We use methods of moment to estimate *σ*^2^_*gwas*_ and *σ*^2^_*ctrl*_ by solving the regression equation,

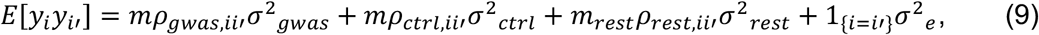

where *m* represents the number of GWAS and control genes; *m*_*rest*_ is the number of remaining genes; *ρ*_*gwas,ii*′_ = (∑_*j*_ *x*_*gwas,ij*_*x*_*gwas,i*′*j*_/*m, ρ*_*ctrl,ii*′_ = (∑_*j*_ *x*_*ctrl,ij*_*x*_*ctrl,i*′*j*_/*m*, and *ρ*_*rest,ii*′_ = (∑_*j*_ *x*_*rest,ij*_*x*_*rest,i*′*j*_/*m*_*rest*_ represent the covariance between individual *i* and *i*′ over the expression of the set of GWAS genes, control genes, and remaining genes; *σ*^2^_*gwas*_, *σ*^2^_*ctrl*_, *σ*^2^_*rest*_, and *σ*^2^_*e*_ are the respective variance components.

We obtain estimates of the variance components and standard errors, using weighted least square regression with 1/*Var*[*y*_*i*_*y*_*i*′_] as the weights to ad*j*ust for heteroscedasticity. Since statistical noise in the independent variables of the regression, *ρ*_*gwas,ii*′_, *ρ*_*ctrl,ii*′_, *ρ*_*rest,ii*′_, attenuates the regression coefficients, resulting in downward bias of the scEPS statistics, we apply a disattenuation factor estimated via bootstrapping [138] to mitigate the downward bias. We provide detailed derivation of the regression weights, standard errors, and disattenuation factors in “Supplementary notes”.

Let 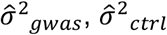, and 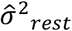 be the estimated *σ*^2^_*gwas*_, *σ*^2^_*ctrl*_, and *σ*^2^_*rest*_, respectively. We obtain estimates of *d, ω*^2^_*gwas*_, *ω*^2^_*ctrl*_, *ω*^2^_*rest*_, and *ω*^2^_*overall*_ as, 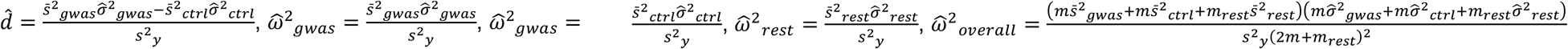, respectively. For neighborhoods with *s*^2^_*y*_ = 0, we set 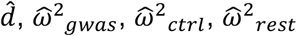, and 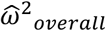 to 0.

### Significance testing of the scEPS statistics

We obtain the variance-covariance matrix for the estimated coefficients, 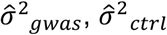, and 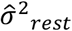, by bootstrapping across entries of the regression equation in Equation (9). Let 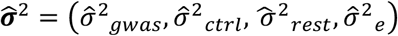 be the vector of estimated variance components, and 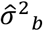 be the estimates obtained using the b-th bootstrap sample, we obtain the variance-covariance matrix for 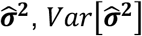, using,

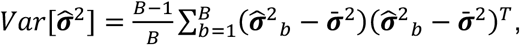

where 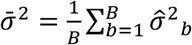. We set, *B*, the number of bootstrap samples to 1,000 by default.

We obtain the standard errors of the estimated 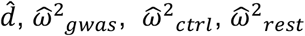, and 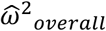, as, 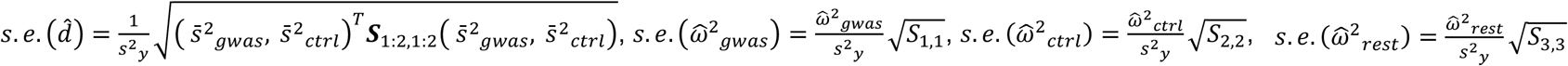, and 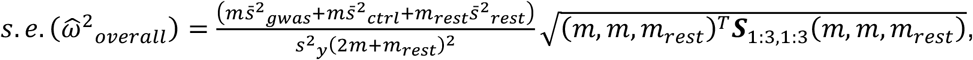 respectively. For neighborhoods with *s*^2^_*y*_ = 0, we set the standard errors of 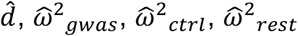, and 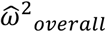to ∞.

We obtain test statistics testing 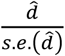 at each neighborhood as 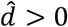, and obtain p-values from t-distributions with *N* − 6 degrees of freedom. We obtain test statistics for 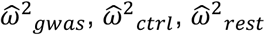, and 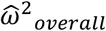 using similar approaches, and obtain p-values from t-distributions with *N* − 5, *N* − 5, *N* − 5, and *N* − 7 degrees of freedom, respectively.

### Aggregating the scEPS statistics

For a group of cell neighborhoods (e.g., from a specific cell type), we calculate the aggregated scEPS statistics as the average scEPS statistics of the cell neighborhoods constituting the group. In detail, let *G* ⊆ *C* represents a group of cell neighborhoods, we obtain aggregated 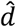 for *G*, 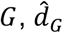, as, 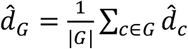. We obtain aggregated 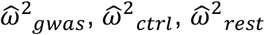, and 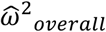 for *G* using similar approaches.

We obtain standard errors for the aggregated scEPS statistics using statistical block bootstrap [139], generating bootstrapped samples based on approximately independent blocks of cell neighborhoods. In detail, we group the cell neighborhoods into 100 (by default) approximately independent blocks, based on the neighborhood abundance matrix, ***Q***^*s*^ (see previous subsection), using mini-batch k-means clustering [140], relying on the heuristics that cell neighborhoods with similar donor compositions have correlated scEPS statistics. We note that bootstrap over individual cell neighborhoods treats each neighborhood as independent and would yield mis-calibrated standard error estimates.

Let *B* ⊆ *P*(*C*) represent the set of independent cell neighborhoods, where *P*(*C*) is the power set for the set of cells *C*. To generate the bootstrapped aggregated scEPS statistics at each bootstrap iteration, we first sample |*B*| sets of cells from *B* with replacement, yielding a set of bootstrapped cells, 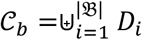, where *b* indexes the block bootstrap iteration; *D*_*i*_ is the *i*-th resampled set of cells; and ⊎ is the multiset sum operation, preserving duplicates. The aggregated 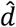 for *G* in the *b*-th iteration of bootstrap, 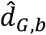, is then 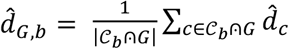, where ⩀ preserves duplicates in *C*_*b*_ in the intersection with *G*. The block bootstrap standard error for 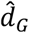 is then, 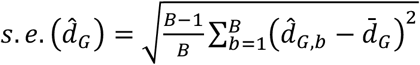, where 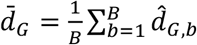, and *B* is the number of bootstrap samples (1,000 by default). The block bootstrap standard errors for other aggregated scEPS statistics can be obtained analogously. We obtain test statistics testing aggregated 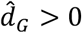 as 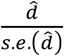 and obtain p-values from standard normal distributions. We obtain test statistics for other aggregated scEPS statistics, 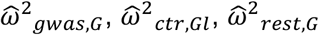, and 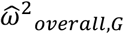, using similar approaches.

### Simulations

We simulated disease phenotypes based on the subset of snRNA-seq data for 37,720 microglia and perivascular macrophages (microglia-PVM) of 81 donors from the Seattle Alzheimer’s Disease Brain Cell Atlas (SEA-AD) Consortium [32]. We reserved the top *m* Alzheimer’s disease (AD) GWAS genes, obtained by applying MAGMA [13] to the summary statistics of a large-scale AD GWAS [37] (see subsections below), as the set of GWAS genes in the scEPS model, to capture realistic expression patterns of AD disease genes.

In each simulation, we first defined a cell neighborhood anchored at a randomly selected cell, using approaches described in previous subsections, and randomly selected a set of control genes that match the size and mean expression of the GWAS genes. Next, we simulated the phenotypes for the donors represented in the neighborhood based on a set of desired simulation parameters. Let *η*^2^_*total*_ be the desired total variance in phenotype explained by all genes, and *λ*_*gwas*_ be the desired enrichment of variance in phenotype explained by each GWAS gene. The variance of the effect size of each GWAS gene is then 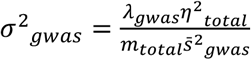, where *m*_*total*_ is the total number of genes; and 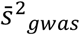 is the average variance of the expression of GWAS genes. Analogously, the variance of the effect size of each control and remaining gene is 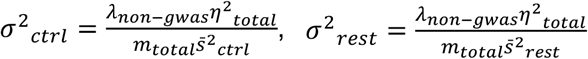, respectively, where 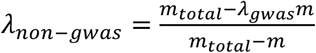 represents the depletion of variance in phenotype explained by each non-GWAS gene (i.e., control or remaining gene); 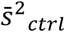 and 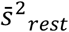 are the average variance of the expression of control and remaining genes, respectively. We simulated phenotypes using the linear model, ***y*** = ***X***_*gwas*_***β*** + ***X***_*ctrl*_***α*** + ***X***_*rest*_***γ*** + ***ϵ***, where ***X***_*gwas*_, ***X***_*ctrl*_, and ***X***_*rest*_ are the column-centered matrices of pseudo-bulk expression of the GWAS, control, and remaining genes, respectively; ***β, α***, and ***γ*** are the corresponding effect sizes; and ***ϵ*** represents environmental effects. We randomly selected *p*_*gwas,causal*_ fraction of GWAS genes and *p*_*non*−*gwas,causal*_ fraction of control and remaining genes as the causal genes with non-zero effect sizes, and drew effect sizes for each of these genes from the Gaussian distributions, 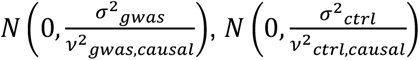,and 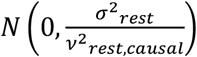, respectively, where *v*^2^_*gwas,causal*_,*v*^2^_*ctrl,causal*_, and *v*^2^_*rest,causal*_ represent the sum of the variances of the expression of the causal GWAS, control, and remaining genes, respectively. We drew the environmental effect for each donor from the the Gaussian distribution *N*(0, 1 − *η*^2^_*total*_).

We generated 2,000 simulations for each combination of simulation parameters (i.e., *λ*_*gwas*_, *η*^2^_*total*_, *m, p*_*gwas,causal*_, *p*_*non*−*gwas,causal*_. We assessed the impact of each simulation parameter on the performance of scEPS across a set of simulations by varying the parameter while fixing all other simulation parameters at their default values. Specifically, we assessed the impact of each simulation parameter, by choosing *λ*_*gwas*_ from {1.0, 4.0, 7.0, 10.0, 13.0} with *λ*_*gwas*_ = 7.0 as the default, *η*^2^_*total*_ from {0.0, 0.2, 0.4, 0.6} with *η*^2^_*total*_ = 0.4 as the default, *m* from {200, 600, 1000, 1400} with *m* = 600 as the default, *p*_*gwas,causal*_ and *p*_*non*−*gwas,causal*_ from {0.05,0.1, 0.2, 1.0} with *p*_*gwas,causal*_ = *p*_*non*−*gwas,causal*_ = 0.05 as the default.

### SEA-AD brain snRNA-seq and genotype data

We obtained brain single-nucleus RNA-seq (snRNA-seq) data, consisting of 1,378,211 brain cells from 89 donors, from the Seattle Alzheimer’s Disease Brain Cell Atlas (SEA-AD) consortium [32] (see “Data availability”). Detailed procedures for QC, cell-type labeling, and latent space embedding are described in ref. [32] Briefly, cells with fewer than 500 detected genes were removed from the brain cell atlas; scANVI [141] was used to predict cell-type labels for each single nucleus; scVI [142] was used to calculate single nucleus embedding in the harmonized latent space; doublets and cells with high fraction of mitochondrial reads were filtered [32]. We further restricted our analyses to the 1,203,090 brain cells from 81 donors of European ancestry (40 with dementia), to avoid the impact of ancestry in the analyses of PRSs. We provide visualizations of the SEA-AD brain snRNA-seq data in Supplementary Figure 29–30. We obtained imputed SNP array genotype data across 11,9705,313 SNPs for the donors in the SEA-AD consortium from NIAGADS Data Sharing Service (see “Data availability”). We further restricted our analyses of PRSs to the 41 control donors of European ancestry (see Table 1). We applied filtering and quality control to the genotype data using PGS Catalog Calculator [38] with default settings (see below).

### Translational Genomics Research Institute (TGen) lung scRNA-seq and genotype data

We obtained the TGen lung scRNA-seq data, consisting of 475,047 lung cells from 120 donors, from ref. [33](see “Data availability”). Detailed procedures for tissue processing, QC, cell-type labeling, and latent space embedding of the original TGen lung cell atlas data are described in ref. [33]. The TGen lung cell atlas data consists of both control donors and donors of 9 different diagnoses of respiratory disorders, including Idiopathic Pulmonary Fibrosis (IPF) and Interstitial Lung Disease (ILD), etc. We restricted our analyses to the 28 IPF and 31 control donors of European ancestry (228,086 cells). We calculated log-normalized gene expression with taget_sum=10000, obtained PCA using the top 2,000 highly variable genes, and applied Harmony [143] to integrate the lung scRNA-seq data across experimental batches. We provide visualizations of the TGen snRNA-seq data in Supplementary Figure 32. We obtained imputed SNP array genotype data across 8,481,051 SNPs for the donors in the TGen lung cell atlas data from dbGAP (see “Data availability”). We further restricted our analyses of PRSs to the 31 control donors of European ancestry (see Table 1). We applied filtering and quality control to the genotype data using PGS Catalog Calculator [38] with default settings (see below).

### ROSMAP brain snRNA-seq data

We obtained the Religious Orders Study/Memory and Aging Project (ROSMAP) [64] brain snRNA-seq data, consisting of 1,249,214 brain cells from 437 donors, from ref. [144] (see “Data availability”). Detailed QC procedures of the ROSMAP brain snRNA-seq data are described in ref. [144]. Briefly, cells with fewer than 500 unique molecular identifiers (UMIs) were removed [144]; genes expressed in fewer than 50 cells were removed [144]; and cells with more than 10% mitochondrial genes were removed. [144]. We did not filter for donors of European ancestry in the ROSMAP data as we did for the SEA-AD or TGen data, since we did not perform analysis of disease PRSs using the ROSMAP data. We down sampled the number of cells at random to 100,000 for astrocyte, excitatory neurons, and inhibitory neurons, due to memory constraints. We calculated log-normalized gene expression with taget_sum=10000, obtained PCA using the top 2,000 highly variable genes, and applied Harmony [143] to integrate the lung scRNA-seq data across experimental batches. We provide visualizations of the ROSMAP snRNA-seq data in Supplementary Figure 61.

### Calculating disease PRS

We calculated PRS, based on the genotype data of the donors in the SEA-AD brain cell atlas data and TGen lung cell atlas data, using the PGS Catalog Calculator [38] under default setting. In detail, we removed multi-allelic and strand-ambiguous SNPs prior to calculating PRS and ensured that at least 75% of the SNPs with PRS weights are available in the genotype data.

For each disease, we calculated PRS using publicly available pre-computed PRS weights from the PGS Catalog [38]. We used the raw scores, representing the sum of genotypes weighted by PRS weights, from the PGS Catalog Calculator for downstream analyses. We restricted our analyses of PRSs to donors of European ancestry only, as PRSs for non-European donors are known to be less accurate [40, 41, 42]. For diseases with more than one publicly available PRS weights, we selected the PRS yielding the highest scEPS *ω*^2^_*overall*_ statistics for downstream analyses. We report the list of disease PRSs analyzed in this study in Supplementary Table 7.

### scEPS analysis of 4 neurological and 4 respiratory disorders

We analyzed cognitive status (CS, defined as clinical diagnosis of dementia), and PRSs of Alzheimer’s disease (AD), multiple sclerosis (MS), and Parkinson’s disease (PD), integrating the SEA-AD brain cell atlas data with corresponding GWAS summary statistics data (Table 1). Because the SEA-AD cohort is an AD-specific cohort, we used the GWAS genes derived from the AD GWAS summary statistics data [37] in the analysis of CS. We also treated the analyses of CS and AD PRS (control donors only), as representing the active/symptomatic and preclinical/asymptomatic stages of AD, respectively.

We also analyzed Idiopathic Pulmonary Fibrosis (IPF), and PRSs of IPF, Chronic Obstructive Pulmonary Disease (COPD), and FEV1/FVC, integrating the TGen lung cell atlas data with corresponding GWAS summary statistics data (Table 1). Prior to running scEPS, we normalized the total read count of each cell in the brain and lung cell atlas data to 10,000, and applied log transformation. We also regressed out batch effects from the gene expression in each individual cell using the regress_out function of the scanpy package [145].

We performed scEPS analyses under the default settings (see subsection above). In the analyses of brain cell atlas data, we analyzed each of the 24 SEA-AD brain cell types independently in parallel, constructing separate k-NN graphs for each cell type, based on the batch-harmonized scVI embeddings. This reduced the memory requirement and run time. We note that since cells of different cell types are generally disjoint on the k-NN graph, analyzing them separately would have minimal impact on the results. In the analyses of lung cell atlas data, we analyzed all lung cell types together, constructing a joint k-NN graph for all cells, based on harmony [143] batch-corrected PCA. In the analysis of disease PRSs, we only used the cells from the control donors, to minimize the consequential effect of the disease on gene expression. In all scEPS analyses, we adjusted for age and sex as covariates at global level; we also adjusted for cell abundance of each donor at neighborhood level.

We estimated the scEPS statistics (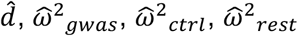, and 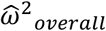) for each cell neighborhood in the brain and lung cell atlas data. We also obtained aggregated scEPS statistics for the 139 subtypes in the brain cell atlas data, and the 43 subtypes in the lung cell atlas data.

### Biological and cellular processes implicated by scEPS

We investigated the biological and cellular processes implicated by scEPS using gene set enrichment analysis (GSEA). First, we calculated the correlations (Spearman’s *ρ*) between the log-normalized expression of each gene with the *d* statistics across cells, in which the gene is expressed. We then applied GSEApy [71] to perform GSEA based on the ranked list of genes ordered by their correlations with the *d* statistics. We analyzed 2,785 gene sets from Reactome [70], which provides a both granular and hierarchical representation of biological and cellular processes.

We performed GSEA at two different levels. First, we performed global GSEA, based on the correlations across all cells, identifying the dominant disease-associated biological processes across cell types. We also performed cell-type-specific GSEA, based on the correlations across cells of each cell type, identifying context-dependent disease-associated biological processes.

### CNA*, a modification of CNA

We implemented CNA*, a modified version of CNA, to estimate the variance in disease phenotype explained by variations in neighborhood abundance. Specifically, let ***Q*** ∈ ℝ^|*C*|×*M*^ represent the neighborhood abundance matrix, across |*C*| cell neighborhoods for *M* individuals, let ***q***_*c*_ ∈ ℝ^*M*^ represent the neighborhood abundance across the *M* individuals at the neighborhood index by cell *c*, and let ***y*** ∈ ℝ^*M*^ represent the vector of phenotypes for the *M* individuals, CNA* aims to estimate the variance in ***y*** attributable to variations in ***q***_*c*_. We note that the metrics of disease association of CNA* and scEPS are both defined based on variance explained and are therefore comparable with each other.

We define cell neighborhoods and calculate neighborhood abundance using the same approach as described in ref. [34]. We model the disease phenotype as a linear function of neighborhood abundance, i.e., ***y*** = ***q***_*c*_*β*_*c*_ + ***ϵ***_*c*_, where *β*_*c*_ represents the effect size of the neighborhood abundance, ***q***_*c*_, on the phenotype, ***y***; and ***ϵ***_*c*_ represent the residual effects, specific to the neighborhood. We assume that ***y*** and ***q***_*c*_ are standardized to have mean 0 and unit variance, and that *β*_*c*_ is a random variable with *E*[*β*_*c*_] = 0 and *Var*[*β*_*c*_] = *σ*^2^_*na,c*_. The variance in ***y*** attributable to variations in ***q***_*c*_ is then *Var*[***q***_*c*_*β*_*c*_] = *σ*^2^_*na,c*_.

We estimate *σ*^2^_*na,c*_ using methods of moment. Briefly, it can be shown that *E*[*y*_*i*_*y*_*j*_] = *σ*^2^_*na,c*_*q*_*c,i*_*q*_*c,j*_ for *i* ≠ *j*. And an estimate of *σ*^2^_*na,c*_, 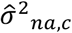, can be obtained via linear regression. We estimate the standard error of 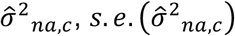, via statistical bootstrap (1,000 bootstrapped samples by default). We obtain test statistics testing 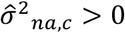 as 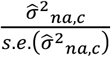, and obtain p-values under a t-distribution with *M* − 1 degrees of freedom.

We aggregate 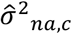 across a group of cell neighborhoods, *G* ⊆ *C*, by calculating the average 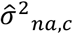 across the cell neighborhoods in *G*, i.e., 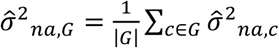, where 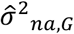 is the aggregated estimate. We calculate the standard error for 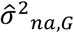 using statistical block bootstrap, similar to the approach for calculating the standard errors for the aggregated scEPS statistics (see subsection above). Briefly, let 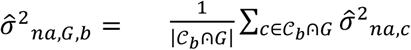 be the bootstrapped aggregated estimate for the *b*-th iteration, where *C*_*b*_ is the set of bootstrapped cell neighborhoods. The block bootstrap standard error for 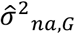 is then, 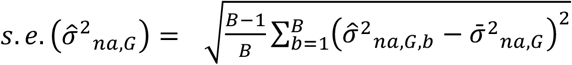, where 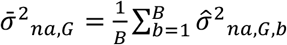, and *B* is the number of bootstrap samples (1,000 by default). We obtain test statistics testing aggregated 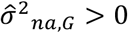 as 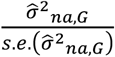, and obtain p-values from standard normal distributions.

### CNA* analysis of 4 neurological and 4 respiratory disorders

We applied CNA*, under the default setting (see subsection above), to analyze CS and PRSs of AD, MS, and PD using the SEA-AD brain cell atlas data, and to analyze IPF and PRSs of IPF, COPD, and FEV1/FVC using the TGen lung cell atlas data; these are the same disease phenotypes as analyzed by scEPS (Table 1). We constructed k-NN graphs across all cells, using the batch-harmonized scVI embeddings and the harmony batch-corrected PCA, for the brain and lung cell atlas, respectively.

In all CNA* analyses, we adjusted for age and sex as covariates, consistent with the scEPS analyses. In the analyses of disease PRSs, we only used the cells from the control donors, to minimize the consequential effect of the disease on cell abundance and to be consistent with scEPS analyses.

We obtained 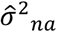 and test statistics for each individual cell neighborhood. We also obtained aggregated 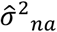 and test statistics for the 139 subtypes in the brain cell atlas data, and the 43 subtypes in the lung cell atlas data.

### scDRS analysis of 4 neurological and 4 respiratory disorders

We applied scDRS to analyze AD, MS, and PD, using the SEA-AD brain cell atlas data, and to analyze IPF, COPD, and FEV1/FVC using the TGen lung cell atlas data, all starting from the raw count data. We used the same GWAS summary statistics data as used by scEPS (Table 1) to select GWAS genes. To be consistent with scEPS analyses, we applied scDRS to analyze the same set of GWAS genes as analyzed by scEPS (Table 1), instead of the top 1,000 MAGMA prioritized GWAS genes by default.

For each disease, we ran scDRS using cells from all (cases and controls) donors, which we compared with scEPS and CNA* analyses of clinical diagnoses for CS and IPF. We also ran scDRS using cells from control donors only, which we compared with scEPS and CNA* analyses of PRSs of AD, MS, PD, IPF, COPD, and FEV1/FVC.

We obtained scDRS raw scores and test statistics at individual cell level. We also obtained aggregated scDRS raw scores and test statistics for the 143 subtypes in the brain cell atlas data, and the 43 subtypes in the lung cell atlas data.

## Data availability

The SEA-AD snRNA-seq data for brain cells is available on Chan Zuckerberg CELLxGENE Discover at https://cellxgene.cziscience.com/collections/1ca90a2d-2943-483d-b678-b809bf464c30. The genotype data for the SEA-AD brain cell atlas donors is available on NIAGADS Data Sharing Service (accession number: NG00174.v1). The Translational Genomics Research Institute (TGen) scRNA-seq data for lung cells is available on NCBI Gene Expression Omnibus (accession number: GSE227136). The genotype data for the TGen lung cell atlas donors is available on dbGaP (accession number: phs003521.v1.p1). The ROSMAP snRNA-seq data for brain cells is available on Synapse (Project SynID: syn2580853). scEPS statistics at individual cell neighborhood resolution for the 4 neurological and 4 respiratory disorders analyzed in this study can be downloaded at https://figshare.com/s/9855b32b6179ecf9ceb0.

## Data Availability

scEPS statistics at individual cell neighborhood resolution for the 4 neurological and 4 respiratory disorders analyzed in this study will be available at https://figshare.com/s/9855b32b6179ecf9ceb0 upon publication.

## Code availability

Python code implementing the scEPS method is available at https://github.com/Genentech/sceps. Python code implementing the CNA* method is available at https://github.com/Genentech/cna_star. Python code implementing the simulation procedure is available at https://github.com/Genentech/sceps_sim.

## Acknowledgements

The authors are grateful to M. Crow, L. Orozco, O. Mayba, B. Friedman, and 2 Genentech internal reviewers for helpful discussions.

## Author contributions

H.S. designed the computational methods with input from all co-authors. L.Z., O.W., and H.S., performed the computational analyses. L.Z., O.W., and H.S. wrote the paper with assistance from all authors.

