## Supplementary Information for "scEPS integrates genetic and single-cell disease atlas data to provide granular mechanistic insights into complex human diseases"

#### Supplementary notes

##### Deriving the scEPS model

We model the disease outcome of individual $i$, $y_i$, given the pseudo-bulk gene expression at a neighborhood of cells, $x_i$, using a linear model (dropping index for cell neighborhood for notational simplicity),

$$y_i = \sum_j x_{gwas,ij}\alpha_j + \sum_j x_{ctrl,ij}\beta_j + \sum_j x_{rest,ij}\gamma_j + \epsilon_i, \text{ (1)}$$

where $x_{gwas,ij}$, $x_{ctrl,ij}$, and $x_{rest,ij}$ represent individual i-th's expression of the j-th genetically nominated disease gene, control gene, and remaining gene, respectively; $\alpha_j$, $\beta_j$, and $\gamma_j$ represent the corresponding effect sizes, with mean 0, and $Var[\alpha_j] = \sigma^2{}_{gwas}$, $Var[\beta_j] = \sigma^2{}_{ctrl}$, $Var[\gamma_j] = \sigma^2{}_{rest}$; $\epsilon_i$ represents environmental effects, with mean 0 and $Var[\epsilon_i] = \sigma^2{}_e$. We assume that both the disease outcome and gene expression are mean centered but not standardized across individuals, so that the effect sizes, $\alpha_j$, $\beta_j$, and $\gamma_j$, represent the increase in disease risk per unit increase in gene expression, relative to the population mean.

We elected not to model standardized gene expression and effect sizes for both interpretability and practicality. First, modeling standardized gene expression and effect sizes entails a coupling between the effect sizes and variances of the unstandardized gene expression, such that genes with lower expression variances have higher magnitude of effect sizes than genes with higher expression variances. Specifically, the relationship between the effect size of standardized gene expression, $\beta_{std}$, and its unstandardized counterpart, $\beta_{unstd}$, for a gene with standard deviation, $\sigma$, is

$$\beta_{unstd} = \beta_{std}\sigma.$$

This relationship stipulates that for the same $\beta_{std}$, genes with lower standard deviation have higher magnitudes of $\beta_{unstd}$. This implicit assumption entailed by modeling standardized gene expression complicates the interpretation of the scEPS model parameters. Second, normalizing gene expression values by their standard deviations creates statistical noise in the estimation process, especially for genes with standard deviation close to 0, leading to large standard errors in the estimated model parameters. Modeling unstandardized gene expression avoids the normalization of the standard deviation of gene expression, resulting in smaller standard errors in estimated model parameters.

The scEPS model parameters consist of $\sigma^2{}_{gwas}$, $\sigma^2{}_{ctrl}$, $\sigma^2{}_{rest}$, and $\sigma^2{}_e$, the variance components governing the distribution of $\alpha_j$, $\beta_j$, $\gamma_j$, and $\epsilon_i$, respectively. Although the effect sizes of individual genes on the disease outcome (i.e., $\alpha_j$, $\beta_j$, and $\gamma_j$) are involved in defining the scEPS model in Equation (1), they are nuance parameters and not part of the scEPS model parameters.

### Defining the scEPS statistics

We define scEPS statistics based on the scEPS model in Equation (1). We determine that the cell neighborhood is critical for a disease if the expression of genetically nominated genes explain more variance in disease than match control genes, i.e., $Var\left[\sum_j x_{gwas,ij}\alpha_j\right] > Var\left[\sum_j x_{ctrl,ij}\beta_j\right]$, which is equivalent to testing,

$$\frac{1}{m} Var\left[\sum_j x_{gwas,ij}\alpha_j\right] - \frac{1}{m} Var\left[\sum_j x_{ctrl,ij}\beta_j\right] > 0,$$

where $m$ is the number of genetically nominated genes and matched control genes. We simplify the left-hand side of the inequality as, $\bar{s}^2{}_{gwas}\sigma^2{}_{gwas} - \bar{s}^2{}_{ctrl}\sigma^2{}_{ctrl}$, where $\bar{s}^2{}_{gwas}$ and $\bar{s}^2{}_{ctrl}$ represent the average variance of the expression of genetically nominated genes and control genes, respectively. We normalize the statistic by the variance of the disease outcome in the neighborhood, $s^2{}_y$, so that results are comparable across cell neighborhoods, yielding the scEPS $d$ *statistic*,

$$d = \frac{\bar{s}^2{}_{gwas}\sigma^2{}_{gwas} - \bar{s}^2{}_{ctrl}\sigma^2{}_{ctrl}}{s^2{}_y}.$$

In cases where $s^2{}_y$ (i.e., no variation in disease outcome in the neighborhood), we set the scEPS $d$ statistic to 0. Effectively, the scEPS $d$ statistic quantifies the difference in variance in disease outcome attributable to the expression of *each* genetically nominated gene vs. *each* matched control gene at a cell neighborhood.

We also define the following statistics:

- $\omega^2{}_{gwas} = \frac{\bar{s}^2{}_{gwas}\sigma^2{}_{gwas}}{s^2{}_y}$, the variance in disease outcomes attributable to the expression of each genetically nominated gene;
- $\omega^2{}_{ctrl} = \frac{\bar{s}^2{}_{ctrl}\sigma^2{}_{ctrl}}{s^2{}_y}$, the variance in disease outcomes attributable to the expression of each control gene matched with genetically nominated genes on mean expression;
- $\omega^2{}_{rest} = \frac{\bar{s}^2{}_{rest}\sigma^2{}_{rest}}{s^2{}_y}$, the variance in disease outcomes attributable to the expression of each remaining gene, where $\bar{s}^2{}_{rest}$ is the average variance of the remaining genes;
- $\omega^2{}_{overall} = \frac{\bar{s}^2{}_{overall}\sigma^2{}_{overall}}{s^2{}_y}$, the variance in disease outcomes attributable to each gene, regardless of whether they are genetically nominated or not.

### Estimating the scEPS model parameters using methods of moments

We estimate the scEPS model parameters, $\sigma^2{}_{gwas}, \sigma^2{}_{ctrl}$, $\sigma^2{}_{rest}$, and $\sigma^2{}_e$ using methods of moment (MoM).
Let

$$y_i = \sum_j x_{gwas,ij}\alpha_j + \sum_j x_{ctrl,ij}\beta_j + \sum_j x_{rest,ij}\gamma_j + \epsilon_i,$$
$$y_{i'} = \sum_j x_{gwas,i'j}\alpha_j + \sum_j x_{ctrl,i'j}\beta_j + \sum_j x_{rest,i'j}\gamma_j + \epsilon_{i'}.$$

It can be shown that

$$E[y_i y_{i'}] = E\left[\left(\sum_j x_{gwas,ij}\alpha_j\right)\left(\sum_j x_{gwas,i'j}\alpha_j\right)\right] + E\left[\left(\sum_j x_{ctrl,ij}\alpha_j\right)\left(\sum_j x_{ctrl,i'j}\alpha_j\right)\right]$$
$$+ E\left[\left(\sum_j x_{rest,ij}\alpha_j\right)\left(\sum_j x_{rest,i'j}\alpha_j\right)\right] + E[\epsilon_i\epsilon_{i'}]$$
$$= E[\alpha_j\alpha_j]\sum_j x_{gwas,ij}x_{gwas,i'j} + E[\beta_j\beta_j]\sum_j x_{ctrl,ij}x_{ctrl,i'j} + E[\gamma_j\gamma_j]\sum_j x_{rest,ij}x_{rest,i'j} + 1_{\{i=i'\}}\sigma^2{}_e$$
$$= m\rho_{gwas,ii'}\sigma^2{}_{gwas} + m\rho_{ctrl,ii'}\sigma^2{}_{ctrl} + m_{rest}\rho_{rest,ii'}\sigma^2{}_{rest} + 1_{\{i=i'\}}\sigma^2{}_e, \quad (2)$$

where $m$ represents the number of genetically nominated and matched control genes; $m_{rest}$ is the number of remaining genes; $\rho_{gwas,ii'} = \sum_j x_{gwas,ij} x_{gwas,i'j}/m$, $\rho_{ctrl,ii'} = \sum_j x_{ctrl,ij} x_{ctrl,i'j}/m$, and $\rho_{rest,ii'} = \sum_j x_{rest,ij} x_{rest,i'j}/m_{rest}$ represent the covariance between individual $i$ and $i'$ over the expression of the set of genetically nominated disease genes, control genes, and remaining genes. Estimating the scEPS model parameters amounts to solving the system of linear equations defined by $y_i y_{i'}$, $\rho_{gwas,ii'}$, $\rho_{ctrl,ii'}$, and $\rho_{rest,ii'}$.

Although maximum likelihood (ML) based approaches tend to be more statistically efficient, yielding estimates with lower standard errors, we elected not to use ML for both computational efficiency and robustness reasons. ML typically employs iterative procedures for model fitting that is not guaranteed to converge, increasing running time and potentially resulting in neighborhoods with sub-optimal estimates. Despite larger standard errors, MoM is both computationally efficient and highly robust.

#### Regression weights

Differences in the variances of the response variable, $y_i y_{i'}$, in the MoM regression equation in Equation (2) results in heteroskedasticity, yielding reduced statistical efficiency (higher standard errors) of the estimates of the scEPS model parameters. We account for heteroskedasticity using weighted least square regression, with weights set to $1/Var[y_i y_{i'}]$.

It can be shown that

$$\begin{aligned} Var[y_i y_{i'}] &= Cov\left[{y^2}_i, {y^2}_{i'}\right] + Var[y_i]Var[y_{i'}] - Cov[y_i, y_{i'}]^2 \\ &= 2Cov[y_i, y_{i'}]^2 + Var[y_i]Var[y_{i'}] - Cov[y_i, y_{i'}]^2 \\ &= Cov[y_i, y_{i'}]^2 + Var[y_i]Var[y_{i'}] \end{aligned}$$

where $Var[y_i] = Var[y_{i'}] = {s^2}_y$ is the sample variance of the disease outcome in the cell neighborhood.

We estimate $Cov[y_i, y_{i'}]$ using a 2-step approach. First, we perform a univariate ordinary least square regression to obtain an estimate of the variance of the effect size of the expression of each gene on disease outcome, ${\sigma^2}_{overall}$, solving,

$$E[y_i y_{i'}] = m_{all} \rho_{ii'} {\sigma^2}_{overall}, \quad (3)$$

restricting to $i \neq i'$, where $m_{all}$ is the total number of genes; $\rho_{ii'} = \left(\sum_j x_{ij} x_{i'j}\right)/m_{all}$ represents the covariance between individual $i$ and $i'$ over the expression of all genes. Let ${\hat{\sigma}^2}_{overall,init}$ be initial the estimate of ${\sigma^2}_{overall}$ through ordinary least square regression, we further regularize ${\hat{\sigma}^2}_{overall,init}$ by limiting its value to $\left[0, \frac{Var[y]}{m_{all} {s^2}_{overall}}\right]$, where ${s^2}_{overall}$ is the average variance of the expression across all genes, yielding ${\hat{\sigma}^2}_{overall}$. We then plug in ${\hat{\sigma}^2}_{overall}$ into Equation (3), yielding,

$$Cov[y_i, y_{,}] = m_{all} \rho_{ii'} {\hat{\sigma}^2}_{overall}.$$

Finally, for $i \neq i'$, we calculate the regression weights using,

$$w_{ii'} = (m_{all} \rho_{ii'} {\hat{\sigma}^2}_{overall})^2 + {s^2}_y;$$

and for $i = i'$, we calculate the regression weights using,

$$w_{ii} = 2{s^2}_y.$$

#### Disattenuation factors

Statistical noises in the vectors, $\boldsymbol{\rho}_{gwas} = (\rho_{gwas,ii'})$, $\boldsymbol{\rho}_{ctrl} = (\rho_{ctrl,ii'})$, and $\boldsymbol{\rho}_{rest} = (\rho_{rest,ii'})$in the design matrix of the MoM equation in Equation (2) attenuates the estimates of ${\sigma^2}_{gwas}$, ${\sigma^2}_{ctrl}$, and ${\sigma^2}_{rest}$ , resulting in downward biases in the scEPS statistics. We alleviate these downward biases by applying a disattenuation factor, calculated using a bootstrap based approach, to each of the estimated scEPS model parameters.

Here, we derive the steps to estimate the disattenuation factor for ${\sigma^2}_{gwas}$, following the procedure described in ref. [138] – disattenuation factors for ${\sigma^2}_{ctrl}$ and ${\sigma^2}_{rest}$ can be obtained using analogous procedures. We first obtain, $\boldsymbol{P}_{gwas} \in \mathbb{R}^{m\times 1000}$, each column of which represents a bootstrapped $\boldsymbol{\rho}_{gwas}$, calculated by sampling genes with replacement. Given the bootstrapped samples in $\boldsymbol{P}_{gwas}$, we then estimate the proportion of noise in the observed $\rho_{gwas}$ using,

$$r_{noise} = \frac{{\sigma^2}_b}{{\sigma^2}_b+{\sigma^2}_w},$$

where ${\sigma^2}_b = Var\left[\boldsymbol{P}_{gwas,\cdot,\cdot} - E\left[\boldsymbol{P}_{gwas,\cdot,\cdot}\right]\right]$ represents the variation in $\boldsymbol{\rho}_{gwas}$ driven by variations in the sampled genes; ${\sigma^2}_w = Var\left[\boldsymbol{P}_{gwas,:,\cdot} - E\left[\boldsymbol{P}_{gwas,:,\cdot}\right]\right]$ represents variations in $\boldsymbol{\rho}_{gwas}$ across pairs of samples. We calculate the disattenuation for the estimated ${\sigma^2}_{gwas}$ using,

$$f_{disatt} = \frac{1}{r_{noise}} = 1 + \frac{{\sigma^2}_w}{{\sigma^2}_b}.$$

We treat $f_{disatt}$ as a fixed constant in driving the variances for the estimated scEPS model parameters – variations in $f_{disatt}$ is negligible with 1,000 bootstrapped samples.

### Estimating and testing the statistical significance of the scEPS statistics at individual cell neighborhood

Let ${\hat{\sigma}^2}_{gwas}$, ${\hat{\sigma}^2}_{ctrl}$, and ${\hat{\sigma}^2}_{rest}$ be the scEPS model parameters estimated via weighted least square regression, disattenuated to account for regression dilution. We estimate each scEPS statistics defined in the previous subsection, by plugging in ${\hat{\sigma}^2}_{gwas}$, ${\hat{\sigma}^2}_{ctrl}$, and ${\hat{\sigma}^2}_{rest}$ into the definitions of the statistic.

We derive and implement an analytical as well as a permutation-based approach to test the statistical significance of the scEPS statistics. We use the more computationally efficient analytical approach as the default approach; results from simulations suggest that test statistics obtained from either approach are comparable.

#### Analytical approach

At each cell neighborhood, we group all genes into $B$ (10 by default) bins, based on the mean and variance of the neighborhood-specific pseudo-bulk expression across individuals. We then sample control genes from each bin, in proportion to the number of genetically nominated genes in the bin, prioritizing selecting genes with lower MAGMA gene-level association statistics, yielding a set of control genes with the same size as that of GWAS disease genes.

Let $\hat{\boldsymbol{\sigma}}^2 = \left({\hat{\sigma}^2}_{gwas}, {\hat{\sigma}^2}_{ctrl}, {\hat{\sigma}^2}_{rest}, {\hat{\sigma}^2}_{e}\right)$ be the vector of estimated scEPS model parameters, and $\boldsymbol{S} \in \mathbb{R}^{4\times 4}$ the corresponding variance-covariance matrix, from the weighted least square regression. The standard error for the scEPS $d$ statistic is

$$s.e.\left(\hat{d}\right) = \frac{\sqrt{\left({\bar{s}^2}_{gwas}, -{\bar{s}^2}_{ctrl}\right)^T \boldsymbol{S}_{1:2,1:2}\left({\bar{s}^2}_{gwas}, -{\bar{s}^2}_{ctrl}\right)}}{{s^2}_y}$$

the standard errors for the estimated scEPS ${\omega^2}_{gwas}$, ${\omega^2}_{ctrl}$, and ${\omega^2}_{rest}$ statistics are

$$s.e.\left(\widehat{\omega}^2{}_{gwas}\right) = \frac{\sqrt{\bar{s}^2{}_{gwas} S_{1,1}}}{s^2{}_y},\ s.e.\left(\widehat{\omega}^2{}_{ctrl}\right) = \frac{\sqrt{\bar{s}^2{}_{ctrl} S_{2,2}}}{s^2{}_y},\ s.e.\left(\widehat{\omega}^2{}_{rest}\right) = \frac{\sqrt{\bar{s}^2{}_{rest} S_{3,3}}}{s^2{}_y},$$

respectively; the standard error for the estimated scEPS $\omega^2{}_{overall}$ statistics is

$$s.e.\left(\widehat{\omega}^2{}_{overall}\right) = \frac{\sqrt{\left(\bar{s}^2{}_{gwas}, \bar{s}^2{}_{ctrl}, \bar{s}^2{}_{rest}\right)^T \boldsymbol{S}_{1:3,1:3} \left(\bar{s}^2{}_{gwas}, \bar{s}^2{}_{ctrl}, \bar{s}^2{}_{rest}\right)}}{s^2{}_y}.$$

We calculate the Z-score testing the null hypothesis of $d = 0$ using $\frac{\hat{d}}{s.e.(\hat{d})}$ and obtain 2-tailed p-value from a $t$ distribution with $N - 5$ degrees of freedom, where $N$ is the number of donors. We calculate the Z-scores testing the null hypotheses of $\omega^2{}_{gwas} = 0$, $\omega^2{}_{ctrl} = 0$, and $\omega^2{}_{rest} = 0$, using $\frac{\widehat{\omega}^2{}_{gwas}}{s.e.(\widehat{\omega}^2{}_{gwas})}$, $\frac{\widehat{\omega}^2{}_{ctrl}}{s.e.(\widehat{\omega}^2{}_{ctrl})}$, and $\frac{\widehat{\omega}^2{}_{rest}}{s.e.(\widehat{\omega}^2{}_{rest})}$, respectively, and obtain 2-tailed p-values from $t$ distributions with $N - 4$ degrees of freedom. We calculate the Z-score testing the null hypothesis of $\omega^2{}_{overall} = 0$ using $\frac{\widehat{\omega}^2{}_{overall}}{s.e.(\widehat{\omega}^2{}_{overall})}$ and obtain 2-tailed p-value from a $t$ distribution with $N - 6$ degrees of freedom.

##### Permutation-based approach

We generate a null distribution for each of the scEPS statistics by randomly permuting the disease outcomes of the donors, randomly sampling a different set of control genes for each permutation of disease outcomes.

Let $\hat{\tau}$ represent an estimated scEPS statistic using the original unpermuted outcomes, and $\hat{T}$ represent the set of estimated scEPS statistic using permuted outcomes ($|\hat{T}| = 1000$ by default). We obtain p-values testing the null hypothesis of $\tau = 0$ using,

$$p = \frac{1 + \sum_{i=1}^{|\hat{T}|} 1_{\{|\hat{T}_i| > \hat{\tau}\}}}{1 + |\hat{T}|}.$$

#### Relationship between scEPS statistics and genetic covariance when analyzing PRS

We show that the scEPS statistics are related to the genome-wide genetic covariance between gene expression and complex disease when the disease phenotype, $\boldsymbol{y}$, is the disease polygenic risk score (PRS), i.e., $\boldsymbol{y} = \boldsymbol{Gz}$, where $\boldsymbol{G} \in \mathbb{R}^{N \times M}$ is the genotype matrix for the $N$ donors in the scRNA-seq data across $M$ SNPs used for calculating the PRS, and $\boldsymbol{z} \in \Re^M$ is the PRS weights (e.g., obtained using LassoSum2, PRS-CS, etc.).

We consider the linear model for the relationship between the mean centered gene expression $\boldsymbol{X} = \left[\boldsymbol{X}_{gwas}\ \boldsymbol{X}_{ctrl}\ \boldsymbol{X}_{rest}\right] \in \mathbb{R}^{(m+m+m_{rest}) \times N}$ and PRS at a cell neighborhood: $\boldsymbol{Gz} = \boldsymbol{X\xi} + \boldsymbol{\epsilon}$, where $\boldsymbol{\xi} = [\boldsymbol{\alpha};\ \boldsymbol{\beta};\ \boldsymbol{\gamma}] \in \mathbb{R}^{(m+m+m_{rest})}$ is the expression-to-disease effect size vector; $\boldsymbol{\epsilon} \in \mathbb{R}^N$ is the vector of environmental effects. For convenience of illustration, we show how to estimate the $\sigma^2{}_{gwas}$ parameter as $N \to \infty$ ; other parameters $\sigma^2{}_{ctrl}$ and $\sigma^2{}_{rest}$ can be estimated analogously. As $N \to \infty$, one can estimate $\sigma^2{}_{gwas}$ as

$$\hat{\sigma}^2{}_{gwas} = Var\left[\hat{\alpha}_j\right] = \frac{1}{m} \sum_{j=1}^{2m+m_{rest}} \mathbf{1}_{\{j \in G_{gwas}\}} \hat{\xi}_j{}^2,$$

where $m$ is the number of GWAS genes; $(2m + m_{rest})$ is the total number of genes; $\mathbf{1}_{\{j \in G_{gwas}\}} = 1$ if gene $j$ is in $G_{gwas}$, the set of GWAS genes, and 0 otherwise. And $\hat{\xi}_j$ is

$$\hat{\xi}_j = (\boldsymbol{X}^T \boldsymbol{X})^{-1} (\boldsymbol{X}^T \boldsymbol{Gz})_j. \quad (4)$$

We express $\boldsymbol{X}$ as $\boldsymbol{X} = \boldsymbol{G}\boldsymbol{B} + \boldsymbol{E}$, where $\boldsymbol{B} \in \mathbb{R}^{M\times(2m+m_{rest})}$ is the matrix of effect sizes of $M$ SNPs on the expression of all $(2m + m_{rest})$ genes, and $\boldsymbol{E} \in \mathbb{R}^{N\times(2m+m_{rest})}$ is the matrix of environmental effects on gene expression. We can then express $\xi_j$ in Equation (4) as,

$$\hat{\xi}_j = (\boldsymbol{X}^T\boldsymbol{X})^{-1}[(\boldsymbol{G}\boldsymbol{B}+\boldsymbol{E})^T\boldsymbol{G}\boldsymbol{z}]_j = (\boldsymbol{X}^T\boldsymbol{X})^{-1}[\boldsymbol{B}^T\boldsymbol{G}^T\boldsymbol{G}\boldsymbol{z} + \boldsymbol{E}^T\boldsymbol{G}\boldsymbol{z}]_j = (\boldsymbol{X}^T\boldsymbol{X})^{-1}[M\boldsymbol{B}^T\boldsymbol{V}\boldsymbol{z} + \boldsymbol{E}^T\boldsymbol{G}\boldsymbol{z}]_j,$$

where $\boldsymbol{V}^{M\times M}$ is the linkage disequilibrium (LD) matrix for the $M$ SNPs; $\rho_{g,j} = (\boldsymbol{B}^T\boldsymbol{V}\boldsymbol{z})$ is the genome-wide genetic covariance between gene $j$ and the complex disease; $\rho_{e,j} = \frac{1}{N}(\boldsymbol{E}^T\boldsymbol{G}\boldsymbol{z})_j$ is the covariance between environmental effects on the expression of gene $j$ and disease PRS, assumed to be zero.

In the special case, where $\boldsymbol{X}^T\boldsymbol{X}$ is a diagonal matrix (i.e., the expression of pairs of genes is independent),

$$\hat{\xi}_j = \frac{M\rho_{g,j}}{(2m+m_{rest})Var[\boldsymbol{X}_j]} + \frac{N\rho_{e,j}}{(2m+m_{rest})Var[\boldsymbol{X}_j]} = \frac{M\rho_{g,j}}{(2m+m_{rest})Var[\boldsymbol{X}_j]},$$

and

$$\hat{\sigma}^2{}_{gwas} = \frac{1}{m}\sum_{j=1}^{(2m+m_{rest})} \mathbf{1}_{\{j\in G_{gwas}\}} \left[\frac{M\rho_{g,j}}{(2m+m_{rest})Var[\boldsymbol{X}_j]}\right]^2.$$

Thus, the $\sigma^2{}_{gwas}$ statistic, under the assumption that the expression of pairs of genes is independent, is a function of the squared genome-wide genetic covariance between gene expression and complex disease. In the more general case, where $\boldsymbol{X}^T\boldsymbol{X}$ is not diagonal (i.e., the expression of pairs of genes is correlated), the $\sigma^2{}_{gwas}$ statistic is then a function of the squared genome-wide genetic covariance between the decorrelated/whitened gene expression, $(\boldsymbol{X}^T\boldsymbol{X})^{-1}$, and complex disease.

The scEPS $d$ statistic, $d = \frac{\bar{s}^2{}_{gwas}\sigma^2{}_{gwas} - \bar{s}^2{}_{ctrl}\sigma^2{}_{ctrl}}{s^2{}_y}$, effectively tests for the difference, scaled by the mean variance of gene expression, between the average squared genome-wide genetic covariance between the expression of GWAS genes and randomly selected matched control genes. Ideally, replacing the mean variance of gene expression with the mean heritability of gene expression would be more congruent with genetic covariance. However, estimating the heritability for each gene at each cell neighborhood can be computationally costly, and is beyond the scope of this work.

### Relationship between scEPS statistics and disease models

We consider 4 representative models linking SNPs, gene expression, and disease phenotype, as illustrated in Supplementary Figure 66: causality model, pleiotropy model, reverse causality model, and confounder model. Other more complicated models (e.g., models involving gene-environment interactions and/or feedback loops) are also possible but are beyond the scope of this work. Below, we illustrate the properties of the scEPS $d$ statistics, obtained from analyzing disease diagnosis as well as disease PRS of healthy donors.

Under the causality model (Supplementary Figure 66**a**), gene expression causally impacts the disease, resulting in differential gene expression across both the disease diagnosis and disease PRS – both the overall and genetic component of expression impacts the disease under the causality model. Thus, the variance in disease phenotype explained by expression is non-zero for both disease diagnosis and disease PRS under the causality model. Positive $d$ statistics would suggest that the causal effects of the expression of GWAS genes on disease diagnosis/PRS are stronger than matched control genes.

Under the pleiotropy model (Supplementary Figure 66**b**), the disease phenotype and gene expression are both impacted by a shared genetic factor (e.g., expression of another gene, protein, etc.). Changes in the shared genetic factor would result in coordinated changes in both gene expression and disease phenotype, inducing differential gene expression across both disease diagnosis and disease PRS, and thus non-zero variance explained by gene expression. Positive $d$ statistic would suggest that the coordinated effects of the shared heritable factor on gene expression and disease phenotype are stronger and more correlated for GWAS genes than matched control genes.

Under the reverse causality model (Supplementary Figure 66**c**), gene expression is impacted by the disease phenotype, resulting in differential gene expression and positive variance explained by gene expression for both disease phenotype and disease PRS under the reverse causality model. Positive $d$ statistics would suggest that the causal effects of the disease phenotype on gene expression are stronger at GWAS genes than matched control genes. We note that when scEPS is applied to the disease PRS of healthy donors, the effect of reverse causality can be reduced, as a diagnosable disease is not manifested in the donors. However, the scEPS statistics can still be a result of the "pre-disease", changes in cellular functions/processes before the onset of the disease. Thus, positive $d$ statistics would not imply a causal association.

Under the confounder model (Supplementary Figure 66**d**), gene expression and disease phenotype are independently impacted by genetic variations, and are both causally impacted by a shared non-heritable confounder (e.g., exposure to a certain environment). Changes in the shared non-heritable factor would result in coordinated changes in both gene expression and disease diagnosis, inducing non-zero variance explained by gene expression in disease diagnosis; positive $d$ statistics would imply a stronger and more correlated effect of the non-heritable confounder on the expression and disease diagnosis. However, the non-heritable confounder would not result in coordinated changes in the gene expression and the disease PRS (i.e., genetic component of the disease), and the gene expression would not explain variance in the disease PRS. Thus, under the confounder model, the scEPS $d$ statistics would be zero.

In summary, the scEPS $d$ statistics obtained from analyzing disease PRS of healthy donors can distinguish the confounder model from the remaining models and can mitigate the effect of reverse causality but cannot distinguish the pleiotropy model from the causality model. Although the $d$ statistics obtained from analyzing disease diagnosis cannot distinguish the 4 models, they highlight cell populations where GWAS genes are more strongly associated with the disease than matched control genes.

### Supplementary Tables

| Method | Disease association based on [a] | Directly models disease [b] | Incorporate GWAS [c] | Models up or down regulation of expression [d] | Resolution [e] |
|---|---|---|---|---|---|
| scEPS (this work) | variance in disease explained by gene expression | Yes | Yes | both up and down regulation | cell neighborhood anchored at individual cells |
| CNA [34] | covariance between cell abundance and disease at each cell neighborhood | Yes | No | not applicable | cell neighborhood anchored at individual cells |
| Milo [28] | covariance between cell abundance and disease at each cell neighborhood | Yes | No | not applicable | cell neighborhood |
| scDRS [24] | overexpression of GWAS genes | No | Yes | up regulation only | individual cell |
| sc-linker [31] | heritability enrichment of disease-associated genes / gene expression programs | No | Yes | both up and down regulation | cellular processes/programs |
| LDSC-SEG [27] | heritability enrichment of specifically expressed genes | No | Yes | up regulation only | cell type |

Supplementary Table 1: **Comparison between different methods for identifying disease-associated cell populations.** We compare the methods based on: (**a**) how each method defines association between a cell population and disease, (**b**) whether the method directly models the disease, (c) whether the method incorporates GWAS data, (**d**) whether the method assesses contributions of up-/down-regulated genes in cases/controls, and (**e**) resolution of the results (i.e., individual cell vs. cell type).

Supplementary Table 2: **Numerical results of the simulations for assessing the performance of scEPS for individual cell neighborhoods.** (**a**, **b**) Results of simulations, in which 1,000 (default) and 100, respectively, bootstrap samples were used in statistical testing.

See attached Excel file.

Supplementary Table 3: **Numerical results of the simulations for assessing the performance of scEPS in estimating aggregated statistics across cell neighborhoods.** (**a**, **b**, **c**) Results of simulations, in which scEPS statistics were aggregated across 20, 100 (default), and 500, respectively, randomly selected cell neighborhoods. In all simulations, a set of 100 approximately independent cell neighborhood blocks was used in statistical block bootstrap for statistical testing.

See attached Excel file.

Supplementary Table 4: **Numerical results of the simulations for assessing the impact of the number of bootstrap blocks in aggregating scEPS statistics.** (**a**, **b**) Results of simulations, in which 50 and 150, respectively, approximately independent cell neighborhood blocks were used in statistical block bootstrap. In all simulations, scEPS statistics were aggregated across 100 randomly selected cell neighborhoods.

See attached Excel file.

Supplementary Table 5: **Numerical results of the simulations for assessing the impact of model misspecification on the performance of scEPS for individual cell neighborhoods.** Results of simulations, in which the top 600 GWAS genes were used for simulating the phenotype, and the top 300 (**a**) and 900 (**b**) GWAS genes were analyzed by scEPS. (**c**) Results of simulations, in which 50% of the cells in each cell neighborhood were randomly removed.

See attached Excel file.

Supplementary Table 6: **Genetic correlations among the 3 neurological and 3 respiratory disorders.** We report the pairwise genetic correlations for the 3 neurological and 3 respiratory disorders obtained using cross-trait LDSC [146].

See attached Excel file.

Supplementary Table 7: **Aggregated scEPS $\boldsymbol{\omega^2}_{\boldsymbol{overall}}$ statistics across all cells for disease PRSs.** (**a**) Aggregated $\omega^2{}_{overall}$ statistics for PRSs of AD, MS, and PD. (**b**) Aggregated $\omega^2{}_{overall}$ statistics for PRSs of IPF, COPD, and FEV1/FVC. For each disease, we used the PRSs with the highest aggregated $\omega^2{}_{overall}$ (marked with "*") in our main analyses. All analyses involving disease PRSs are restricted to using control donors only.

See attached Excel file.

Supplementary Table 8: **Average size of the cell neighborhoods.** (**a**, **b**) Average size (number of cells) of the cell neighborhoods analyzed by scEPS in the SEA-AD brain cell atlas and TGen lung cell atlas, respectively.

See attached Excel file.

Supplementary Table 9: **Aggregated scEPS statistics of 4 neurological disorders.** (**a**, **c**, **e**, **g**) Aggregated scEPS statistics across all cell neighborhoods, for CS, and PRSs of AD, MS and PD, respectively. (**b**, **d**, **f**, **h**) Aggregated scEPS statistics across cell neighborhoods for 139 brain cell subtypes, for CS, and PRSs of AD, MS and PD, respectively.

See attached Excel file.

Supplementary Table 10: **Aggregated scEPS statistics of 4 respiratory disorders.** (**a**, **c**, **e**, **g**) Aggregated scEPS statistics across all cell neighborhoods, for IPF, and PRSs of IPF, COPD and FEV1/FVC, respectively. (**b**, **d**, **f**, **h**) Aggregated scEPS statistics across cell neighborhoods for 43 lung cell subtypes, for IPF, and PRSs of IPF, COPD and FEV1/FVC, respectively.

See attached Excel file.

Supplementary Table 11: **scEPS statistics of 4 neurological disorders obtained using pseudo-bulked gene expression of brain cell types.** (**a**, **b**, **c**, **d**) Results for CS, and PRSs of AD, MS and PD, respectively. All analyses involving disease PRSs are restricted to using control donors only.

See attached Excel file.

Supplementary Table 12: **scEPS statistics of 4 respiratory disorders obtained using pseudo-bulked gene expression of lung cell types.** (**a**, **b**, **c**, **d**) Results for IPF, and PRSs of IPF, COPD and FEV1/FVC, respectively. All analyses involving disease PRSs are restricted to using control donors only.

See attached Excel file.

Supplementary Table 13: **Number of significant (FDR < 0.05) disease-cell-type associations identified using neighborhood vs. pseudo-bulk based approaches.** Number of significant disease-associated cell types identified using neighborhood vs. pseudo-bulk approaches for the 4 neurological (**a**) and 4 respiratory (**b**) disorders, respectively.

See attached Excel file.

Supplementary Table 14: **Aggregated scEPS statistics of 4 neurological disorders obtained with COPD GWAS.** (**a**, **b**, **c**, **d**) Results for CS, and PRSs of AD, MS and PD, respectively. All analyses involving disease PRSs are restricted to using control donors only.

See attached Excel file.

Supplementary Table 15: **Aggregated scEPS statistics of 4 respiratory disorders obtained with AD GWAS.** (**a**, **b**, **c**, **d**) Results for IPF, and PRSs of IPF, COPD and FEV1/FVC, respectively. All analyses involving disease PRSs are restricted to using control donors only.

See attached Excel file.

Supplementary Table 16: **Aggregated scEPS $d$ statistics obtained with matched vs. mismatched GWASs of 4 respiratory and 4 neurological disorders.** (**a**, **b**) Results for the 4 neurological and 4 respiratory disorders, respectively. P-values testing the difference between aggregated $d$ across analyses using matched vs. mismatched GWAS were obtained based on 1,000 permutations.

See attached Excel file.

Supplementary Table 17: **Gene set enrichment analysis of top AD and COPD GWAS genes analyzed by scEPS.** (**a**) Reactome pathways implicated using the top 570 AD GWAS genes analyzed by scEPS. (**b**) Reactome pathways implicated using the top 845 COPD GWAS genes analyzed by scEPS. "Enrichment Score (ES)" quantifies the degree of overrepresentation of pathway genes in the ranked gene list. "Normalized Enrichment Score (NES)" normalizes the raw ES based on the ES of pathway gene sets of similar sizes. "Tag %" and "Gene %" represent the percentages of genes from the pathway gene set and entire gene set, respectively, encountered before the enrichment score reaches maximum deviation from 0.

See attached Excel file.

Supplementary Table 18: **Aggregated scEPS statistics of CS using ROSMAP brain cell snRNA-seq data.** (**a**) Aggregated scEPS statistics across all cell neighborhoods. (**b**) Aggregated scEPS statistics across 71 brain cell subtypes.

See attached Excel file.

Supplementary Table 19: **Reactome pathways implicated via GSEA of genes correlated with scEPS $d$ statistics for 4 neurological disorders.** (**a**, **b**, **c**, **d**) Reactome pathways implicated via global GSEA for CS, AD PRS, MS PRS, and PD PRS, respectively. (**e**, **f**, **g**, **h**) Reactome pathways implicated via cell-type level GSEA for CS, AD PRS, MS PRS, and PD PRS, respectively. "Enrichment Score (ES)" quantifies the degree of overrepresentation of pathway genes in the ranked gene list. "Normalized Enrichment Score (NES)" normalizes the raw ES based on the ES of pathway gene sets of similar sizes. "Tag %" and "Gene %" represent the percentages of genes from the pathway gene set and entire gene set, respectively, encountered before the enrichment score reaches maximum deviation from 0.

See attached Excel file.

Supplementary Table 20: **Reactome pathways implicated via GSEA of genes correlated with scEPS $d$ statistics for 4 respiratory disorders.** (**a**, **b**, **c**, **d**) Reactome pathways implicated via global GSEA for CS, AD PRS, MS PRS, and PD PRS, respectively. (**e**, **f**, **g**, **h**) Reactome pathways implicated via cell-type level GSEA for CS, AD PRS, MS PRS, and PD PRS, respectively. "Enrichment Score (ES)" quantifies the degree of overrepresentation of pathway genes in the ranked gene list. "Normalized Enrichment Score (NES)" normalizes the raw ES based on the ES of pathway gene sets of similar sizes. "Tag %" and "Gene %" represent the percentages of genes from the pathway gene set and entire gene set, respectively, encountered before the enrichment score reaches maximum deviation from 0.

See attached Excel file.

Supplementary Table 21: **Aggregated CNA* $\sigma^2_{na}$ across cell neighborhoods of 139 brain cell subtypes for 4 neurological disorders.** (**a**, **b**, **c**, **d**) Results for CS, and PRSs of AD, MS, and PD, respectively. All analyses involving disease PRSs are restricted to using control donors only.

See attached Excel file.

Supplementary Table 22: **Aggregated CNA* $\sigma^2_{na}$ across cell neighborhoods of 43 lung cell subtypes for 4 respiratory disorders.** (**a**, **b**, **c**, **d**) Results for IPF, and PRSs of IPF, COPD, and FEV1/FVC, respectively. All analyses involving disease PRSs are restricted to using control donors only.

See attached Excel file.

Supplementary Table 23: **Aggregated scDRS raw scores across cells of 139 brain cell subtypes for 4 neurological disorders.** (**a**) Results for AD using cells from all donors. (**b**, **c**, **d**) Results for AD, MS, and PD, respectively, using cells from control donors only.

See attached Excel file.

Supplementary Table 24: **Aggregated scDRS raw scores across cells of 43 lung cell subtypes for 4 respiratory disorders.** (**a**) Results for IPF using cells from all donors. (**b**, **c**, **d**) Results for IPF, COPD, and FEV1/FVC, respectively, using cells from control donors only.

See attached Excel file.

### Supplementary Figures

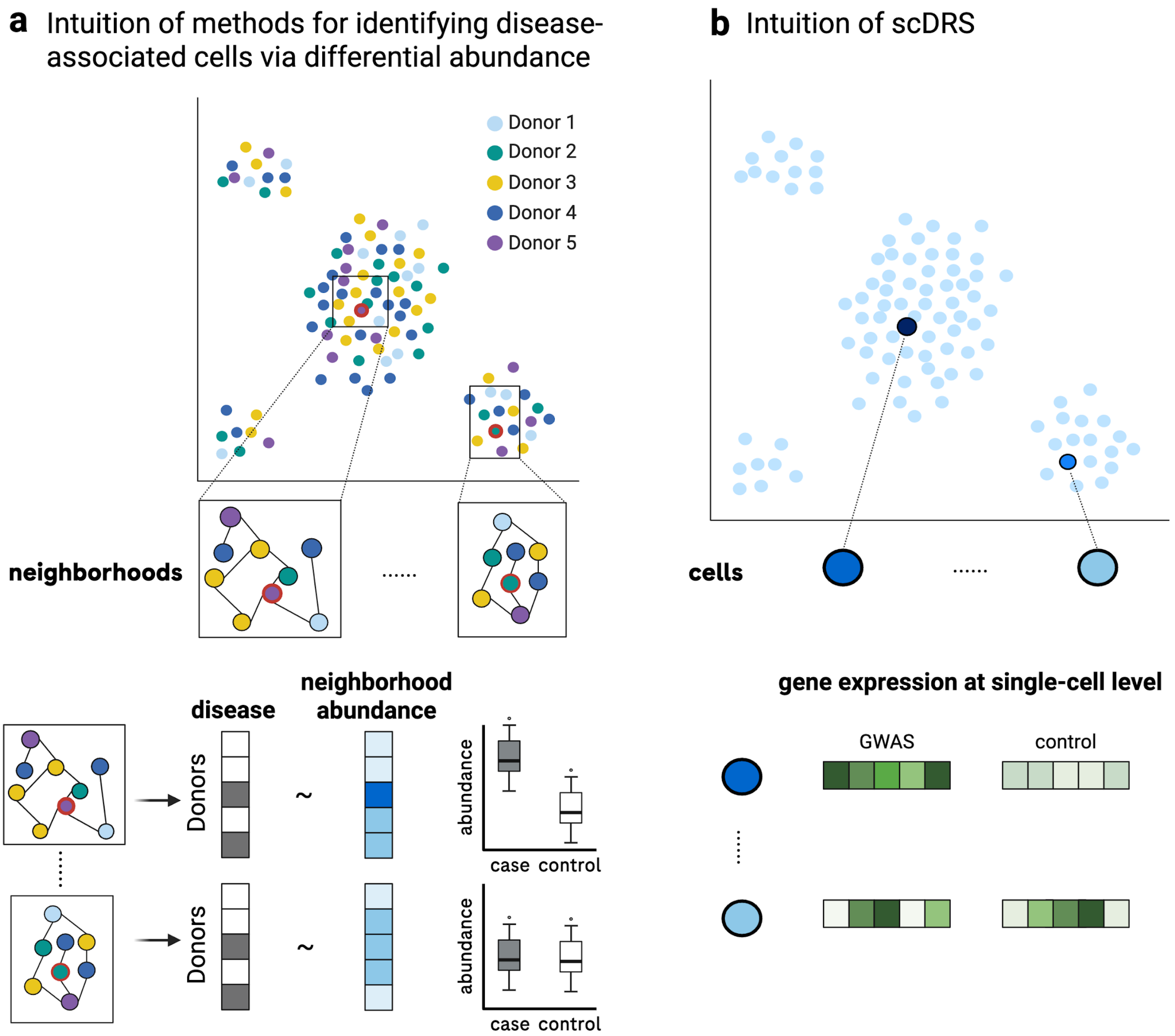


Supplementary Figure 1: **Intuition of neighborhood-resolution differential abundance methods and scDRS.** (**a**) Intuition of methods (e.g., CNA, Milo) that identify disease-associated cell populations by testing differential abundance at cell neighborhood resolution. These methods first define cell neighborhoods based on the k-NN graph for a set of cells. These methods then test for differential cell abundance across cases vs. controls in each cell neighborhood. Both CNA and Milo do not incorporate GWAS genes. (**b**) Intuition of scDRS. scDRS identifies disease-associated cell populations, by testing for whether disease GWAS genes are specifically expressed in individual cells. scDRS determines that a cell is associated with a disease, if the expression of GWAS genes is significantly higher than a set of randomly selected mean and variance matched control genes. scDRS does not directly model the disease outcome.

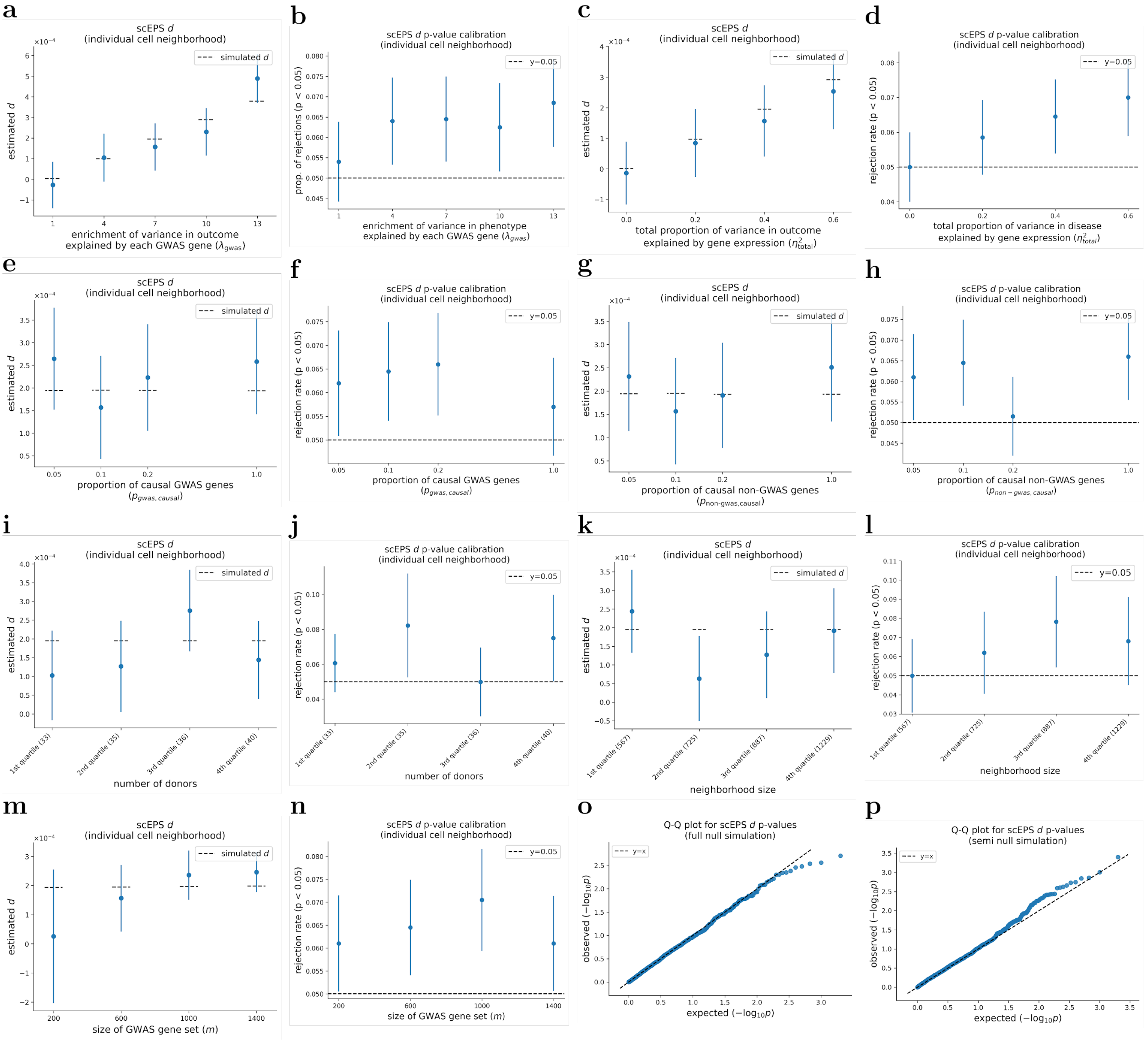


Supplementary Figure 2: **Performance of scEPS in estimating $d$ statistics for individual cell neighborhoods.** We report the average estimated $d$ and proportions of rejected null hypotheses, respectively, across simulated $\lambda^2{}_{gwas}$ in (**a**, **b**), across simulated $\eta^2{}_{total}$ in (**c**, **d**), across $p_{gwas,causal}$ in (**e**, **f**), across $p_{non-gwas,causal}$ in (**g**, **h**), across quartiles of number of donors in the neighborhoods in (**i**, **j**), across quartiles of neighborhood sizes in (**k**, **l**), and across sizes of GWAS gene sets in (**m**, **n**). (**o**, **p**) Q-Q plots showing the expected vs. observed -$\log_{10}$ p-values for $d$ across 2,000 full null ($\eta^2{}_{total} = 0$) and semi null ($\eta^2{}_{total} = 0.4$, $\lambda_{gwas} = 1.0$) simulations, respectively. In simulations where one parameter was varied, all other parameters were fixed at their default values, with $\lambda_{gwas} = 7.0$, $\eta^2{}_{total} = 0.4$, $m_{sim} = 600$, and $p_{gwas,causal} = p_{non-gwas,causal} = 0.05$. Mean and standard errors were obtained based on 2,000 simulations. Error bars represent 1.96× standard errors on both times. Numerical results are reported in Supplementary Table 2.

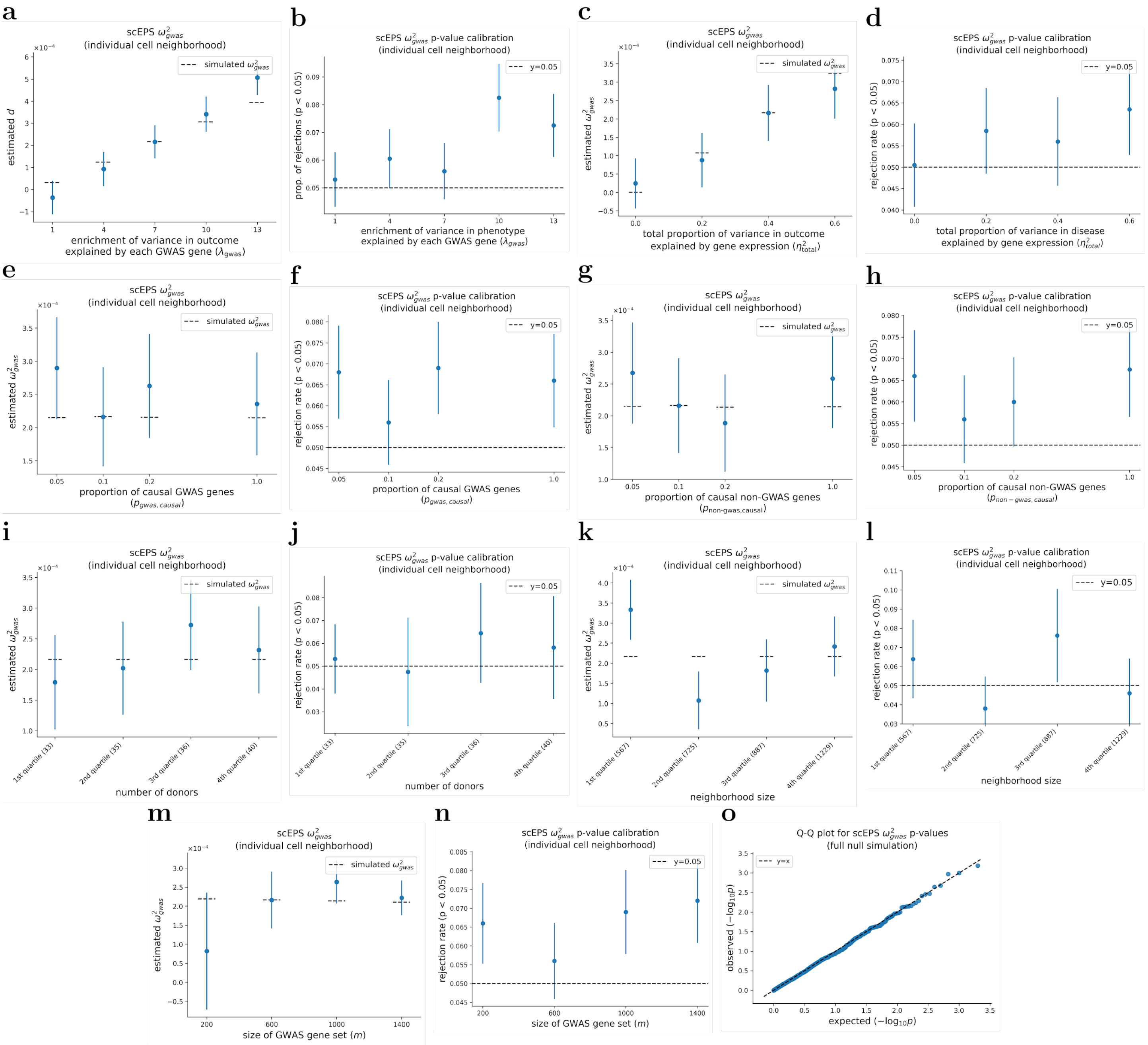


Supplementary Figure 3: **Performance of scEPS in estimating $\omega^2{}_{gwas}$ statistics for individual cell neighborhoods.** We report the average estimated $\omega^2{}_{gwas}$ and proportions of rejected null hypotheses, respectively, across simulated $\lambda^2{}_{gwas}$ in (**a**, **b**), across simulated $\eta^2{}_{total}$ in (**c**, **d**), across $p_{gwas,causal}$ in (**e**, **f**), across $p_{non-gwas,causal}$ in (**g**, **h**), across quartiles of numbers of donors in the neighborhoods in (**i**, **j**), across quartiles of neighborhood sizes in (**k**, **l**), and across sizes of GWAS gene sets in (**m**, **n**). (**o**) Q-Q plot showing the expected vs. observed -$\log_{10}$ p-values for $\omega^2{}_{gwsa}$ across 2,000 full null simulations ($\eta^2{}_{total} = 0$). In simulations where one parameter was varied, the other parameters were fixed at their default values, with $\lambda_{gwas} = 7.0$, $\eta^2{}_{total} = 0.4$, $m_{sim} = 600$, and $p_{gwas,causal} = p_{non-gwas,causal} = 0.05$. Mean and standard errors were obtained based on 2,000 simulations. Error bars represent 1.96× standard errors on both times. Numerical results are reported in Supplementary Table 2.

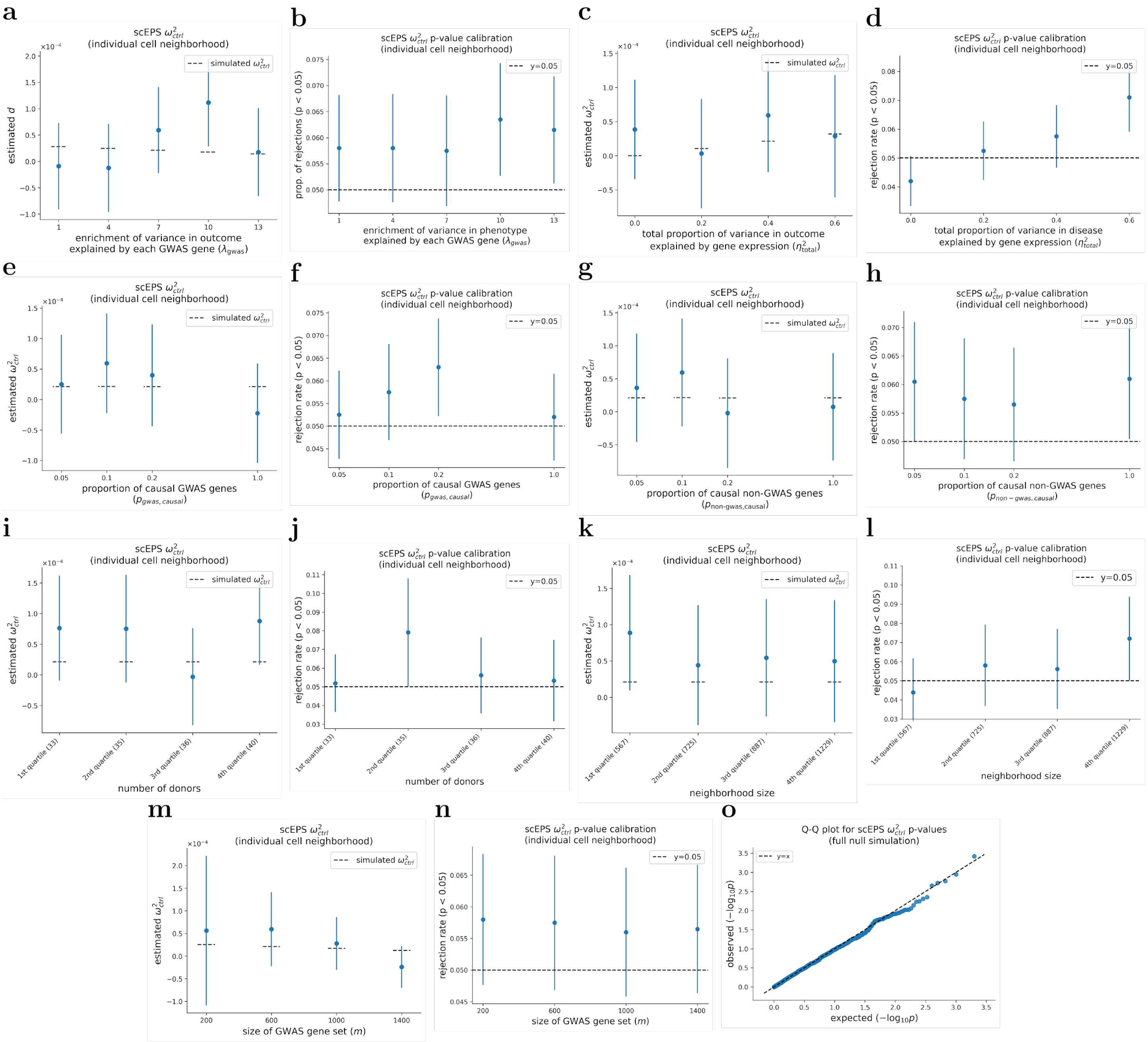


Supplementary Figure 4: **Performance of scEPS in estimating $\omega^2{}_{ctrl}$ statistics for individual cell neighborhoods.** We report the average estimated $\omega^2{}_{ctrl}$ and proportions of rejected null hypotheses, respectively, across simulated $\lambda^2{}_{gwas}$ in (**a**, **b**), across simulated $\eta^2{}_{total}$ in (**c**, **d**), across $p_{gwas,causal}$ in (**e**, **f**), across $p_{non-gwas,causal}$ in (**g**, **h**), across quartiles of numbers of donors in the neighborhoods in (**i**, **j**), across quartiles of neighborhood sizes in (**k**, **l**), and across sizes of GWAS gene sets in (**m**, **n**). (**o**) Q-Q plot showing the expected vs. observed -log$_{10}$ p-values for $\omega^2{}_{ctrl}$ across 2,000 full null simulations ($\eta^2{}_{total} = 0$). In simulations where one parameter was varied, the other parameters were fixed at their default values, with $\lambda_{gwas} = 7.0$, $\eta^2{}_{total} = 0.4$, $m_{sim} = 600$, and $p_{gwas,causal} = p_{non-gwas,causal} = 0.05$. Mean and standard errors were obtained based on 2,000 simulations. Error bars represent 1.96×standard errors on both times. Numerical results are reported in Supplementary Table 2.

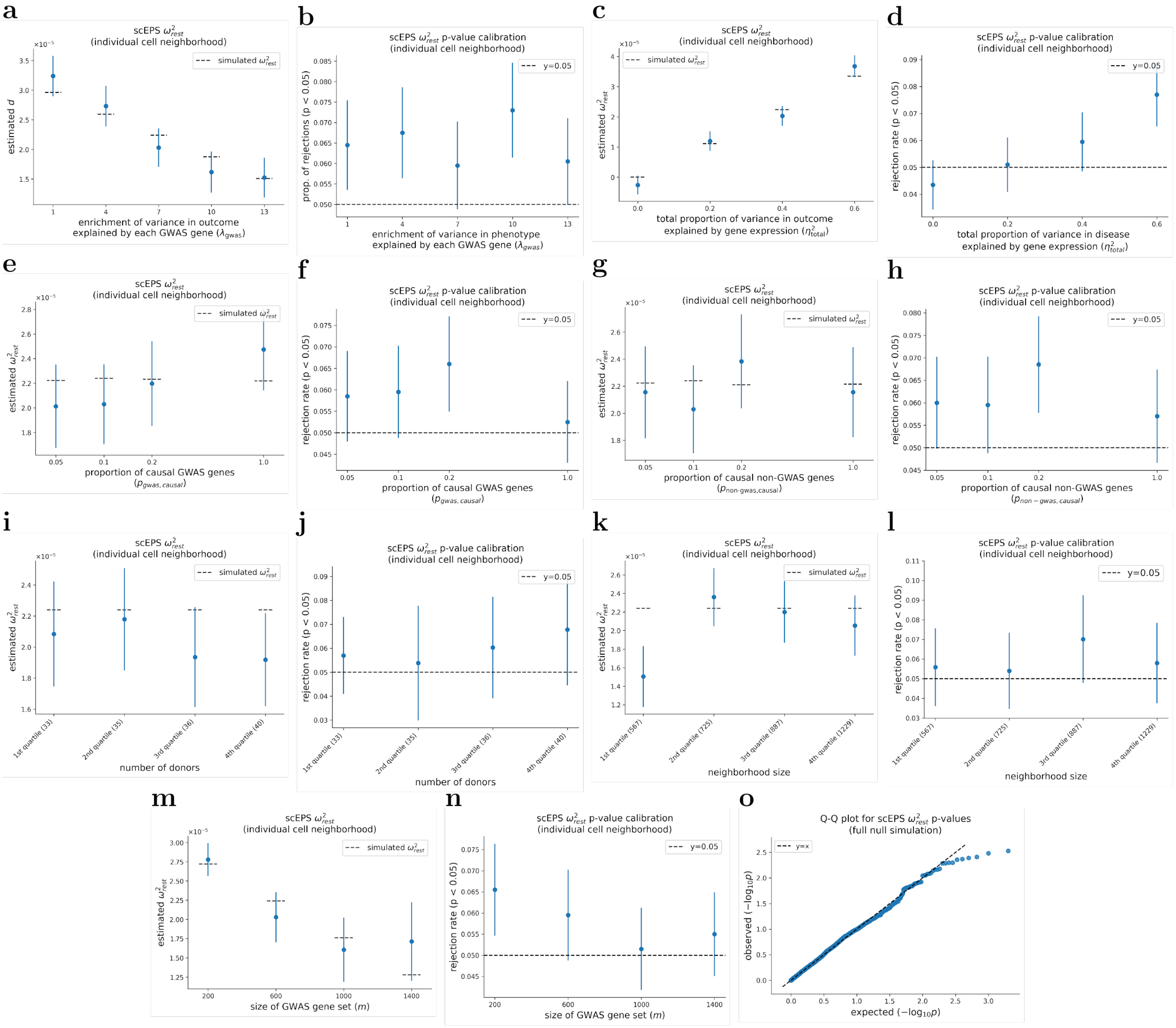


Supplementary Figure 5: **Performance of scEPS in estimating $\omega^2{}_{rest}$ statistics for individual cell neighborhoods.** We report the average estimated $\omega^2{}_{rest}$ and proportions of rejected null hypotheses, respectively, across simulated $\lambda^2{}_{gwas}$ in (**a**, **b**), across simulated $\eta^2{}_{total}$ in (**c**, **d**), across $p_{gwas,causal}$ in (**e**, **f**), across $p_{non-gwas,causal}$ in (**g**, **h**), across quartiles of numbers of donors in the neighborhoods in (**i**, **j**), across quartiles of neighborhood sizes in (**k**, **l**), and across sizes of GWAS gene sets in (**m**, **n**). (**o**) Q-Q plot showing the expected vs. observed -log$_{10}$ p-values for $\omega^2{}_{rest}$ across 2,000 full null simulations ($\eta^2{}_{total} = 0$). In simulations where one parameter was varied, the other parameters were fixed at their default values, with $\lambda_{gwas} = 7.0$, $\eta^2{}_{total} = 0.4$, $m_{sim} = 600$, and $p_{gwas,causal} = p_{non-gwas,causal} = 0.05$. Mean and standard errors were obtained based on 2,000 simulations. Error bars represent 1.96× standard errors on both times. Numerical results are reported in Supplementary Table 2.

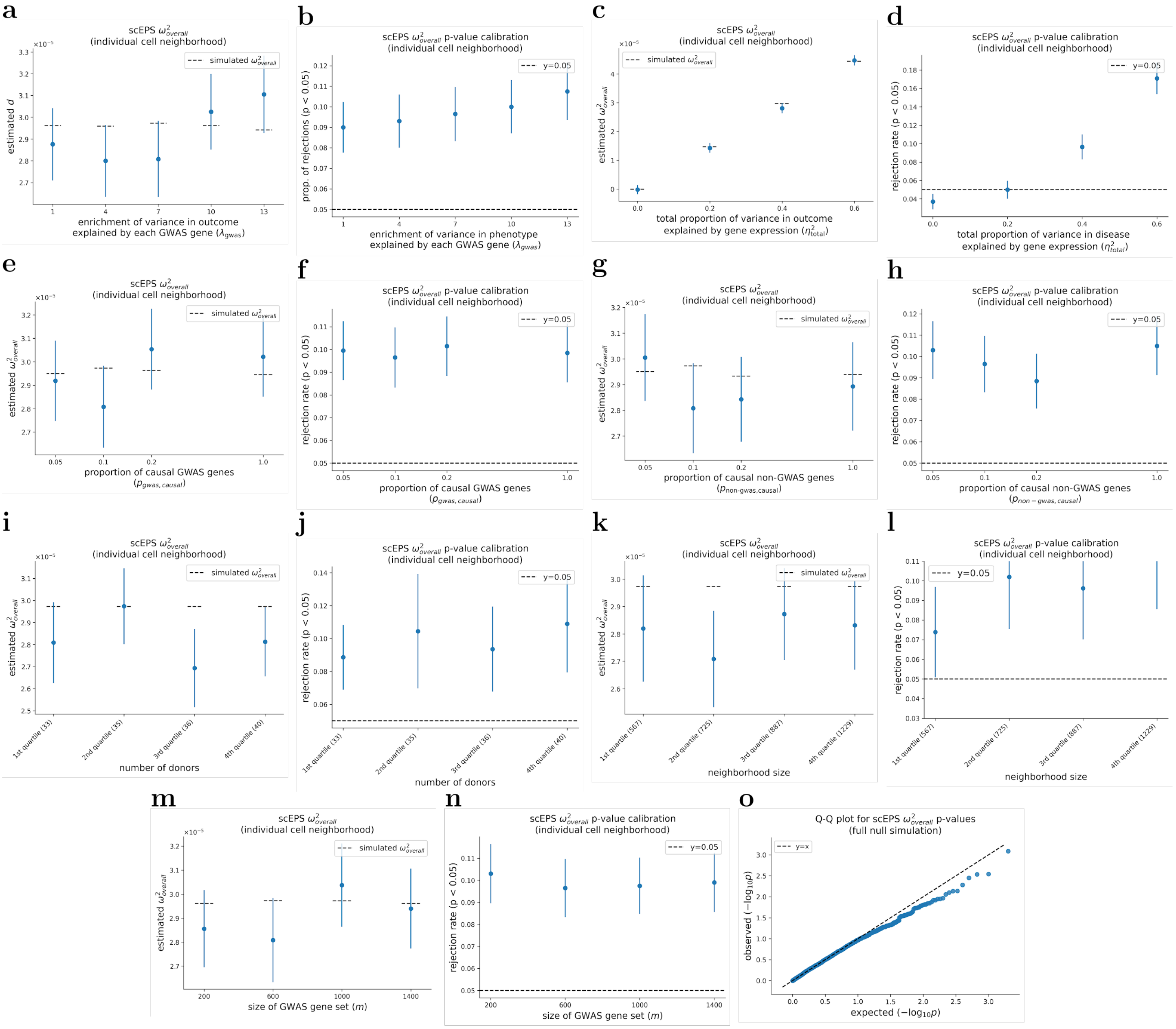


Supplementary Figure 6: **Performance of scEPS in estimating $\omega^2_{overall}$ statistics for individual cell neighborhoods.** We report the average estimated $\omega^2_{overall}$ and proportions of rejected null hypotheses, respectively, across simulated $\lambda^2_{gwas}$ in (**a**, **b**), across simulated $\eta^2_{total}$ in (**c**, **d**), across $p_{gwas,causal}$ in (**e**, **f**), across $p_{non-gwas,causal}$ in (**g**, **h**), across quartiles of numbers of donors in the neighborhoods in (**i**, **j**), across quartiles of neighborhood sizes in (**k**, **l**), and across sizes of GWAS gene sets in (**m**, **n**). (**o**) Q-Q plot showing the expected vs. observed -log$_{10}$ p-values for $\omega^2_{overall}$ across 2,000 full null simulations ($\eta^2_{total} = 0$). In simulations where one parameter was varied, the other parameters were fixed at their default values, with $\lambda_{gwas} = 7.0$, $\eta^2_{total} = 0.4$, $m_{sim} = 600$, and $p_{gwas,causal} = p_{non-gwas,causal} = 0.05$. Mean and standard errors were obtained based on 2,000 simulations. Error bars represent 1.96× standard errors on both times. Numerical results are reported in Supplementary Table 2.

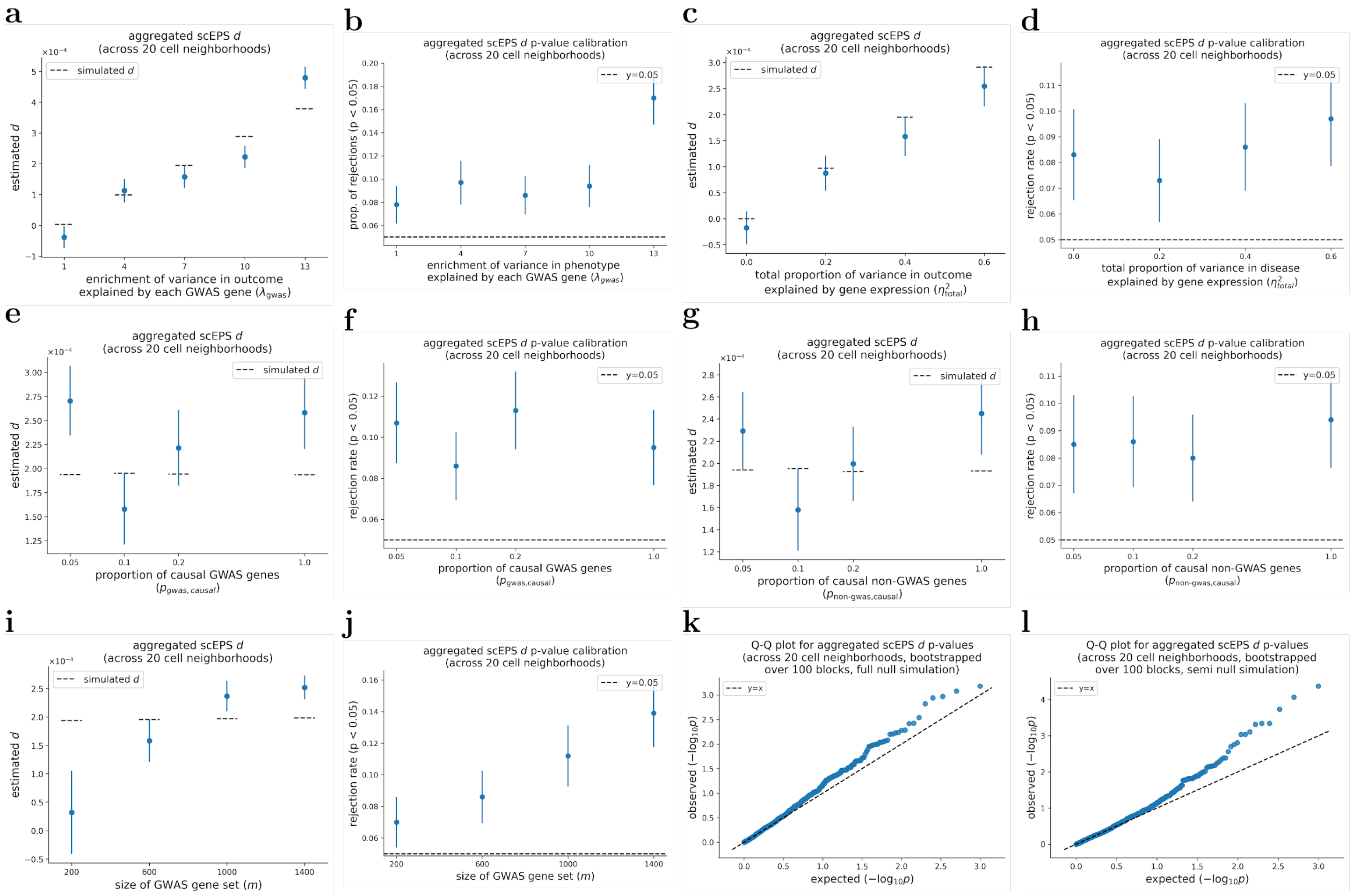


Supplementary Figure 7: **Performance of scEPS in estimating aggregated $d$ across 20 cell neighborhoods.** We report the average estimates of aggregated $d$ across 20 cell neighborhoods and proportions of rejected null hypotheses, respectively, across simulated $\lambda^2{}_{gwas}$ in (**a**, **b**), across simulated $\eta^2{}_{total}$ in (**c**, **d**), across $p_{gwas,causal}$ in (**e**, **f**), across $p_{non-gwas,causal}$ in (**g**, **h**), and across sizes of GWAS gene sets in (**i**, **j**). (**k**, **l**) Q-Q plots showing the expected vs. observed -log$_{10}$ p-values for aggregated $d$ across 1,000 full null ($\eta^2{}_{total} = 0$) and semi null ($\eta^2{}_{total} = 0.4$, $\lambda_{gwas} = 1.0$) simulations, respectively. In simulations where one parameter was varied, the other parameters were fixed at their default values, with $\lambda_{gwas} = 7.0$, $\eta^2{}_{total} = 0.4$, $m_{sim} = 600$, and $p_{gwas,causal} = p_{non-gwas,causal} = 0.05$. Mean and standard errors were obtained based on 1,000 simulations. P-values testing the statistical significance of aggregated $d$ was obtained by bootstrapping over 100 approximately independent cell neighborhood blocks**.** Error bars represent 1.96× standard errors on both times. Numerical results are reported in Supplementary Table 3.

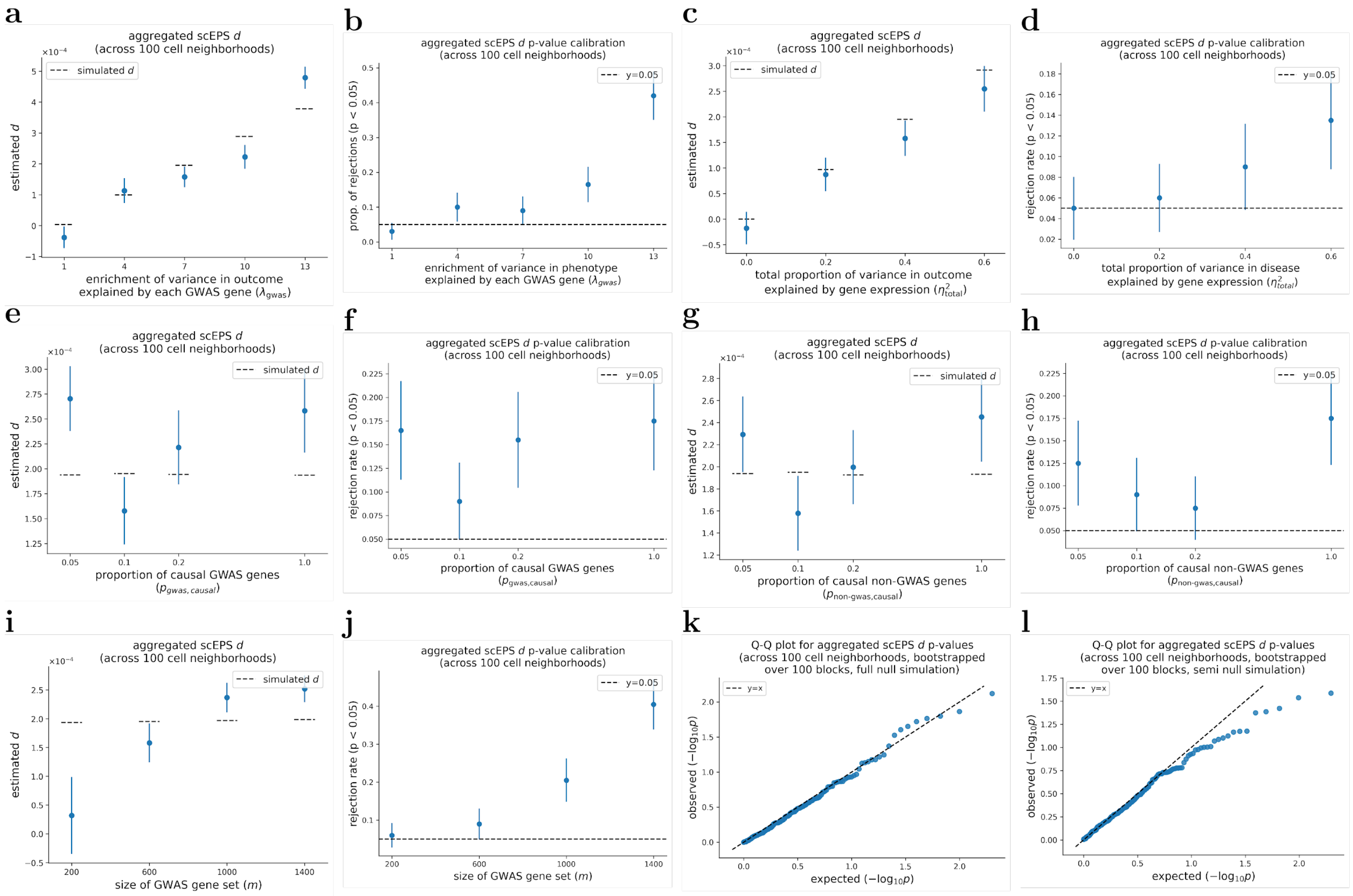


Supplementary Figure 8: **Performance of scEPS in estimating aggregated $d$ statistics across 100 cell neighborhoods.** We report the average estimates of aggregated $d$ across 100 cell neighborhoods and proportions of rejected null hypotheses, respectively, across simulated $\lambda^2{}_{gwas}$ in (**a**, **b**), across simulated $\eta^2{}_{total}$ in (**c**, **d**), across $p_{gwas,causal}$ in (**e**, **f**), across $p_{non-gwas,causal}$ in (**g**, **h**), and across sizes of GWAS gene sets in (**i**, **j**). (**k**, **l**) Q-Q plots showing the expected vs. observed -$\log_{10}$ p-values for aggregated $d$ across 200 full null ($\eta^2{}_{total} = 0$) and semi null ($\eta^2{}_{total} = 0.4$, $\lambda_{gwas} = 1.0$) simulations, respectively. In simulations where one parameter was varied, the other parameters were fixed at their default values, with $\lambda_{gwas} = 7.0$, $\eta^2{}_{total} = 0.4$, $m_{sim} = 600$, and $p_{gwas,causal} = p_{non-gwas,causal} = 0.05$. Mean and standard errors were obtained based on 200 simulations. P-values testing the statistical significance of aggregated $d$ was obtained by bootstrapping over 100 approximately independent cell neighborhood blocks**.** Error bars represent 1.96× standard errors on both times. Numerical results are reported in Supplementary Table 3.

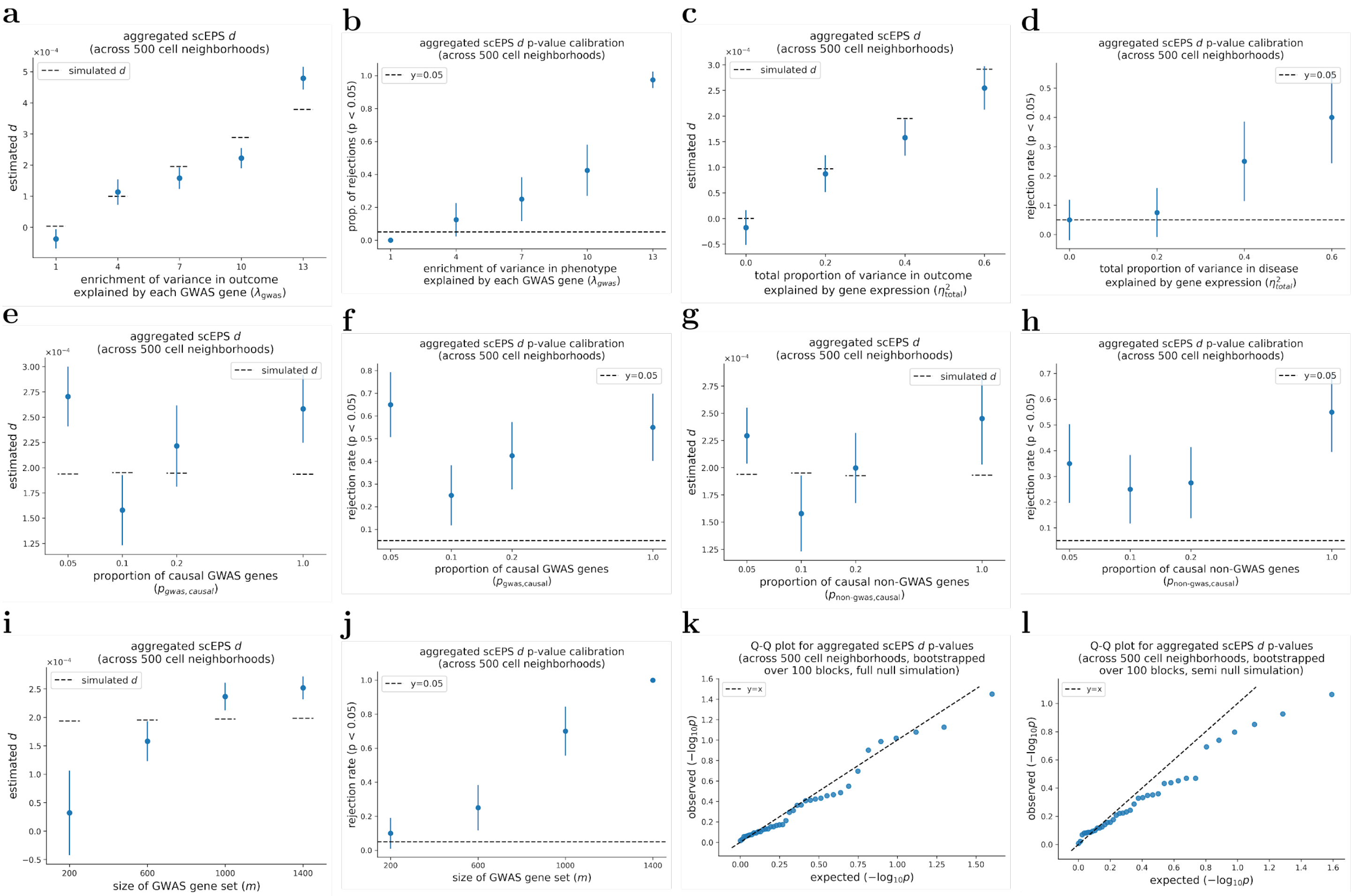


Supplementary Figure 9: **Performance of scEPS in estimating aggregated $d$ statistics across 500 cell neighborhoods.** We report the average estimates of aggregated $d$ across 500 cell neighborhoods and proportions of rejected null hypotheses, respectively, across simulated $\lambda^{2}{}_{gwas}$ in (**a**, **b**), across simulated $\eta^{2}{}_{total}$ in (**c**, **d**), across $p_{gwas,causal}$ in (**e**, **f**), across $p_{non-gwas,causal}$ in (**g**, **h**), and across sizes of GWAS gene sets in (**i**, **j**). (**k**, **l**) Q-Q plots showing the expected vs. observed -log$_{10}$ p-values for aggregated $d$ across 40 full null ($\eta^{2}{}_{total} = 0$) and semi null ($\eta^{2}{}_{total} = 0.4$, $\lambda_{gwas} = 1.0$) simulations, respectively. In simulations where one parameter was varied, the other parameters were fixed at their default values, with $\lambda_{gwas} = 7.0$, $\eta^{2}{}_{total} = 0.4$, $m_{sim} = 600$, and $p_{gwas,causal} = p_{non-gwas,causal} = 0.05$. Mean and standard errors were obtained based on 40 simulations. P-values testing the statistical significance of aggregated $d$ was obtained by bootstrapping over 100 approximately independent cell neighborhood blocks**.** Error bars represent 1.96× standard errors on both times. Numerical results are reported in Supplementary Table 3.

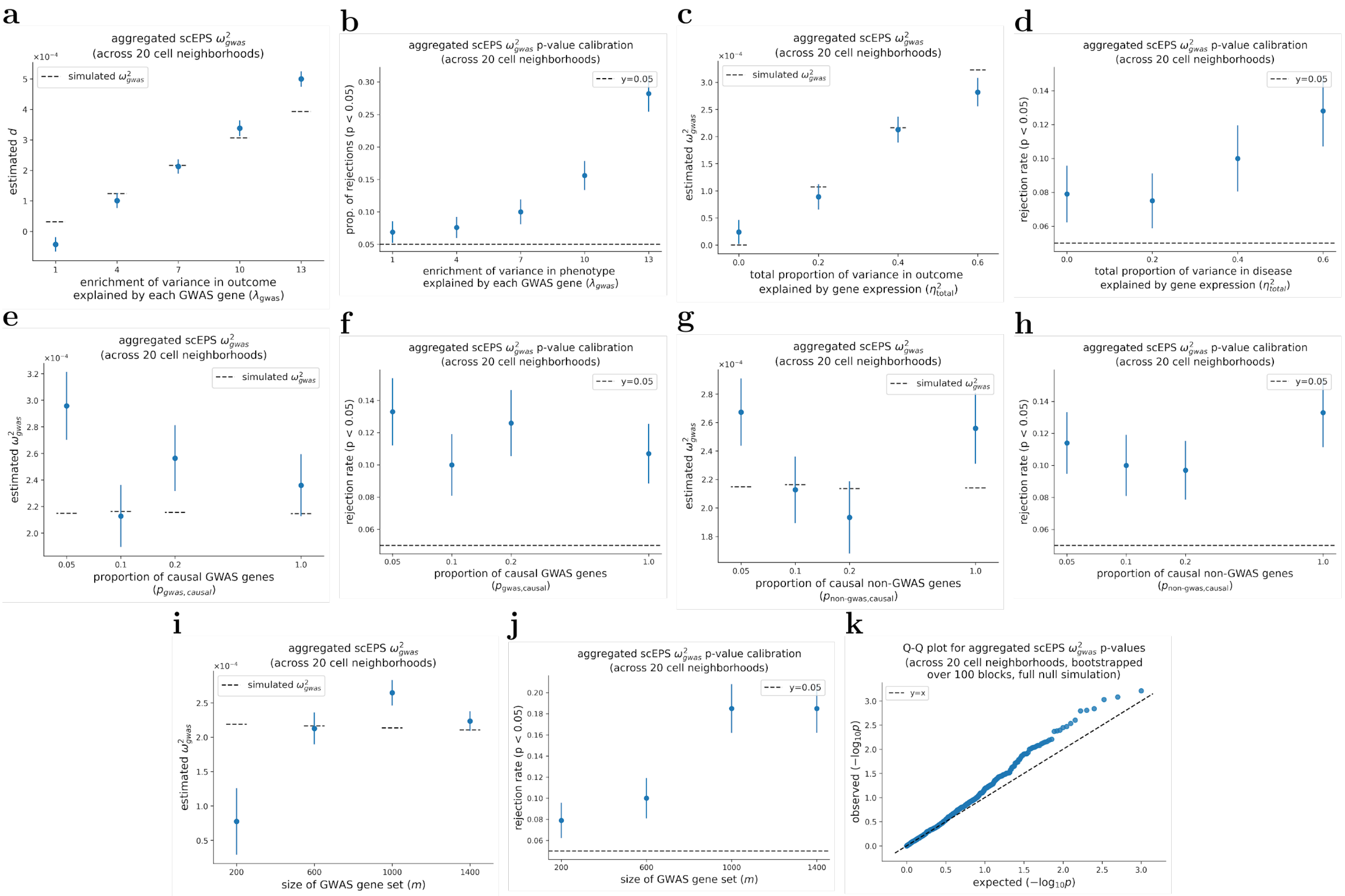


Supplementary Figure 10: **Performance of scEPS in estimating aggregated $\omega^2{}_{gwas}$ statistics across 20 cell neighborhoods.** We report the average estimates of aggregated $\omega^2{}_{gwas}$ across 20 cell neighborhoods and proportions of rejected null hypotheses, respectively, across simulated $\lambda^2{}_{gwas}$ in (**a**, **b**), across simulated $\eta^2{}_{total}$ in (**c**, **d**), across $p_{gwas,causal}$ in (**e**, **f**), across $p_{non-gwas,causal}$ in (**g**, **h**), and across sizes of GWAS gene sets in (**i**, **j**). (**k**) Q-Q plots showing the expected vs. observed -$\log_{10}$ p-values for aggregated $\omega^2{}_{gwas}$ across 1,000 full null ($\eta^2{}_{total} = 0$) simulations. In simulations where one parameter was varied, the other parameters were fixed at their default values, with $\lambda_{gwas} = 7.0$, $\eta^2{}_{total} = 0.4$, $m_{sim} = 600$, and $p_{gwas,causal} = p_{non-gwas,causal} = 0.05$. Mean and standard errors were obtained based on 1,000 simulations. P-values testing the statistical significance of aggregated $\omega^2{}_{gwas}$ was obtained by bootstrapping over 100 approximately independent cell neighborhood blocks. Error bars represent 1.96× standard errors on both times. Numerical results are reported in Supplementary Table 3.

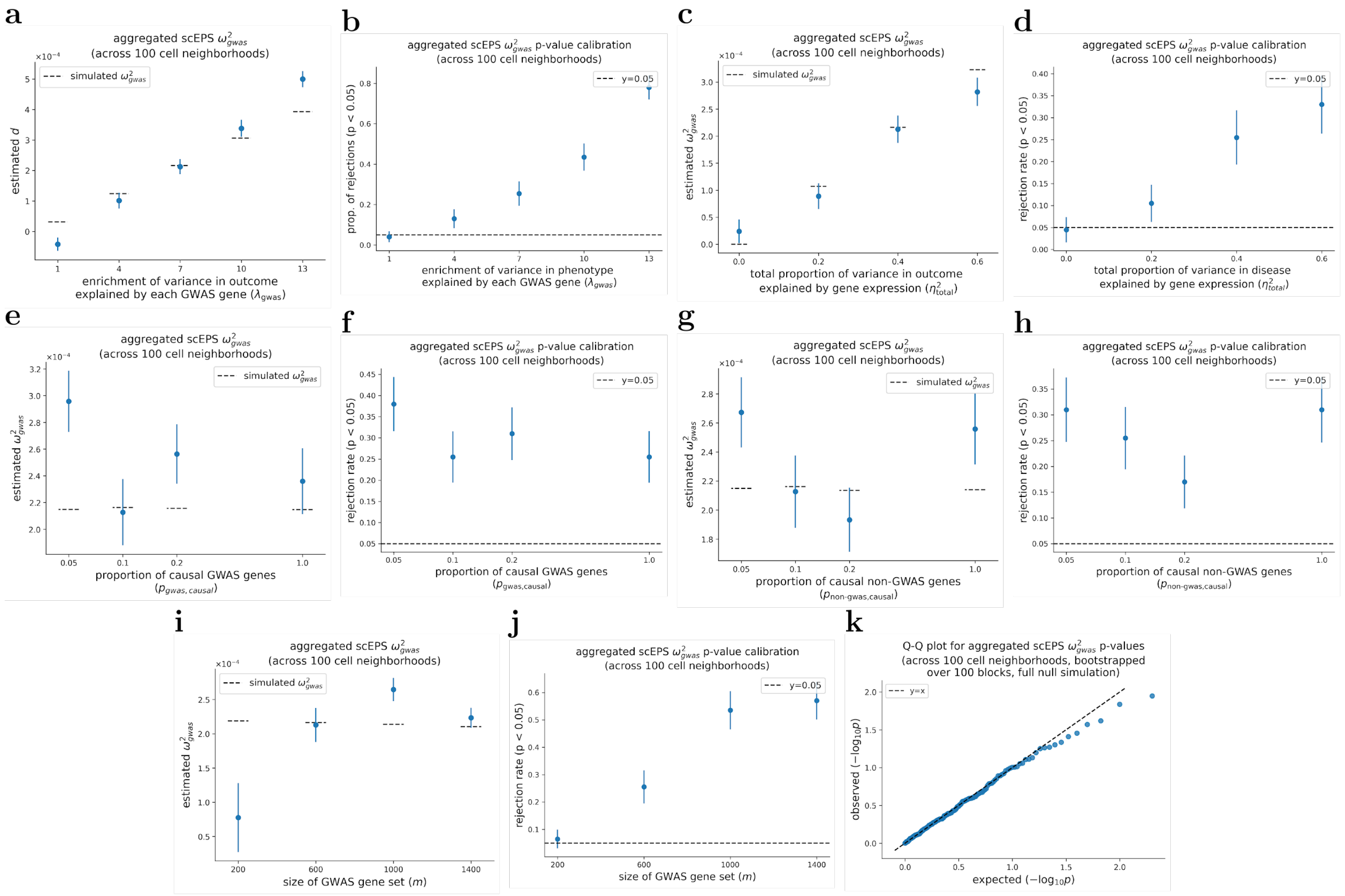


Supplementary Figure 11: **Performance of scEPS in estimating aggregated $\omega^2_{gwas}$ statistics across 100 cell neighborhoods.** We report the average estimates of aggregated $\omega^2_{gwas}$ across 100 cell neighborhoods and proportions of rejected null hypotheses, respectively, across simulated $\lambda^2_{gwas}$ in (**a**, **b**), across simulated $\eta^2_{total}$ in (**c**, **d**), across $p_{gwas,causal}$ in (**e**, **f**), across $p_{non-gwas,causal}$ in (**g**, **h**), and across sizes of GWAS gene sets in (**i**, **j**). (**k**) Q-Q plots showing the expected vs. observed -log$_{10}$ p-values for aggregated $\omega^2_{gwas}$ across 200 full null ($\eta^2_{total} = 0$) simulations. In simulations where one parameter was varied, the other parameters were fixed at their default values, with $\lambda_{gwas} = 7.0$, $\eta^2_{total} = 0.4$, $m_{sim} = 600$, and $p_{gwas,causal} = p_{non-gwas,causal} = 0.05$. Mean and standard errors were obtained based on 200 simulations. P-values testing the statistical significance of aggregated $\omega^2_{gwas}$ was obtained by bootstrapping over 100 approximately independent cell neighborhood blocks. Error bars represent 1.96× standard errors on both times. Numerical results are reported in Supplementary Table 3.

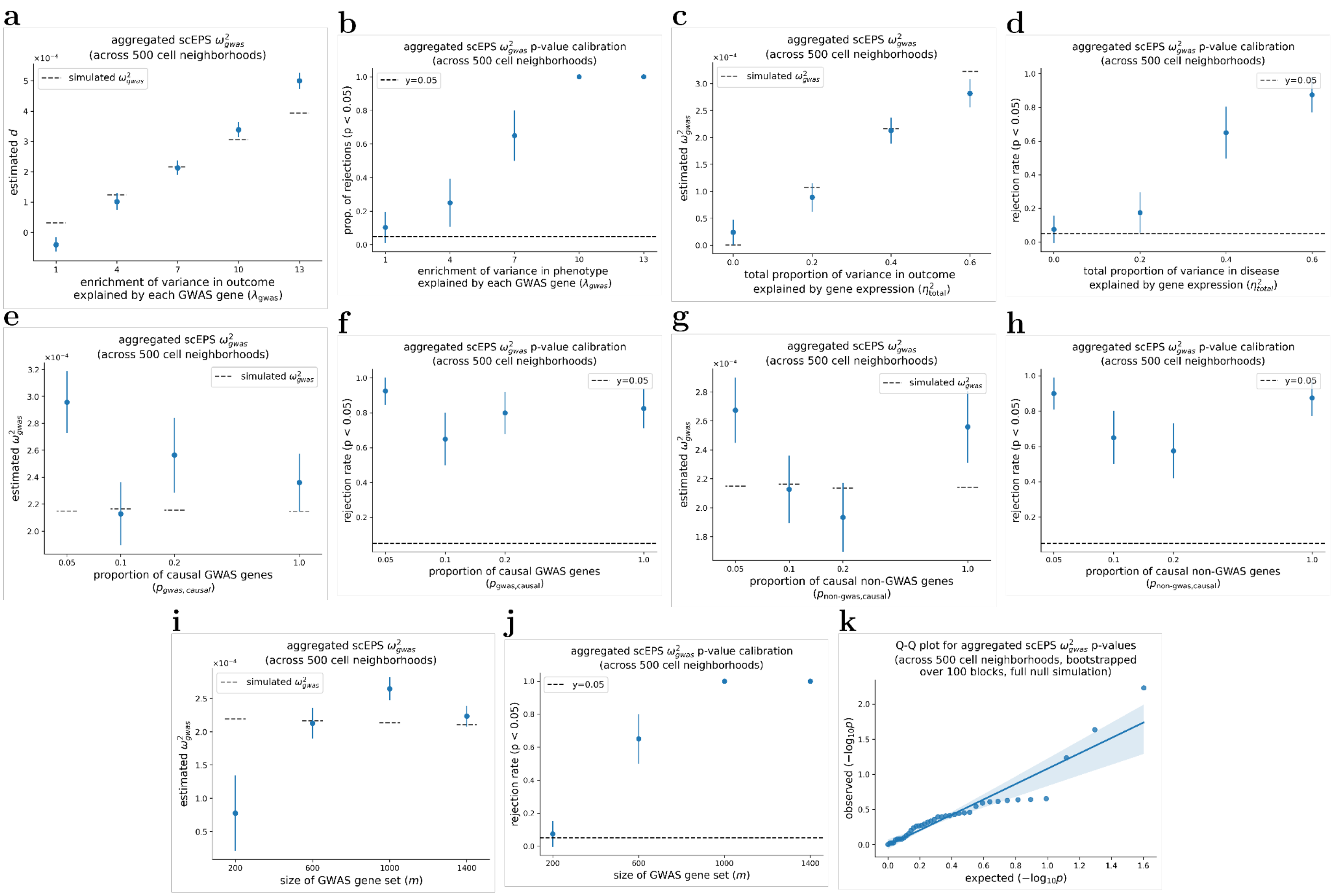


Supplementary Figure 12: **Performance of scEPS in estimating aggregated $\omega^2{}_{gwas}$ statistics across 500 cell neighborhoods.** We report the average estimates of aggregated $\omega^2{}_{gwas}$ across 500 cell neighborhoods and proportions of rejected null hypotheses, respectively, across simulated $\lambda^2{}_{gwas}$ in (**a**, **b**), across simulated $\eta^2{}_{total}$ in (**c**, **d**), across $p_{gwas,causal}$ in (**e**, **f**), across $p_{non-gwas,causal}$ in (**g**, **h**), and across sizes of GWAS gene sets in (**i**, **j**). (**k**) Q-Q plots showing the expected vs. observed -log$_{10}$ p-values for aggregated $\omega^2{}_{gwas}$ across 40 full null ($\eta^2{}_{total} = 0$) simulations. In simulations where one parameter was varied, the other parameters were fixed at their default values, with $\lambda_{gwas} = 7.0$, $\eta^2{}_{total} = 0.4$, $m_{sim} = 600$, and $p_{gwas,causal} = p_{non-gwas,causal} = 0.05$. Mean and standard errors were obtained based on 40 simulations. P-values testing the statistical significance of aggregated $\omega^2{}_{gwas}$ was obtained by bootstrapping over 100 approximately independent cell neighborhood blocks**.** Error bars represent 1.96× standard errors on both times. Numerical results are reported in Supplementary Table 3.

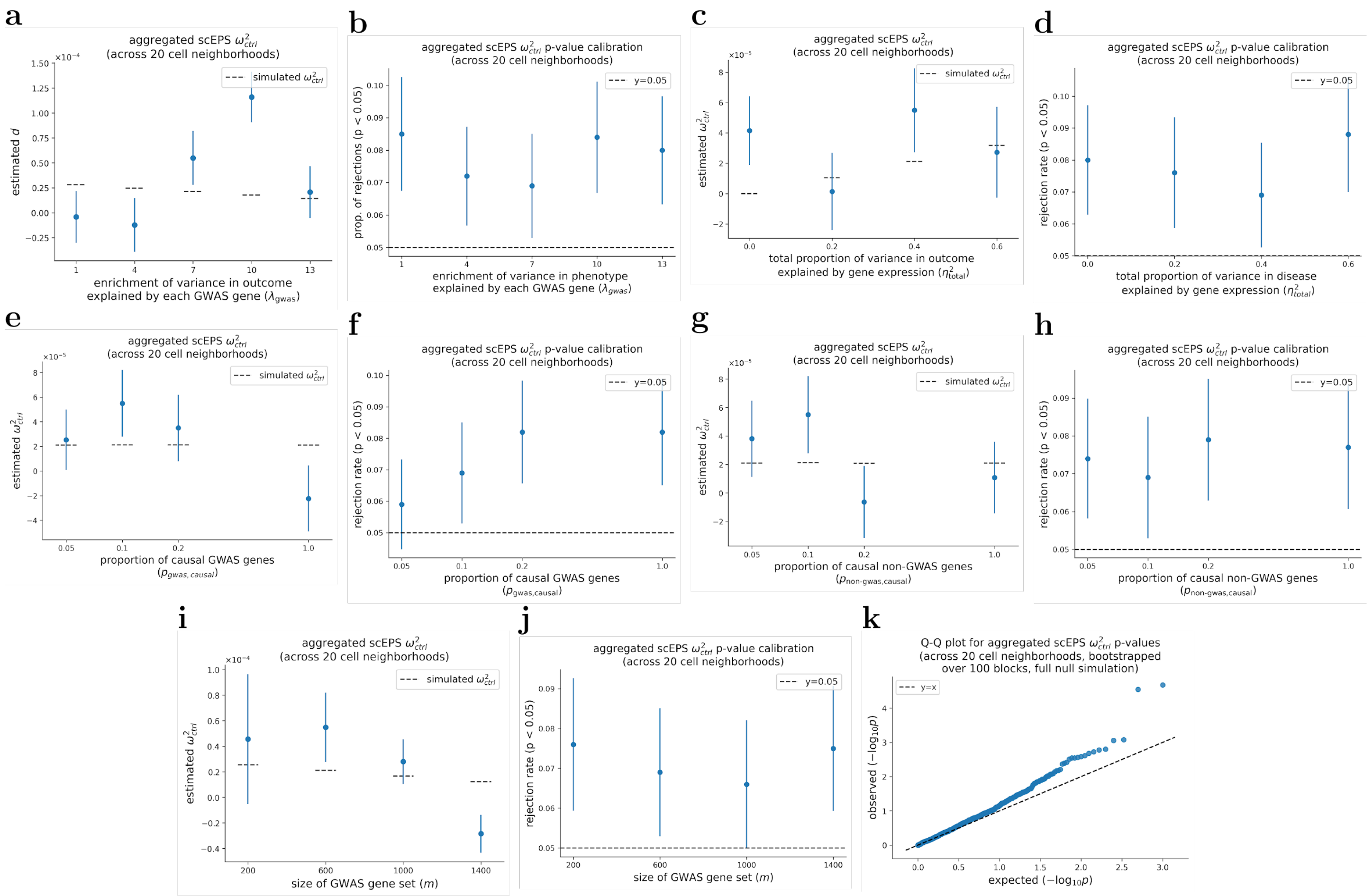


Supplementary Figure 13: **Performance of scEPS in estimating aggregated $\omega^2{}_{ctrl}$ statistics across 20 cell neighborhoods.** We report the average estimates of aggregated $\omega^2{}_{ctrl}$ across 20 cell neighborhoods and proportions of rejected null hypotheses, respectively, across simulated $\lambda^2{}_{gwas}$ in (**a**, **b**), across simulated $\eta^2{}_{total}$ in (**c**, **d**), across $p_{gwas,causal}$ in (**e**, **f**), across $p_{non-gwas,causal}$ in (**g**, **h**), and across sizes of GWAS gene sets in (**i**, **j**). (**k**) Q-Q plots showing the expected vs. observed -log$_{10}$ p-values for aggregated $\omega^2{}_{ctrl}$ across 1,000 full null ($\eta^2{}_{total} = 0$) simulations. In simulations where one parameter was varied, the other parameters were fixed at their default values, with $\lambda_{gwas} = 7.0$, $\eta^2{}_{total} = 0.4$, $m_{sim} = 600$, and $p_{gwas,causal} = p_{non-gwas,causal} = 0.05$. Mean and standard errors were obtained based on 1,000 simulations. P-values testing the statistical significance of aggregated $\omega^2{}_{ctrl}$ was obtained by bootstrapping over 100 approximately independent cell neighborhood blocks**.** Error bars represent 1.96× standard errors on both times. Numerical results are reported in Supplementary Table 3.

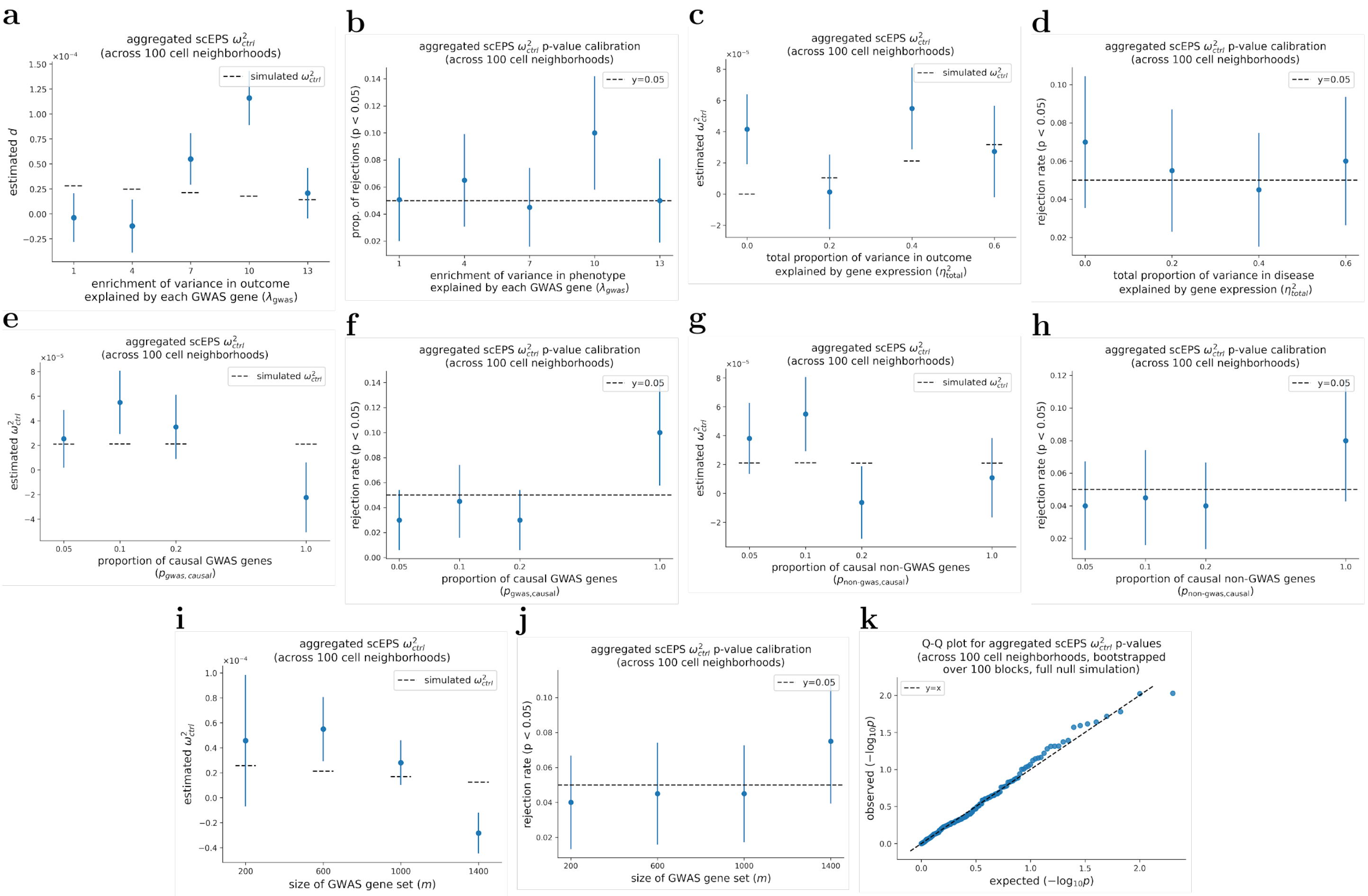


Supplementary Figure 14: **Performance of scEPS in estimating aggregated $\omega^2{}_{ctrl}$ statistics across 100 cell neighborhoods.** We report the average estimates of aggregated $\omega^2{}_{ctrl}$ across 100 cell neighborhoods and proportions of rejected null hypotheses, respectively, across simulated $\lambda^2{}_{gwas}$ in (**a**, **b**), across simulated $\eta^2{}_{total}$ in (**c**, **d**), across $p_{gwas,causal}$ in (**e**, **f**), across $p_{non-gwas,causal}$ in (**g**, **h**), and across sizes of GWAS gene sets in (**i**, **j**). (**k**) Q-Q plots showing the expected vs. observed -log$_{10}$ p-values for aggregated $\omega^2{}_{ctrl}$ across 200 full null ($\eta^2{}_{total} = 0$) simulations. In simulations where one parameter was varied, the other parameters were fixed at their default values, with $\lambda_{gwas} = 7.0$, $\eta^2{}_{total} = 0.4$, $m_{sim} = 600$, and $p_{gwas,causal} = p_{non-gwas,causal} = 0.05$. Mean and standard errors were obtained based on 200 simulations. P-values testing the statistical significance of aggregated $\omega^2{}_{ctrl}$ was obtained by bootstrapping over 100 approximately independent cell neighborhood blocks. Error bars represent 1.96× standard errors on both times. Numerical results are reported in Supplementary Table 3.

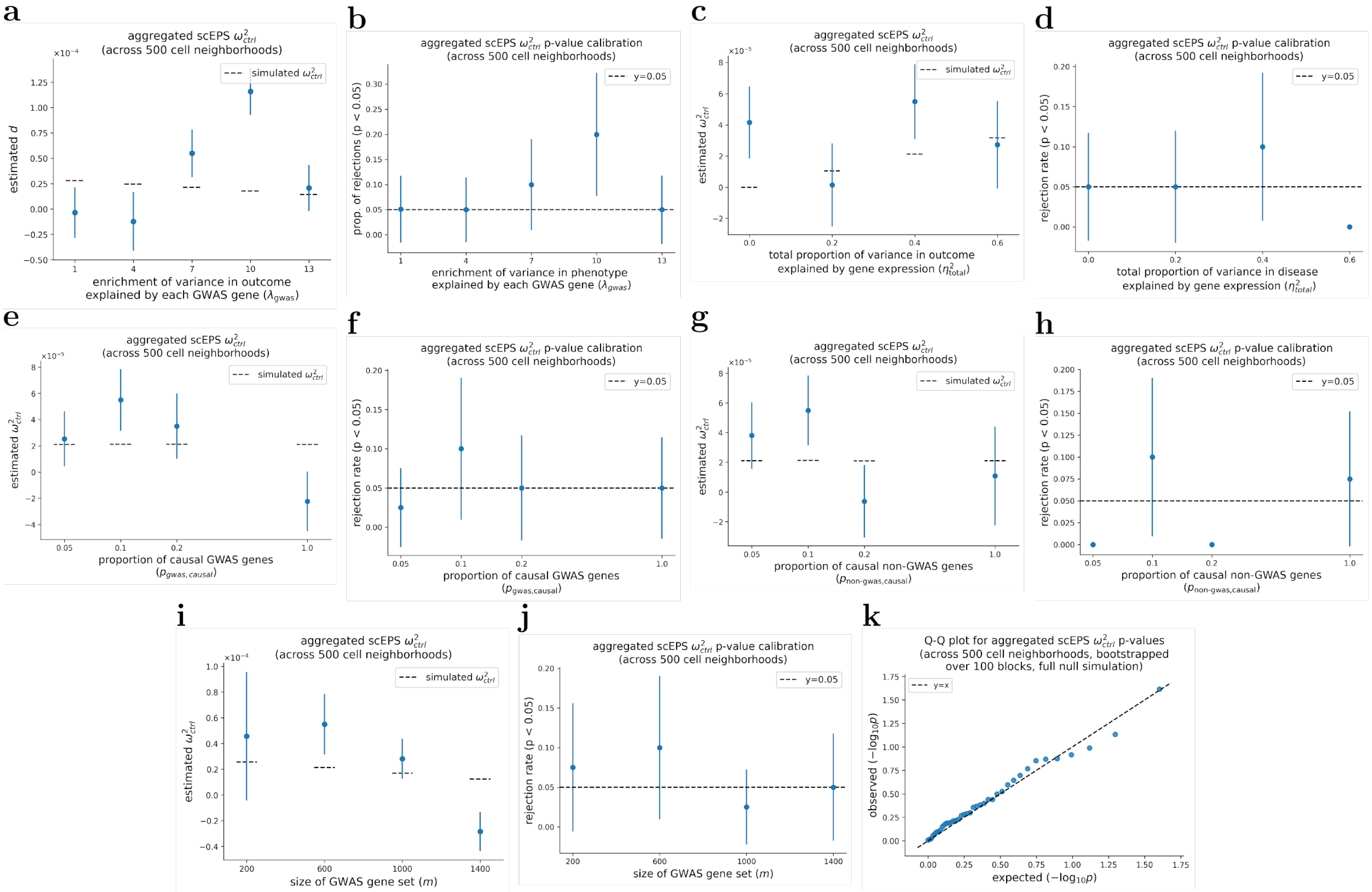


Supplementary Figure 15: **Performance of scEPS in estimating aggregated $\omega^2{}_{ctrl}$ statistics across 500 cell neighborhoods.** We report the average estimates of aggregated $\omega^2{}_{ctrl}$ across 500 cell neighborhoods and proportions of rejected null hypotheses, respectively, across simulated $\lambda^2{}_{gwas}$ in (**a**, **b**), across simulated $\eta^2{}_{total}$ in (**c**, **d**), across $p_{gwas,causal}$ in (**e**, **f**), across $p_{non-gwas,causal}$ in (**g**, **h**), and across sizes of GWAS gene sets in (**i**, **j**). (**k**) Q-Q plots showing the expected vs. observed -log$_{10}$ p-values for aggregated $\omega^2{}_{ctrl}$ across 40 full null ($\eta^2{}_{total} = 0$) simulations. In simulations where one parameter was varied, the other parameters were fixed at their default values, with $\lambda_{gwas} = 7.0$, $\eta^2{}_{total} = 0.4$, $m_{sim} = 600$, and $p_{gwas,causal} = p_{non-gwas,causal} = 0.05$. Mean and standard errors were obtained based on 40 simulations. P-values testing the statistical significance of aggregated $\omega^2{}_{ctrl}$ was obtained by bootstrapping over 100 approximately independent cell neighborhood blocks**.** Error bars represent 1.96× standard errors on both times. Numerical results are reported in Supplementary Table 3.

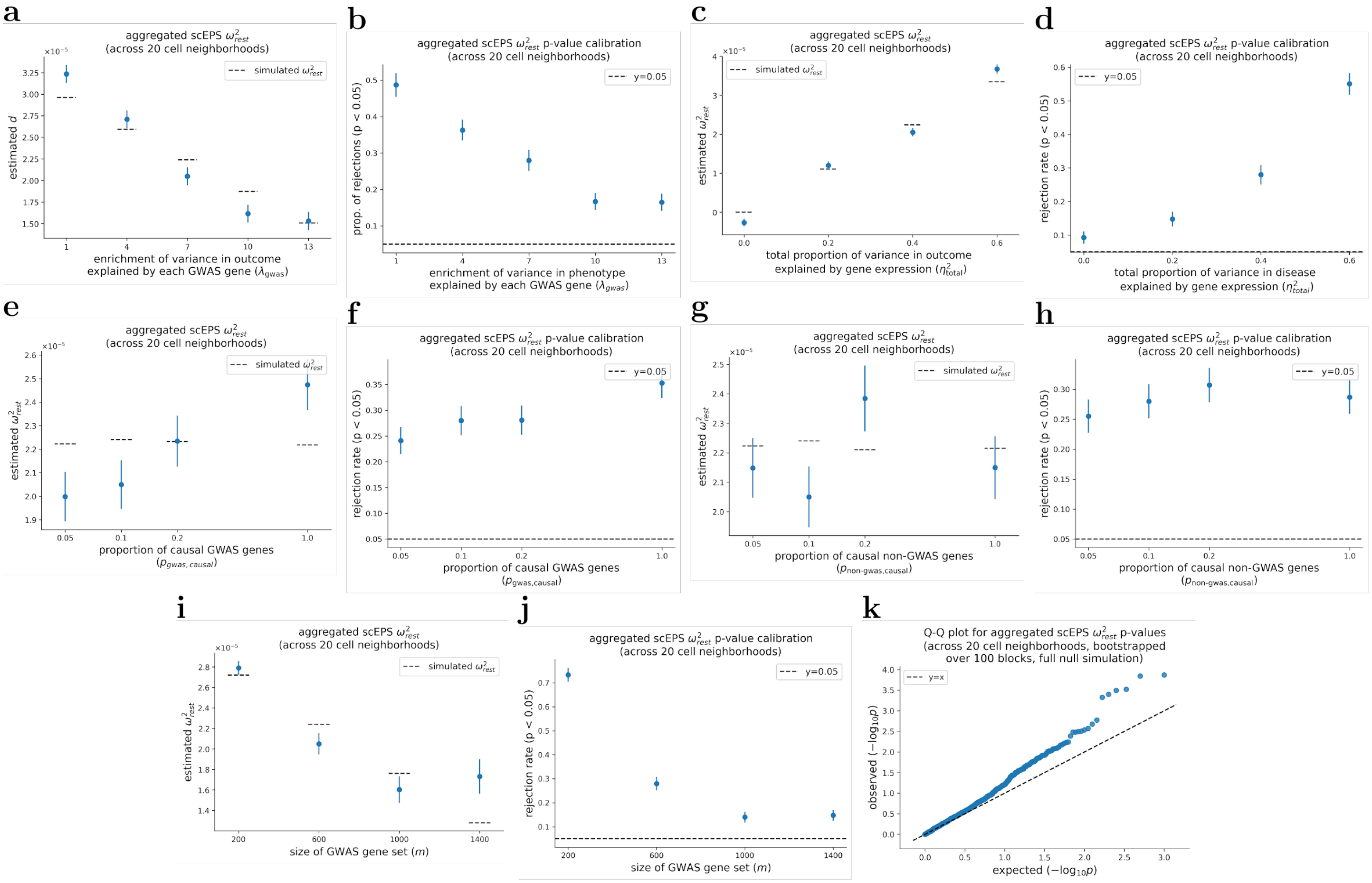


Supplementary Figure 16: **Performance of scEPS in estimating aggregated $\omega^2{}_{rest}$ statistics across 20 cell neighborhoods.** We report the average estimates of aggregated $\omega^2{}_{rest}$ across 20 cell neighborhoods and proportions of rejected null hypotheses, respectively, across simulated $\lambda^2{}_{gwas}$ in (**a**, **b**), across simulated $\eta^2{}_{total}$ in (**c**, **d**), across $p_{gwas,causal}$ in (**e**, **f**), across $p_{non-gwas,causal}$ in (**g**, **h**), and across sizes of GWAS gene sets in (**i**, **j**). (**k**) Q-Q plots showing the expected vs. observed -log$_{10}$ p-values for aggregated $\omega^2{}_{rest}$ across 1,000 full null ($\eta^2{}_{total} = 0$) simulations. In simulations where one parameter was varied, the other parameters were fixed at their default values, with $\lambda_{gwas} = 7.0$, $\eta^2{}_{total} = 0.4$, $m_{sim} = 600$, and $p_{gwas,causal} = p_{non-gwas,causal} = 0.05$. Mean and standard errors were obtained based on 1,000 simulations. P-values testing the statistical significance of aggregated $\omega^2{}_{rest}$ was obtained by bootstrapping over 100 approximately independent cell neighborhood blocks**.** Error bars represent 1.96× standard errors on both times. Numerical results are reported in Supplementary Table 3.

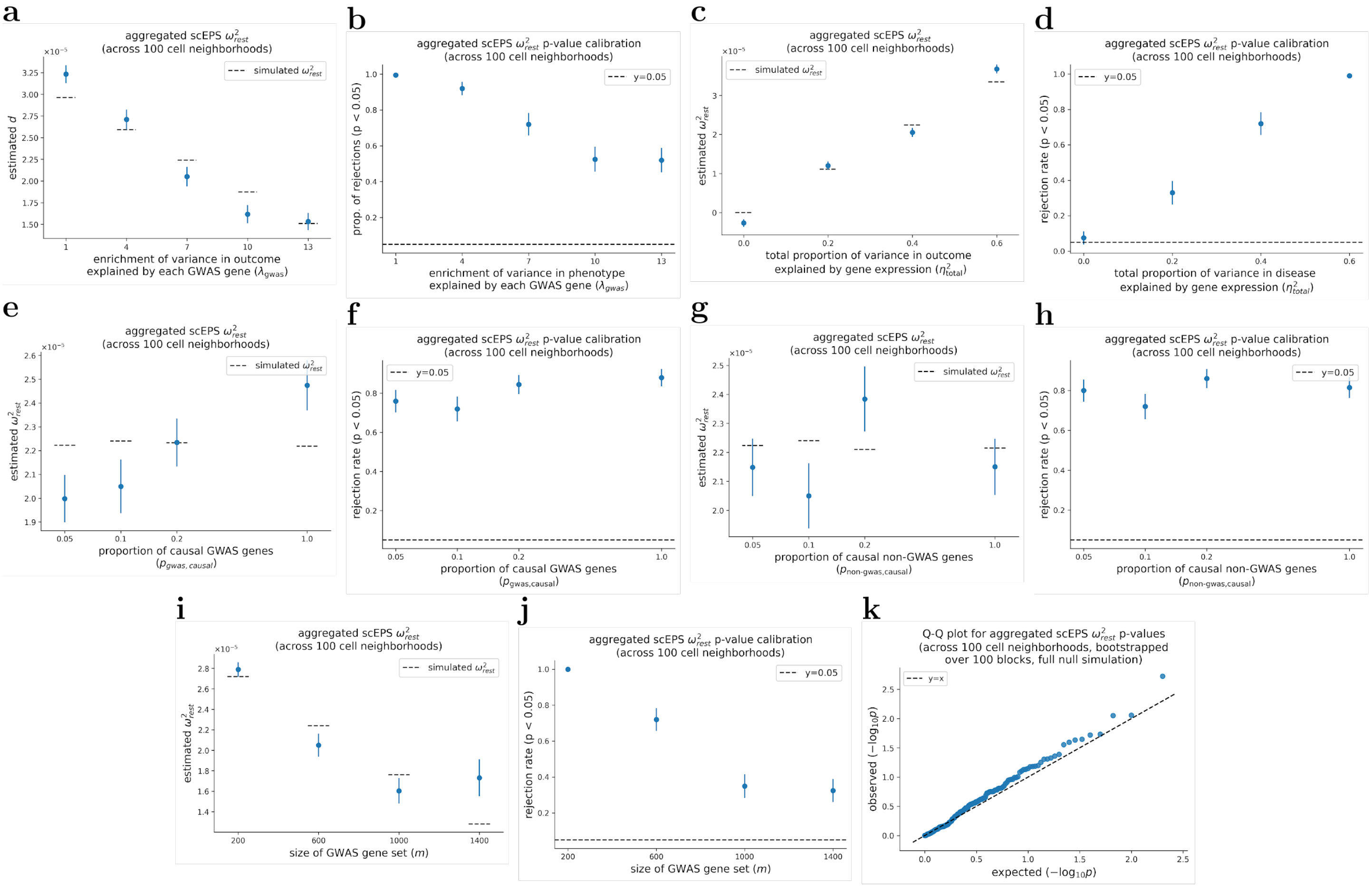


Supplementary Figure 17: **Performance of scEPS in estimating aggregated $\omega^2{}_{rest}$ statistics across 100 cell neighborhoods.** We report the average estimates of aggregated $\omega^2{}_{rest}$ across 100 cell neighborhoods and proportions of rejected null hypotheses, respectively, across simulated $\lambda^2{}_{gwas}$ in (**a**, **b**), across simulated $\eta^2{}_{total}$ in (**c**, **d**), across $p_{gwas,causal}$ in (**e**, **f**), across $p_{non-gwas,causal}$ in (**g**, **h**), and across sizes of GWAS gene sets in (**i**, **j**). (**k**) Q-Q plots showing the expected vs. observed -$\log_{10}$ p-values for aggregated $\omega^2{}_{rest}$ across 200 full null ($\eta^2{}_{total} = 0$) simulations. In simulations where one parameter was varied, the other parameters were fixed at their default values, with $\lambda_{gwas} = 7.0$, $\eta^2{}_{total} = 0.4$, $m_{sim} = 600$, and $p_{gwas,causal} = p_{non-gwas,causal} = 0.05$. Mean and standard errors were obtained based on 200 simulations. P-values testing the statistical significance of aggregated $\omega^2{}_{rest}$ was obtained by bootstrapping over 100 approximately independent cell neighborhood blocks**.** Error bars represent 1.96× standard errors on both times. Numerical results are reported in Supplementary Table 3.

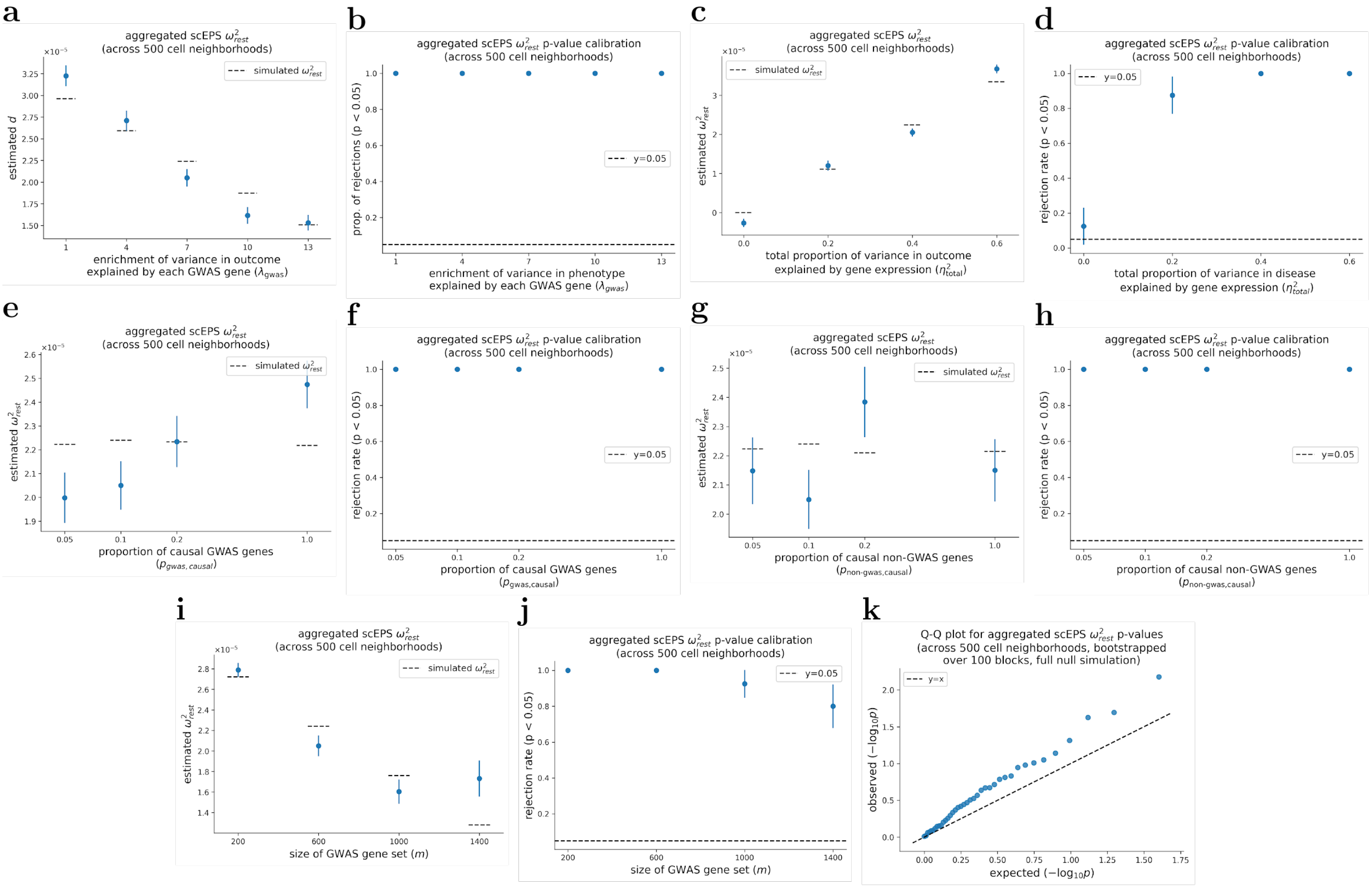


Supplementary Figure 18: **Performance of scEPS in estimating aggregated $\omega^2{}_{rest}$ statistics across 500 cell neighborhoods.** We report the average estimates of aggregated $\omega^2{}_{rest}$ across 500 cell neighborhoods and proportions of rejected null hypotheses, respectively, across simulated $\lambda^2{}_{gwas}$ in (**a**, **b**), across simulated $\eta^2{}_{total}$ in (**c**, **d**), across $p_{gwas,causal}$ in (**e**, **f**), across $p_{non-gwas,causal}$ in (**g**, **h**), and across sizes of GWAS gene sets in (**i**, **j**). (**k**) Q-Q plots showing the expected vs. observed -log$_{10}$ p-values for aggregated $\omega^2{}_{rest}$ across 40 full null ($\eta^2{}_{total} = 0$) simulations. In simulations where one parameter was varied, the other parameters were fixed at their default values, with $\lambda_{gwas} = 7.0$, $\eta^2{}_{total} = 0.4$, $m_{sim} = 600$, and $p_{gwas,causal} = p_{non-gwas,causal} = 0.05$. Mean and standard errors were obtained based on 40 simulations. P-values testing the statistical significance of aggregated $\omega^2{}_{rest}$ was obtained by bootstrapping over 100 approximately independent cell neighborhood blocks**.** Error bars represent 1.96× standard errors on both times. Numerical results are reported in Supplementary Table 3.

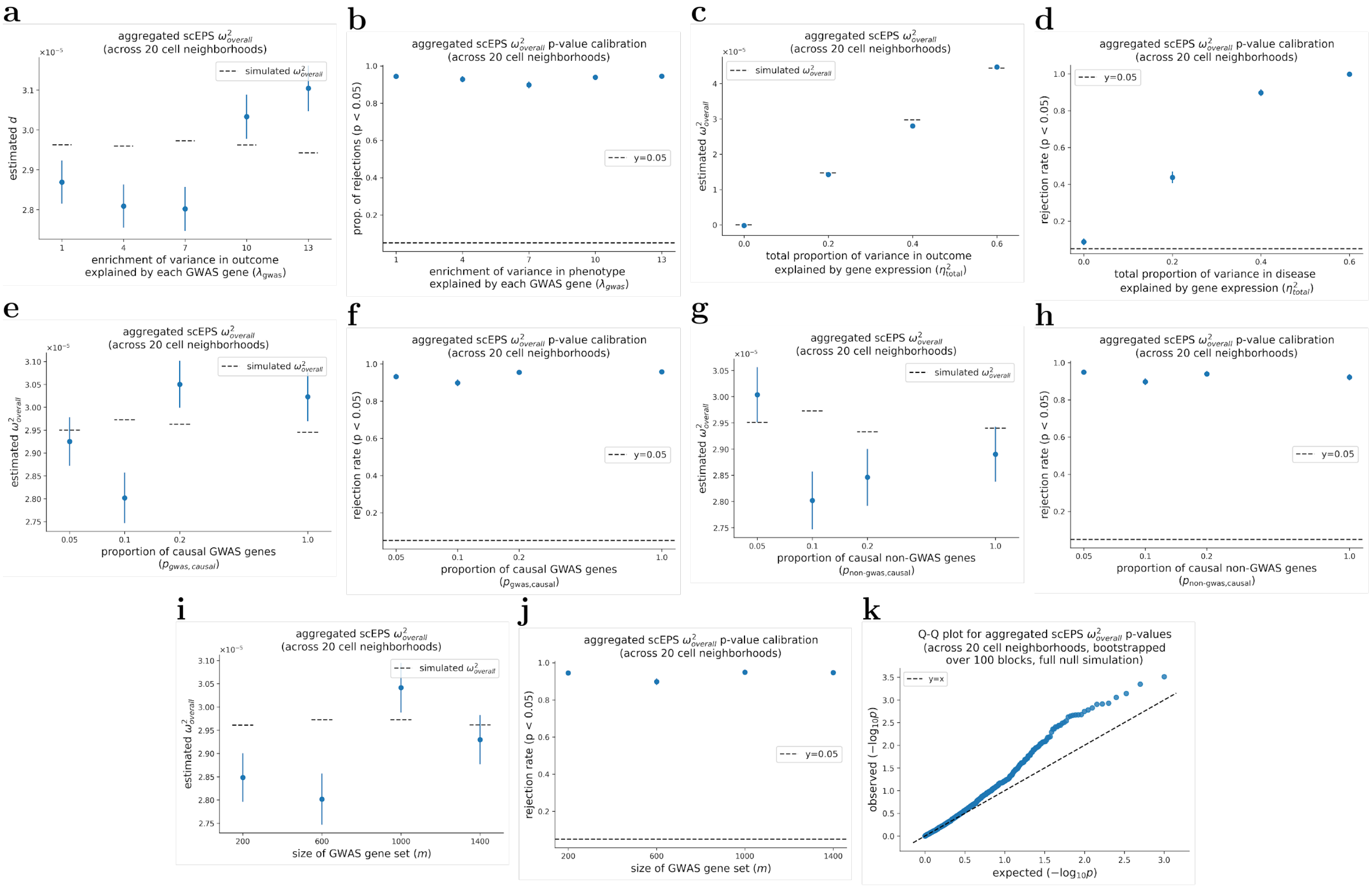


Supplementary Figure 19: **Performance of scEPS in estimating aggregated $\omega^2{}_{overall}$ statistics across 20 cell neighborhoods.** We report the average estimates of aggregated $\omega^2{}_{overall}$ across 20 cell neighborhoods and proportions of rejected null hypotheses, respectively, across simulated $\lambda^2{}_{gwas}$ in (**a**, **b**), across simulated $\eta^2{}_{total}$ in (**c**, **d**), across $p_{gwas,causal}$ in (**e**, **f**), across $p_{non-gwas,causal}$ in (**g**, **h**), and across sizes of GWAS gene sets in (**i**, **j**). (**k**) Q-Q plots showing the expected vs. observed -log$_{10}$ p-values for aggregated $\omega^2{}_{overall}$ across 1,000 full null ($\eta^2{}_{total} = 0$) simulations. In simulations where one parameter was varied, the other parameters were fixed at their default values, with $\lambda_{gwas} = 7.0$, $\eta^2{}_{total} = 0.4$, $m_{sim} = 600$, and $p_{gwas,causal} = p_{non-gwas,causal} = 0.05$. Mean and standard errors were obtained based on 1,000 simulations. P-values testing the statistical significance of aggregated $\omega^2{}_{overall}$ was obtained by bootstrapping over 100 approximately independent cell neighborhood blocks**.** Error bars represent 1.96× standard errors on both times. Numerical results are reported in Supplementary Table 3.

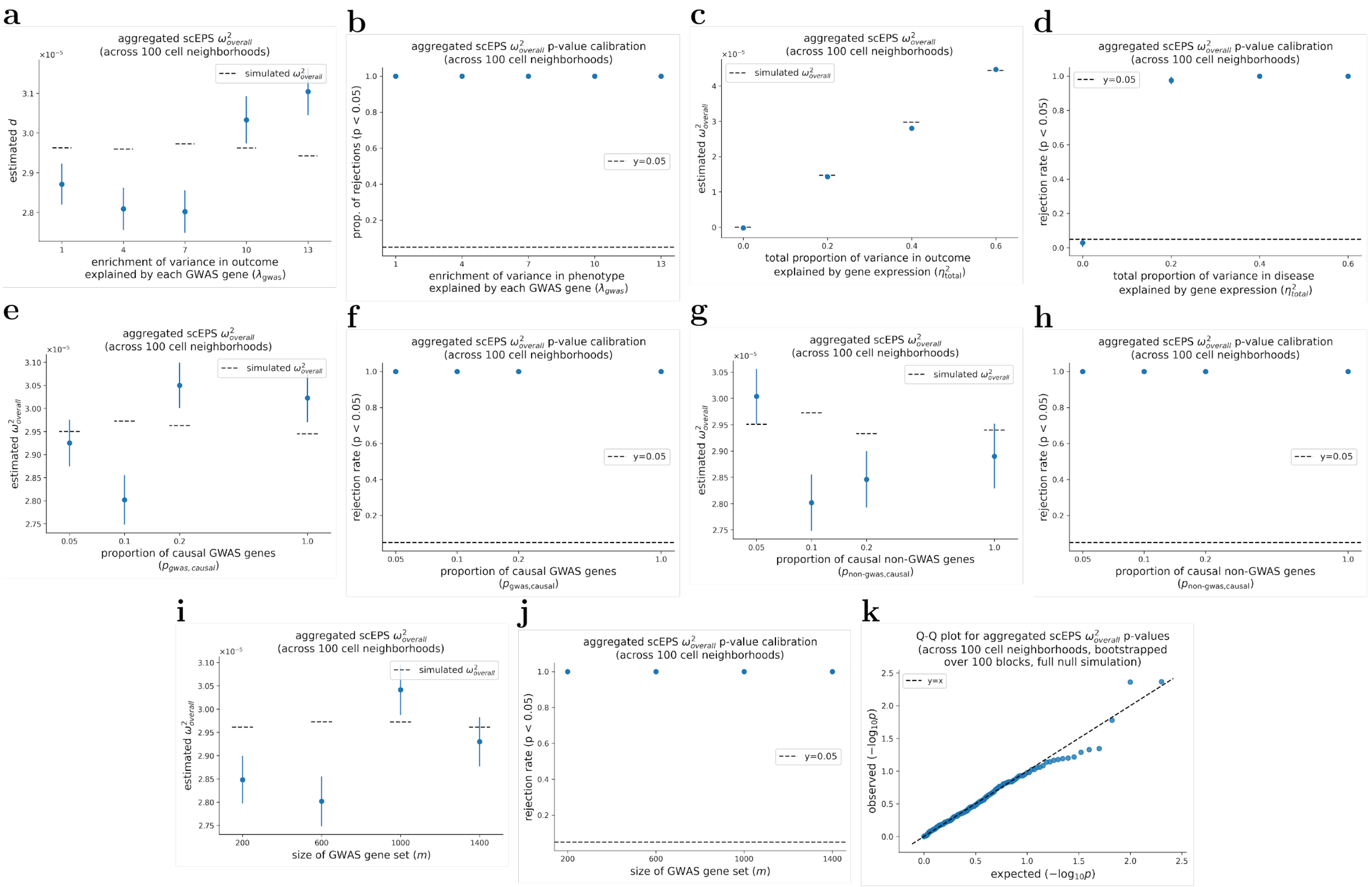


Supplementary Figure 20: **Performance of scEPS in estimating aggregated $\omega^2{}_{overall}$ statistics across 100 cell neighborhoods.** We report the average estimates of aggregated $\omega^2{}_{overall}$ across 100 cell neighborhoods and proportions of rejected null hypotheses, respectively, across simulated $\lambda^2{}_{gwas}$ in (**a**, **b**), across simulated $\eta^2{}_{total}$ in (**c**, **d**), across $p_{gwas,causal}$ in (**e**, **f**), across $p_{non-gwas,causal}$ in (**g**, **h**), and across sizes of GWAS gene sets in (**i**, **j**). (**k**) Q-Q plots showing the expected vs. observed -log$_{10}$ p-values for aggregated $\omega^2{}_{overall}$ across 200 full null ($\eta^2{}_{total} = 0$) simulations. In simulations where one parameter was varied, the other parameters were fixed at their default values, with $\lambda_{gwas} = 7.0$, $\eta^2{}_{total} = 0.4$, $m_{sim} = 600$, and $p_{gwas,causal} = p_{non-gwas,causal} = 0.05$. Mean and standard errors were obtained based on 200 simulations. P-values testing the statistical significance of aggregated $\omega^2{}_{overall}$ was obtained by bootstrapping over 100 approximately independent cell neighborhood blocks**.** Error bars represent 1.96× standard errors on both times. Numerical results are reported in Supplementary Table 3.

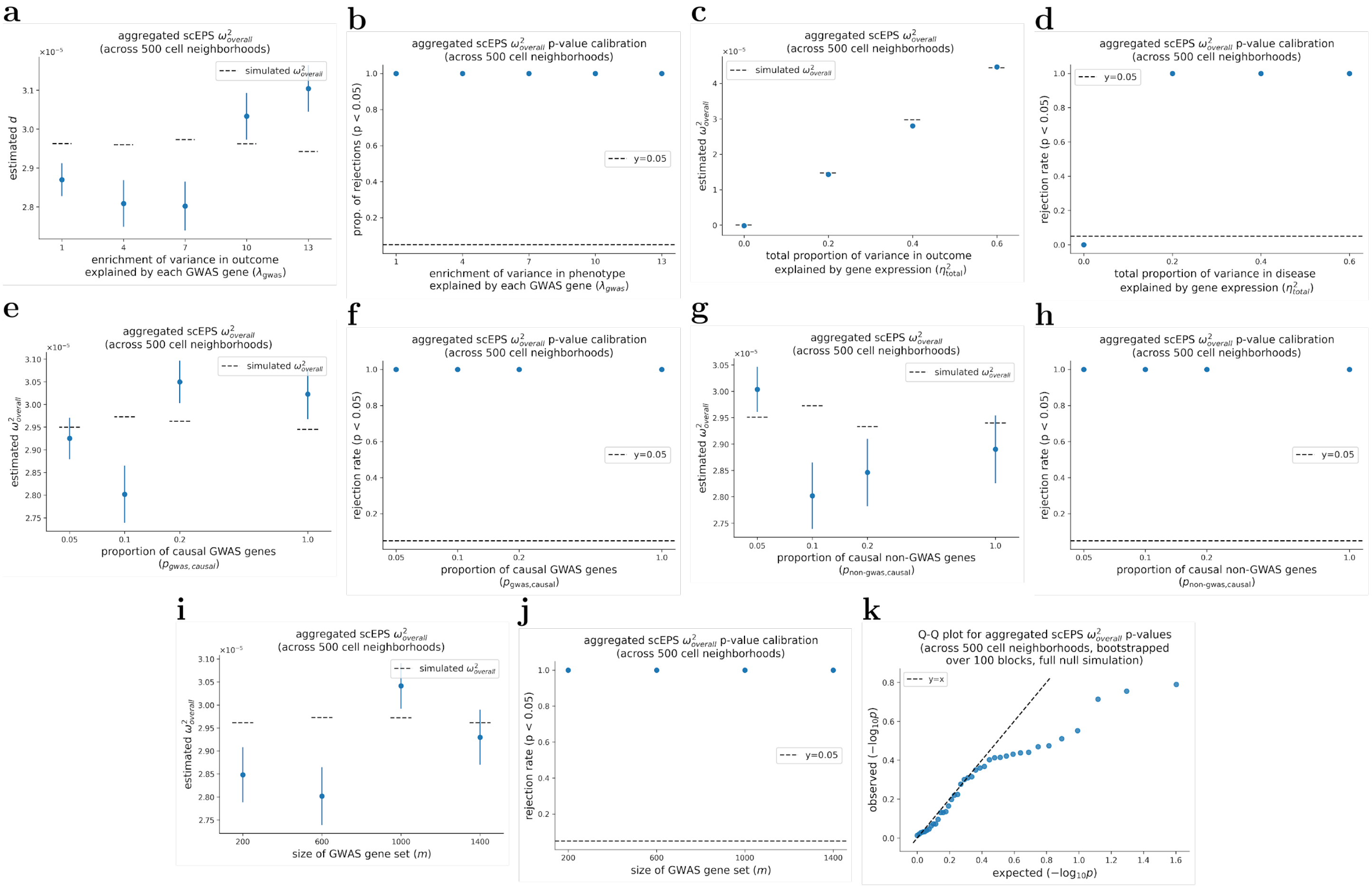


Supplementary Figure 21: **Performance of scEPS in estimating aggregated $\boldsymbol{\omega^2}_{\boldsymbol{overall}}$ statistics across 500 cell neighborhoods.** We report the average estimates of aggregated $\omega^2{}_{overall}$ across 500 cell neighborhoods and proportions of rejected null hypotheses, respectively, across simulated $\lambda^2{}_{gwas}$ in (**a**, **b**), across simulated $\eta^2{}_{total}$ in (**c**, **d**), across $p_{gwas,causal}$ in (**e**, **f**), across $p_{non-gwas,causal}$ in (**g**, **h**), and across sizes of GWAS gene sets in (**i**, **j**). (**k**) Q-Q plots showing the expected vs. observed -log$_{10}$ p-values for aggregated $\omega^2{}_{overall}$ across 40 full null ($\eta^2{}_{total} = 0$) simulations. In simulations where one parameter was varied, the other parameters were fixed at their default values, with $\lambda_{gwas} = 7.0$, $\eta^2{}_{total} = 0.4$, $m_{sim} = 600$, and $p_{gwas,causal} = p_{non-gwas,causal} = 0.05$. Mean and standard errors were obtained based on 40 simulations. P-values testing the statistical significance of aggregated $\omega^2{}_{overall}$ was obtained by bootstrapping over 100 approximately independent cell neighborhood blocks**.** Error bars represent 1.96× standard errors on both times. Numerical results are reported in Supplementary Table 3.

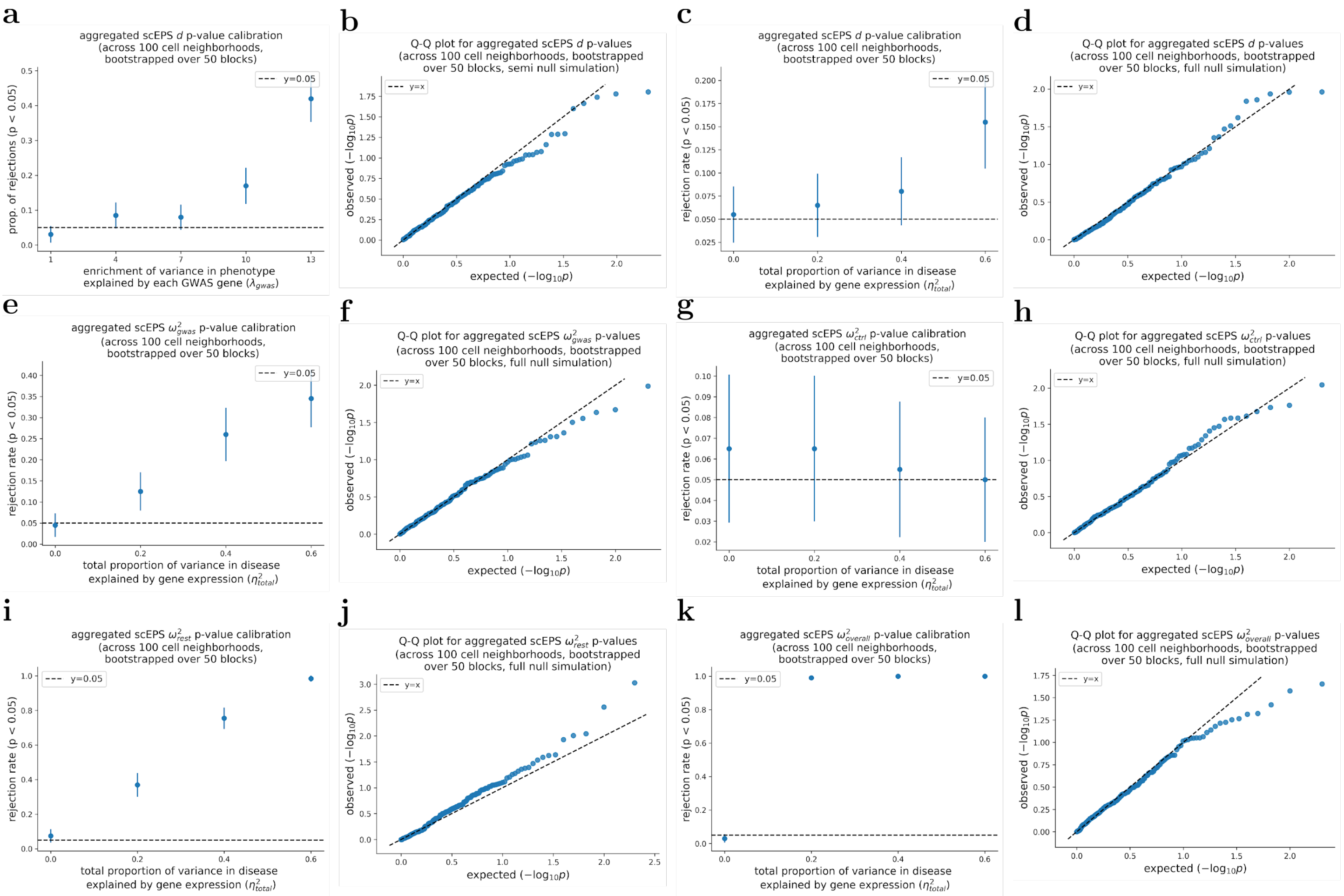


Supplementary Figure 22: **Calibration of scEPS' p-values for aggregated statistics across 100 cell neighborhoods obtained using 50 cell neighborhood blocks.** (**a**) We report the proportion of rejected null hypotheses testing aggregated $d$ across simulated $\lambda^2{}_{gwas}$. (**b**) Q-Q plot showing the expected vs. observed -$\log_{10}$ p-values for aggregated $d$ across 200 semi null (${\eta^2}_{total} = 0.4$, $\lambda_{gwas} = 1.0$) simulations. (**c**, **e**, **g**, **i**, **k**) We report the proportion of rejected null hypotheses testing aggregated $d$, ${\omega^2}_{gwas}$, ${\omega^2}_{ctrl}$, ${\omega^2}_{rest}$, and ${\omega^2}_{overall}$, respectively, across simulated ${\eta^2}_{total}$. (**d**, **f**, **h**, **j**, **l**) Q-Q plot showing the expected vs. observed -$\log_{10}$ p-values for aggregated $d$, ${\omega^2}_{gwas}$, ${\omega^2}_{ctrl}$, ${\omega^2}_{rest}$, and ${\omega^2}_{overall}$, respectively, across 200 full null (${\eta^2}_{total} = 0$) simulations. In simulations where one parameter was varied, the other parameters were fixed at their default values, with $\lambda_{gwas} = 7.0$, ${\eta^2}_{total} = 0.4$, $m_{sim} = 600$, and $p_{gwas,causal} = p_{non-gwas,causal} = 0.05$. Mean and standard errors were obtained based on 200 simulations. P-values testing the statistical significance of aggregated ${\omega^2}_{overall}$ was obtained by bootstrapping over 100 approximately independent cell neighborhood blocks**.** Error bars represent 1.96× standard errors on both times. Numerical results are reported in Supplementary Table 4.

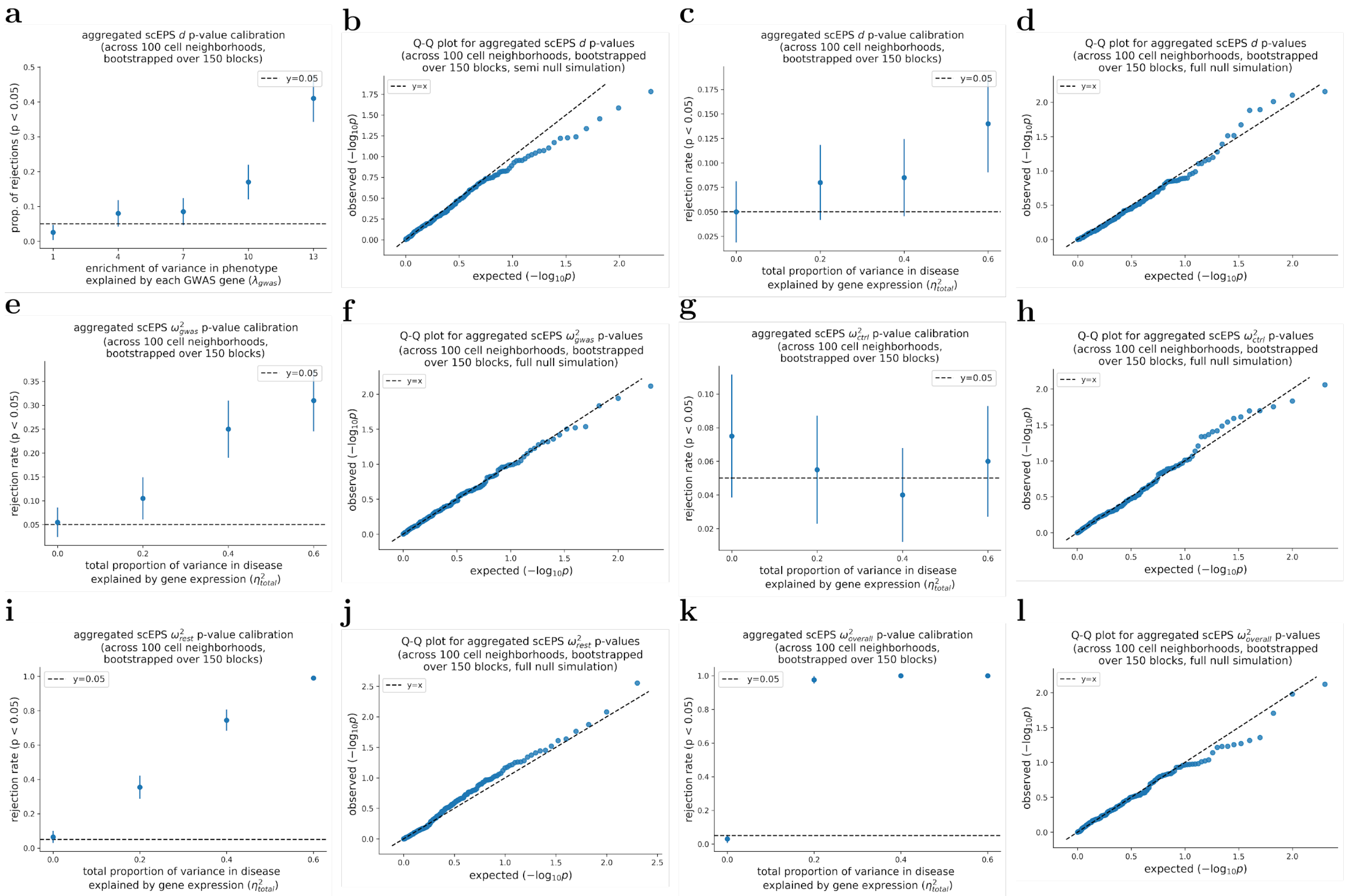


Supplementary Figure 23: **Calibration of scEPS' p-values for aggregated statistics across 100 cell neighborhoods obtained using 150 cell neighborhood blocks.** (**a**) We report the proportion of rejected null hypotheses testing aggregated $d$ across simulated $\lambda^2{}_{gwas}$. (**b**) Q-Q plot showing the expected vs. observed -log$_{10}$ p-values for aggregated $d$ across 200 semi null ($\eta^2{}_{total} = 0.4$, $\lambda_{gwas} = 1.0$) simulations. (**c**, **e**, **g**, **i**, **k**) We report the proportion of rejected null hypotheses testing aggregated $d$, $\omega^2{}_{gwas}$, $\omega^2{}_{ctrl}$, $\omega^2{}_{rest}$, and $\omega^2{}_{overall}$, respectively, across simulated $\eta^2{}_{total}$. (**d**, **f**, **h**, **j**, **l**) Q-Q plot showing the expected vs. observed -log$_{10}$ p-values for aggregated $d$, $\omega^2{}_{gwas}$, $\omega^2{}_{ctrl}$, $\omega^2{}_{rest}$, and $\omega^2{}_{overall}$ , respectively, across 200 full null ($\eta^2{}_{total} = 0$) simulations. In simulations where one parameter was varied, the other parameters were fixed at their default values, with $\lambda_{gwas} = 7.0$, $\eta^2{}_{total} = 0.4$, $m_{sim} = 600$, and $p_{gwas,causal} = p_{non-gwas,causal} = 0.05$. Mean and standard errors were obtained based on 200 simulations. P-values testing the statistical significance of aggregated $\omega^2{}_{overall}$ was obtained by bootstrapping over 100 approximately independent cell neighborhood blocks**.** Error bars represent 1.96× standard errors on both times. Numerical results are reported in Supplementary Table 4.

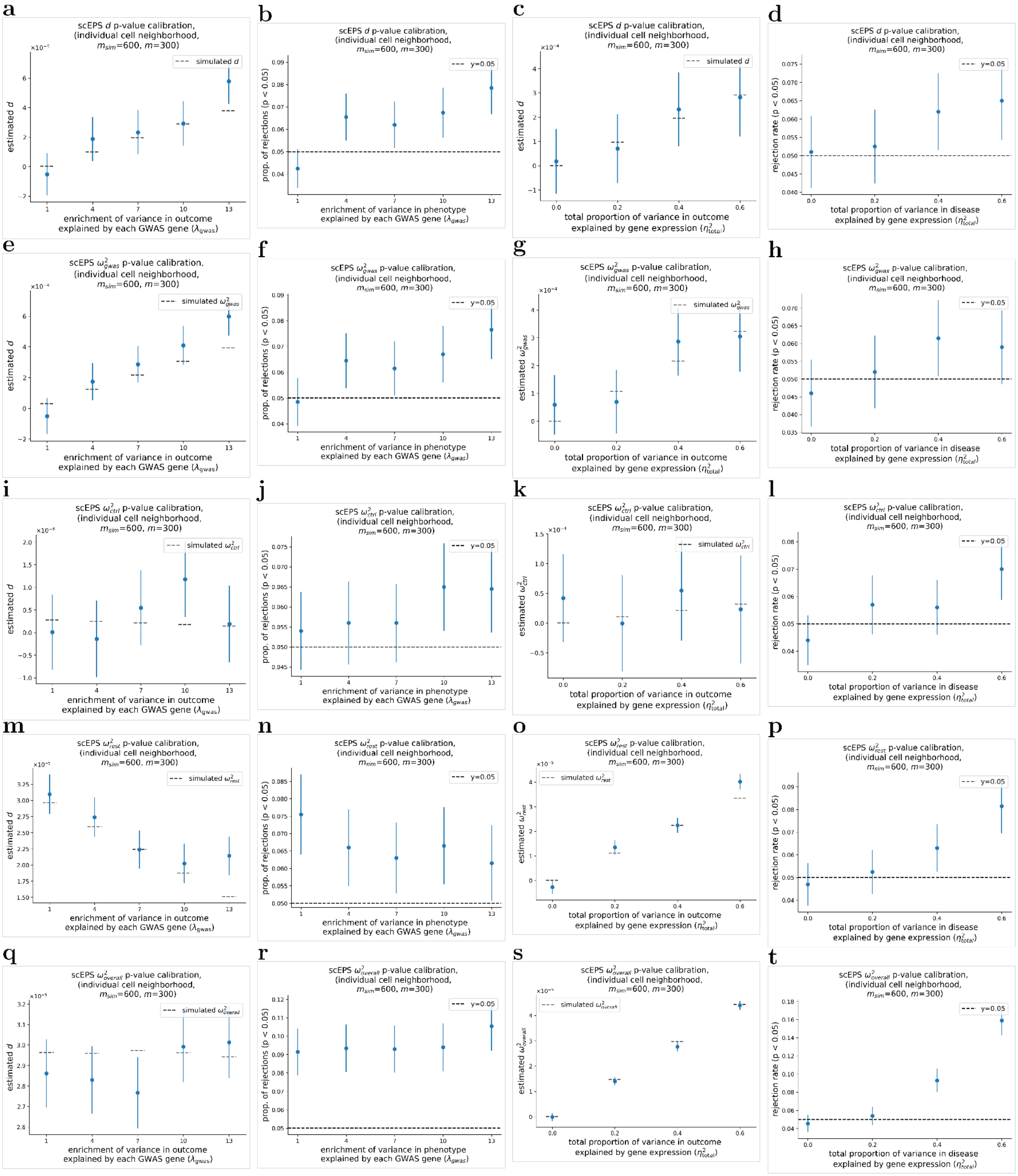


Supplementary Figure 24: **Performance of scEPS in simulations with model misspecification, where $\boldsymbol{m_{sim}} = \mathbf{600}$ and $\boldsymbol{m} = \mathbf{300}$.** (**a**, **e**, **i**, **m**, **q**) We report the average estimated $d$, ${\omega^2}_{gwas}$, ${\omega^2}_{ctrl}$, ${\omega^2}_{rest}$, and ${\omega^2}_{overall}$, respectively, across simulated $\lambda_{gwas}$. (**b**, **f**, **j**, **n**, **r**) We report the proportion of rejected null hypotheses testing $d$, ${\omega^2}_{gwas}$, ${\omega^2}_{ctrl}$, ${\omega^2}_{rest}$, and ${\omega^2}_{overall}$, respectively, across simulated $\lambda_{gwas}$. (**c**, **g**, **k**, **o**, **s**) We report the average estimated $d$, ${\omega^2}_{gwas}$, ${\omega^2}_{ctrl}$, ${\omega^2}_{rest}$, and ${\omega^2}_{overall}$ across simulated ${\eta^2}_{total}$. (**d**, **h**, **l**, **p**, **t**)

We report the proportion of rejected null hypotheses testing $d$, ${\omega^2}_{gwas}$, ${\omega^2}_{ctrl}$, ${\omega^2}_{rest}$, and ${\omega^2}_{overall}$, respectively, across simulated ${\eta^2}_{total}$. In simulations where one parameter was varied, the other parameters were fixed at their default values, with $\lambda_{gwas} = 7.0$, ${\eta^2}_{total} = 0.4$, $m_{sim} = 600$, and $p_{gwas,causal} = p_{non-gwas,causal} = 0.05$. Mean and standard errors were obtained based on 2,000 simulations. Error bars represent 1.96× standard errors on both times. Numerical results are reported in Supplementary Table 5.

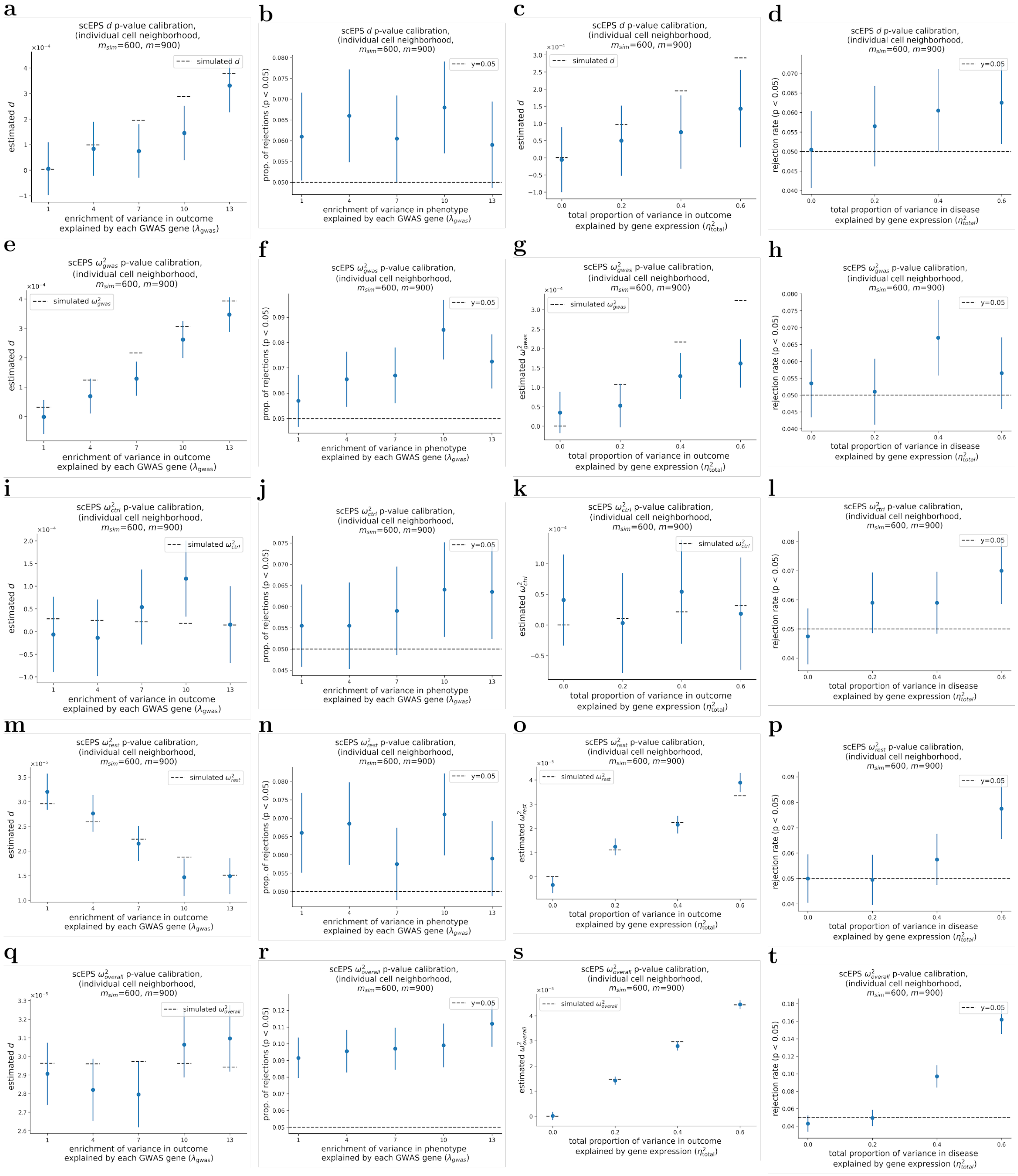


Supplementary Figure 25: **Performance of scEPS in simulations with model misspecification, where $m_{sim} = 600$ and $m = 900$.** (**a**, **e**, **i**, **m**, **q**) We report the average estimated $d$, $\omega^2{}_{gwas}$, $\omega^2{}_{ctrl}$, $\omega^2{}_{rest}$, and $\omega^2{}_{overall}$, respectively, across simulated $\lambda_{gwas}$. (**b**, **f**, **j**, **n**, **r**) We report the proportion of rejected null hypotheses testing $d$, $\omega^2{}_{gwas}$, $\omega^2{}_{ctrl}$, $\omega^2{}_{rest}$, and $\omega^2{}_{overall}$, respectively across simulated $\lambda_{gwas}$. (**c**, **g**, **k**, **o**, **s**) We report the average estimated $d$, $\omega^2{}_{gwas}$, $\omega^2{}_{ctrl}$, $\omega^2{}_{rest}$, and $\omega^2{}_{overall}$ , respectively, across simulated

$\eta^2{}_{total}$. (**d**, **h**, **l**, **p**, **t**) We report the proportion of rejected null hypotheses testing $d$, $\omega^2{}_{gwas}$, $\omega^2{}_{ctrl}$, $\omega^2{}_{rest}$, and $\omega^2{}_{overall}$, respectively, across simulated $\eta^2{}_{total}$. In simulations where one parameter was varied, the other parameters were fixed at their default values, with $\lambda_{gwas} = 7.0$, $\eta^2{}_{total} = 0.4$, $m_{sim} \quad = 600$, and $p_{gwas,causal} = p_{non-gwas,causal} = 0.05$. Mean and standard errors were obtained based on 2,000 simulations. Error bars represent 1.96× standard errors on both times. Numerical results are reported in Supplementary Table 5.

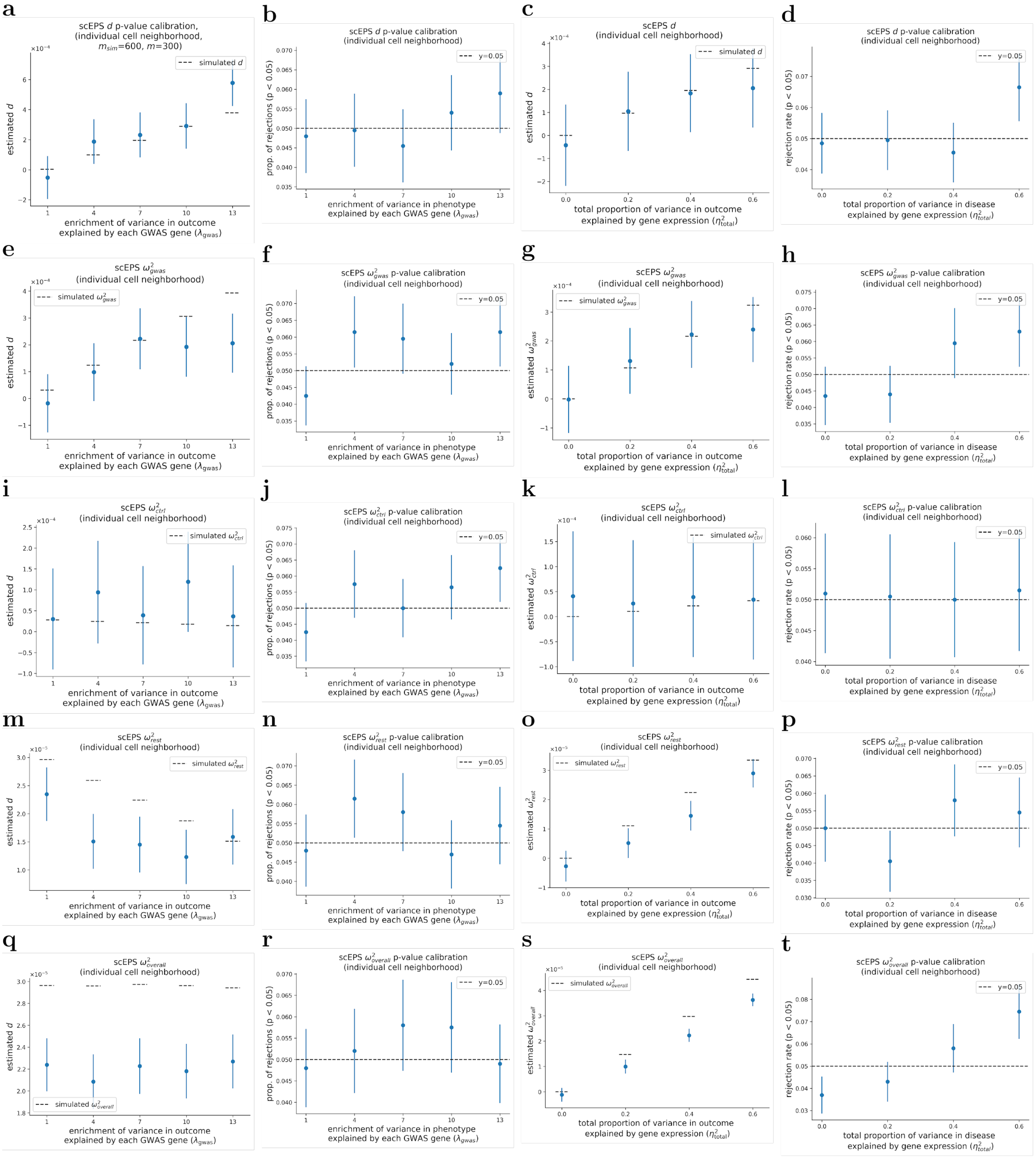


Supplementary Figure 26: **Performance of scEPS in simulations, where 50% of the cells in the cell neighborhoods were randomly removed.** (**a**, **e**, **i**, **m**, **q**) We report the average estimated $d$, $\omega^2{}_{gwas}$, $\omega^2{}_{ctrl}$, $\omega^2{}_{rest}$, and $\omega^2{}_{overall}$ , respectively, across simulated $\lambda_{gwas}$. (**b**, **f**, **j**, **n**, **r**) We report the proportion of rejected null hypotheses testing $d$, $\omega^2{}_{gwas}$, $\omega^2{}_{ctrl}$, $\omega^2{}_{rest}$, and $\omega^2{}_{overall}$, respectively, across simulated $\lambda_{gwas}$. (**c**, **g**, **k**, **o**, **s**) We report the average estimated $d$, $\omega^2{}_{gwas}$, $\omega^2{}_{ctrl}$, $\omega^2{}_{rest}$, and $\omega^2{}_{overall}$, respectively, across simulated $\eta^2{}_{total}$. (**d**, **h**, **l**, **p**, **t**) We report the proportion of rejected null hypotheses testing $d$, $\omega^2{}_{gwas}$, $\omega^2{}_{ctrl}$, $\omega^2{}_{rest}$, and

${\omega^2}_{overall}$, respectively across simulated ${\eta^2}_{total}$. In simulations where one parameter was varied, the other parameters were fixed at their default values, with $\lambda_{gwas} = 7.0$, ${\eta^2}_{total} = 0.4$, $m_{sim} \quad = 600$, and $p_{gwas,causal} = p_{non-gwas,causal} = 0.05$. Mean and standard errors were obtained based on 2,000 simulations. Error bars represent 1.96× standard errors on both times. Numerical results are reported in Supplementary Table 5.

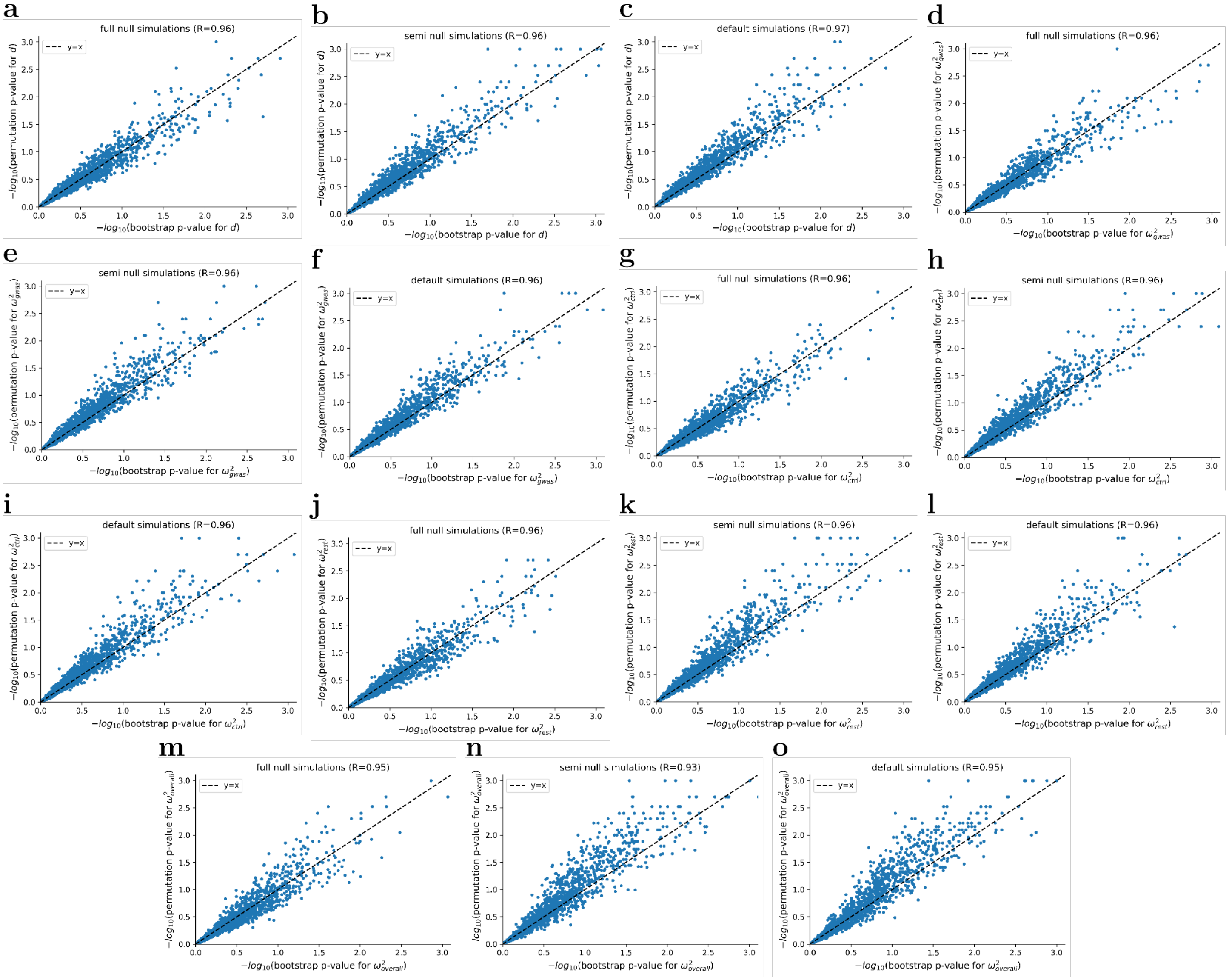


Supplementary Figure 27: **Comparison of p-values testing scEPS statistics obtained using bootstrap and permutation.** (**a**, **b**, **c**) Results for $d$ under full null ($\eta^2{}_{total} = 0$), semi null ($\eta^2{}_{total} = 0.4$, $\lambda_{gwas} = 1.0$), and default ($\lambda_{gwas} = 7.0$, $\eta^2{}_{total} = 0.4$) simulations, respectively. (**d**, **e**, **f**) Results for $\omega^2{}_{gwas}$ under full null, semi null, and default simulations, respectively. (**g**, **h**, **i**) Results for $\omega^2{}_{ctrl}$ under full null, semi null, and default simulations, respectively. (**j**, **k**, **l**) Results for $\omega^2{}_{rest}$ under full null, semi null, and default simulations, respectively. (**m**, **n**, **o**) Results for $\omega^2{}_{overall}$ under full null, semi null, and default simulations, respectively. In all simulations we set $m_{sim} = 600$, and $p_{gwas,causal} = p_{non-gwas,causal} = 0.05$.

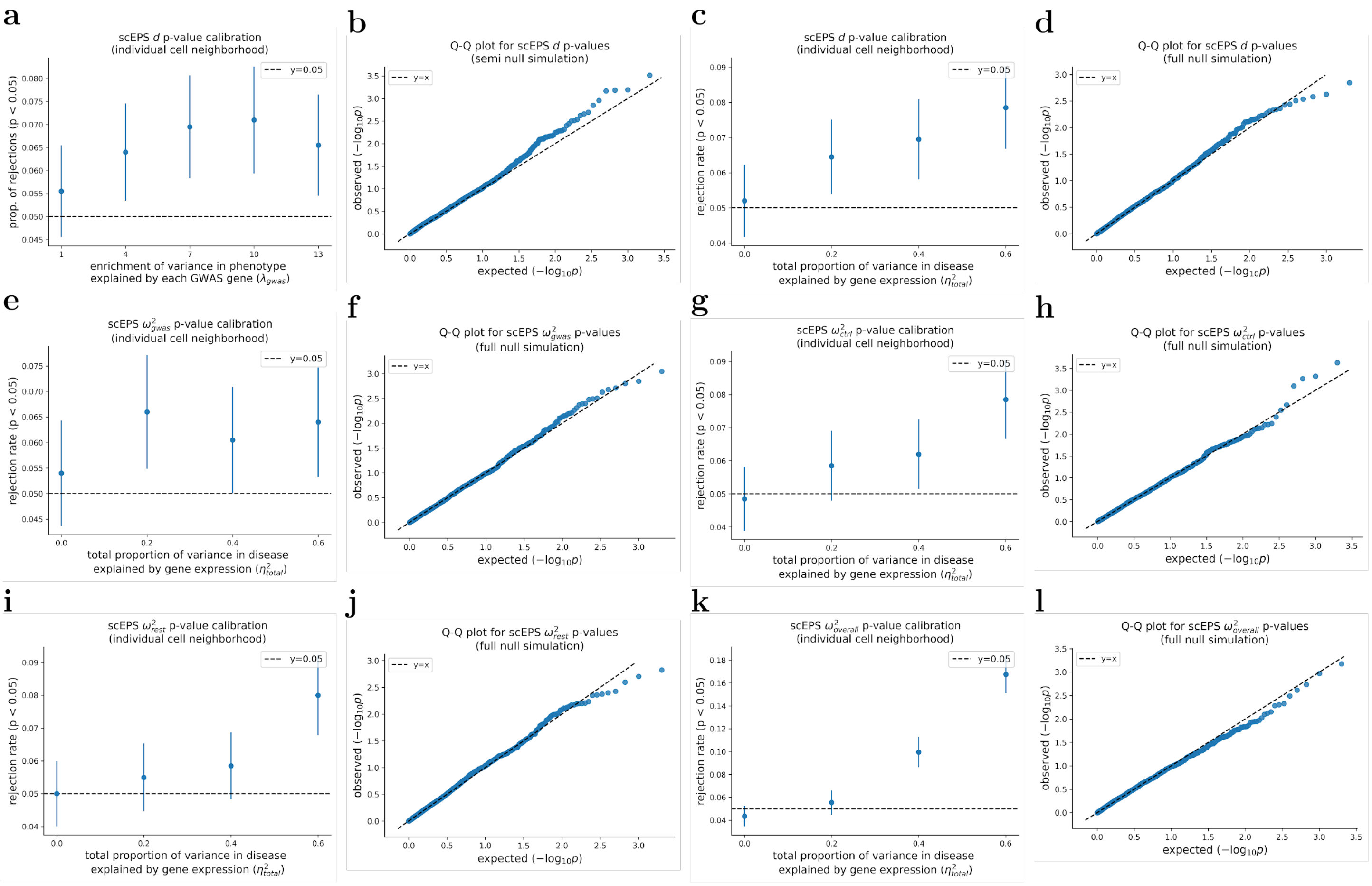


Supplementary Figure 28: **Calibration of scEPS' p-values for individual cell neighborhoods obtained using 100 bootstrap samples.** (**a**) We report the proportion of rejected null hypotheses testing the $d$ statistics across simulated $\lambda_{gwas}$. (**b**) Q-Q plot showing the expected vs. observed $-\log_{10}$ p-values for aggregated $d$ across 200 semi null ($\eta^2{}_{total} = 0.4$, $\lambda_{gwas} = 1.0$) simulations. (**c**, **e**, **g**, **i**, **k**) We report the proportion of rejected null hypotheses testing $d$, $\omega^2{}_{gwas}$, $\omega^2{}_{ctrl}$, $\omega^2{}_{rest}$, and $\omega^2{}_{overall}$, respectively, across simulated $\eta^2{}_{total}$. (**d**, **f**, **h**, **j**, **l**) Q-Q plot showing the expected vs. observed $-\log_{10}$ p-values testing $d$, $\omega^2{}_{gwas}$, $\omega^2{}_{ctrl}$, $\omega^2{}_{rest}$, and $\omega^2{}_{overall}$, respectively, across 2,000 full null ($\eta^2{}_{total} = 0$) simulations. In simulations where one parameter was varied, the other parameters were fixed at their default values, with $\lambda_{gwas} = 7.0$, $\eta^2{}_{total} = 0.4$, $m_{sim} = 600$, and $p_{gwas,causal} = p_{non-gwas,causal} = 0.05$. Mean and standard errors were obtained based on 2,000 simulations. Numerical results are reported in Supplementary Table 2.

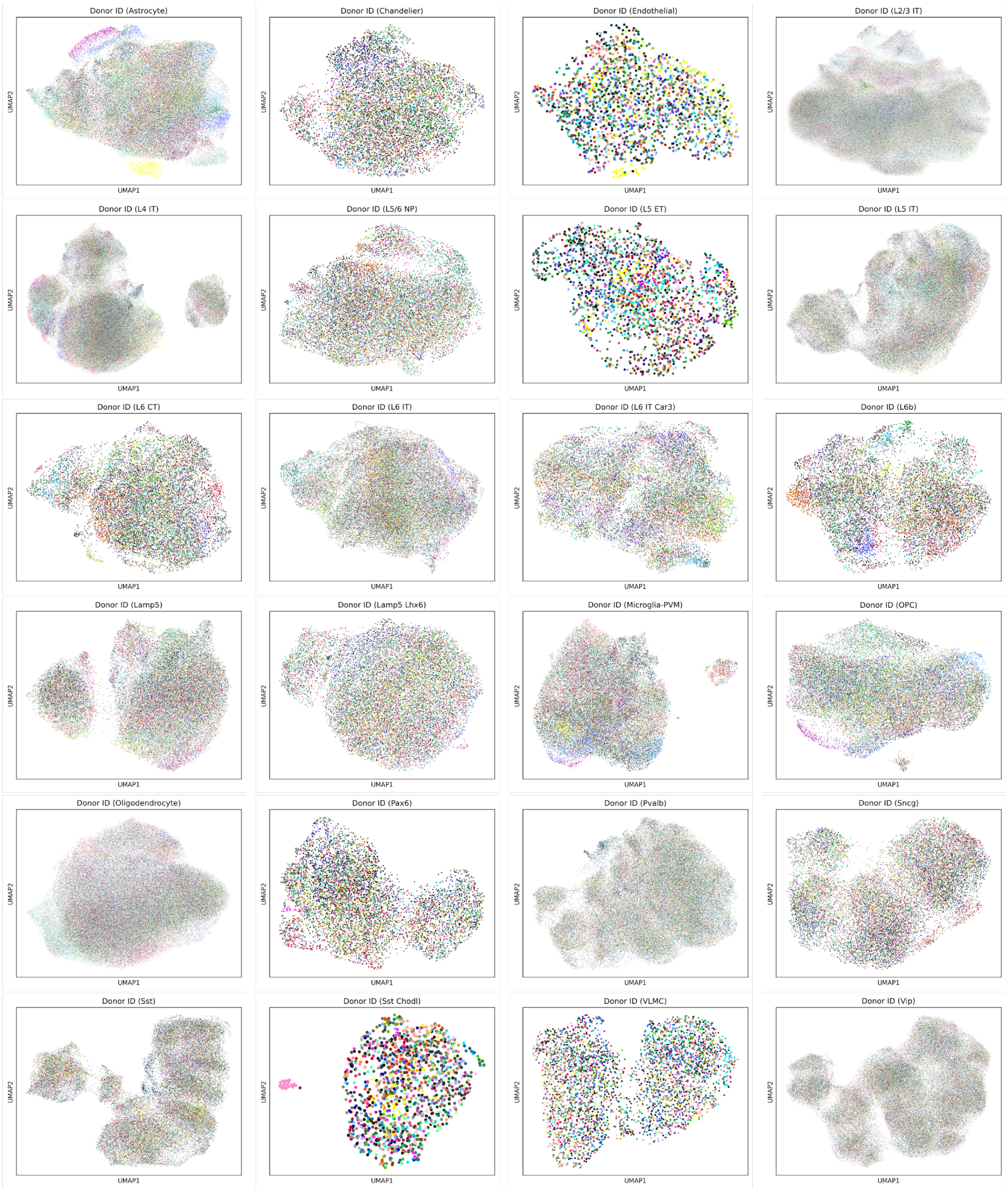


Supplementary Figure 29: **UMAP plots visualizing the cells from the 81 donors in the SEA-AD brain cell atlas data.** Each plot represents one of the 24 cell types annotated in the SEA-AD brain cell atlas data. Different colors in each plot represent different donors.

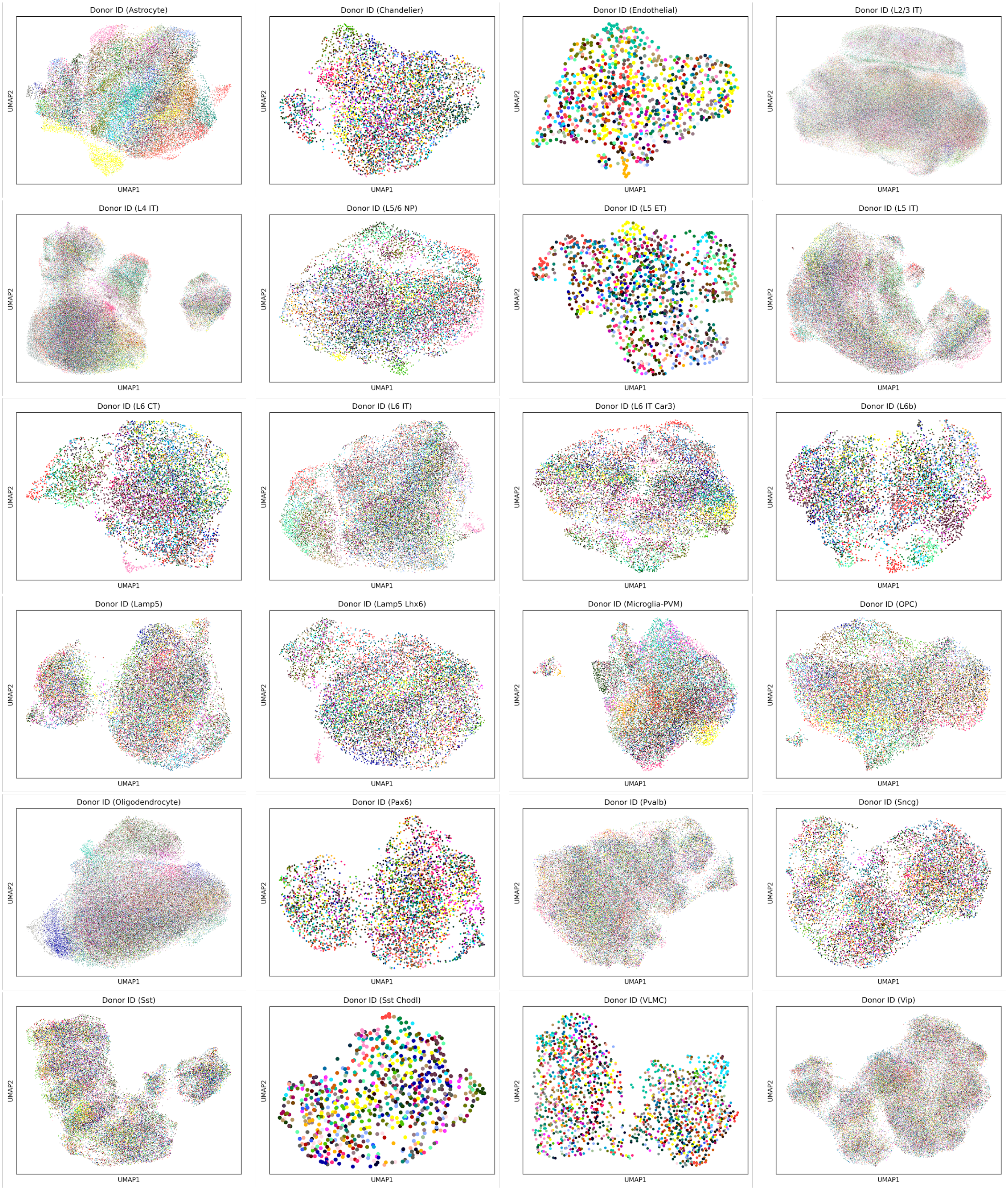


Supplementary Figure 30: **UMAP plots visualizing the cells from the 41 control donors in the SEA-AD brain cell atlas data.** Each plot represents one of the 24 cell types annotated in the SEA-AD brain cell atlas data. Different colors in each plot represent different donors.

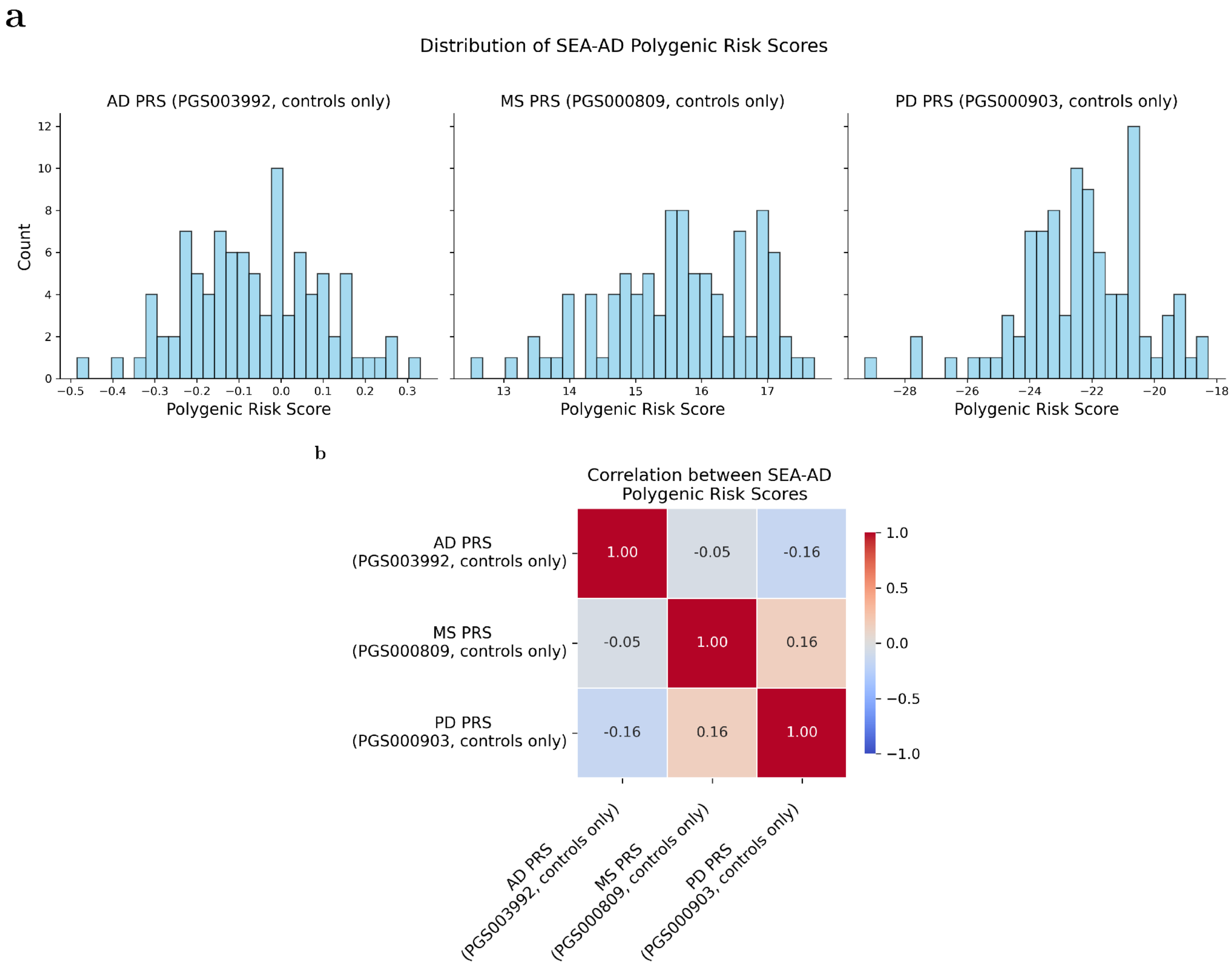


Supplementary Figure 31: **Distribution of polygenic risk scores of 3 neurological disorders.** (**a**) Histograms showing the distribution of the PRSs for AD, MS, and PD, respectively, in control donors. (**b**) Correlation between the PRSs for AD, MS, and PD, across control donors.

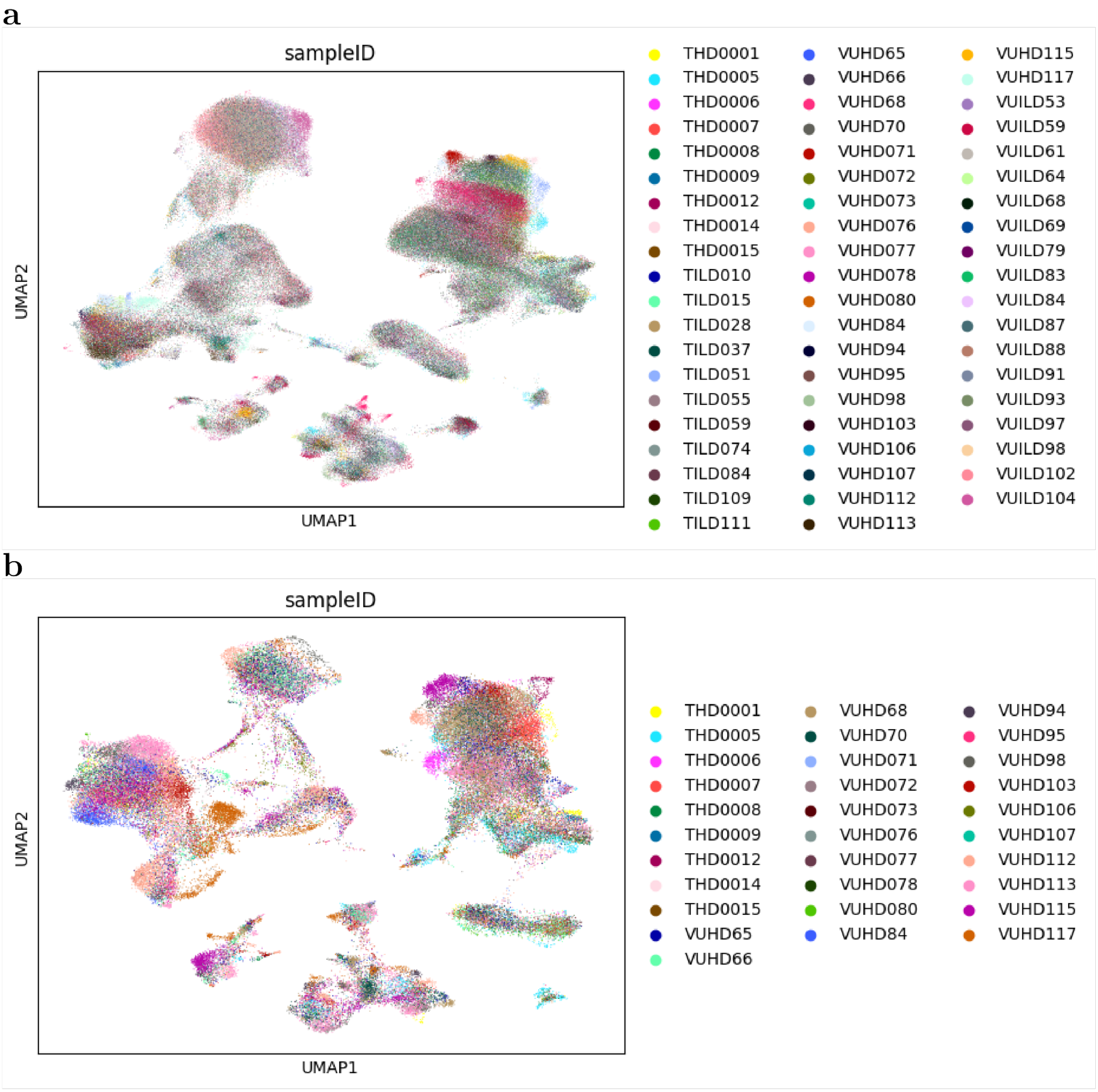


Supplementary Figure 32: **UMAP plots visualizing the cells from TGen lung cell atlas data.** (**a**) UMAP plot for cells from all the 59 donors. (**b**) UMAP plot for cells from the 31 controls donors only. Different colors in each plot represent different donors.

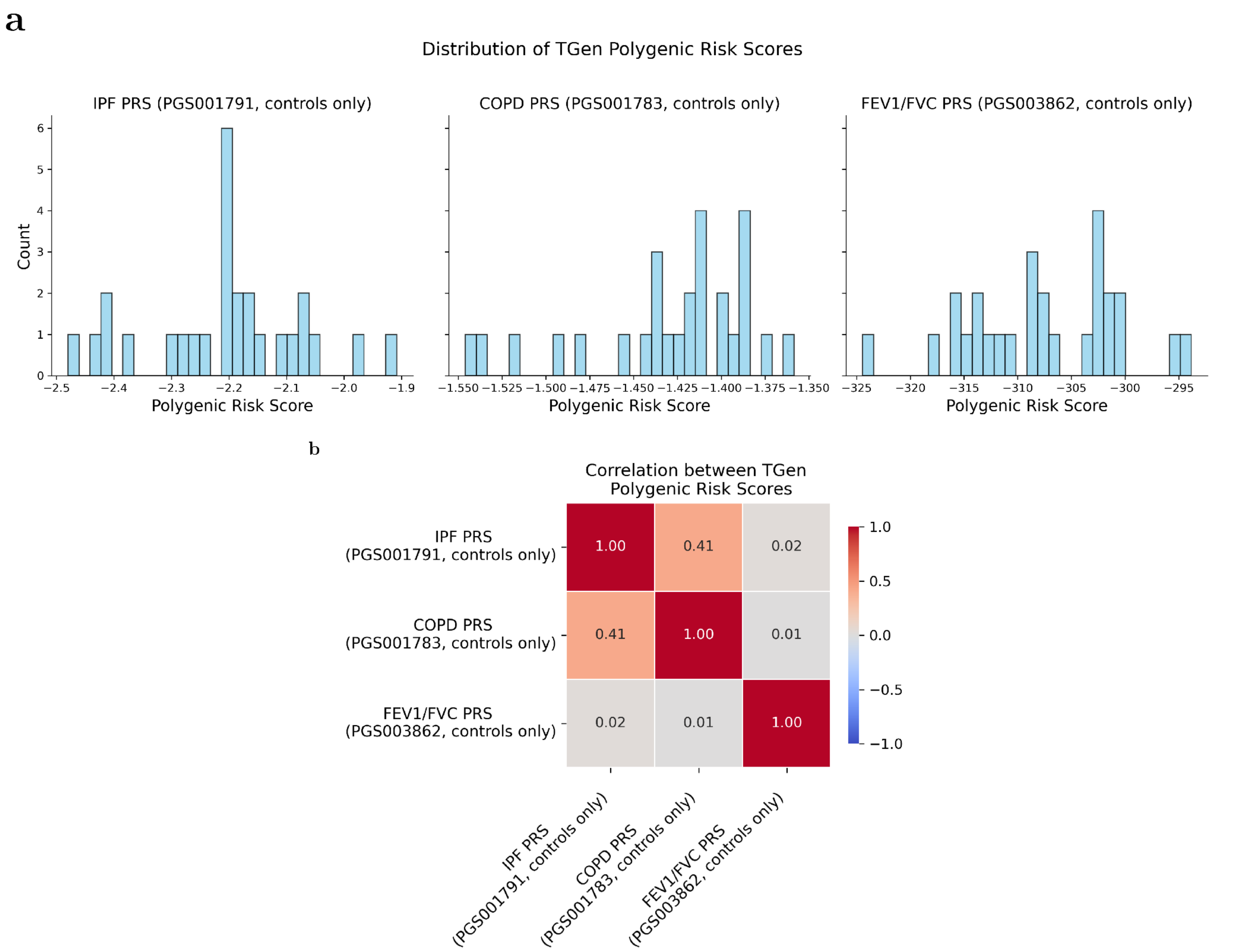


Supplementary Figure 33: **Distribution of polygenic risk scores of 3 respiratory disorders.** (**a**) Histograms showing the distribution of the PRSs for IPF, COPD, and FEV1/FVC, respectively, in control donors. (**b**) Correlation between the PRSs for IPF, COPD, and FEV1/FVC, across control donors.

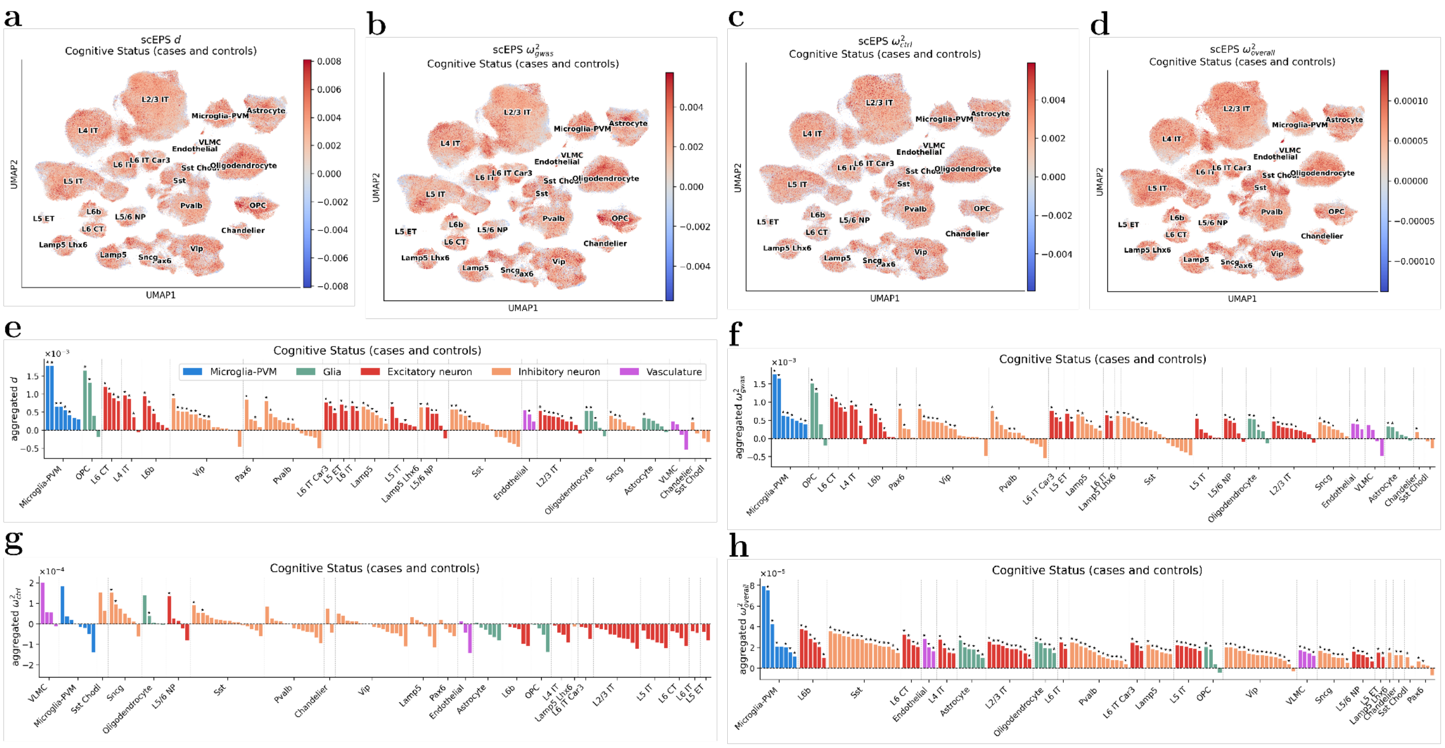


Supplementary Figure 34: **Results from the scEPS analysis of Cognitive Status using the SEA-AD brian cell atlas data.** (**a**, **b**, **c**, **d**) UMAP plots showing the scEPS $d$, $\omega^2{}_{gwas}$, $\omega^2{}_{ctrl}$, and $\omega^2{}_{overall}$ statistics for individual cell neighborhoods for CS, respectively. (**e**, **f**, **g**, **h**) We report the aggregated scEPS $d$, $\omega^2{}_{gwas}$, $\omega^2{}_{ctrl}$, and $\omega^2{}_{overall}$ statistics for CS for 139 brain cell subtypes, respectively. "★" denotes statistical significance: FDR < 0.1 and positivity for aggregated $\omega^2{}_{gwas}$, $\omega^2{}_{ctrl}$, and $\omega^2{}_{overall}$. For aggregated $d$ to reach statistical significance, we also require aggregated $\omega^2{}_{gwas} > 0$. Numerical results are reported in Supplementary Table 9.

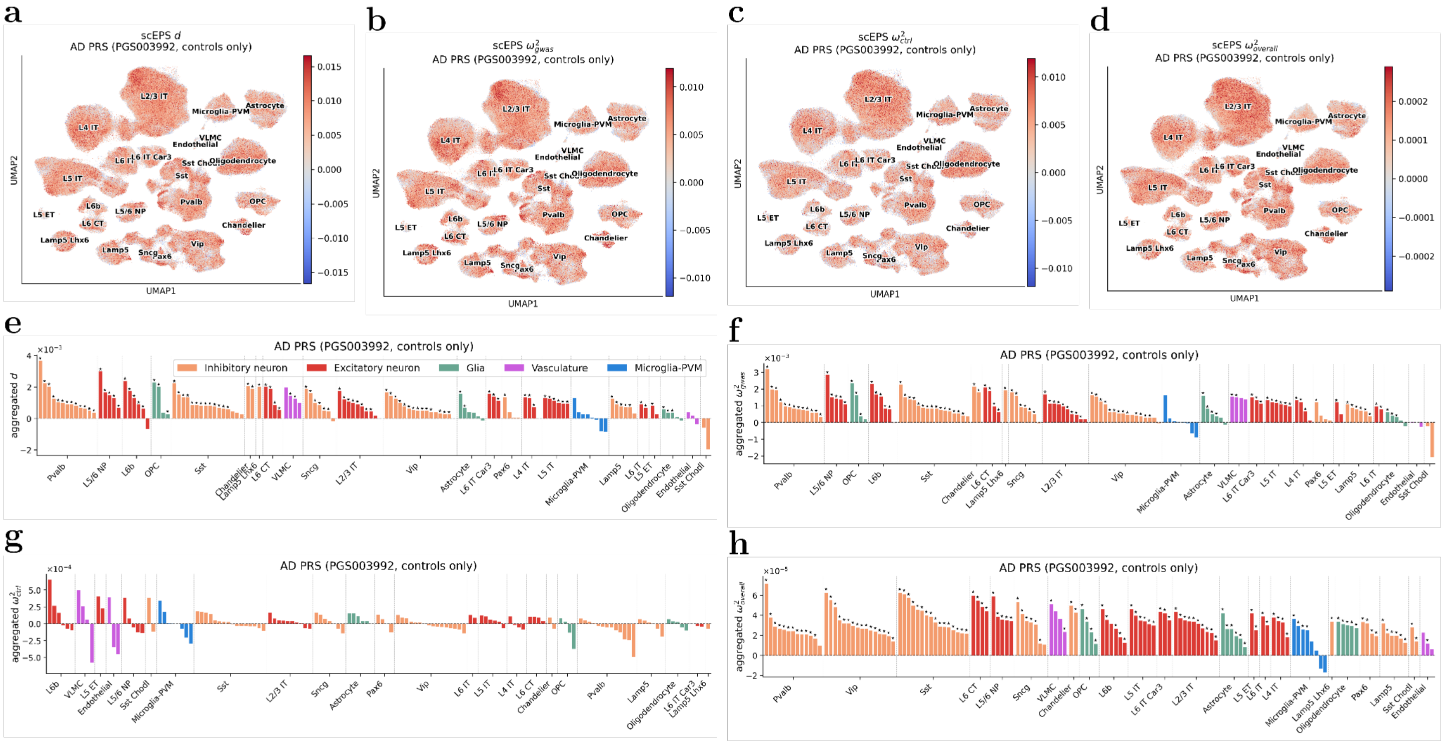


Supplementary Figure 35: **Results from the scEPS analysis of AD PRS in controls using the SEA-AD brain cell atlas data.** (**a**, **b**, **c**, **d**) UMAP plots showing the scEPS $d$, ${\omega^2}_{gwas}$, ${\omega^2}_{ctrl}$, and ${\omega^2}_{overall}$ statistics for individual cell neighborhoods for AD PRS, respectively. (**e**, **f**, **g**, **h**) We report the aggregated scEPS $d$, ${\omega^2}_{gwas}$, ${\omega^2}_{ctrl}$, and ${\omega^2}_{overall}$ statistics for AD PRS for 139 brain cell subtypes, respectively. "★" denotes statistical significance: FDR < 0.1 and positivity for aggregated ${\omega^2}_{gwas}$, ${\omega^2}_{ctrl}$, and ${\omega^2}_{overall}$. For aggregated $d$ to reach statistical significance, we also require aggregated ${\omega^2}_{gwas} > 0$. Numerical results are reported in Supplementary Table 9.

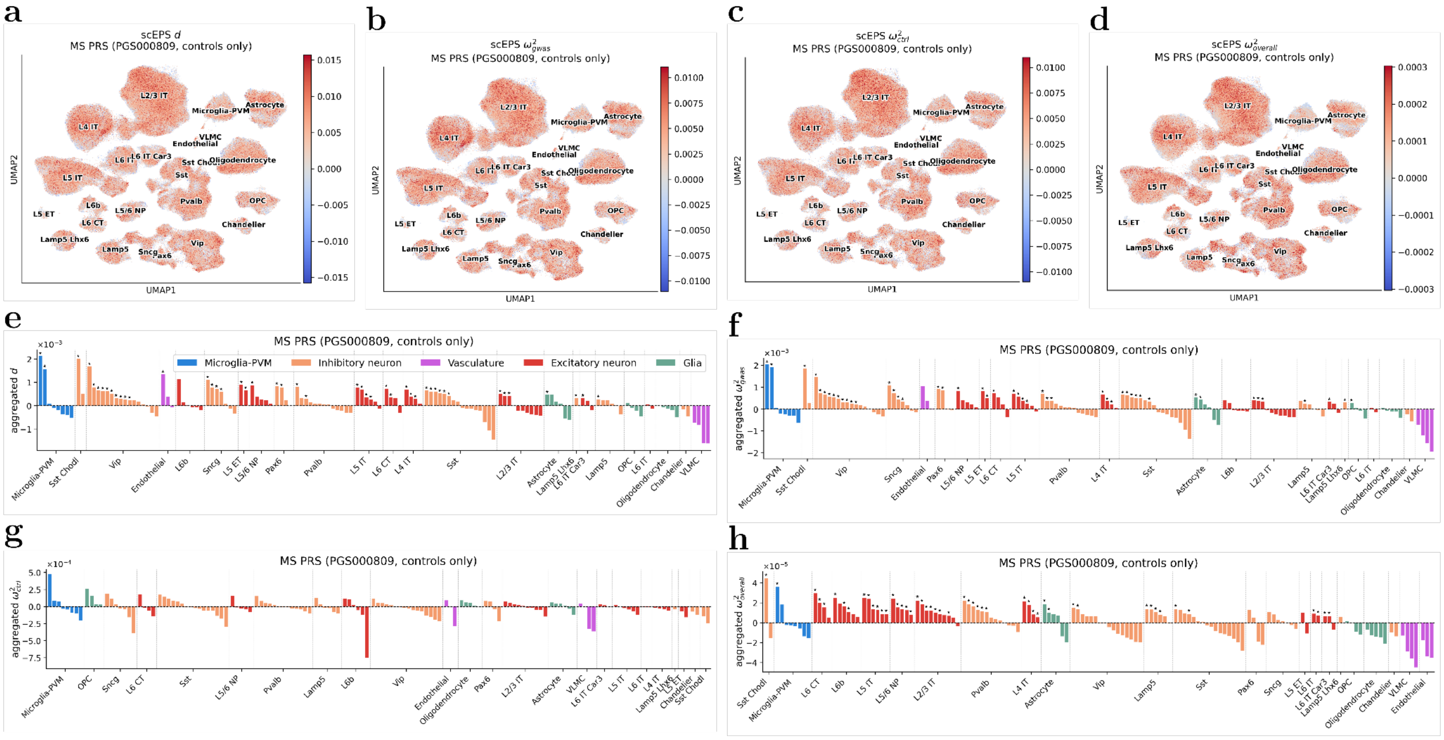


Supplementary Figure 36: **Results from the scEPS analysis of MS PRS in controls using the SEA-AD brain cell atlas data.** (**a**, **b**, **c**, **d**) UMAP plots showing the scEPS $d$, ${\omega^2}_{gwas}$, ${\omega^2}_{ctrl}$, and ${\omega^2}_{overall}$ statistics for individual cell neighborhoods for MS PRS, respectively. (**e**, **f**, **g**, **h**) We report the aggregated scEPS $d$, ${\omega^2}_{gwas}$, ${\omega^2}_{ctrl}$, and ${\omega^2}_{overall}$ statistics for MS PRS for 139 brain cell subtypes, respectively. "★" denotes statistical significance: FDR < 0.1 and positivity for aggregated ${\omega^2}_{gwas}$, ${\omega^2}_{ctrl}$, and ${\omega^2}_{overall}$. For aggregated $d$ to reach statistical significance, we also require aggregated ${\omega^2}_{gwas} > 0$. Numerical results are reported in Supplementary Table 9.

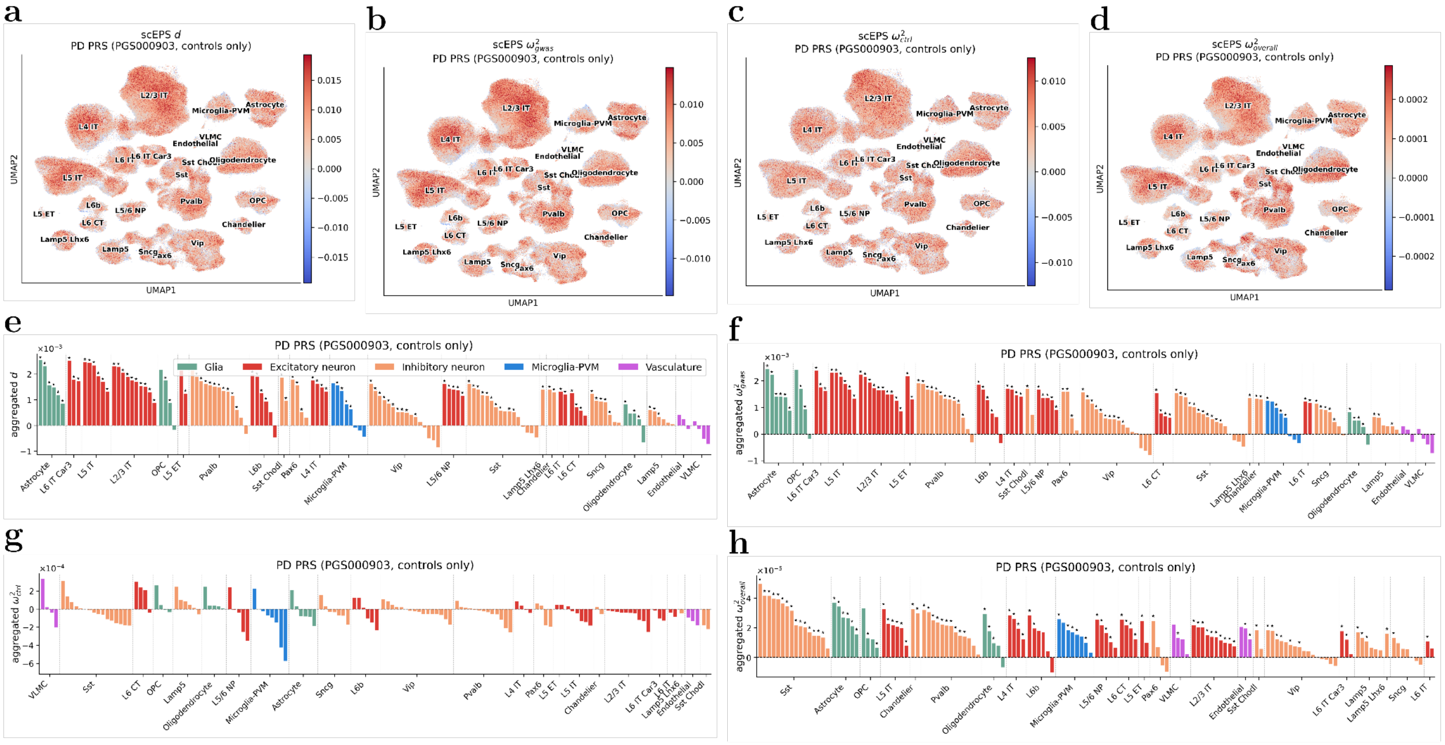


Supplementary Figure 37: **Results from the scEPS analysis of PD PRS in controls using the SEA-AD brain cell atlas data.** (**a**, **b**, **c**, **d**) UMAP plots showing the scEPS $d$, $\omega^2{}_{gwas}$, $\omega^2{}_{ctrl}$, and $\omega^2{}_{overall}$ statistics for individual cell neighborhoods for PD PRS, respectively. (**e**, **f**, **g**, **h**) We report the aggregated scEPS $d$, $\omega^2{}_{gwas}$, $\omega^2{}_{ctrl}$, and $\omega^2{}_{overall}$ statistics for PD PRS for 139 brain cell subtypes, respectively. "★" denotes statistical significance: FDR < 0.1 and positivity for aggregated $\omega^2{}_{gwas}$, $\omega^2{}_{ctrl}$, and $\omega^2{}_{overall}$. For aggregated $d$ to reach statistical significance, we also require aggregated $\omega^2{}_{gwas} > 0$. Numerical results are reported in Supplementary Table 9.

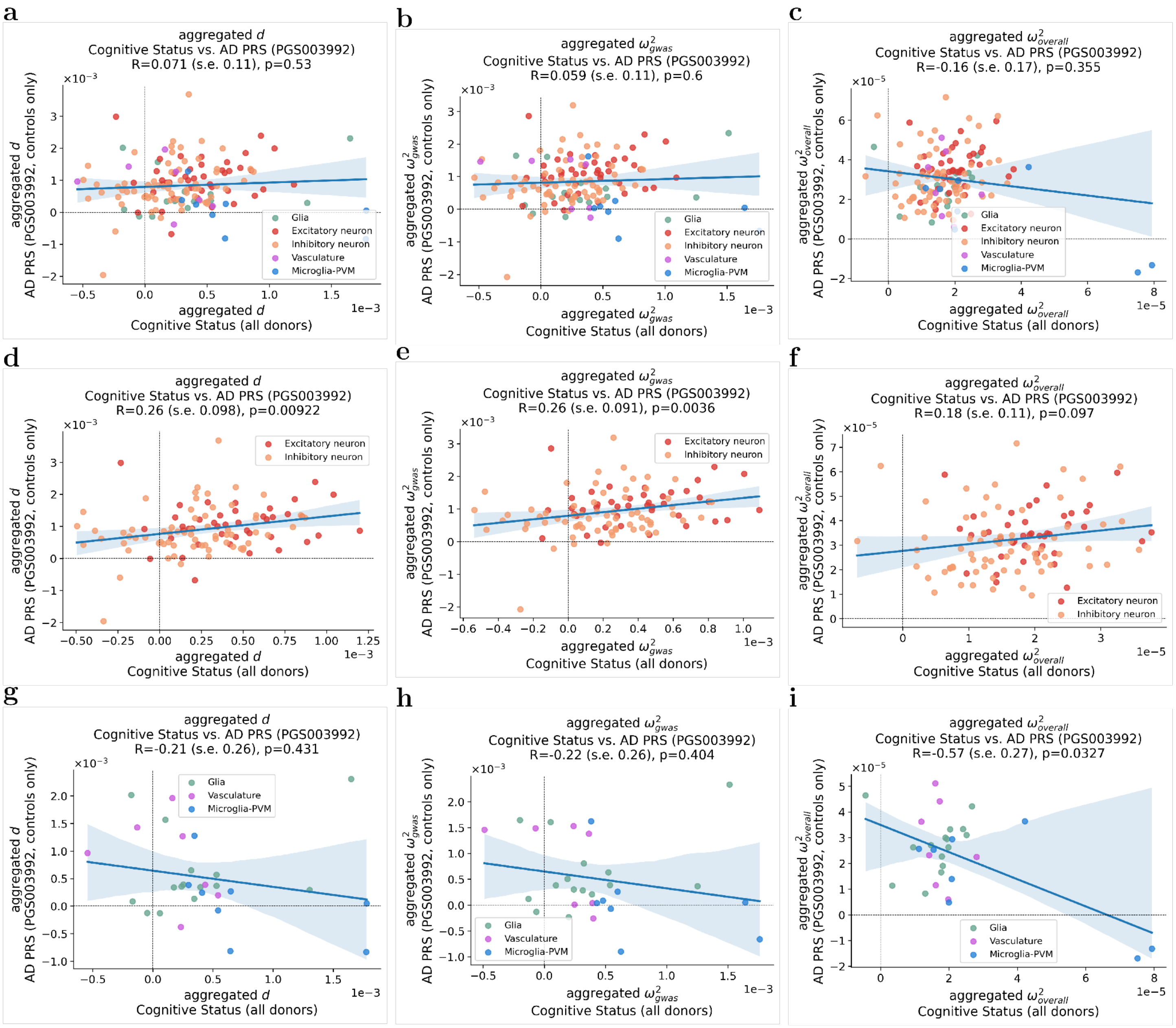


Supplementary Figure 38: **Comparisons of aggregated scEPS statistics of CS and AD PRS.** (**a**, **b**, **c**) Comparisons of aggregated $d$, $\omega^2{}_{gwas}$, and $\omega^2{}_{overall}$ between CS and AD PRS, across all SEA-AD brain cell subtypes. (**d**, **e**, **f**) Comparisons of aggregated $d$, $\omega^2{}_{gwas}$, and $\omega^2{}_{overall}$ between CS and AD PRS, across SEA-AD neuronal cell subtypes. (**g**, **h**, **i**) Comparisons of aggregated $d$, $\omega^2{}_{gwas}$, and $\omega^2{}_{overall}$ between CS and AD PRS, across SEA-AD glial cell subtypes. Standard errors and p-values testing the R were based on statistical bootstrap using 1,000 bootstrapped samples. Shaded regions of the regression plots represent the 95% confidence interval.

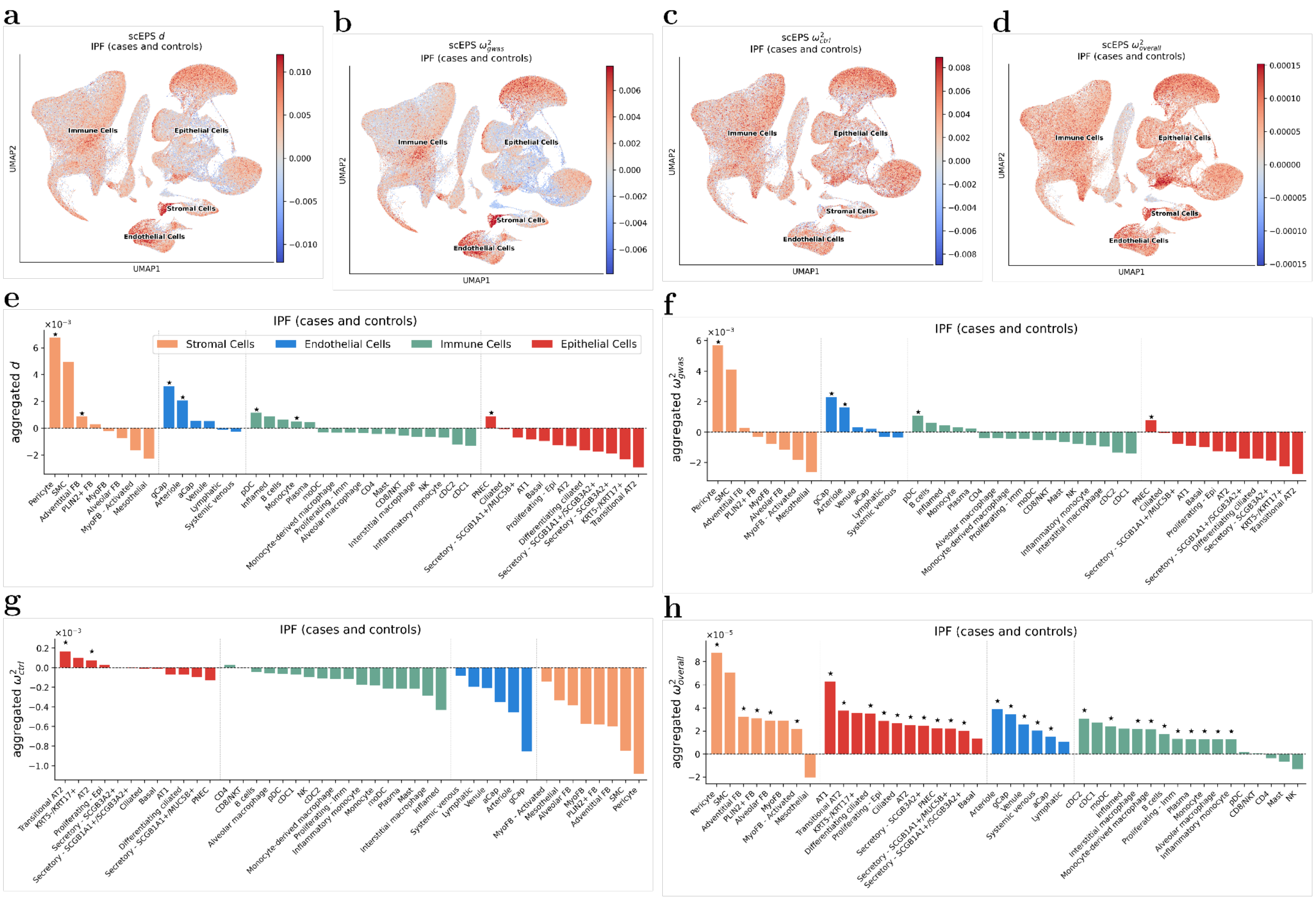


Supplementary Figure 39: **Results from the scEPS analysis of IPF using the TGen lung cell atlas data.** (**a**, **b**, **c**, **d**) UMAP plots showing the scEPS $d$, ${\omega^2}_{gwas}$, ${\omega^2}_{ctrl}$, and ${\omega^2}_{overall}$ statistics for individual cell neighborhoods for IPF, respectively. (**e**, **f**, **g**, **h**) We report the aggregated scEPS $d$, ${\omega^2}_{gwas}$, ${\omega^2}_{ctrl}$, and ${\omega^2}_{overall}$ statistics for IPF for 43 lung cell subtypes, respectively. "★" denotes statistical significance: FDR < 0.1 and positivity for aggregated ${\omega^2}_{gwas}$, ${\omega^2}_{ctrl}$, and ${\omega^2}_{overall}$. For aggregated $d$ to reach statistical significance, we also require aggregated ${\omega^2}_{gwas} > 0$. Numerical results are reported in Supplementary Table 10.

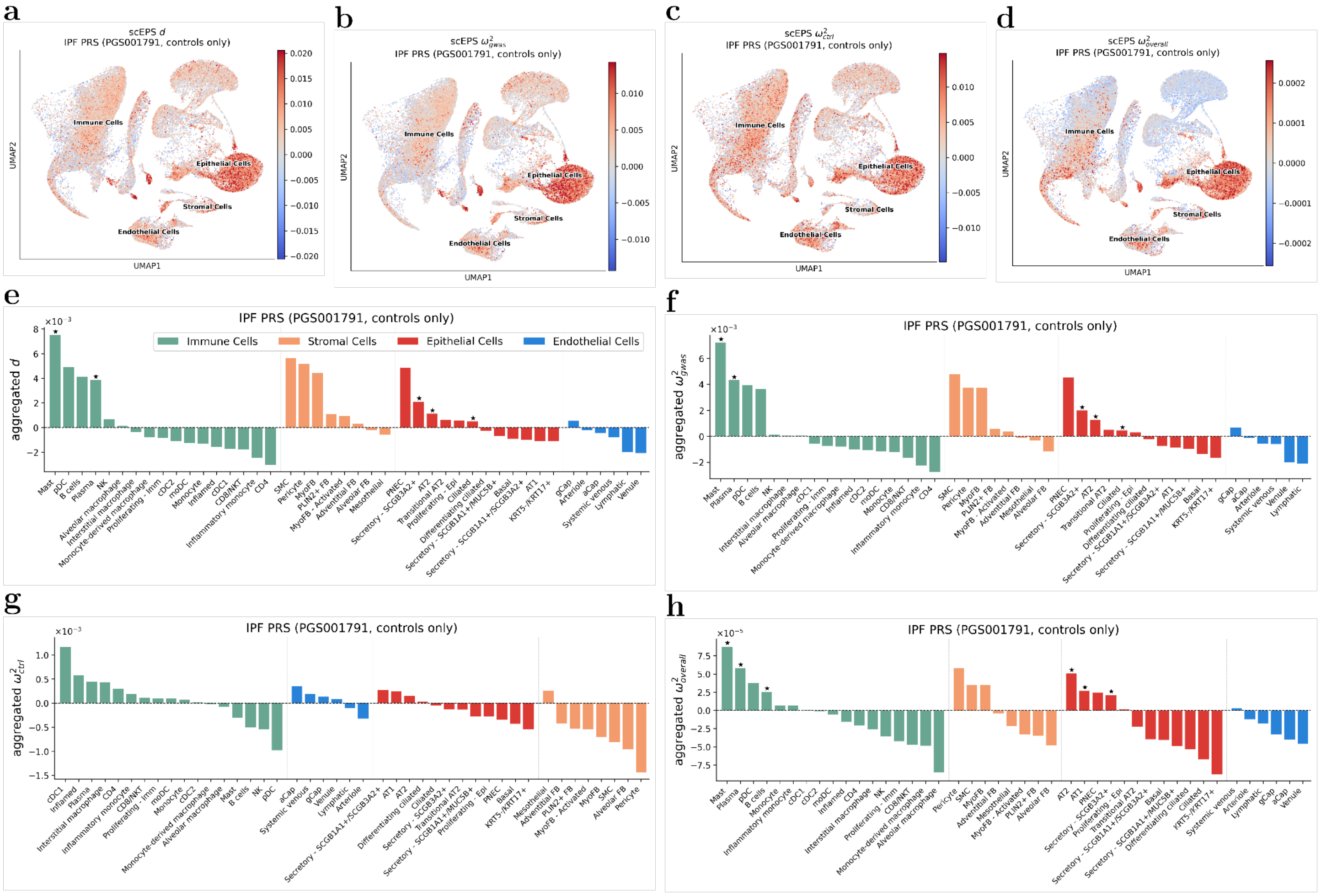


Supplementary Figure 40: **Results from the scEPS analysis of IPF PRS in controls using the TGen lung cell atlas data.** (**a**, **b**, **c**, **d**) UMAP plots showing the scEPS $d$, $\omega^2_{gwas}$, $\omega^2_{ctrl}$, and $\omega^2_{overall}$ statistics for individual cell neighborhoods for IPF PRS, respectively. (**e**, **f**, **g**, **h**) We report the aggregated scEPS $d$, $\omega^2_{gwas}$, $\omega^2_{ctrl}$, and $\omega^2_{overall}$ statistics for IPF PRS for 43 lung cell subtypes, respectively. "★" denotes statistical significance: FDR < 0.1 and positivity for aggregated $\omega^2_{gwas}$, $\omega^2_{ctrl}$, and $\omega^2_{overall}$. For aggregated $d$ to reach statistical significance, we also require aggregated $\omega^2_{gwas} > 0$. Numerical results are reported in Supplementary Table 10.

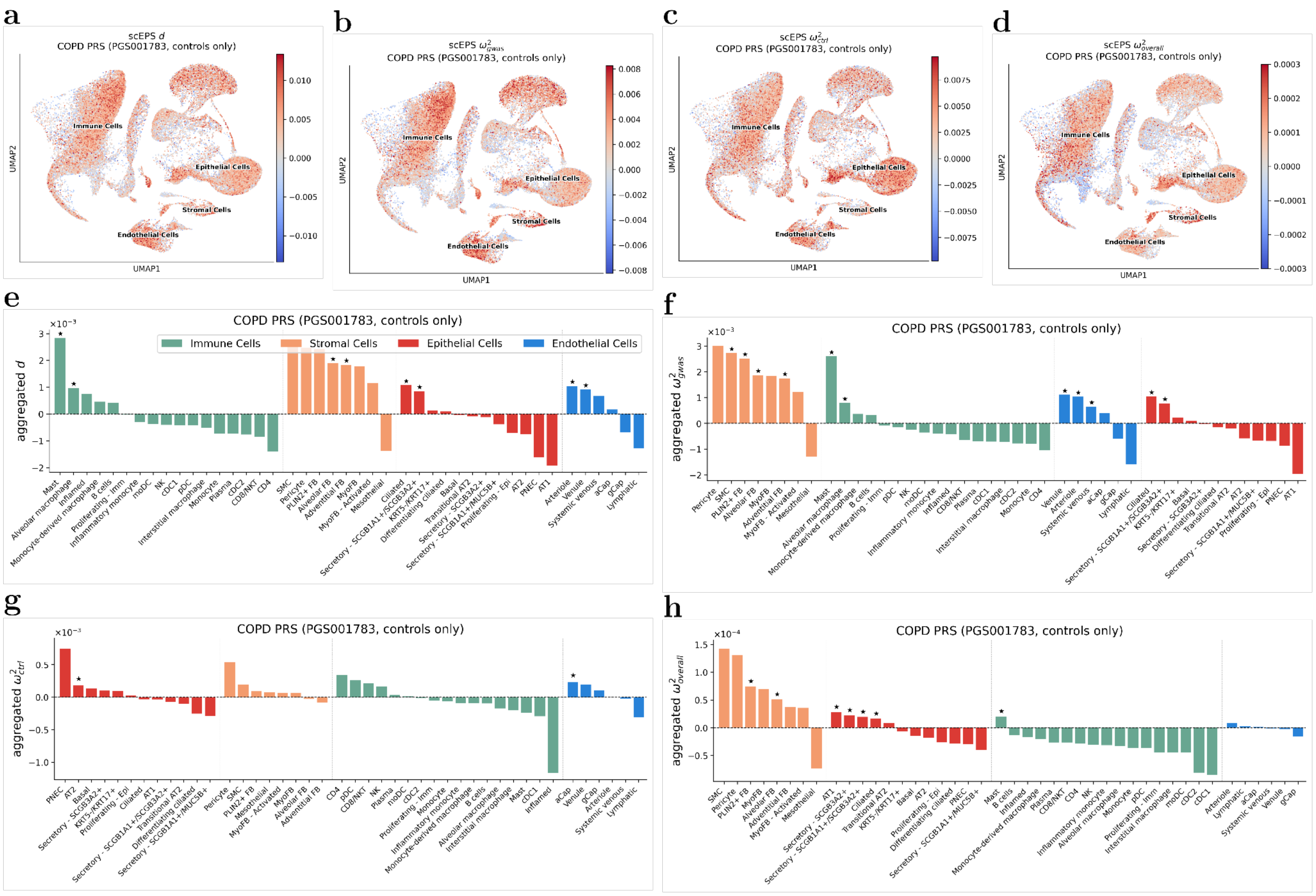


Supplementary Figure 41: **Results from the scEPS analysis of COPD PRS in controls using the TGen lung cell atlas data.** (**a**, **b**, **c**, **d**) UMAP plots showing the scEPS $d$, $\omega^2_{gwas}$, $\omega^2_{ctrl}$, and $\omega^2_{overall}$ statistics for individual cell neighborhoods for COPD PRS, respectively. (**e**, **f**, **g**, **h**) We report the aggregated scEPS $d$, $\omega^2_{gwas}$, $\omega^2_{ctrl}$, and $\omega^2_{overall}$ statistics for COPD PRS for 43 lung cell subtypes, respectively. "★" denotes statistical significance: FDR < 0.1 and positivity for aggregated $\omega^2_{gwas}$, $\omega^2_{ctrl}$, and $\omega^2_{overall}$. For aggregated $d$ to reach statistical significance, we also require aggregated $\omega^2_{gwas} > 0$. Numerical results are reported in Supplementary Table 10.

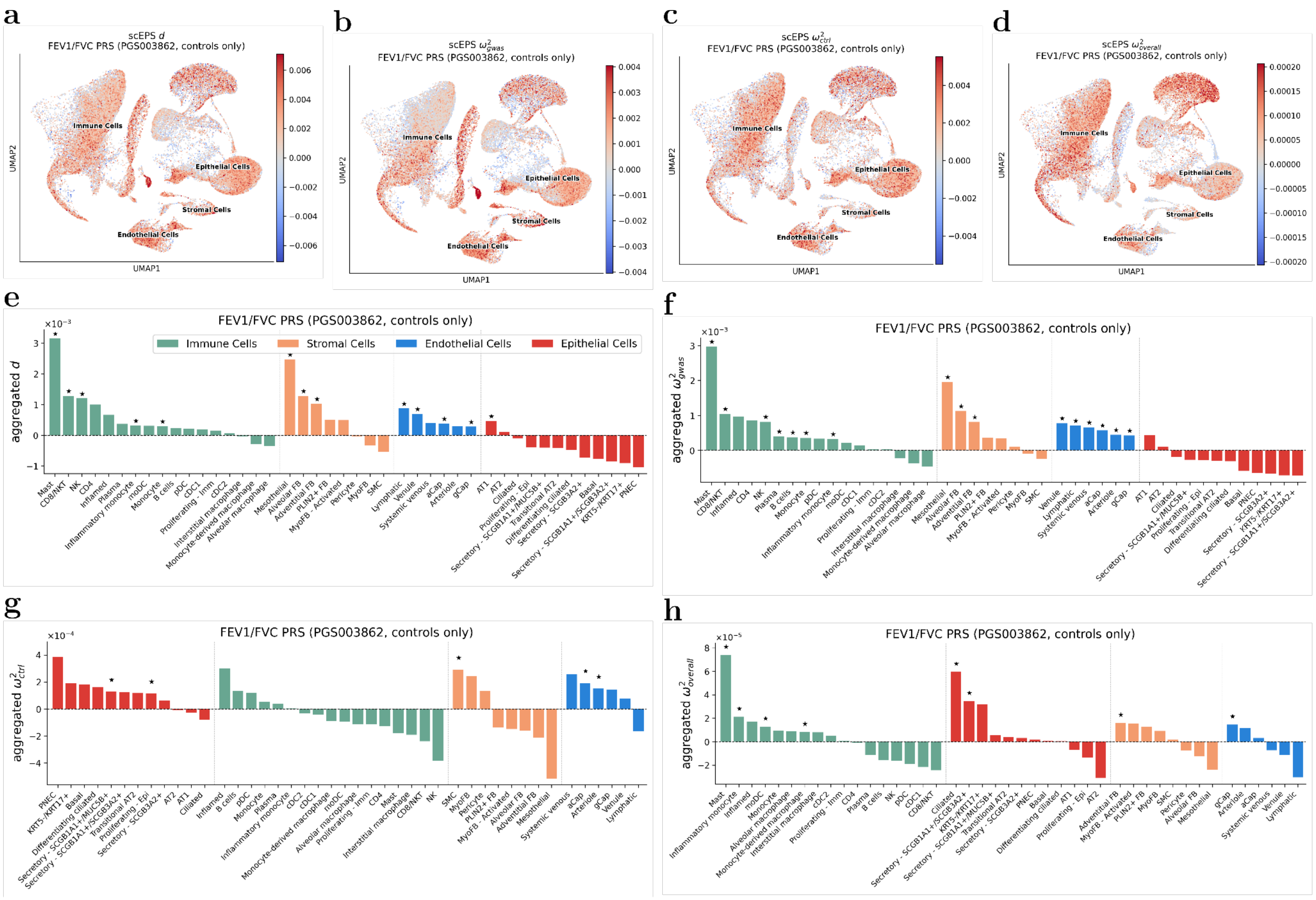


Supplementary Figure 42: **Results from the scEPS analysis of FEV1/FVC PRS in controls using the TGen lung cell atlas data.** (**a**, **b**, **c**, **d**) UMAP plots showing the scEPS $d$, $\omega^2{}_{gwas}$, $\omega^2{}_{ctrl}$, and $\omega^2{}_{overall}$ statistics for individual cell neighborhoods for FEV1/FVC PRS, respectively. (**e**, **f**, **g**, **h**) We report the aggregated scEPS $d$, $\omega^2{}_{gwas}$, $\omega^2{}_{ctrl}$, and $\omega^2{}_{overall}$ statistics for FEV1/FVC PRS for 43 lung cell subtypes, respectively. "★" denotes statistical significance: FDR < 0.1 and positivity for aggregated $\omega^2{}_{gwas}$, $\omega^2{}_{ctrl}$, and $\omega^2{}_{overall}$. For aggregated $d$ to reach statistical significance, we also require aggregated $\omega^2{}_{gwas} > 0$. Numerical results are reported in Supplementary Table 10.

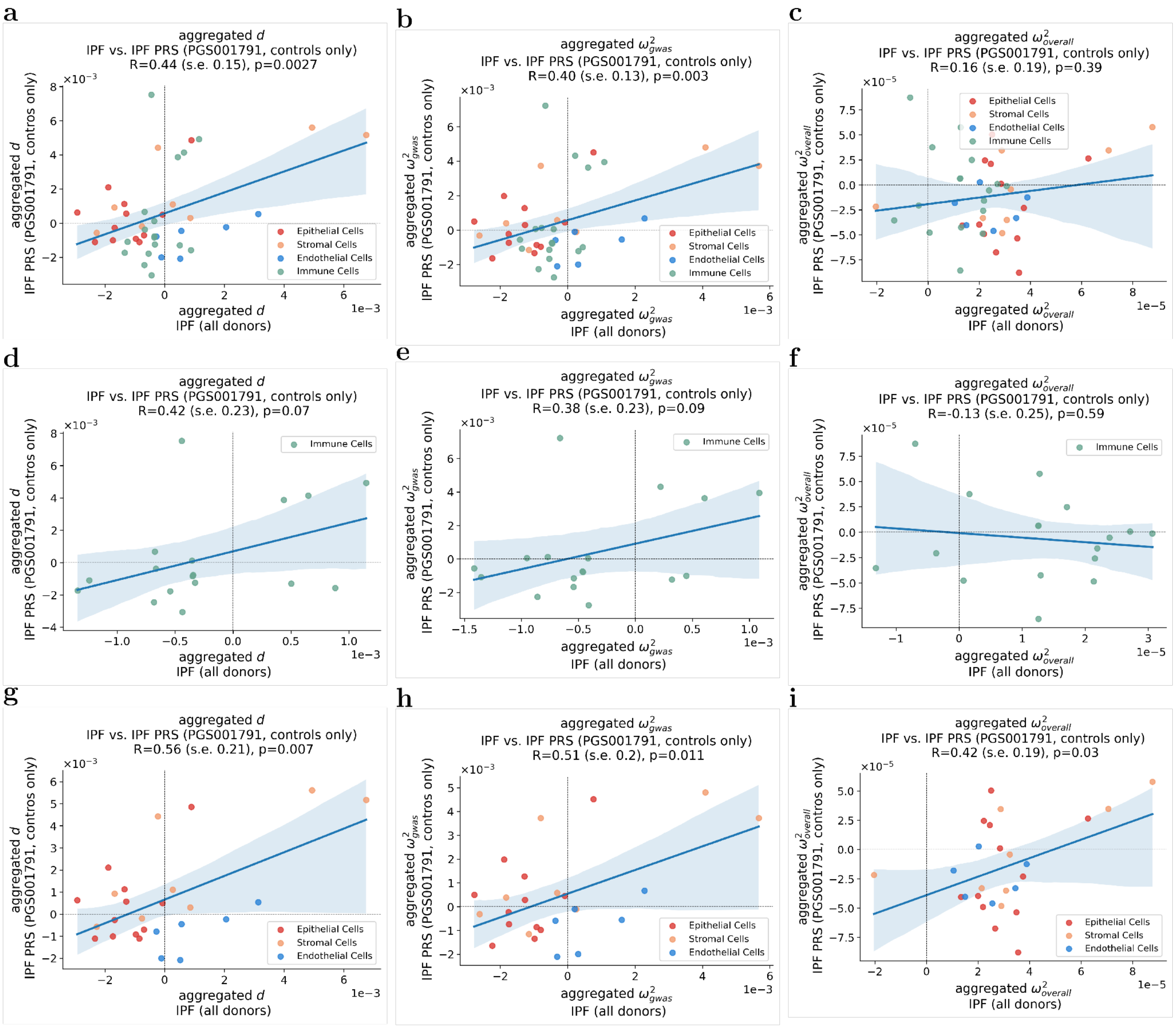


Supplementary Figure 43: **Comparisons of aggregated scEPS statistics for IPF and IPF PRS.** (**a**, **b**, **c**) Comparisons of aggregated $d$, $\omega^2{}_{gwas}$, and $\omega^2{}_{overall}$ between IPF and IPF PRS, across all TGen lung cell subtypes. (**d**, **e**, **f**) Comparisons of aggregated $d$, $\omega^2{}_{gwas}$, and $\omega^2{}_{overall}$ between IPF and IPF PRS, across lug immune cell subtypes. (**g**, **h**, **i**) Comparisons of aggregated $d$, $\omega^2{}_{gwas}$, and $\omega^2{}_{overall}$ between IPF and IPF PRS, across non-immune cell subtypes. Standard errors and p-values testing the R were based on statistical bootstrap using 1,000 bootstrapped samples. Shaded regions of the regression plots represent the 95% confidence interval.

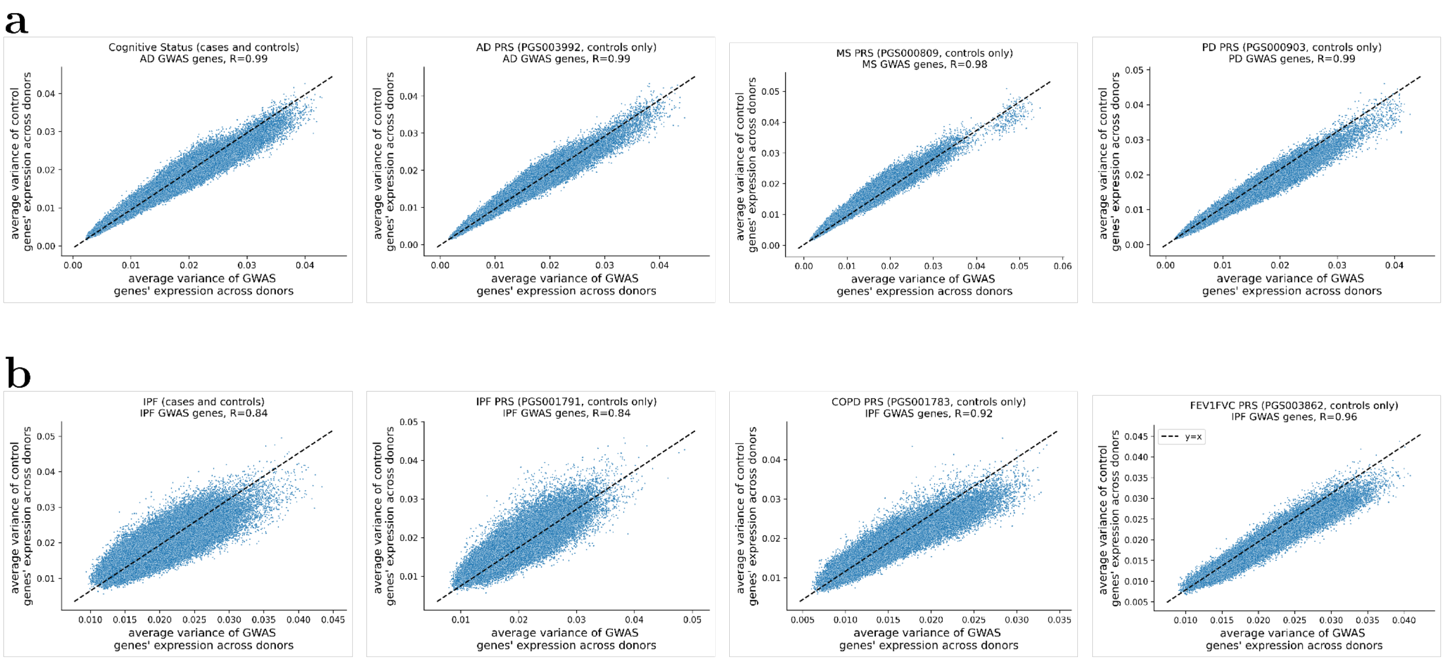


Supplementary Figure 44: **Comparison of the variance of the expression of GWAS genes and control genes.** (**a**, **b**) Results for the 4 neurological and 4 respiratory disorders, respectively. Each dot in the scatter plots represents a cell neighborhood. Variance of the gene expression was calculated across donors and averaged across genes.

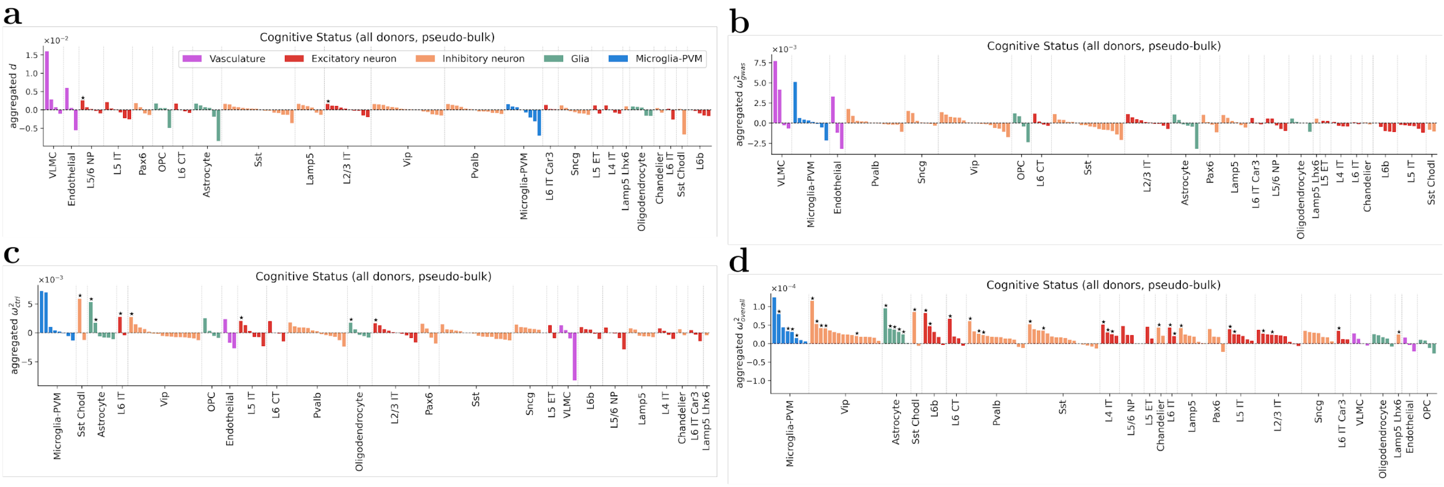


Supplementary Figure 45: **scEPS statistics across for CS obtained using pseudo-bulked genes expression of 138 brain cell types.** (**a**, **b**, **c**, **d**) We report the scEPS $d$, ${\omega^2}_{gwas}$, ${\omega^2}_{ctrl}$, and ${\omega^2}_{overall}$ statistics obtained using the pseudo-bulk gene expression data, respectively. “★” denotes statistical significance: FDR < 0.1 and positivity for ${\omega^2}_{gwas}$, ${\omega^2}_{ctrl}$, and ${\omega^2}_{overall}$. For $d$ to reach statistical significance, we also require ${\omega^2}_{gwas} > 0$. Numerical results are reported in Supplementary Table 11.

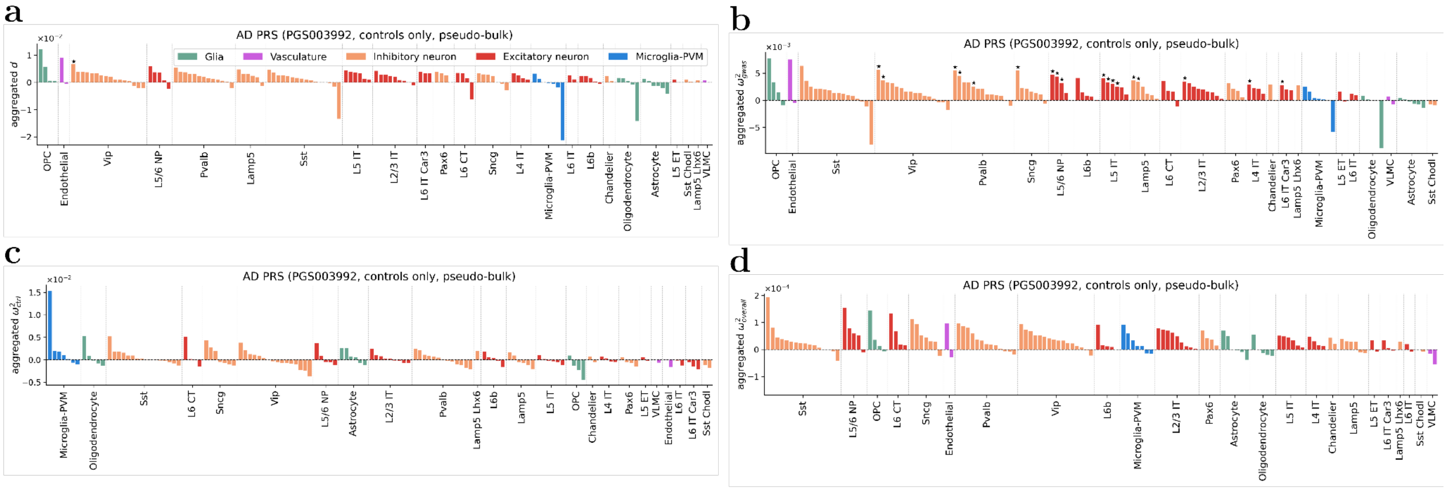


Supplementary Figure 46: **scEPS statistics of CS obtained using pseudo-bulked gene expression of 133 brain cell types.** (**a**, **b**, **c**, **d**) We report the scEPS $d$, ${\omega^2}_{gwas}$, ${\omega^2}_{ctrl}$, and ${\omega^2}_{overall}$ statistics obtained using the pseudo-bulk gene expression data, respectively. "★" denotes statistical significance: FDR < 0.1 and positivity for ${\omega^2}_{gwas}$, ${\omega^2}_{ctrl}$, and ${\omega^2}_{overall}$. For $d$ to reach statistical significance, we also require ${\omega^2}_{gwas} > 0$. Numerical results are reported in Supplementary Table 11.

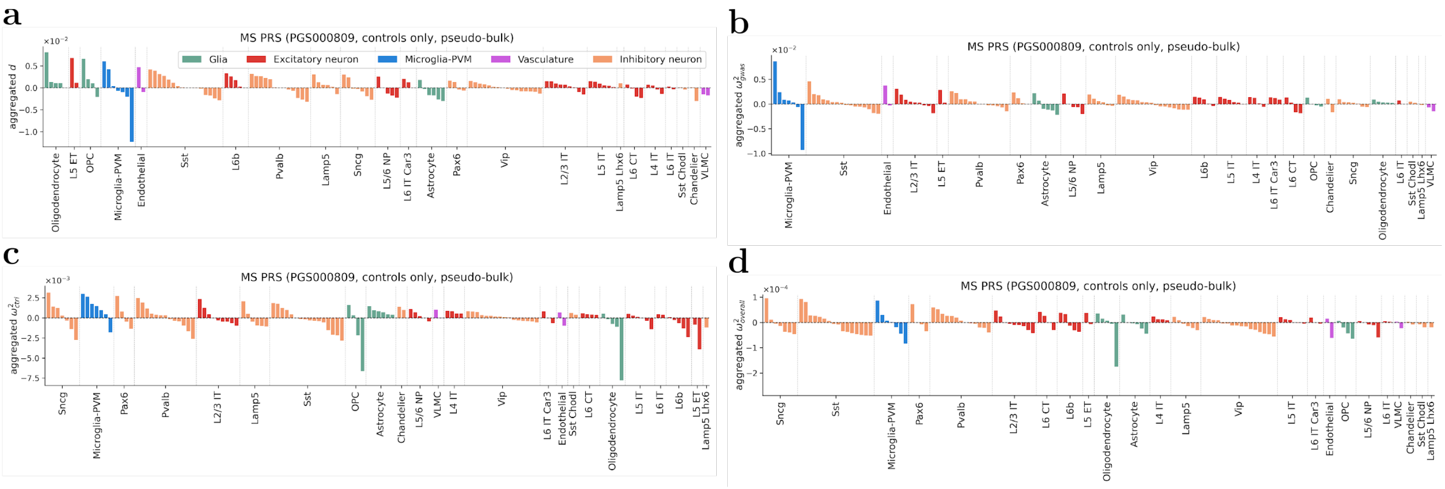


Supplementary Figure 47: **scEPS statistics of MS PRS obtained using pseudo-bulked gene expression of 133 brain cell types.** (**a**, **b**, **c**, **d**) We report the scEPS $d$, $\omega^2{}_{gwas}$, $\omega^2{}_{ctrl}$, and $\omega^2{}_{overall}$ statistics obtained using the pseudo-bulk gene expression data, respectively. "★" denotes statistical significance: FDR < 0.1 and positivity for $\omega^2{}_{gwas}$, $\omega^2{}_{ctrl}$, and $\omega^2{}_{overall}$. For $d$ to reach statistical significance, we also require $\omega^2{}_{gwas} > 0$. Numerical results are reported in Supplementary Table 11.

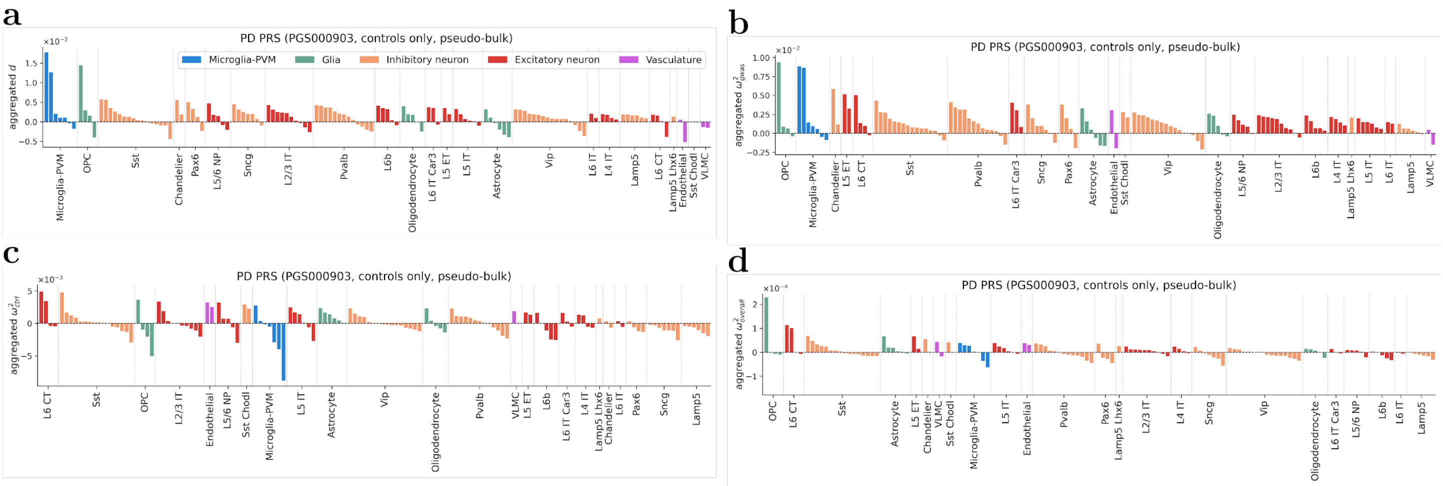


Supplementary Figure 48: **scEPS statistics of PD PRS obtained using pseudo-bulked gene expression of 134 brain cell types.** (**a**, **b**, **c**, **d**) We report the scEPS $d$, ${\omega^2}_{gwas}$, ${\omega^2}_{ctrl}$, and ${\omega^2}_{overall}$ statistics obtained using the pseudo-bulk gene expression data, respectively. "★" denotes statistical significance: FDR < 0.1 and positivity for ${\omega^2}_{gwas}$, ${\omega^2}_{ctrl}$, and ${\omega^2}_{overall}$. For $d$ to reach statistical significance, we also require ${\omega^2}_{gwas} > 0$. Numerical results are reported in Supplementary Table 11.

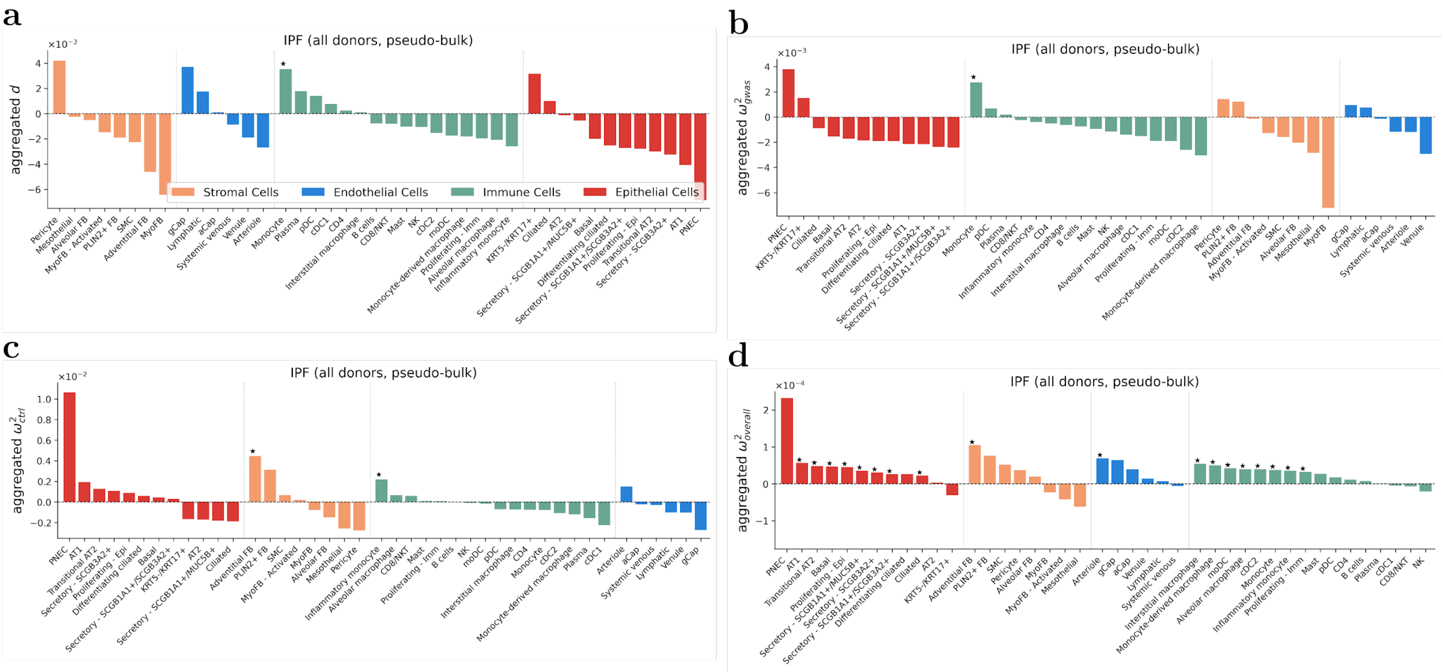


Supplementary Figure 49: **scEPS statistics of IPF obtained using pseudo-bulked gene expression of 42 lung cell types.** (**a**, **b**, **c**, **d**) We report the scEPS $d$, ${\omega^2}_{gwas}$, ${\omega^2}_{ctrl}$, and ${\omega^2}_{overall}$ statistics obtained using the pseudo-bulk gene expression data, respectively. "★" denotes statistical significance: FDR < 0.1 and positivity for ${\omega^2}_{gwas}$, ${\omega^2}_{ctrl}$, and ${\omega^2}_{overall}$. For $d$ to reach statistical significance, we also require ${\omega^2}_{gwas} > 0$. Numerical results are reported in Supplementary Table 12.

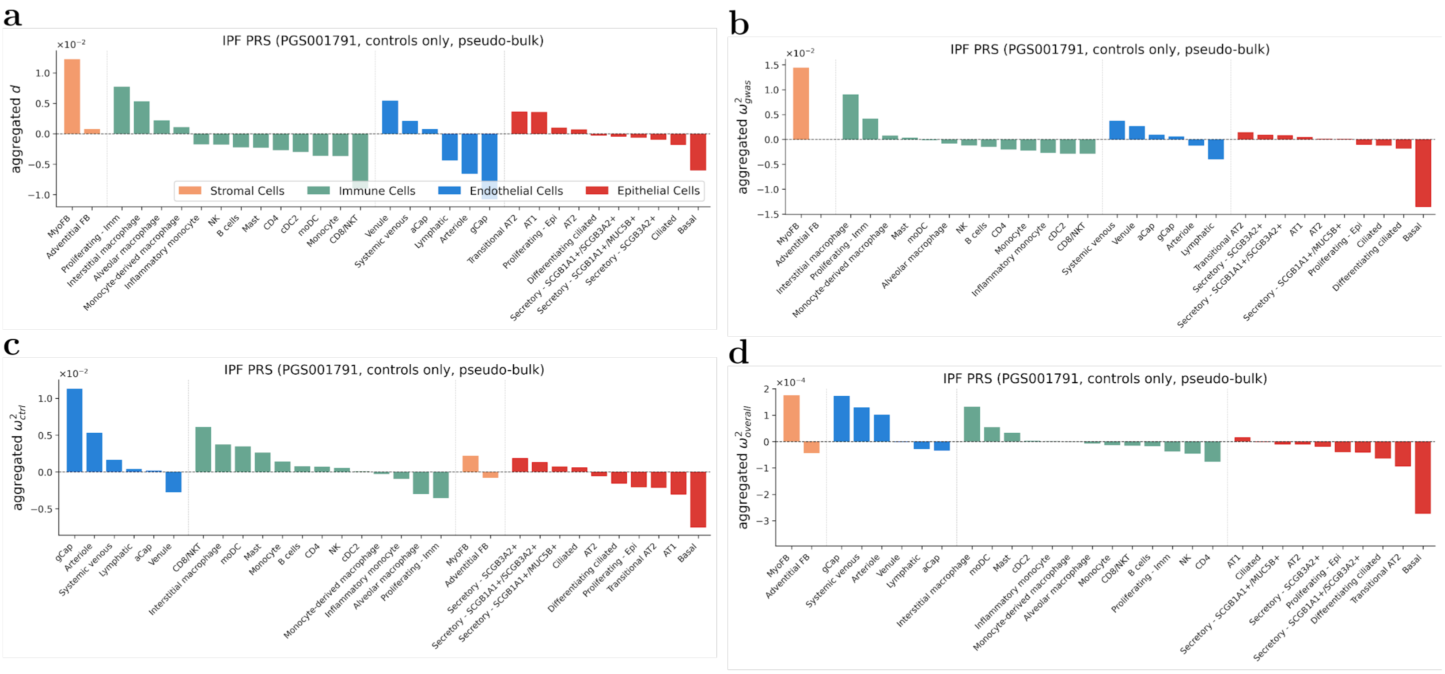


Supplementary Figure 50: **scEPS statistics of IPF PRS obtained using pseudo-bulked gene expression of 31 lung cell types.** (**a**, **b**, **c**, **d**) We report the scEPS $d$, ${\omega^2}_{gwas}$, ${\omega^2}_{ctrl}$, and ${\omega^2}_{overall}$ statistics obtained using the pseudo-bulk gene expression data, respectively. "★" denotes statistical significance: FDR < 0.1 and positivity for ${\omega^2}_{gwas}$, ${\omega^2}_{ctrl}$, and ${\omega^2}_{overall}$. For $d$ to reach statistical significance, we also require ${\omega^2}_{gwas} > 0$. Numerical results are reported in Supplementary Table 12.

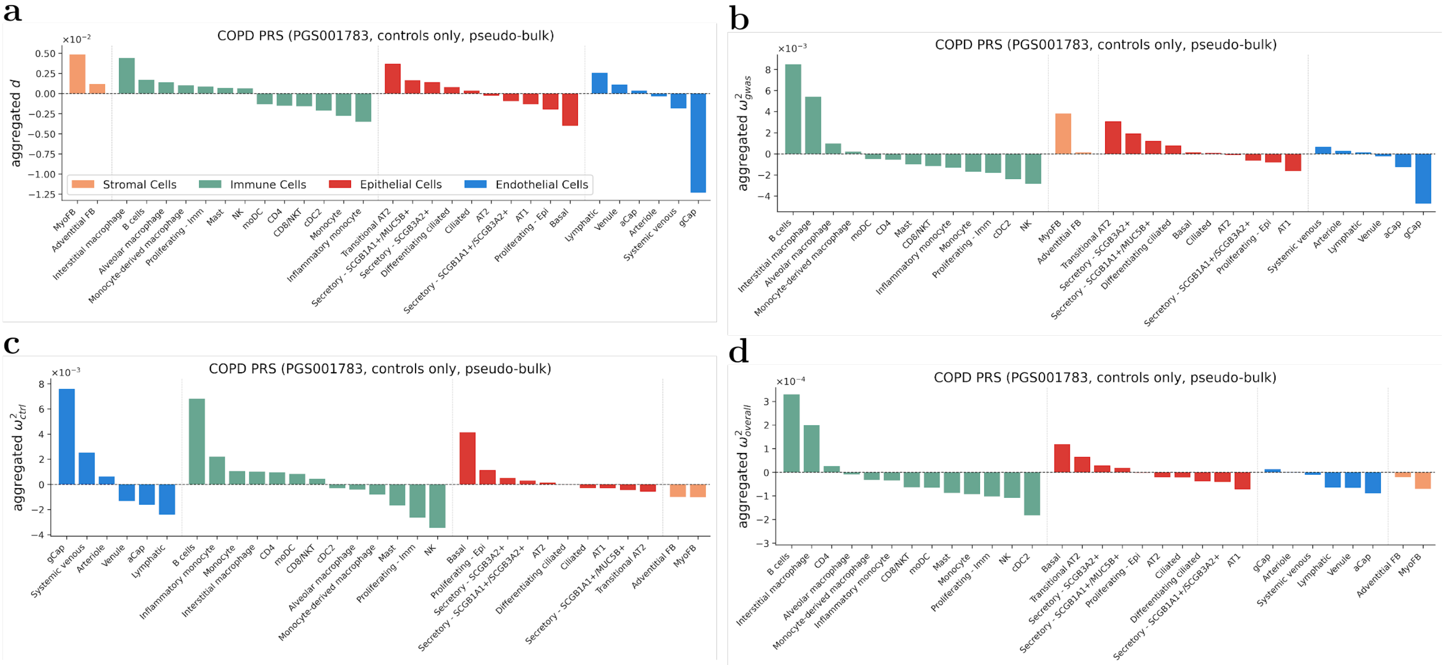


Supplementary Figure 51: **scEPS statistics of IPF PRS obtained using pseudo-bulked gene expression of 31 lung cell types.** (**a**, **b**, **c**, **d**) We report the scEPS $d$, ${\omega^2}_{gwas}$, ${\omega^2}_{ctrl}$, and ${\omega^2}_{overall}$ statistics obtained using the pseudo-bulk gene expression data, respectively. "★" denotes statistical significance: FDR < 0.1 and positivity for ${\omega^2}_{gwas}$, ${\omega^2}_{ctrl}$, and ${\omega^2}_{overall}$. For $d$ to reach statistical significance, we also require ${\omega^2}_{gwas} > 0$. Numerical results are reported in Supplementary Table 12.

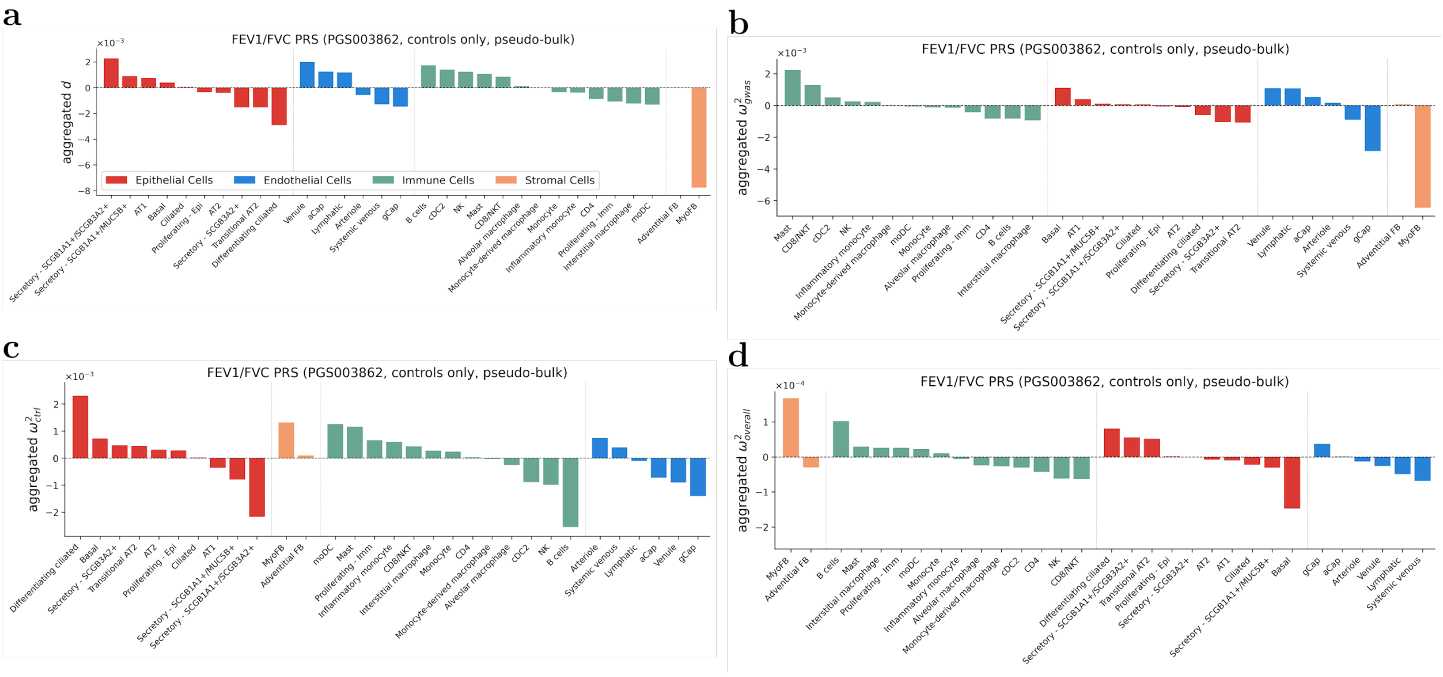


Supplementary Figure 52: **scEPS statistics of FEV1/FVC PRS obtained using pseudo-bulked gene expression of 31 lung cell types.** (**a**, **b**, **c**, **d**) We report the scEPS $d$, ${\omega^2}_{gwas}$, ${\omega^2}_{ctrl}$, and ${\omega^2}_{overall}$ statistics obtained using the pseudo-bulk gene expression data, respectively. "★" denotes statistical significance: FDR < 0.1 and positivity for ${\omega^2}_{gwas}$, ${\omega^2}_{ctrl}$, and ${\omega^2}_{overall}$. For $d$ to reach statistical significance, we also require ${\omega^2}_{gwas} > 0$. Numerical results are reported in Supplementary Table 12.

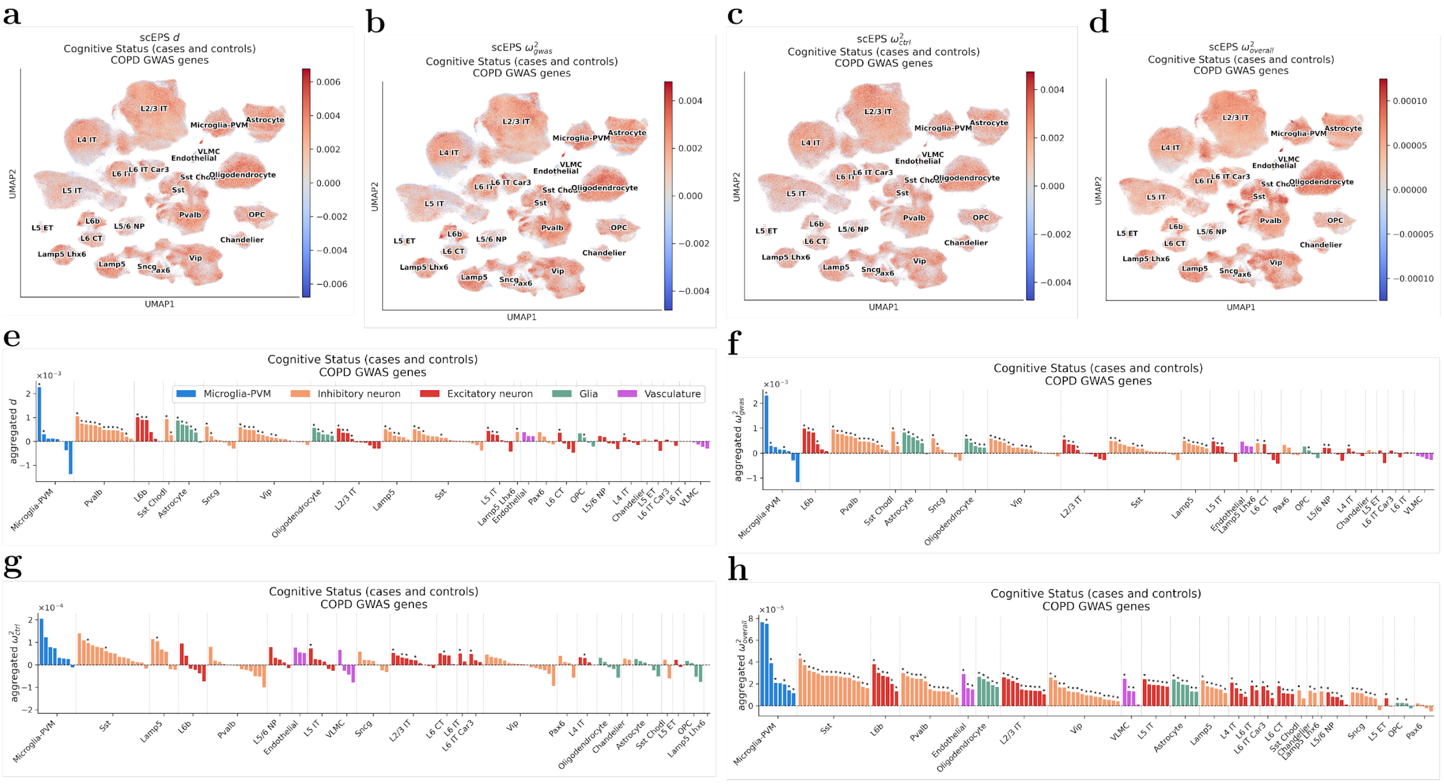


Supplementary Figure 53: **Results from the scEPS analysis of CS using COPD GWAS.** (**a**, **b**, **c**, **d**) UMAP plots showing the scEPS $d$, ${\omega^2}_{gwas}$, ${\omega^2}_{ctrl}$, and ${\omega^2}_{overall}$ statistics for individual cell neighborhoods for CS, respectively. (**e**, **f**, **g**, **h**) We report the aggregated scEPS $d$, ${\omega^2}_{gwas}$, ${\omega^2}_{ctrl}$, and ${\omega^2}_{overall}$ statistics for CS for 139 brain cell subtypes, respectively. "★" denotes statistical significance: FDR < 0.1 and positivity for aggregated ${\omega^2}_{gwas}$, ${\omega^2}_{ctrl}$, and ${\omega^2}_{overall}$. For aggregated $d$ to reach statistical significance, we also require aggregated ${\omega^2}_{gwas} > 0$. Numerical results are reported in Supplementary Table 14.

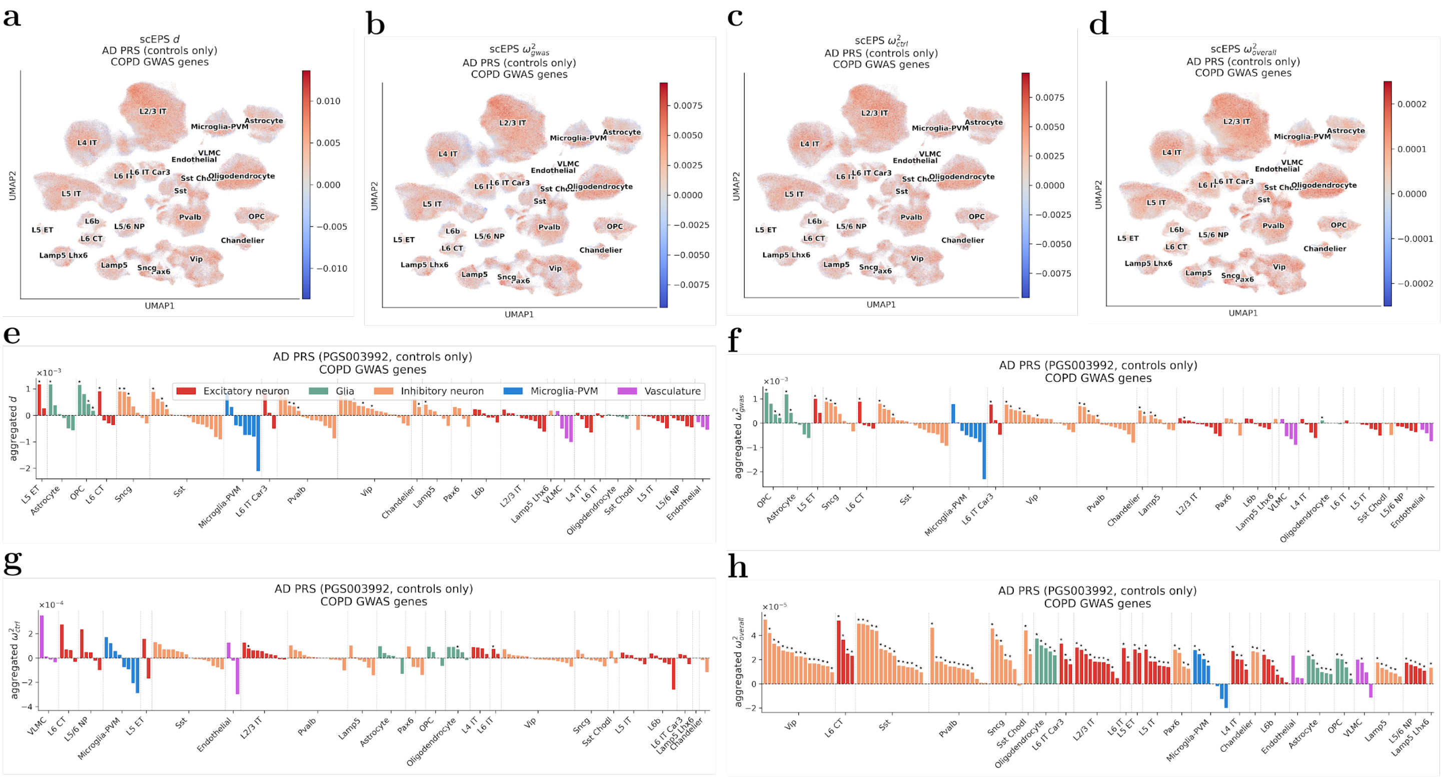


Supplementary Figure 54: **Results from the scEPS analysis of AD PRS (controls only) using COPD GWAS.** (**a**, **b**, **c**, **d**) UMAP plots showing the scEPS $d$, ${\omega^2}_{gwas}$, ${\omega^2}_{ctrl}$, and ${\omega^2}_{overall}$ statistics for individual cell neighborhoods for AD PRS, respectively. (**e**, **f**, **g**, **h**) We report the aggregated scEPS $d$, ${\omega^2}_{gwas}$, ${\omega^2}_{ctrl}$, and ${\omega^2}_{overall}$ statistics for AD PRS for 139 brain cell subtypes, respectively. "★" denotes statistical significance: FDR < 0.1 and positivity for aggregated ${\omega^2}_{gwas}$, ${\omega^2}_{ctrl}$, and ${\omega^2}_{overall}$. For aggregated $d$ to reach statistical significance, we also require aggregated ${\omega^2}_{gwas} > 0$. Numerical results are reported in Supplementary Table 14.

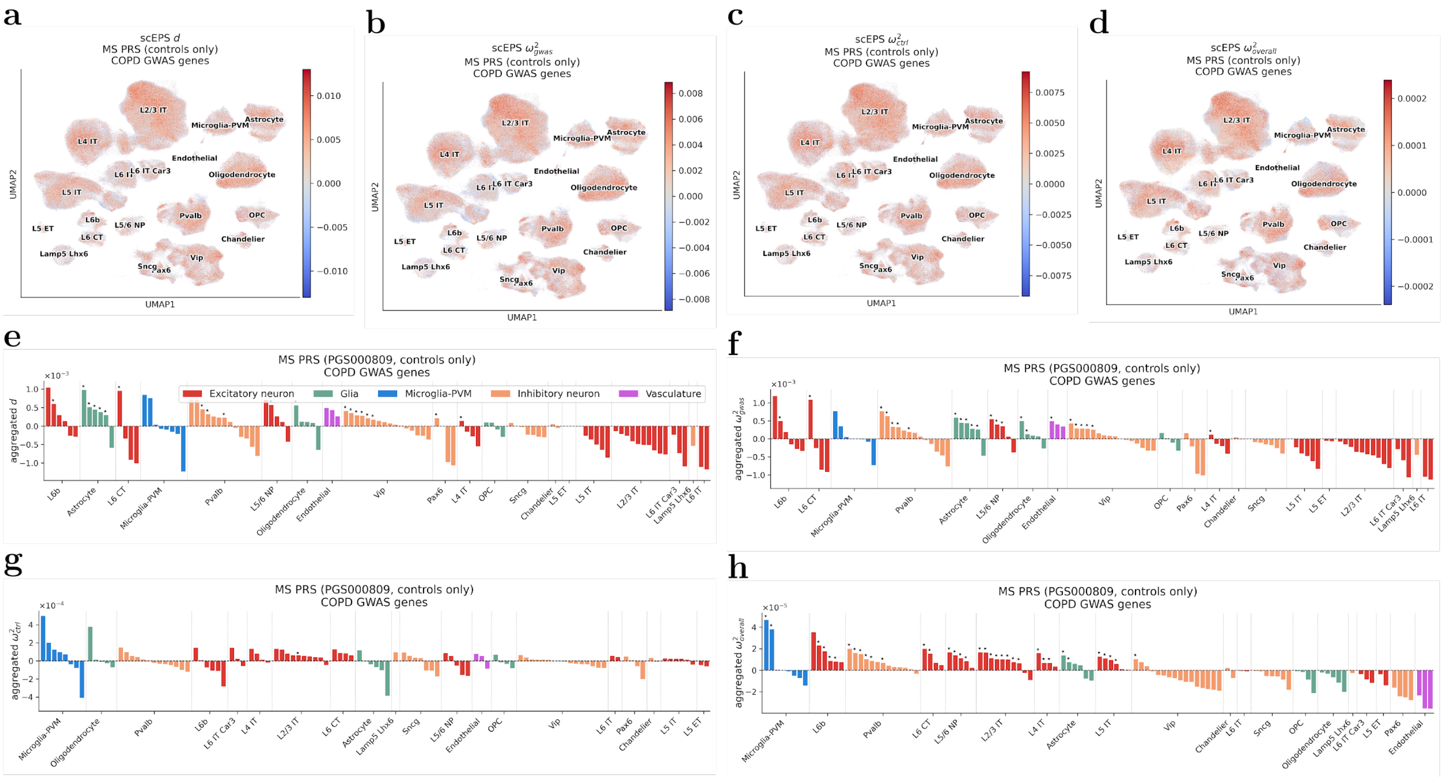


Supplementary Figure 55: **Results from the scEPS analysis of MS PRS (controls only) using COPD GWAS.** (**a**, **b**, **c**, **d**) UMAP plots showing the scEPS $d$, $\omega^2{}_{gwas}$, $\omega^2{}_{ctrl}$, and $\omega^2{}_{overall}$ statistics for individual cell neighborhoods for MS PRS, respectively. (**e**, **f**, **g**, **h**) We report the aggregated scEPS $d$, $\omega^2{}_{gwas}$, $\omega^2{}_{ctrl}$, and $\omega^2{}_{overall}$ statistics for MS PRS for 139 brain cell subtypes, respectively. "★" denotes statistical significance: FDR < 0.1 and positivity for aggregated $\omega^2{}_{gwas}$, $\omega^2{}_{ctrl}$, and $\omega^2{}_{overall}$. For aggregated $d$ to reach statistical significance, we also require aggregated $\omega^2{}_{gwas} > 0$. Numerical results are reported in Supplementary Table 14.

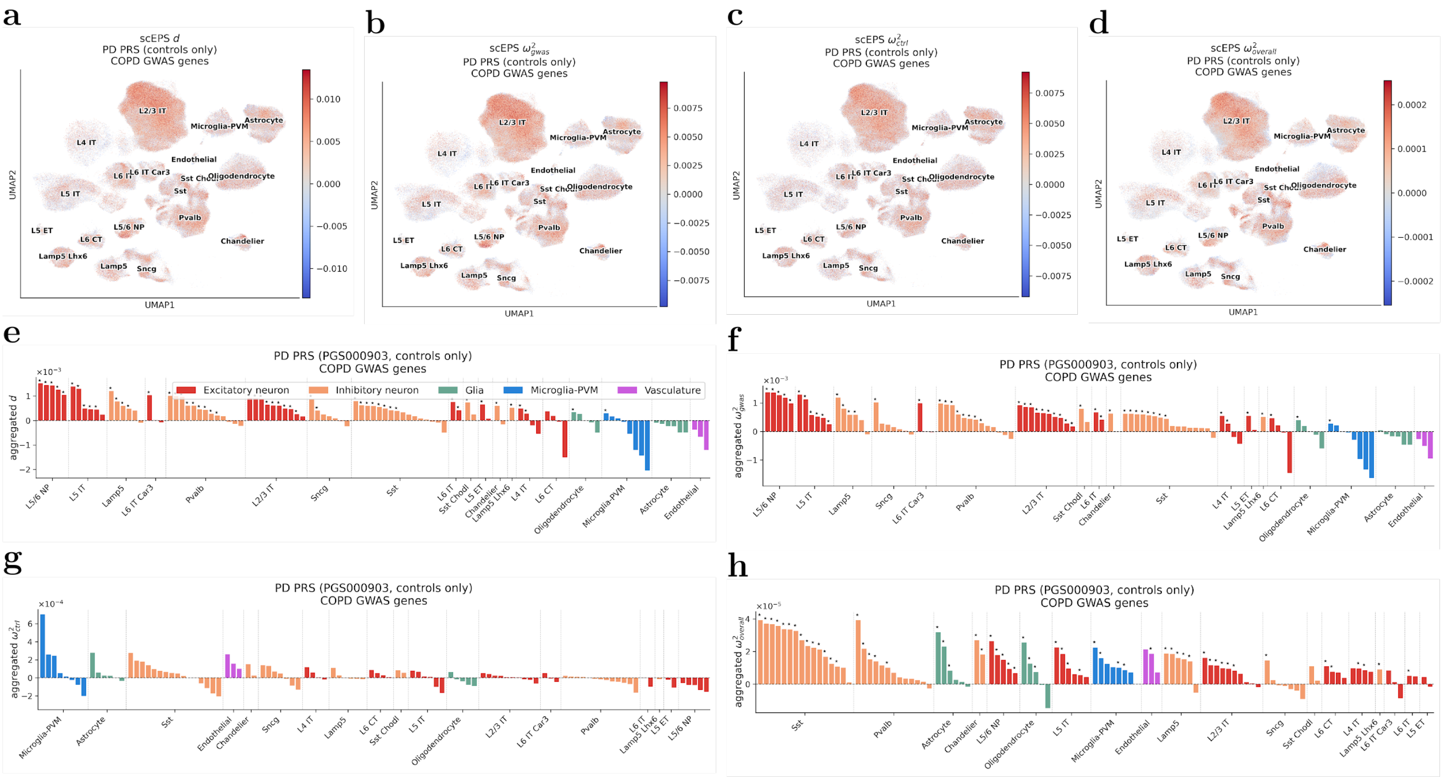


Supplementary Figure 56: **Results from the scEPS analysis of PD PRS (controls only) using COPD GWAS genes.** (**a**, **b**, **c**, **d**) UMAP plots showing the scEPS $d$, $\omega^2{}_{gwas}$, $\omega^2{}_{ctrl}$, and $\omega^2{}_{overall}$ statistics for individual cell neighborhoods for PD PRS, respectively. (**e**, **f**, **g**, **h**) We report the aggregated scEPS $d$, $\omega^2{}_{gwas}$, $\omega^2{}_{ctrl}$, and $\omega^2{}_{overall}$ statistics for PD PRS for 139 brain cell subtypes, respectively. "★" denotes statistical significance: FDR < 0.1 and positivity for aggregated $\omega^2{}_{gwas}$, $\omega^2{}_{ctrl}$, and $\omega^2{}_{overall}$. For aggregated $d$ to reach statistical significance, we also require aggregated $\omega^2{}_{gwas} > 0$. Numerical results are reported in Supplementary Table 14.

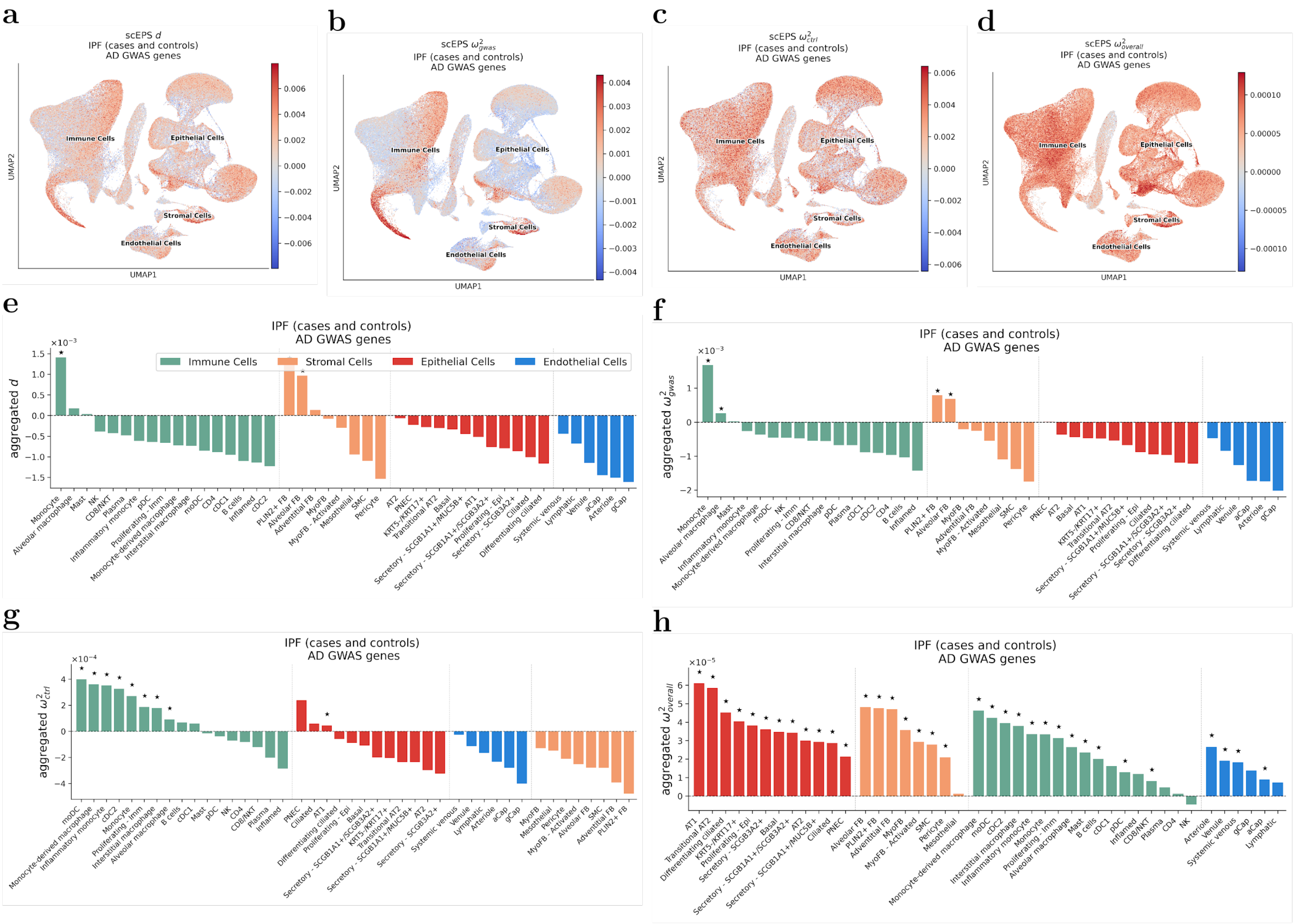


Supplementary Figure 57: **Results from the scEPS analysis of IPF using AD GWAS.** (**a**, **b**, **c**, **d**) UMAP plots showing the scEPS $d$, $\omega^2{}_{gwas}$, $\omega^2{}_{ctrl}$, and $\omega^2{}_{overall}$ statistics for individual cell neighborhoods for IPF, respectively. (**e**, **f**, **g**, **h**) We report the aggregated scEPS $d$, $\omega^2{}_{gwas}$, $\omega^2{}_{ctrl}$, and $\omega^2{}_{overall}$ statistics for IPF for 43 lung cell subtypes, respectively. "★" denotes statistical significance: FDR < 0.1 and positivity for aggregated $\omega^2{}_{gwas}$, $\omega^2{}_{ctrl}$, and $\omega^2{}_{overall}$. For aggregated $d$ to reach statistical significance, we also require aggregated $\omega^2{}_{gwas} > 0$. Numerical results are reported in Supplementary Table 15.

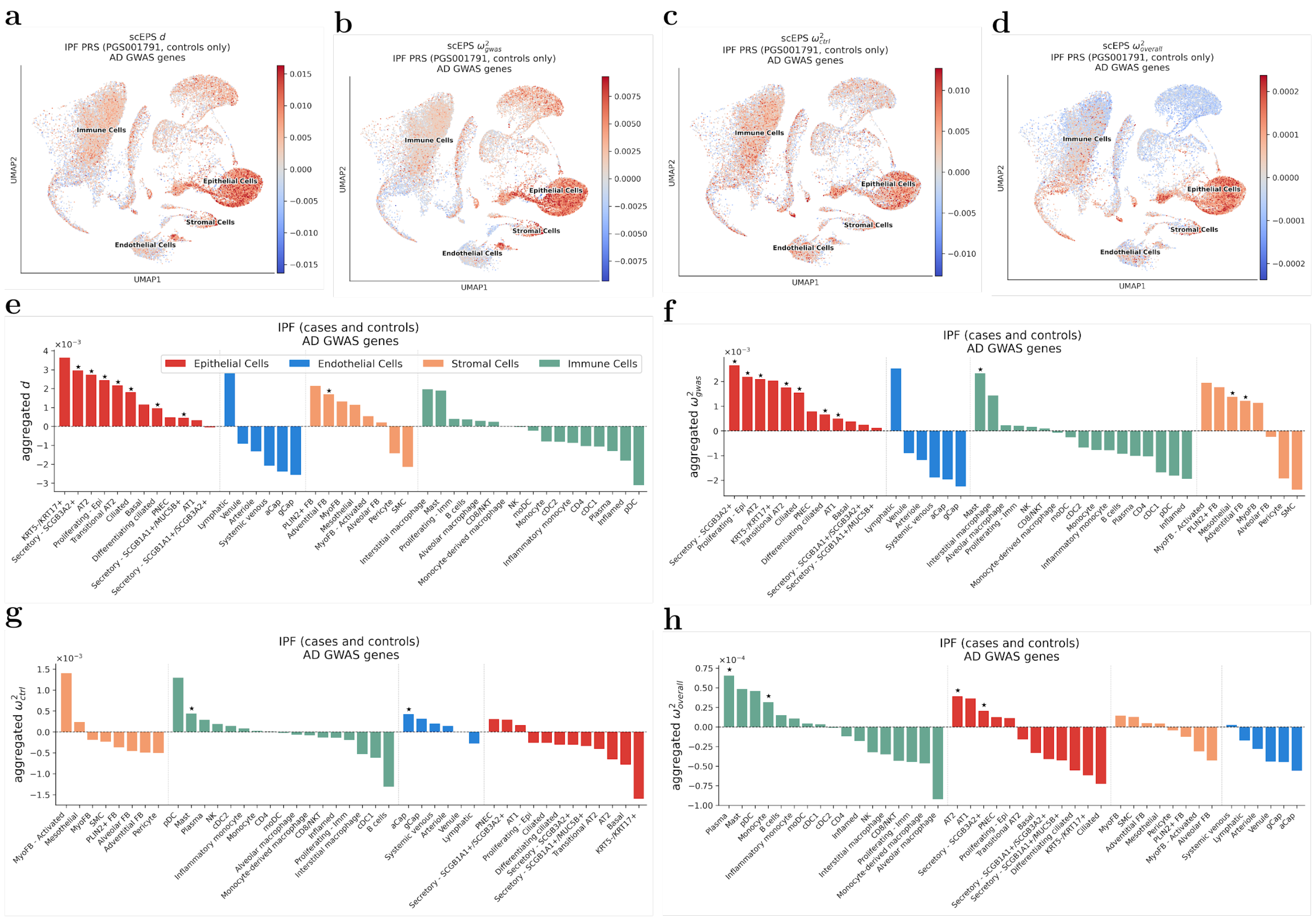


Supplementary Figure 58: **Results from the scEPS analysis of IPF PRS (controls only) using AD GWAS.** (**a**, **b**, **c**, **d**) UMAP plots showing the scEPS $d$, $\omega^2{}_{gwas}$, $\omega^2{}_{ctrl}$, and $\omega^2{}_{overall}$ statistics for individual cell neighborhoods for IPF PRS, respectively. (**e**, **f**, **g**, **h**) We report the aggregated scEPS $d$, $\omega^2{}_{gwas}$, $\omega^2{}_{ctrl}$, and $\omega^2{}_{overall}$ statistics for IPF PRS for 43 lung cell subtypes, respectively. "★" denotes statistical significance: FDR < 0.1 and positivity for aggregated $\omega^2{}_{gwas}$, $\omega^2{}_{ctrl}$, and $\omega^2{}_{overall}$. For aggregated $d$ to reach statistical significance, we also require aggregated $\omega^2{}_{gwas} > 0$. Numerical results are reported in Supplementary Table 15.

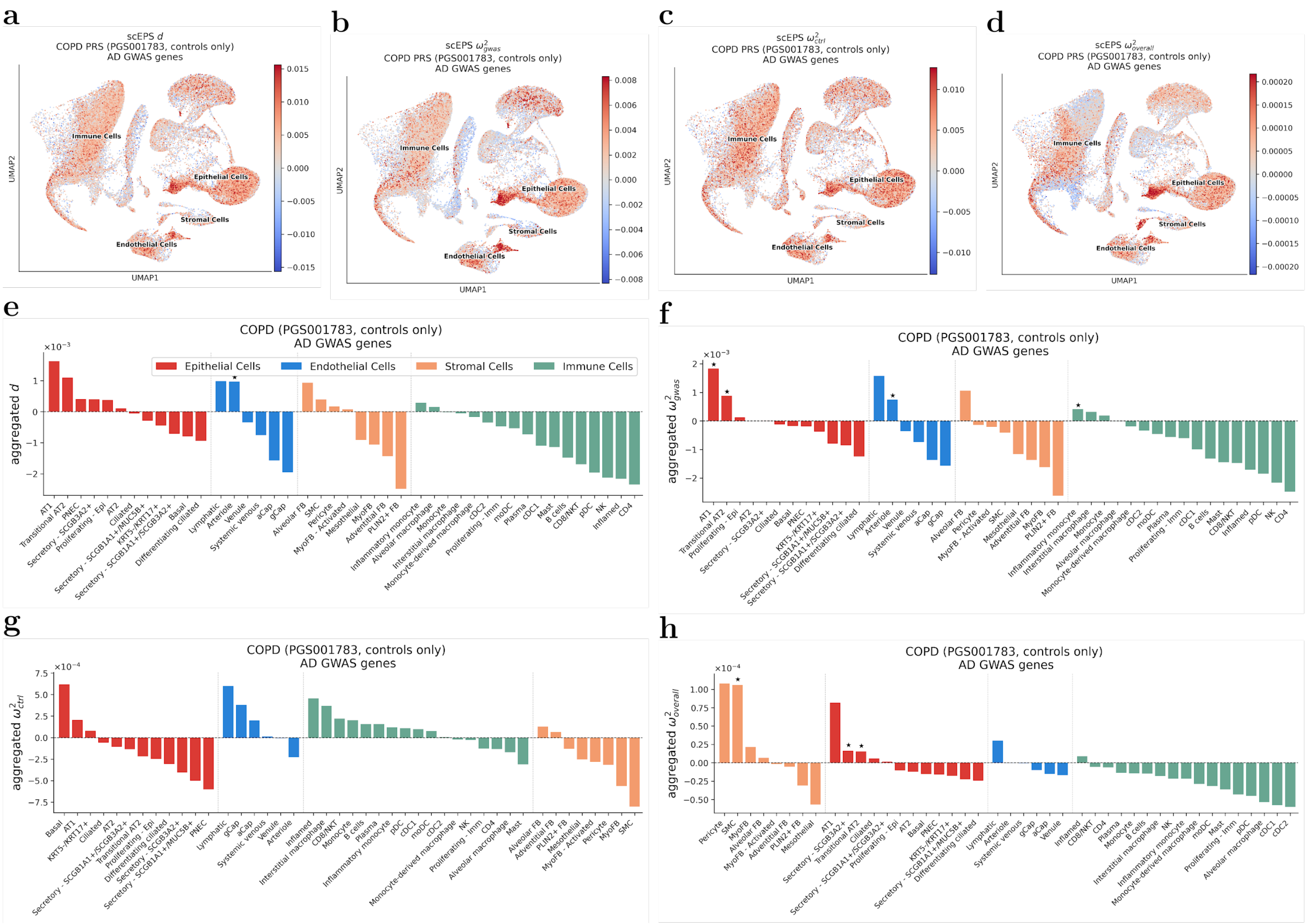


Supplementary Figure 59: **Results from the scEPS analysis of COPD PRS (controls only) using AD GWAS.** (**a**, **b**, **c**, **d**) UMAP plots showing the scEPS $d$, ${\omega^2}_{gwas}$, ${\omega^2}_{ctrl}$, and ${\omega^2}_{overall}$ statistics for individual cell neighborhoods for COPD PRS, respectively. (**e**, **f**, **g**, **h**) We report the aggregated scEPS $d$, ${\omega^2}_{gwas}$, ${\omega^2}_{ctrl}$, and ${\omega^2}_{overall}$ statistics for COPD PRS for 43 lung cell subtypes, respectively. "★" denotes statistical significance: FDR < 0.1 and positivity for aggregated ${\omega^2}_{gwas}$, ${\omega^2}_{ctrl}$, and ${\omega^2}_{overall}$. For aggregated $d$ to reach statistical significance, we also require aggregated ${\omega^2}_{gwas} > 0$. Numerical results are reported in Supplementary Table 15.

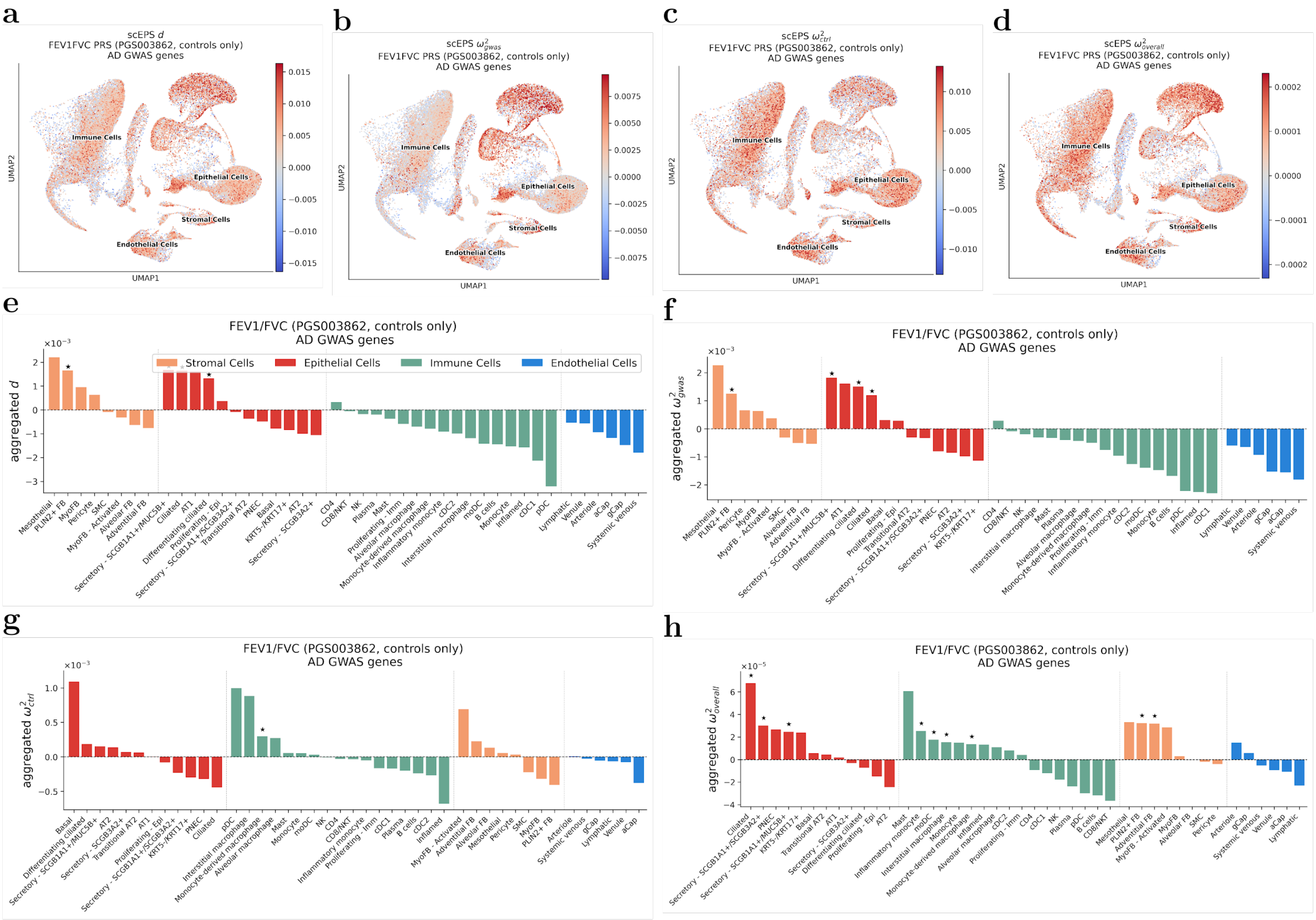


Supplementary Figure 60: **Results from the scEPS analysis of FEV1/FVC PRS (controls only) using AD GWAS.** (**a**, **b**, **c**, **d**) UMAP plots showing the scEPS $d$, $\omega^2{}_{gwas}$, $\omega^2{}_{ctrl}$, and $\omega^2{}_{overall}$ statistics for individual cell neighborhoods for FEV1/FVC PRS, respectively. (**e**, **f**, **g**, **h**) We report the aggregated scEPS $d$, $\omega^2{}_{gwas}$, $\omega^2{}_{ctrl}$, and $\omega^2{}_{overall}$ statistics for FEV1/FVC PRS for 43 lung cell subtypes, respectively. "★" denotes statistical significance: FDR < 0.1 and positivity for aggregated $\omega^2{}_{gwas}$, $\omega^2{}_{ctrl}$, and $\omega^2{}_{overall}$. For aggregated $d$ to reach statistical significance, we also require aggregated $\omega^2{}_{gwas} > 0$ Numerical results are reported in Supplementary Table 15.

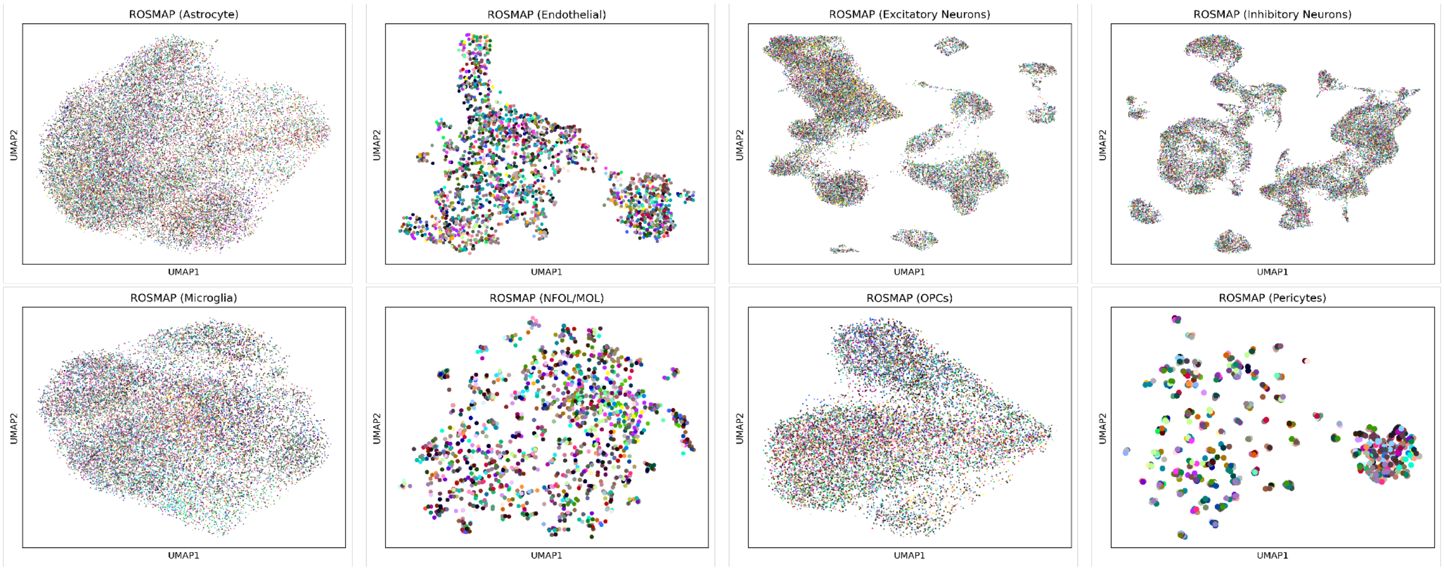


Supplementary Figure 61: **UMAP plots visualizing the cells from the donors in the ROSMAP brain cell atlas data.** Each plot represents one of the 8 cell types annotated in the ROSMAP brain cell atlas data. Different colors in each plot represent different donors. For better visualization, we only visualized cells from 100 randomly selected ROSMAP donors.

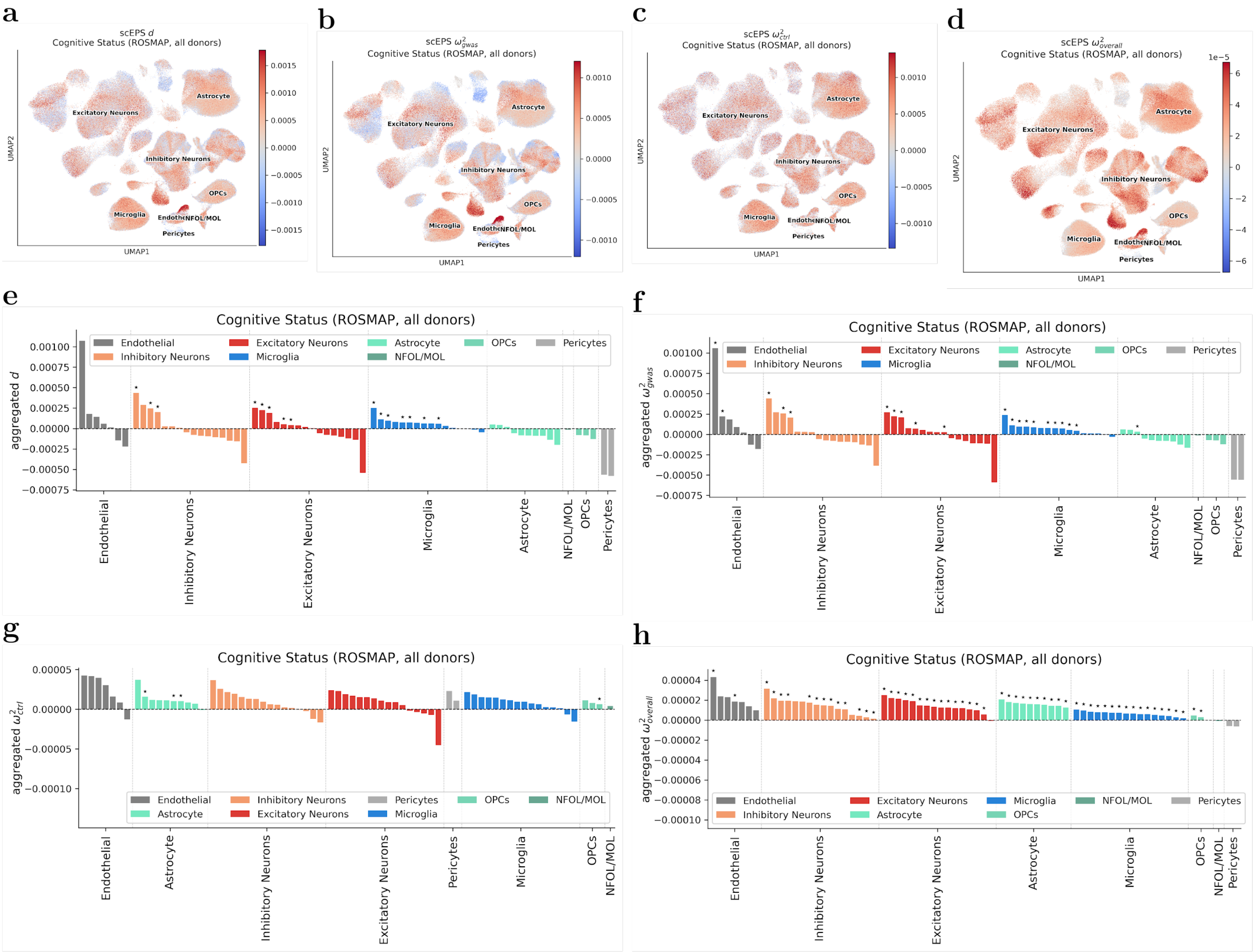


Supplementary Figure 62: **Results from the scEPS analysis of CS using ROSMAP brain cell atlas data.** (**a**, **b**, **c**, **d**) UMAP plots showing the scEPS $d$, ${\omega^2}_{gwas}$, ${\omega^2}_{ctrl}$, and ${\omega^2}_{overall}$ statistics for individual cell neighborhoods for CS, respectively. (**e**, **f**, **g**, **h**) We report the aggregated scEPS $d$, ${\omega^2}_{gwas}$, ${\omega^2}_{ctrl}$, and ${\omega^2}_{overall}$ statistics for CS for 71 ROSMAP brain cell subtypes, respectively. "★" denotes statistical significance: FDR < 0.1 and positivity for aggregated ${\omega^2}_{gwas}$, ${\omega^2}_{ctrl}$, and ${\omega^2}_{overall}$. For aggregated $d$ to reach statistical significance, we also require aggregated ${\omega^2}_{gwas} > 0$. Numerical results are reported in Supplementary Table 18.

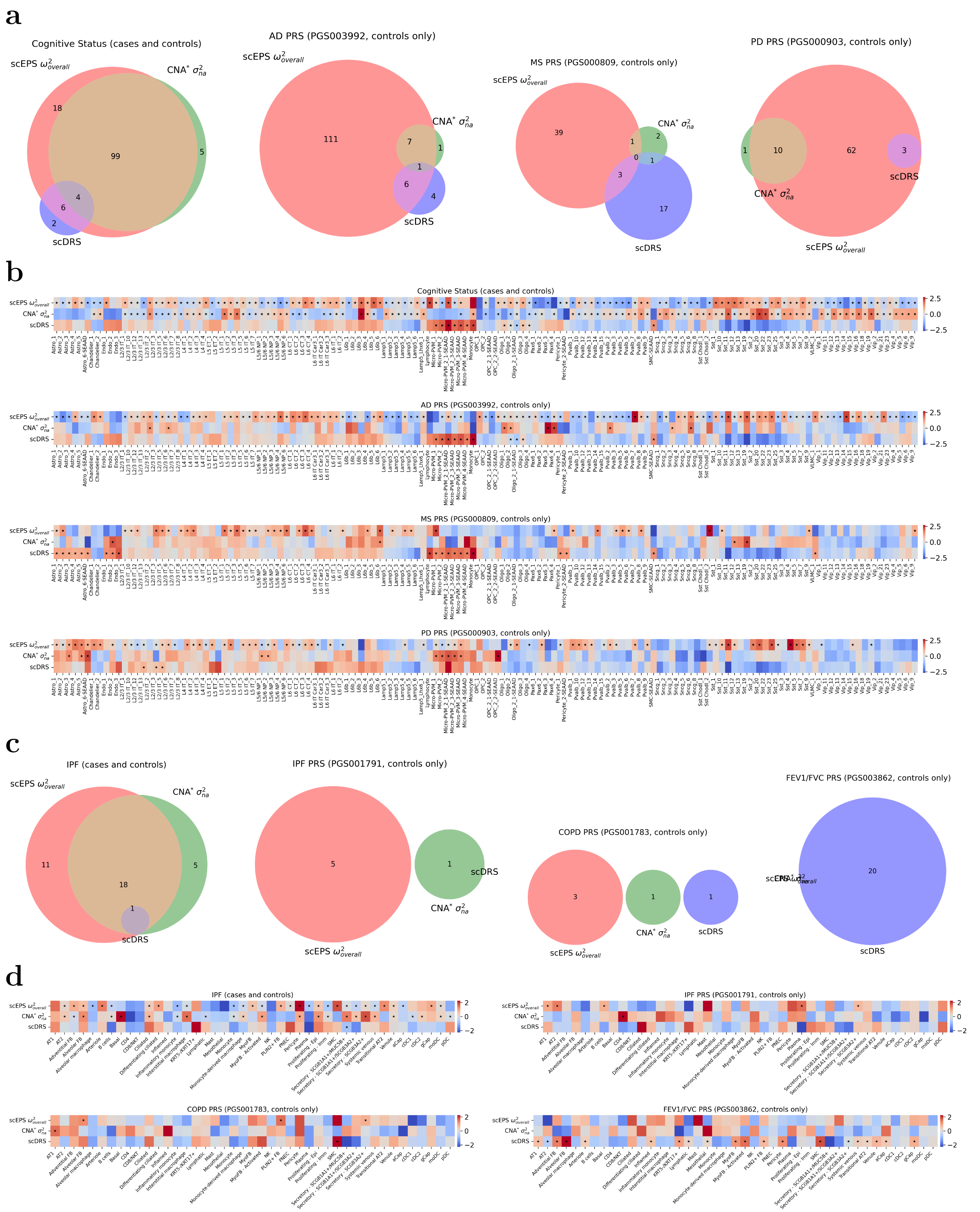


Supplementary Figure 63: **Overlap of disease-associated cell types identified using scEPS $\omega^2_{overall}$ statistics, CNA*, and scDRS.** (**a**) We report the overlap of the significant associations (FDR < 0.1) identified by scEPS (aggregated $\omega^2_{overall}$ > 0), CNA* ($\sigma^2_{na}$ > 0), and scDRS for (left to right) cognitive status, PRSs of AD, MS, and PD, across the 139 brain cell subtypes; we report in (**c**) results for (left to right) IPF, PRSs of IPF, COPD, and FEV1/FVC, across the 43 lung cell subtypes. Note: scDRS did not report any significant association for PD. (**b**) Heat maps showing the inverse normal transformed aggregated scEPS *d* statistics, mean CNA* $\sigma^2_{na}$, and mean scDRS raw statistics for (top to bottom) cognitive status, PRSs of AD, MS, and PD, across the 139 brain cell subtypes; we report in (**d**) results for IPF (top left), PRSs of IPF (top right), COPD (bottom left), and FEV1/FVC (bottom right), across the 43 lung cell subtypes. Significant associations are denoted by stars (★) in the heat maps. Numerical results are reported in Supplementary Table 9, 21, 23.

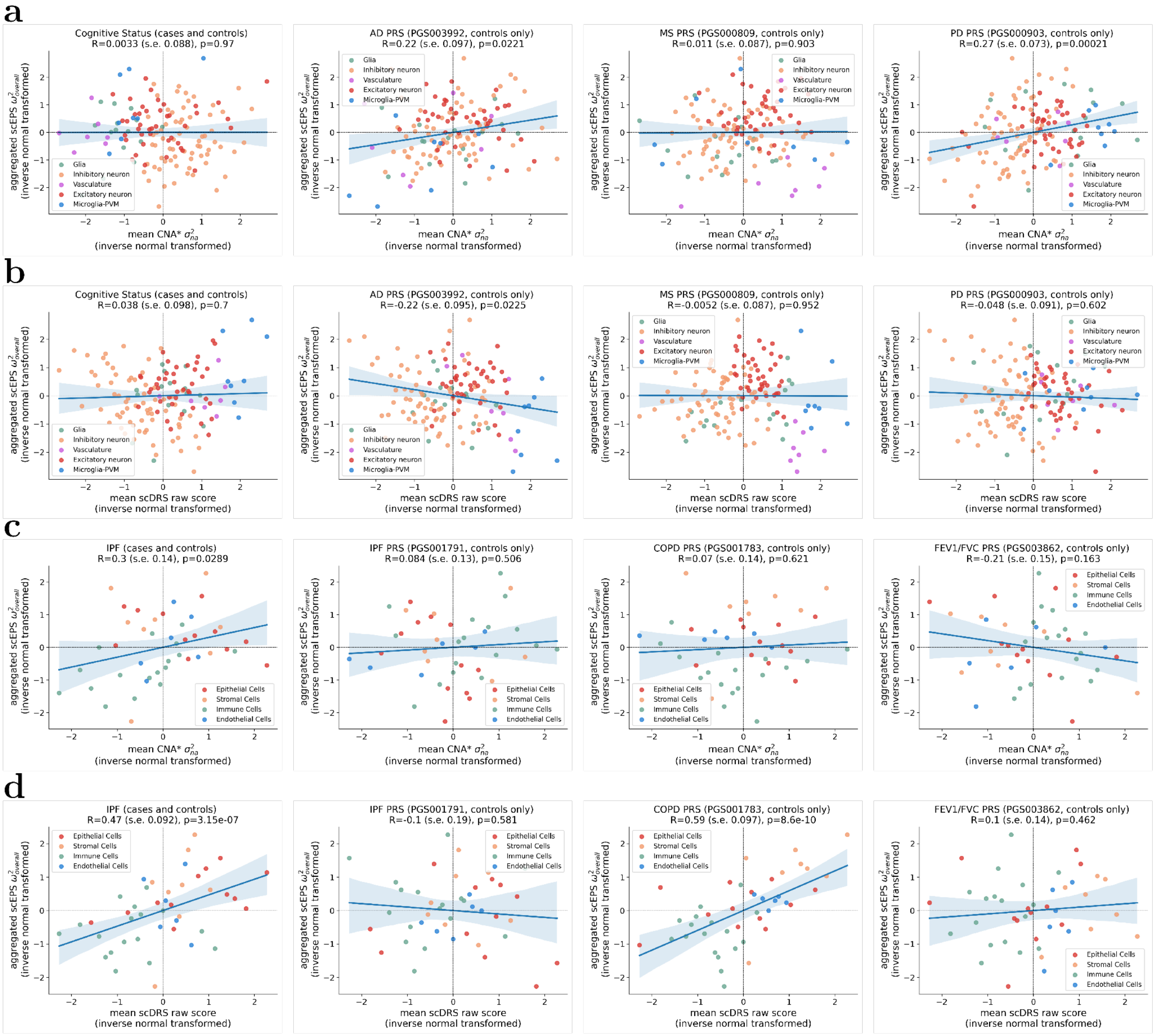


Supplementary Figure 64: **Comparisons between aggregated scEPS $\omega^2{}_{overall}$ statistics vs. CNA* $\sigma^2{}_{na}$ and scDRS raw scores.** (**a**, **b**) Scatter plots showing the inverse normal transformed aggregated $\omega^2{}_{overall}$ statistics vs. CNA* $\sigma^2{}_{na}$ and scDRS raw scores, respectively, across 139 brain cell subtypes, for (left to right) CS, and PRSs of AD, MS, and PD. (**c**, **d**) Similar scatter plots across 43 lung cell subtypes, for (left to right) IPF, and PRSs of IPF, COPD, and FEV1/FVC. Standard errors of the correlations were obtained by statistical bootstrapping across the cell subtypes. Shaded regions of the regression line represent 95% confidence intervals.

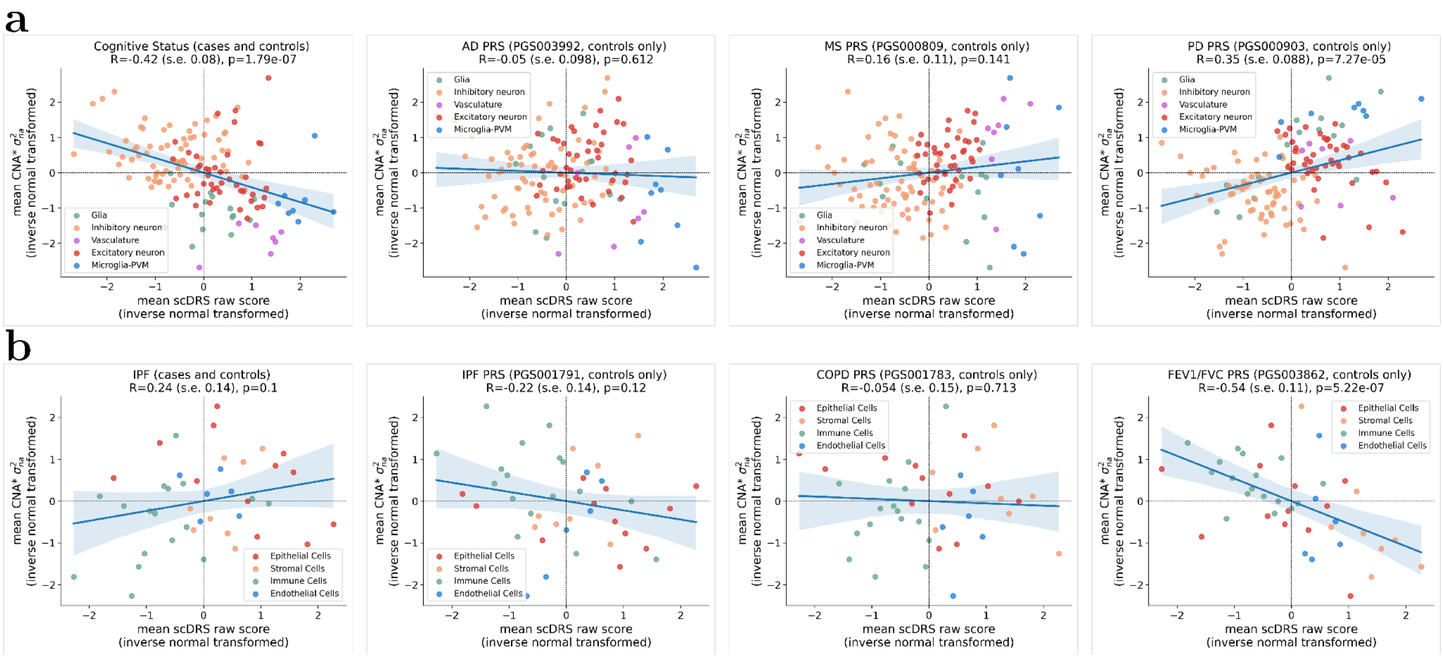


Supplementary Figure 65: **Comparisons between aggregated CNA* $\sigma^2{}_{na}$ vs. scDRS raw scores.** (**a**) Scatter plots showing the inverse normal transformed aggregated CNA* $\sigma^2{}_{na}$ vs. scDRS raw scores, respectively, across 139 brain cell subtypes, for (left to right) CS, and PRSs of AD, MS, and PD. (**b**) Similar scatter plots across 43 lung cell subtypes, for (left to right) IPF, and PRSs of IPF, COPD, and FEV1/FVC. Standard errors of the correlations were obtained by statistical bootstrapping across the cell subtypes. Shaded regions of the regression line represent 95% confidence intervals.

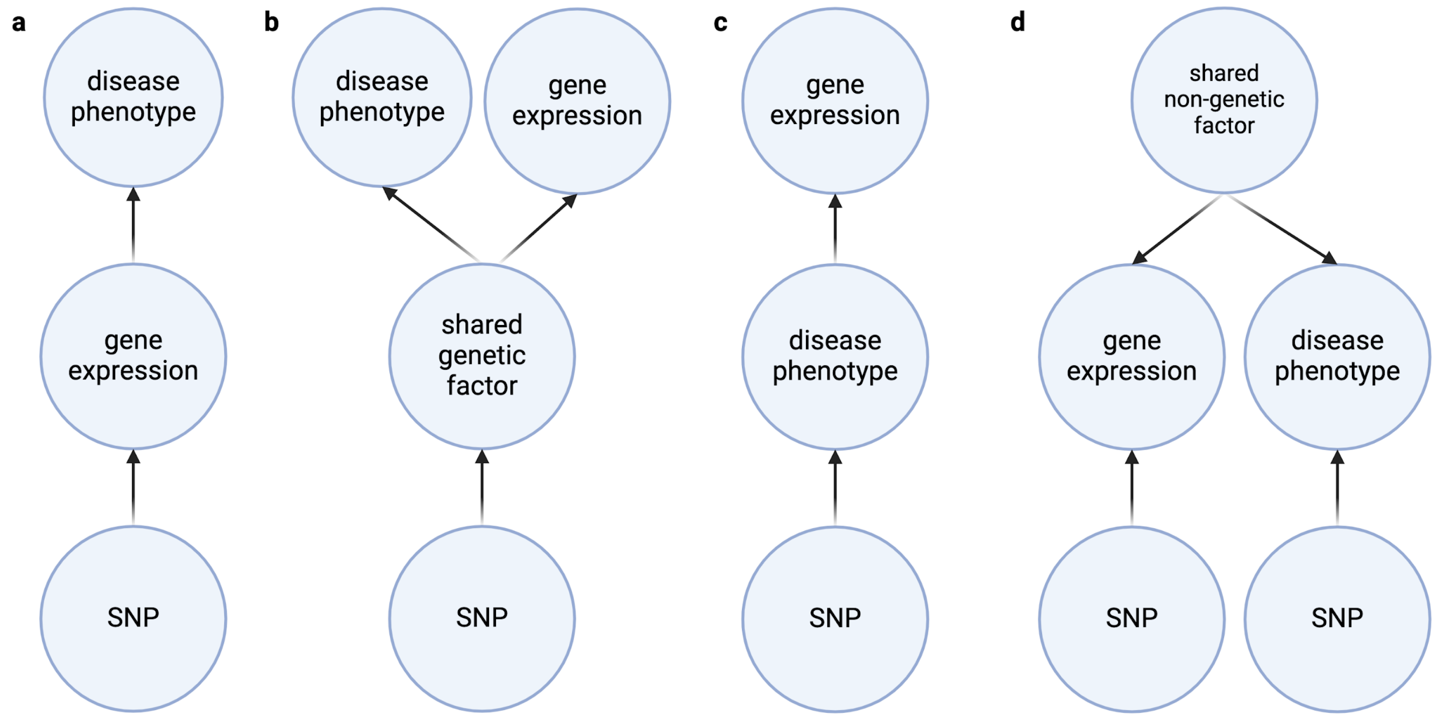


Supplementary Figure 66: **Different models linking genetic variations, gene expression, and disease phenotype.** (**a**) The causality model, in which the gene expression causally impacts the disease. (**b**) The pleiotropy model, in which the disease phenotype and gene expression are both impacted by a shared genetic factor (e.g., expression of another gene, protein, etc.). (**c**) The reverse causality model, in which the gene expression is impacted by the disease phenotype. (**d**) The confounder model, in which the gene expression and disease phenotype are independently impacted by genetic variations, and are both causally impacted by a shared non-heritable confounder (e.g., exposure to a certain environment).
